# Mental and social wellbeing and the UK Coronavirus Job Retention Scheme: evidence from nine longitudinal studies

**DOI:** 10.1101/2021.11.15.21266264

**Authors:** Jacques Wels, Charlotte Booth, Bożena Wielgoszewska, Michael Green, Giorgio Di Gessa, Charlotte F. Huggins, Gareth J. Griffith, Alex S. F. Kwong, Ruth C. E. Bowyer, Jane Maddock, Praveetha Patalay, Richard J. Silverwood, Emla Fitzsimons, Richard Shaw, Ellen J. Thompson, Andrew Steptoe, Alun Hughes, Nishi Chaturvedi, Claire J. Steves, Srinivasa Vittal Katikireddi, George B. Ploubidis

## Abstract

**Background:** The COVID-19 pandemic has led to major economic disruptions. In March 2020, the UK implemented the Coronavirus Job Retention Scheme – known as furlough – to minimize the impact of job losses. We investigate associations between change in employment status and mental and social wellbeing during the early stages of the pandemic.

**Methods:** Data were from 25,670 respondents, aged 17 to 66, across nine UK longitudinal studies. Furlough and other employment changes were defined using employment status pre-pandemic and during the first lockdown (April-June 2020). Mental and social wellbeing outcomes included psychological distress, life satisfaction, self-rated health, social contact, and loneliness. Study-specific modified Poisson regression estimates, adjusting for socio-demographic characteristics and pre-pandemic mental and social wellbeing measures, were pooled using meta-analysis.

**Results:** Compared to those who remained working, furloughed workers were at greater risk of psychological distress (adjusted risk ratio, ARR=1.12; 95% CI: 0.97, 1.29), low life satisfaction (ARR=1.14; 95% CI: 1.07, 1.22), loneliness (ARR=1.12; 95% CI: 1.01, 1.23), and poor self-rated health (ARR=1.26; 95% CI: 1.05, 1.50), but excess risk was less pronounced than that of those no longer employed (e.g., ARR for psychological distress=1.39; 95% CI: 1.21, 1.59) or in stable unemployment (ARR=1.33; 95% CI: 1.09, 1.62).

**Conclusions:** During the early stages of the pandemic, those furloughed had increased risk for poor mental and social wellbeing. However, their excess risk was lower in magnitude than that of those who became or remained unemployed, suggesting that furlough may have partly mitigated poorer outcomes.

## Background

COVID-19 and a series of associated ‘lockdown’ mitigation measures, which included closure of non-essential retail, leisure facilities and schools, have had an adverse impact on the economy in the United Kingdom (UK) and worldwide (Koltai et al., 2020; Office for National Statistics, 2020). There is a well-established relationship between individual employment status and mental health and wellbeing (Di Gessa et al., 2021; Flint et al., 2013; Frasquilho et al., 2016; Parsons et al., 2021; Steele et al., 2013). Existing literature on the effects of economic downturns on population health and health-related behaviours is complex and suggests effects are context-specific and vary across generations and between different demographic and socioeconomic groups (Catalano et al., 2011; Copeland et al., 2015; Valkonen et al., 2000).

Overall, it has been estimated that the prevalence of mental distress in the UK increased from 19.1% pre-pandemic to 30.6% in early lockdown, with greater deteriorations observed in young adults and women (Banks & Xu, 2020; Niedzwiedz et al., 2021). More recent longitudinal research has found raised levels of psychological distress were sustained across subsequent stages of the pandemic, particularly for women and young adults (Patel et al., 2021). However, it is unclear how employment status change is related to mental and social wellbeing in this unique context.

Employment is generally considered to be associated with good health (Benach et al., 2010; Graetz, 1993) and job loss or unemployment with deleterious physical and mental health (Puig-Barrachina et al., 2011), including lower psychological wellbeing (Murphy & Athanasou, 1999) and increased mortality (Roelfs et al., 2011). Men and those in their early and middle career stage can be especially affected (Roelfs et al., 2011), although some studies have found greater effects of unemployment for women (Drivakis, 2015). Unemployment is also sometimes associated with social isolation (Lobo, 2018), which can lead to loneliness (Green et al., 2021).

People with pre-existing mental health problems were more likely to experience employment disruption during the pandemic (Di Gessa et al 2021; Breslau et al., 2021), but it remains unclear how policies introduced to mitigate economic disruption might affect mental health. The UK government launched the Coronavirus Job Retention Scheme (CJRS, widely referred to as ‘furlough ‘) in March 2020, providing employees who were unable to work due to the pandemic with 80% of pay (capped at £2,500 per month) (Adams-Prassl et al., 2020). Furlough differs from becoming unemployed for several reasons. First, furlough schemes reduce uncertainty, as cessation of work is intended to be temporary. Second, a substantial portion of income is maintained. However, while furlough helps maintain many of the advantages of employment, other benefits, such as time structure, collective purpose, social contact, and activity are likely diminished for furloughed workers (Paul & Batinic, 2010). Thus, the implications of the novel UK furlough scheme for mental and social wellbeing remain unclear.

By bringing together data from nine UK longitudinal studies, we investigate how furlough and other employment changes are associated with psychological distress, life satisfaction, self-rated health, social contact, and loneliness, during the early stages of the pandemic, when furlough was at its peak (between 25 and 30 percent of the UK population were furloughed between April and July 2020 and only 10 to 20 percent in the following months (ONS, 2021)). It is plausible that employment distruption will not affect all groups equally, therefore we examine whether associations differ by sex, age, education, and household composition.

## Method

### Participants and design

Participants were 25,670 respondents from nine UK population-based longitudinal studies, who completed surveys both before and during the COVID-19 pandemic. Pandemic data were collected between April-June 2020 and pre-pandemic data constituted the most recent data available for each study prior to the pandemic (median was ~3 years earlier, with a range from 1-14 years). Further details of the design, sampling frame, age range, timing of the pre-pandemic and COVID-19 surveys, response rates, and sample size are in <u>Supplementary File 1</u>.

Five studies were age homogenous birth cohorts: the Millennium Cohort Study (MCS); the children in the Avon Longitudinal Study of Parents and Children (ALSPAC-G1); Next Steps (NS, formerly the Longitudinal Study of Young People in England); the 1970 British Cohort Study (BCS70); and the 1958 National Child Development Study (NCDS). Four age heterogenous studies were included: Understanding Society (USOC); the English Longitudinal Study of Ageing (ELSA); the Scottish Family Health Study: Generation Scotland (GS); and the UK ‘s largest adult twin registry (TwinsUK). Finally, the parents of the ALSPAC-G1 cohort were treated as a fifth age heterogenous study population (ALSPAC-G0). All studies, except TwinsUK and Generation Scotland, are representative of the British population in their target age range (see Supplementary File 1 for further details).

Analytical samples were restricted to working age participants, defined as those aged 16 to 66 (the current state pension age in the UK), who had at least one wellbeing outcome in the COVID-19 survey and relevant pre-pandemic measures for confounder adjustment. Studies were weighted to be representative of their target population, accounting for sampling design and differential non-response (see, for instance, Brown et al. (2020)). Weights were not available for GS.

### Measures

Please, see <u>Supplementary File 2</u> for full details on the measures and variable coding in each study.

#### Exposure: Employment status change

Employment change (or stability) was operationalised by comparing respondents’ self-reported employment status during the initial stages of the pandemic and retrospectively in the months preceding the start of the pandemic. Based on this information, we created six employment change (or stability) categories: stable employed (either as self-employed or an employee, which served as the reference group); furloughed (i.e., from employed to furlough); no longer employed (i.e., from employed to not working, such as job loss or retirement); stable unemployed (i.e., unemployed at both points); became employed (i.e., from not working to employed); and stable non-employed (i.e., not available for employment at either point, including in education, early retirement, caring responsibilities, sick or disabled).

#### Outcomes: Mental health and social wellbeing

We investigated six different mental and social wellbeing outcomes. For each outcome, we created a binary variable using pre-validated cut-off scores where possible. **Psychological distress** was measured using the Kessler-6 (MCS) (Kessler et al., 2002), General Health Questionnaire-12 (NS, USOC) (Goldberg, 1978), Malaise Inventory (BCS, NCDS) (Rutter, 1970), Centre for Epidemiological Studies Depression Scale (ELSA) (Radloff, 1977), Short Mood and Feelings Questionnaire (ALSPAC G0/G1) (Angold et al., 1995), Patient Health Questionnaire (GS) (Kroenke & Spitzer, 2002), and Hospital Anxiety and Depression Scale (Twins UK) (Zigmond & Snaith, 1983). **Life satisfaction** was assessed using the Office for National Statistics (ONS) wellbeing scale that asks participants to rate how satisfied they are with their lives (most studies used a 0-10 scale; USOC used 1-7): those who answered less than 7 (or less than 5 in USOC) were classified as reporting low life satisfaction. **Self-rated health** was measured using responses to a generic question asking participants to rate their health on a five-point ordinal scale (excellent; very good; good; fair; poor): the five items were dichotomised into poor (fair or poor) versus good (excellent, very good or good**). Social contact** (either face-to-face, by telephone, or text message) with family and friends outside the household was coded to distinguish between those reporting daily versus less than daily social contact. **Loneliness** was assessed (MCS, NS, BCS, NCDS, ELSA, TwinsUK) using the short version of the Revised UCLA loneliness scale, with scores of 6 and higher indicating high loneliness (Russell et al., 1980). Additionally, we also considered the direct question “How often do you feel lonely?” rated on a three-point ordinal scale (hardly ever; some of the time; often), as this was asked in two further studies (USOC, GS): we compared those reported often feeling lonely versus less frequent or no feelings of loneliness.

#### Confounders and moderators

Two levels of confounder adjustment were applied. The first level reflected a *basic* adjustment, accounting for sociodemographic characteristics: age (for age heterogeneous studies), sex, ethnicity (White vs. non-White ethnic minority - not available in NCDS and BCS), education (degree vs. no degree – parent education used for MCS), UK nation (England; Scotland; Wales; Northern Ireland), and household composition (living alone; with partner including possible children or others; with others e.g., housemates or other family members but no partner). The second level reflected *full* adjustment, additionally including all available pre-pandemic mental and social wellbeing measures, in order to determine whether differences in outcomes could be attributed to changes taking place during the pandemic.

### Analysis

Within each study, each of the mental and social wellbeing outcomes were regressed on employment status change, using a modified Poisson model with robust standard errors that returns risk ratios for ease of interpretation and to avoid issues related to non-collapsibility of odds ratios (Zou, 2004; Zou & Donner, 2013). A sensitivity analysis was conducted with the continuous version of psychological distress (standardised within studies), using linear regression. We focus on reporting risk ratios comparing stable employment to furlough, no longer employed, and stable unemployment, as the main exposure categories of interest. Results from each study were statistically pooled using a random effects meta-analysis with restricted maximum likelihood (maximum likelihood was used for models that failed to converge). Study-specific estimates were excluded if the number of individuals reporting the outcome of interest was very low (≤ 2). Stratification by sex, age, education, and household composition was assessed with sub-group analyses using the *full* level of confounder adjustment (i.e., controlling for sociodemographic and pre-pandemic mental and social wellbeing). Sub-group differences that were significant at the *p* < .05 threshold are reported in the text. See <u>Supplementary File 3</u> for further information and full model estimates, and <u>Supplementary File 4</u> for figures of the sub-group analyses.

## Results

### Descriptive statistics

Descriptive statistics for mental and social wellbeing outcomes across nine studies are presented in Table 1. During the pandemic, the proportion of participants displaying psychological distress ranged from 7.2% (ALSPAC G0) to 35.7% (NS). The proportion reporting low life satisfaction ranged from 18.2% (ALSPAC G0) to 48.8% (MCS). The proportion reporting poor self-rated health ranged from 6.8% (GS) to 22.0% (ELSA). The proportion reporting less than daily social contact ranged from 5.8% (ALSPAC G0) to 81.2% (USOC). The proportion displaying high loneliness (UCLA scale) ranged from 20.8% (NCDS) to 53.5% (TwinsUK). The proportion reporting feeling often lonely ranged from 4.9% (GS) to 22.6% (MCS).

**Table 1.** Descriptive statistics of mental health and social wellbeing outcomes pre- and during initial stages of the pandemic by study.

|  | MCS | NS | BCS | NCDS | ELSA | USOC | ALSPAC-G0 | ALSPAC-G1 | GS | TWINS-UK |
| --- | --- | --- | --- | --- | --- | --- | --- | --- | --- | --- |
| Age/Age range | 18-20 | 30-31 | 50 | 62 | 52-66 | 17-66 | 50-65 | 27-29 | 27-66 | 22-65 |
|  | % (N) | % (N) | % (N) | % (N) | % (N) | % (N) | % (N) | % (N) | % (N) | % (N) |
| ***Psychological distress*** |  |  |  |  |  |  |  |  |  |  |
| pre-pandemic | 17.8 (338) | 25.4 (432) | 19.1 (493) | 14.4 (508) | 12.9 (272) | 22.2 (1268) | 19.6 (336) | 18.8 (205) | 11.1 (294) | 7.8 (64) |
| during | 19.0 (386) | 35.7 (595) | 17.2 (481) | 12.2 (436) | 22.8 (505) | 33.3 (1991) | 7.2 (108) | 17.3 (182) | 9.8 (243) | 12.6 (105) |
| ***Low life satisfaction*** |  |  |  |  |  |  |  |  |  |  |
| pre-pandemic | - | 9.0 (144) | 22.1 (594) | 22.6 (836) | 29.5 (620) | 29.2 (1619) | 20.4 (346) | 16.4 (166) | 14.1 (374) | - |
| during | 48.8 (863) | 32.1 (525) | 27.8 (813) | 25.6 (993) | 36.0 (807) | 37.6 (2181) | 18.2 (305) | 28.0 (276) | 47.2 (1253) | 40.6 (382) |
| ***Poor self-rated health*** |  |  |  |  |  |  |  |  |  |  |
| pre-pandemic | 7.0 (111) | 9.7 (138) | 19.1 (442) | 16.8 (519) | 22.0 (443) | 20.3 (1055) | - | - | - | 9.9 (85) |
| during | 9.5 (163) | 9.4 (139) | 13.0 (324) | 17.1 (548) | 22.0 (457) | - | - | - | 6.8 (180) | 8.4 (79) |
| ***Less than daily social contact*** |  |  |  |  |  |  |  |  |  |  |
| pre-pandemic | - | - | - | - | 51.6 (1214) | 78.0 (5160) | - | - | 56.8 (1505) | - |
| during | 60.8 (1135) | 64.2 (1024) | 63.3 (2007) | 55.6 (2523) | 65.3 (1524) | 81.2 (5279) | 5.8 (100) | 8.1 (90) | 56.1 (1486) | 11.7 (110) |
| ***High loneliness (UCLA)*** |  |  |  |  |  |  |  |  |  |  |
| pre-pandemic | - | - | - | - | 21.6 (455) | - | - | - | - | - |
| during | 44.0 (782) | 29.9 (480) | 21.2 (623) | 20.8 (809) | 24.4 (528) | - | - | - | - | 53.5 (494) |
| ***Often lonely*** |  |  |  |  |  |  |  |  |  |  |
| pre-pandemic | 13.7 (224) | 8.2 (172) | 7.9 (215) | 8.3 (283) | 6.8 (143) | 9.9 (477) | - | - | 1.0 (26) | - |
| during | 22.6 (403) | 11.0 (187) | 6.9 (180) | 6.8 (231) | 6.8 (136) | 10.4 (465) | - | - | 4.9 (129) | 5.0 (46) |
| Total N | 1,839 | 1,595 | 3,143 | 4,416 | 2,344 | 6,849 | 1,469 | 1,051 | 2,652 | 978 |

**Note:** Data were collected during the early stages of the COVID-19 pandemic (April-June 2020); Pre-pandemic data were collected at different times ranging from 2006-2019 (see Supplementary File 1 for more information); Missing items reflect that no consistent measure was available for that particular study.

### Employment change

Descriptive statistics for employment status change (and stability) across nine studies are displayed in Table 2. The proportion of participants in stable employment ranged from 10.9% (MCS) to 71.5% (ALSPAC G1). The proportion of participants who were furloughed ranged from 5.8% (TwinsUK) to 23.2% (BCS). The proportion of participants no longer employed ranged from 1.7% (BCS) to 7.1% (ALSPAC G0). The proportion of participants who were stable unemployed ranged from 0.5% (BCS, GS) to 8.6% (ALSPAC G0).

**Table 2.** Percent distribution of change in employment status during the pandemic across nine studies.

|  | **MCS** | **NS** | **BCS** | **NCDS** | **ELSA** | **USOC** | **ALSPAC**<br>**G0** | **ALSPAC**<br>**G1** | **GS** | **TwinsUK** |
| --- | --- | --- | --- | --- | --- | --- | --- | --- | --- | --- |
| **Age/Age range** | 18-20 | 30-31 | 50 | 62 | 52-66 | 17-66 | 50-65 | 27-29 | 27-66 | 22-65 |
| Stable employed | 10.9 | 62.2 | 65.4 | 33.9 | 50.3 | 58.7 | 53.8 | 71.5 | 62.5 | 30.6 |
| Furloughed | 14.8 | 22.3 | 23.2 | 19.1 | 13.8 | 14.3 | 13.1 | 15.8 | 8.2 | 5.8 |
| No longer employed | 3.5 | 3.1 | 1.7 | 2.8 | 2.0 | 3.6 | 7.1 | 4.6 | 3.4 | 1.8 |
| Became employed | 1.5 | 1.1 | 0.5 | 0.8 | 0.4 | 1.3 | 3.4 | 2.2 | 0.6 | 0 |
| Stable unemployed | 3.1 | 1.7 | 0.5 | 1.5 | 2.9 | 1.8 | 8.6 | 2.7 | 0.5 | 0.8 |
| Stable non-employed | 66.2 | 9.6 | 8.7 | 41.9 | 30.6 | 20.3 | 14 | 3.2 | 24.9 | 60.9 |
| *N* | 1,839 | 1,595 | 3,143 | 4,416 | 2,344 | 6,849 | 1,469 | 1,051 | 2,652 | 978 |

### Main results

The pooled results suggest a largely common pattern in the way employment change was associated with mental and social wellbeing outcomes (see Figure 1). Compared to those in stable employment, those furloughed, no longer employed, and stable unemployed tended to show excess risk for poor mental and social wellbeing, with magnitude of excess risk being largest for the stable unemployed, followed by those no longer employed, and then those furloughed.

**Figure 1.**
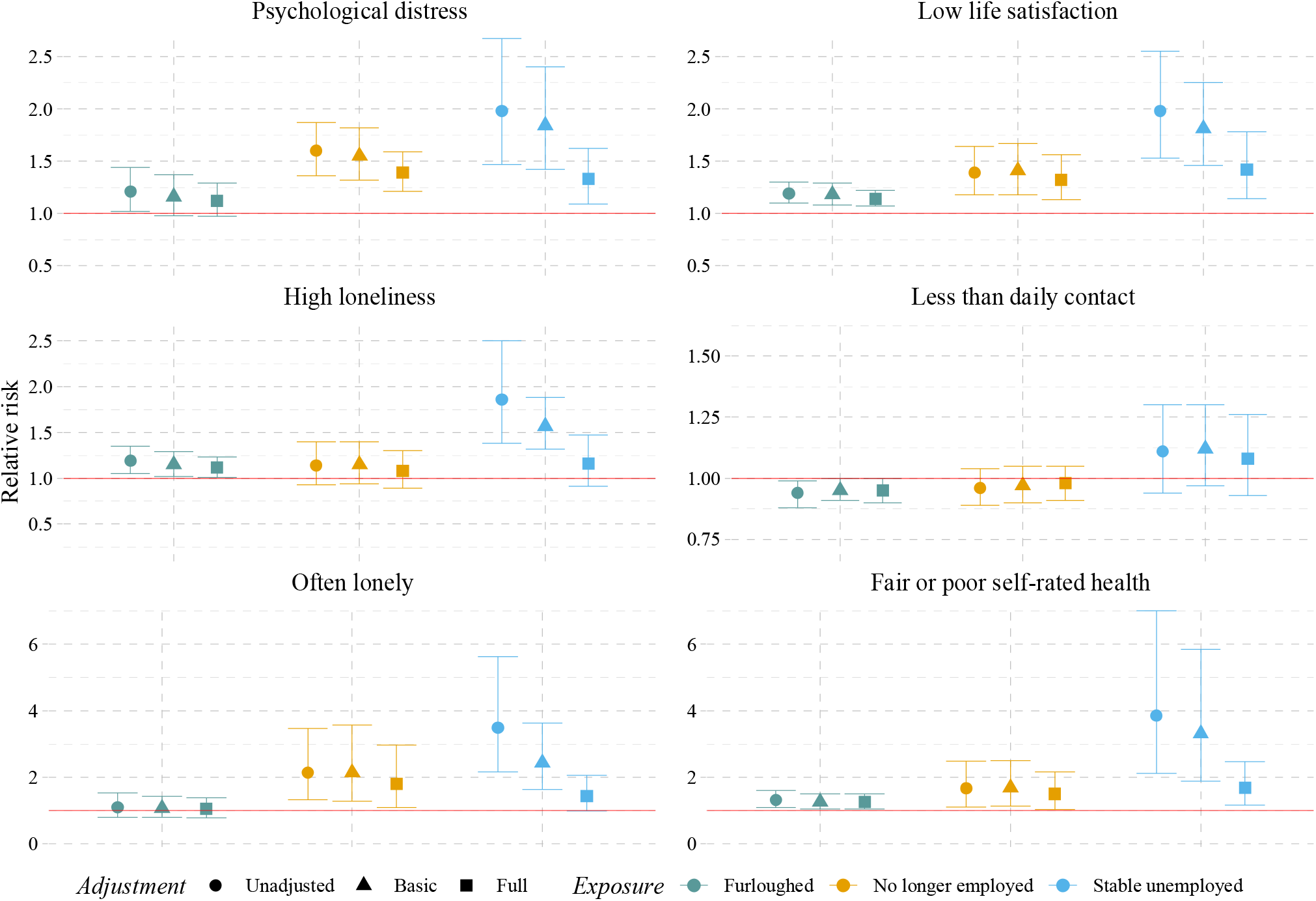
Relative risk of employment status change in mental and social wellbeing. **Caption:** Error bars show 95% confidence intervals; Stable employment is the reference category; Basic adjustment includes: age, sex, ethnicity, education, household composition; Full adjustment includes: psychological distress, life satisfaction, self-rated health, social contact, loneliness.

#### Psychological distress

In unadjusted models, compared to participants in stable employment, those furloughed had higher psychological distress (RR=1.21; 95% CI: 1.02, 1.44; I² = 60%), as did those no longer employed (RR=1.60; 95% CI: 1.36, 1.87; I² = 0%), and those in stable unemployment (RR=1.98; 95% CI: 1.47, 2.67; I² = 50%). In the fully adjusted model, controlling for sociodemographic and pre-pandemic mental and social wellbeing, estimates were attenuated for furlough (ARR=1.12; 95% CI: 0.97, 1.29; I² = 49%), those no longer employed (ARR=1.39; 95% CI: 1.21, 1.59; I² = 0%), and those in stable unemployment (ARR=1.33; 95% CI: 1.09, 1.62; I² = 50%). The sensitivity analysis conducted with the continuous version of psychological distress confirmed these results. Sub-group analyses revealed no differences by sex, education, age, or household composition (see <u>Supplementary File 3</u> for full model estimates).

#### Low life satisfaction

In unadjusted models, compared to participants in stable employment, those furloughed had lower life satisfaction (RR=1.19; 95% CI: 1.10, 1.30; I² = 24%), as did those no longer employed (RR=1.39; 95% CI: 1.18, 1.64; I² = 45%), and those in stable unemployment (RR=1.98; 95% CI: 1.53, 2.55; I² = 76%). Estimates were attenuated in the fully adjusted model, but less so for furlough (ARR=1.14; 95% CI: 1.07, 1.22; I² = 7%) and those no longer employed (ARR=1.32; 95% CI: 1.13, 1.56; I² = 52%), than the stable unemployed (ARR=1.42; 95% CI: 1.14, 1.78; I² = 65%). Sub-group analyses revealed no differences by sex, education, age, or household composition.

##### Poor self-rated health

Compared to stable employment, risk of poor self-rated health was higher in the unadjusted model for furlough (RR=1.32; 95% CI: 1.09, 1.60; I² = 43%), no longer being employed (RR=1.67; 95% CI: 1.11, 2.49; I² = 61%), and stable unemployment (RR=3.85; 95% CI: 2.12, 7.01; I² = 85%). Estimates were attenuated in the fully adjusted model, with a similar pattern of milder attenuation for furlough (ARR=1.26; 95% CI: 1.05, 1.50; I² = 44%) and those no longer employed (ARR=1.50; 95% CI: 1.04, 2.17; I² = 59%), compared to those in stable unemployment (ARR=1.69; 95% CI: 1.16, 2.47; I² = 65%).

Sub-group analyses revealed differences by sex (*p* = .009), where furlough was associated with poorer self-rated health for females (ARR=1.41; 95% CI: 1.11, 1.79; I² = 49%), compared to males (ARR=1.01; 95% CI: 0.97, 1.07; I² = 0%). Differences were also observed by age (*p* = .019), with no longer being employed being more strongly associated with poorer self-rated health among those aged 30-49 years (ARR=2.86; 95% CI: 1.28, 6.36; I² = 0%), compared to those aged 50+ (ARR=1.28; 95% CI: 0.95, 1.71; I² = 42%); estimates for ages 16-29 years were not available due to data sparsity.

#### Less than daily social contact

We observed no differences in the risk of less than daily social contact across employment groups in all models. Sub-group analyses revealed no differences by sex, education, age, or household composition.

#### High loneliness

Compared to stable employment, furlough was associated with higher loneliness in the unadjusted model (RR=1.19; 95% CI: 1.05, 1.35; I² = 27%), no longer being employed showed a similar magnitude association but confidence intervals crossed the null (RR=1.14; 95% CI: 0.93, 1.40; I² = 0%), and there was a stronger association for stable unemployment (RR=1.86; 95% CI: 1.38, 2.50; I² = 50%). Yet, in the fully adjusted model, only those furloughed had increased risk for high loneliness (ARR=1.12; 95% CI: 1.01, 1.23; I² = 0%). Sub-group analyses revealed no differences by sex, education, age, or household composition.

#### Often lonely

In the unadjusted model with the single-item loneliness measure, compared to those in stable employment, there was no clear association with furlough (RR=1.10; 95% CI: 0.80, 1.53; I² = 66%), but those no longer employed were more likely to report feeling lonely (RR=2.14; 95% CI: 1.32, 3.47; I² = 68%), as were those in stable unemployment (RR=3.49; 95% CI: 2.17, 5.63; I² = 61%). Results were attenuated in the fully adjusted model for those no longer employed (ARR=1.80; 95% CI: 1.09, 2.97; I² = 72%) and stable unemployed (ARR=1.43; 95% CI: 0.99, 2.06; I² = 42%).

Sub-group analyses revealed differences by sex (*p* = .051), whereby no longer being employed was strongly associated with feeling lonely for females (ARR=2.39; 95% CI: 1.41, 4.08; I² = 72%), but not males (ARR=1.40; 95% CI: 0.60, 3.30; I² = 60%). There were also differences by household composition (*p* < .001), whereby stable unemployment was more strongly associated with feeling lonely for those living with a partner (and possibly other family members) (ARR=4.04; 95% CI: 2.28, 7.18; I² = 4%), than for those living alone (ARR=2.07; 95% CI: 1.32, 3.25; I² = 60%), or those living with others but no partner (ARR=1.00; 95% CI: 0.69, 1.44; I² = 0%).

## Discussion

Across nine UK longitudinal studies, in their totality representative of the UK population, we found that furlough was associated with a slight decline in mental and social wellbeing compared to stable employment during the early stages of the COVID-19 pandemic. While raised risks of psychological distress, low life satisfaction, poor self-rated health, and loneliness were seen among furloughed people, the excess risk was generally smaller than that associated with no longer being employed or being in stable unemployment. There was little association between employment status and having daily social contact.

Understanding the impacts of furlough is important because it was a key policy measure implemented to mitigate the economic disruption of the pandemic. Due to the UK CJRS furlough scheme, unemployment only rose moderately compared to other countries (Küçük et al., 2021), which is confirmed in our studies, as the number of furloughed workers was more than three times higher than the number of employees who lost their job. Unlike traditional forms of unemployment, the relationship between specific labour market policy interventions, such as furlough, and health is less well-understood (Korpi, 1997). This is partly because job retention schemes, which focus on buffering the impact of economic downturns via subsidised employment, were uncommon in Western countries prior to the COVID-19 outbreak (Puig-Barrachina et al., 2020).

The existing studies on subsidised employment show inconsistent, although mostly beneficial effects on health and wellbeing (Puig-Barrachina et al., 2020; Wels & Hamarat, 2021). For example, focusing on the restaurant industry in the US using cross-sectional data, Bufquin et al. (2021) showed that working employees experienced higher levels of psychological distress, drug, and alcohol use than temporary unemployed workers. However, Korpi (1997) showed that individuals in subsidised employment occupy an intermediate position in terms of wellbeing, where they are better-off than unemployed individuals, but worse-off than those in regular employment, and our findings largely concur with this pattern. A key explanation concerns the nature of these different employment statuses. Furloughed workers had more security than those who were no longer employed, as they were expected to be reinstated into employment as soon as it was safe for them to do so. Furthermore, they still received 80% of their pay (Burchell, 2011; Maier et al., 2006) which could at least partially protect against the long- and short-term health effects of income loss (Björklund & Eriksson, 1998; Dooley et al., 1996). Moreover, we observed when adjusting for pre-pandemic characteristics, that the excess risk associated with stable unemployment was more strongly attenuated than that for furlough or no longer being employed. This indicates that the large magnitude risks associated with stable unemployment may have had more to do with characteristics that were already established before the pandemic.

Previous research shows that economic disruptions during the pandemic were not experienced by all groups equally. Younger workers, low earners, and women were more likely to work in disrupted sectors, and therefore become unemployed or furloughed (Burchell et al., 2020). People in lower skilled jobs, living in more deprived areas, or struggling financially were also more likely to be furloughed (Gray et al., 2021). Women with young children were more likely to be furloughed (Wielgoszewska et al., 2020) and previous studies found that, during the school closure period, women took on a bigger share of housework and childcare responsibilities (Zamarro & Prados, 2021; Zhou et al., 2020). We found little evidence of wellbeing impacts varying between population sub-groups, although there was a slight increase in poorer self-rated health among furloughed women compared to men. The overall lack of gender differences in how employment status change was associated with mental and social wellbeing might be because women who remained employed, as well as those furloughed, experienced increased burdens and stress equally during the pandemic.

As with most observational studies, unobserved confounding could have affected our estimates. Despite being embedded within long standing cohorts, survey responses during the pandemic were lower than typically achieved, and while weighting was employed to correct for this, bias due to selective non-response could not be ruled out (Fernández-Sanlés et al., 2021; Mostafa et al., 2021)). There are other limitations that should also be considered. First, we were not able to achieve full harmonisation of measures across studies, for example, a range of different psychological distress scales were used and questions on social contact differed considerably (which may explain some of the between study differences in prevalence). Second, all cohorts and studies could not contribute to every analysis as the number of cases and available data varied between studies. Third, participation in the furlough scheme was more common during the initial stages of the pandemic than being no longer employed or in stable unemployment, which meant that estimates for the latter groups were based on small numbers with considerable heterogeneity, especially in sub-group analyses. Finally, it is important to recognise that stable employment itself may have changed during the pandemic with potential effects of home working and changes in working practices on mental health and wellbeing, which is an area for future research.

The UK CJRS furlough scheme officially ended on the 30^th^ of September 2021. It might be expected that the economic downturn caused by the COVID-19 pandemic will last beyond the end of the furlough scheme, and potentially beyond the end of the pandemic (Whitehead et al., 2021). With potentially damaging effects on mental health and wellbeing for those who stopped working (via furlough or otherwise), one pertinent question is whether the mental health and wellbeing of those who were furloughed will recover when they move back to their previous employment status. In line with this, another important question is whether those who benefited from the CJRS scheme will be more likely to experience further economic disruptions such as job or income loss in the post-furlough period, as this could exacerbate detrimental effects on health and wellbeing.

## Conclusion

During the initial stages of the COVID-19 pandemic, many people experienced employment disruption, which we found to be associated with change in mental and social wellbeing. Compared to those who remained working, furloughed workers showed a decline with respect to their mental and social wellbeing. However, the negative mental health impacts appeared worse for those who had left employment or remained unemployed. This suggests that furlough may have helped to mitigate some of the detrimental impacts of employment disruption on mental health, but nevertheless, furloughed workers still experienced a modest deterioration in their mental and social wellbeing and may need additional support to recover from pandemic-related disruptions.

## Data Availability

All datasets included in this analysis have established data sharing processes, and for most included studies the anonymised datasets with corresponding documentation can be downloaded for use by researchers from the UK Data Service. All datasets included in this analysis have established data sharing processes, and for most included studies the anonymised datasets with corresponding documentation can be downloaded for use by researchers from the UK Data Service. We have detailed the processes for each dataset in Supplementary File 1.

https://ukdataservice.ac.uk/

## List of abbreviations

ARR: Adjusted Risk Ratio
ALSPAC-G1: Avon Longitudinal Study of Parents and Children
ALSPAC-G0: Parents of ALSPAC-G1.
BCS70: 1970 British Cohort Study
CI: Confidence interval
CJRS: Coronavirus Job Retention Scheme
ELSA: English Longitudinal Study of Ageing
GS: Generation Scotland: the Scottish Family Health Study
MCS: Millennium Cohort Study
NCDS: 1958 National Child Development Study
NS: Next Steps (formerly the Longitudinal Study of Young People in England)
UK: United Kingdom
USOC: Understanding Society

## Additional Files

Supplementary File 1: Study Description

Supplementary File 2: Variable Coding

Supplementary File 3: Model Estimates

Supplementary File 4: Figures for subgroup analyses

## Declarations

### Ethics approval and consent to participate

We have detailed the ethical approval for each study in Supplementary File 1.

### Availability of data and materials

All datasets included in this analysis have established data sharing processes, and for most included studies the anonymised datasets with corresponding documentation can be downloaded for use by researchers from the UK Data Service. We have detailed the processes for each dataset in Supplementary File 1.

## Role of funder

The funders had no role in the methodology, analysis or interpretation of the findings presented in this manuscript.

### Author Contribution Statement

Conceptualised the study and design: Wels; Booth; Wielgoszewska; Green; Di Gessa Ploubidis, Katikireddi

Designed the methodology: Wels; Booth; Wielgoszewska; Green; Di Gessa Ploubidis, Katikireddi, Silverwood; Maddock

Conducted the formal analysis: Wels; Booth; Wielgoszewska; Green; Di Gessa; Huggins; Griffith; S. F. Kwong; Bowyer

Data curation: Wels; Booth; Wielgoszewska; Green; Di Gessa; Huggins; Griffith; S. F. Kwong; Bowyer

Wrote the original draft of the manuscript: Wels; Booth; Wielgoszewska;

Data visualisation: Wels and Di Gessa.

All authors contributed to critical revision of the manuscript.

Supervision: Ploubidis and Katikireddi.

Funding acquired: Patalay, Katikireddi, Ploubidis, Steves, Silverwood, and Chaturvedi.

## Acknowledgements

The contributing studies have been made possible because of the tireless dedication, commitment and enthusiasm of the many people who have taken part. We would like to thank the participants and the numerous team members involved in the studies including interviewers, technicians, researchers, administrators, managers, health professionals and volunteers. We are additionally grateful to our funders for their financial input and support in making this research happen.

GS: Drew Altschul, Chloe Fawns-Ritchie, Archie Campbell, Robin Flaig, David Porteous

ALSPAC: Daniel J Smith, Nicholas J Timpson, Kate Northstone

Understanding Society: Michaela Benzeval

MCS, NS, BCS70, NCDS: Morag Henderson and colleagues in survey, data, and cohort maintenance teams

