## Supplementary File 2 for "Mental and social wellbeing and the UK Coronavirus Job Retention Scheme: evidence from nine longitudinal studies"

**Supplementary File 2 - Variables coding**

| 1. Measures of Psychological Distress | | | | | | |
| --- | --- | --- | --- | --- | --- | --- |
|  | *Covid Surveys* | | | *Pre-Covid Surveys* | | |
| **Study** | **Questions asked (exact wording)  or Scale used (full name of scale, but don't list all items):** | **Range; Cut-off Score** | **When assessed:** | **Questions or Scale (if different, otherwise just put 'same'):** | **Possible Answer categories/range:** | **When assessed:** |
| ELSA | CES - Depression scale | range 0-8; cut-off score 4+ | June/July 2020 | same | 0-8 (can be dichotomised) | 2018/19 |
| MCS | Kessler6 - Anxiety and Depression | range 0-24; cut-off score 13+ | April-May 2020 | same | same | MCS(2018) |
| BCS70 | Malaise Inventory: 9-items psychological distress | range 0-9; cut-off score 4+ | April-May 2021 | same | same | BCS70(2016) |
| NCDS | Malaise Inventory: 9-items psychological distress | range 0-9; cut-off score 4+ | April-May 2021 | same | same | NCDS(2008) |
| NS | GHQ12 - Psychiatric screening | range 0-12; cut-off score 4+ | April-May 2022 | same | same | NS(2015) |
| ALSPAC G1 | Short Mood and Feelings Questionnaire (depressive symptoms) | range 0-26; cut-off score 11+ | March-June 2020 | same | same | 10 occasions (2001-2018) |
| ALSPAC G0 | Short Mood and Feelings Questionnaire (depressive symptoms) | range 0-26; cut off score 11+ | March-June 2020 | EPDS | 0-30 (can be dichotomised) | 1991-2013 (12 occasions) |
| USOC | GHQ12 - Psychiatric screening | range 0-12; cut-off score 4+ | April - June 2020 | same | same | wave 10 (2018-2019)+annually before that too |
| TwinsUK | Hospital Anxiety Despression Scale | range 0-21; cut-off score 11+ | April 2020 - | same | same | 2006, 2017 |
| GS | Patient Health Questionnaire-9 | range 0-27; cut-off score 10+ | April - May 2020 | Whether they had depression pre-covid | binary | 2006 - 2011 |

| 2. Measures of Loneliness | | | |  |  |  |
| --- | --- | --- | --- | --- | --- | --- |
|  | *Covid Surveys* | | | *Pre-Covid Surveys* | | |
| **Study** | **Questions asked (exact wording)  or Scale used (full name of scale, but don't list all items):** | **Possible Answer categories/range:** | **When assessed:** | **Questions or Scale (if different, otherwise just put 'same'):** | **Possible Answer categories/range:** | **When assessed:** |
| ELSA (indirect measure) | 3-item UCLA Loneliness scale | Range 3-9; cut-off score 6 | June-July 2020;  Nov-Dec 2020 | same | same | Wave 9 (2018/19) in self-completion Q |
| ELSA (direct measure) | How often do you feel lonely? | 1. Hardly ever or never 2. Some of the time 3. **Often** | June-July 2020;  Nov-Dec 2020 | same | same | Wave 9 (2018/19) in self-completion Q |
| NCDS, BCS70, NS, & MCS | 3-item UCLA Loneliness scale. | Range 3 to 9, cut-off score 6 | April-May 2020 | no |  |  |
| NCDS, BCS70, NS, & MCS | How often do you feel lonely? | 1. Hardly ever 2. Some of the time 3. **Often** | April-May 2021 | "Have you felt close to other people" (MCS 2018; BCS 2016; NCDS 2013). "People around are willing to listen to me" (NS 2015) |  |  |
| USOC | In the last 4 weeks, how often did you feel lonely? | 1. Hardly ever 2. Some of the time 3. **Often** | April 2020-Jan 2021 | same | same | Wave 9 (2017-2018)+biannually or so before that |
| TwinsUK | 3-item UCLA loneliness scale | Range 3-9; cut-off score 6 | April 2020- | Retrospective within covid wave - same | same | Retrospective April 2020 |
| TwinsUK | How often do you feel alone? | 1. Hardly ever 2. Some of the time 3. **Often** | April 2020- | As above | same | Retrospective April 2021 |
| GS | How often have you felt lonely **during the past week**? | 1. None, or almost none of the time 2. Some of the time 3. **Most of the time** 4. **All, or almost all of the time** 99. Don't know 98. Prefer not to answer | April - May 2020; July - August 2020; February 2021 | Think back to **before COVID-19 measures were introduced** (i.e., January 2020), how often did you feel lonely then? | same | Retrospective April 2020 |

### 3. Measures of Life Satisfaction or General Happiness

|  | *Covid Surveys:* | | | *Pre-Covid Surveys:* | | |
| --- | --- | --- | --- | --- | --- | --- |
| **Study** | **Questions asked (exact wording) or Scale used (full name of scale, but don't list all items):** | **Possible Answer categories/range:** | **When assessed:** | **Questions or Scale (if different, otherwise just put 'same'):** | **Possible Answer categories/range:** | **When assessed:** |
| ELSA | “On a scale of 0 to 10, where 0 is “not at all” and 10 is “very”, how satisfied are you with your life nowadays?” | 0/10 | June/July 2020 | Overall, how satisfied are you with your life nowadays? | 0/10 | Wave 9 (2018/19) in self-completion Q |
|  |  |  | Nov/Dec 2020 |  |  |  |
| ELSA | “On a scale of 0 to 10, where 0 is “not at all” and 10 is “very”, how happy, overall, did you feel yesterday?” | 0/10 | June/July 2020 | Overall, how happy did you feel yesterday? | 0/10 | Wave 9 (2018/19) in self-completion Q |
|  |  |  | Nov/Dec 2020 |  |  |  |
| NCDS, BCS70, NS, & MCS | "Overall, how satisfied are you with your life nowadays, where 0 means 'not at all' and 10 means 'completely" | Range 0-10 | April-May 2020 | "How dissatisfied or satisfied are you about the way your life has turned out so far?" |  | (NS 2015; BCS70 2016; NCDS 2013) |
| USOC | How satisfied are you currently with your life overall? | 1. Completely dissatisfied | May 2020-Jan2021 | minor difference in wording, as there were extra questions using similar response categories (eg assessing satisfaction with health/income/etc before asking about satisfaction with "your life overall" | same | wave 10 (2018-2019)+annually before that |
|  |  | 2. Mostly dissatisfied |  |  |  |  |
|  |  | 3. Somewhat dissatisfied |  |  |  |  |
|  |  | 4. Neither satisfied nor dissatisfied |  |  |  |  |
|  |  | 5. Somewhat satisfied |  |  |  |  |
|  |  | 6. Mostly satisfied |  |  |  |  |
|  |  | 7. Completely satisfied |  |  |  |  |
| TwinsUK | Overall, in the past week, how satisfied have you been with your life? | 0 -10 | April 2020 - | Not really, apart from "General Health Questionnaire\ Have you recently been satisfied with the way you've carried out your task?" |  |  |
| GS | On a scale of 0 (not at all) to 10 (extremely), how satisfied are you with your life nowadays? | 0 -10 | April - May 2020; July - August 2020; February 2021 | Thinking back to just before COVID-19 measures were introduced (i.e., January 2020), how satisfied were you with your life then? | same | April - May 2020 |

### 4. Measures of Social Contacts

|  |  | *Covid Surveys* | | | *Pre-Covid Surveys* | | |
| --- | --- | --- | --- | --- | --- | --- | --- |
| **Study** |  | **Questions asked** | **Possible Answer categories/range:** | **When assessed:** | **Questions or Scale** | **Possible Answer categories/range:** | **When assessed:** |
| **ELSA** | Contact family | “In the past month, how often have you done the following with any of your immediate family (parents, children, grandchildren, and brothers and sisters), not counting any who live with you?” | For each activity 1.Speak on the phone; 2.Video-calling; 3.Write or email; 4.Send or receive text messages | June-July 2020 | On average, how often do you do each of the following with any of your children,not counting any who live with you? | For each activity A.Meet up B.Speak on the phone; C.Write or email; D.Send or receive text messages | Wave 9 (2018/19) in self-completion Q |
|  |  |  | 1.Daily; 2.3 to 6 times a week; 3.Once or twice a week; 4.Less than once a week or never | Nov-Dec 2020 | On average, how often do you do each of the following with any family members, not counting any who live with you? | 1.+3 times/week; 2.Once/Twice week; 3.Once or twice a month; 4.Every few months; 5.Once or Twice a year; 6.Less than once a year or never |  |
| **ELSA** | Contact friends | “In the past month, how often have you done the following with other relatives and/or friends?” | For each activity 1.Speak on the phone; 2.Video-calling; 3.Write or email; 4.Send or receive text messages | June-July 2020; | On average, how often do you do each of the following with any of your friends, not counting any who live with you? | For each activity A.Meet up B.Speak on the phone; C.Write or email; D.Send or receive text messages | Wave 9 (2018/19) in self-completion Q |
|  |  |  | 1.Daily; 2.3 to 6 times a week; 3.Once or twice a week; 4.Less than once a week or never | Nov-Dec 2020 |  | 1.+3 times/week; 2.Once/Twice week; 3.Once or twice a month; 4.Every few months; 5.Once or Twice a year; 6.Less than once a year or never |  |
| **NCDS, BCS70, NS, & MCS** | In-person contact | "In the last seven days, on how many days did you meet up in person with any of your family or friends who do not live with you?" | 0=Never; 1= 1 day; 2 = 2-3 days; 3 = 4-6 days; 4 = Every day | April-May 2020 | "How often have you met friends (& met family)?" ( NS 2015; BCS70 2016; NCDS 2013), "Frequency spending time with friends" (MCS 2018). |  |  |
| **NCDS, BCS70, NS, & MCS** | Digital contact | "In the last seven days, on how many days did you talk to family or friends you do not live with via phone or video calls?" ... "how many days did you keep in contact with family or friends you do not live with by email or text or other electronic messaging?" | 0=Never; 1= 1 day; 2 = 2-3 days; 3 = 4-6 days; 4 = Every day | April-May 2020 | no |  |  |
| USOC |  | In the last 4 weeks, how often have you met in person with friends and family who do not live with you? | 1. Daily | June & Nov 2020 | Thinking back to earlier this year, before the outbreak of the coronavirus pandemic. In | same | Jan/Feb 2020 (Asked during June survey) |
|  |  |  | 2. Several times per week |  | January/February 2020, how often did you meet in person with friends and family who do |  |  |
|  |  |  | 3. At least once per week |  | not live with you? |  |  |
|  |  |  | 4. Several times per month |  |  |  |  |
|  |  |  | 5. At least once per month |  |  |  |  |
|  |  |  | 6. Less often |  |  |  |  |
|  |  |  | 7. Never |  |  |  |  |
| USOC |  | In the last 4 weeks, how often have you spoken to friends or family who do not live with you on the phone or in a video call (e.g. Facetime, Zoom, Skype)? | 1. Daily | June & Nov 2020 | Still thinking about January/February 2020, how often did you speak to friends or family who do not live with you on the phone or in a video call (e.g. Facetime, Zoom, Skype)? | same | Jan/Feb 2020 (Asked during June survey) |
|  |  |  | 2. Several times per week |  |  |  |  |
|  |  |  | 3. At least once per week |  |  |  |  |
|  |  |  | 4. Several times per month |  |  |  |  |
|  |  |  | 5. At least once per month |  |  |  |  |
|  |  |  | 6. Less often |  |  |  |  |
|  |  |  | 7. Never |  |  |  |  |
| USOC |  | In the last 4 weeks, how often have you engaged in text chats with friends or family who do not live with you, for example using text messaging, Instagram, Facebook, or WhatsApp? | 1. Daily | June & Nov 2020 | Still thinking about January/February 2020, how often did you engage in text chats with friends or family who do not live with you, for example using text messaging, Instagram, Facebook, or WhatsApp? | same | Jan/Feb 2020 (Asked during June survey) |
|  |  |  | 2. Several times per week |  |  |  |  |
|  |  |  | 3. At least once per week |  |  |  |  |
|  |  |  | 4. Several times per month |  |  |  |  |
|  |  |  | 5. At least once per month |  |  |  |  |
|  |  |  | 6. Less often |  |  |  |  |
|  |  |  | 7. Never |  |  |  |  |
| ALSPAC G1 |  | How many people, apart from those you live with, did | Face to face (in person): 0-4, 5-17, 18-29, 30-39, 40-49, 50-59, 60-69, 70+; | Jun-20 |  |  |  |
|  |  | you speak to yesterday in the following ways (for | Over the phone (talking but no video image): 0-4, 5-17, 18-29, 30-39, 40-49, 50-59, 60-69, 70+; |  |  |  |  |
|  |  | personal and for work reasons) from each of the | Via video media (e.g. Skype, Factime; with video images of person you spoke to): 0-4, 5-17, 18-29, 30-39, 40-49, 50-59, 60-69, 70+; |  |  |  |  |
|  |  | following age groups (approximate ages are fine) | With physical contact (e.g. handshake/hug/kiss/personal care etc.): 0-4, 5-17, 18-29, 30-39, 40-49, 50-59, 60-69, 70+; |  |  |  |  |
| ALSPAC G0 |  | How many people, apart from those you live with, did | Face to face (in person): 0-4, 5-17, 18-29, 30-39, 40-49, 50-59, 60-69, 70+; | Jun-20 |  |  |  |
|  |  | you speak to yesterday in the following ways (for | Over the phone (talking but no video image): 0-4, 5-17, 18-29, 30-39, 40-49, 50-59, 60-69, 70+; |  |  |  |  |
|  |  | personal and for work reasons) from each of the | Via video media (e.g. Skype, Factime; with video images of person you spoke to): 0-4, 5-17, 18-29, 30-39, 40-49, 50-59, 60-69, 70+; |  |  |  |  |
|  |  | following age groups (approximate ages are fine) | With physical contact (e.g. handshake/hug/kiss/personal care etc.): 0-4, 5-17, 18-29, 30-39, 40-49, 50-59, 60-69, 70+; |  |  |  |  |
| TwinsUK |  | In the past 7 days, how many days have you had face-to-face contact with another person for 15 minutes or more (including someone you live with)? | scale 1 -7 | April 2020 - | Is this more, less or about the same as before this period of self-isolation related to COVID-19 began? | More, about the same, less |  |
| TwinsUK |  | In the past 7 days, how many days have you had a phone or video call with another person for 15 minutes or more? | scale 1 -8 | April 2020 - | Is this more, less or about the same as before this period of self-isolation related to COVID-19 began? | More, about the same, less |  |
| TwinsUK |  | On a scale of 1-5, are you interacting with people in any form (e.g. in person, over the phone, via voice chat etc.) more or less due to COVID-19 isolation? | 1 Many less people than usual, 2- A few less people than usual, 3- No change, 4- A few more people than usual, 5- Many more people than usual | April 2020 - |  |  |  |
|  |  | The way you interact might've changed e.g. more telephone than face-to-face, but we are interested only in the frequency of contact. |  |  |  |  |  |
| TwinsUK |  | In the past week, have any of the following aspects of your life changed? Time spent talking to family/friends via phone or technology | Decreased, stayed the same, increased, NA/I don't do this | April 2020 - |  |  |  |
|  |  | Time spent digitally socialising (e.g. group chats, watching movies in groups online) |  |  |  |  |  |
|  |  | Time spent talking to work colleagues |  |  |  |  |  |
| TwinsUK |  | In the last week, how frequently have you been in contact with your twin (this can be in any form, e.g. via email, phone text message etc.) |  | April 2020 - |  |  |  |
|  |  |  | More than once a day |  |  |  |  |
|  |  |  | Once a day |  |  |  |  |
|  |  |  | Several times a week |  |  |  |  |
|  |  |  | Once or twice a week |  |  |  |  |
|  |  |  | Not at all |  |  |  |  |
|  |  |  | Not applicable |  |  |  |  |
| TwinsUK |  |  |  | April 2020 - | Every day | Every day |  |
|  |  |  |  |  |  | Three or more times a week |  |
|  |  |  |  |  |  | Once or twice a week |  |
|  |  |  |  |  |  | Once or twice a month |  |
|  |  |  |  |  |  | Less than once a month |  |
|  |  |  |  |  | Three or more times a week |  |  |
|  |  |  |  |  | Once or twice a week |  |  |
|  |  |  |  |  | Once or twice a month |  |  |
|  |  |  |  |  | Less than once a month |  |  |
| GS |  | Now that COVID-19 measures are in place, how regularly do you do these activities now | 1. Never | Apri - May 2020 | Just before the COVID-19 measures were introduced (i.e., January 2020), how regularly did you:- Meet with family members face-to-face | Same | April - May 2020 |
|  |  | - Meet with family members face-to-face | 2. Rarely |  | - Meet with friends face-to-face |  |  |
|  |  | - Meet with friends face-to-face | 3. Less than once a week |  | - Call family members |  |  |
|  |  | - Call family members | 4. 1-2 days a week |  | - Call friends |  |  |
|  |  | - Call friends | 5. 3-4 days a week |  | - Video call with family members (e.g., Skype, Facetime) |  |  |
|  |  | - Video call with family members (e.g., Skype, Facetime) | 6. 3-4 days a week |  | - Video call with friends (e.g., Skype, Facetime) |  |  |
|  |  | - Video call with friends (e.g., Skype, Facetime) | 7. Every day/almost every day |  | - Text or instant message (e.g., WhatsApp, Facebook Messenger) with family members |  |  |
|  |  | - Text or instant message (e.g., WhatsApp, Facebook Messenger) with family members |  |  | - Text or instant message (e.g., WhatsApp, Facebook Messenger) with friends |  |  |
|  |  | - Text or instant message (e.g., WhatsApp, Facebook Messenger) with friends |  |  |  |  |  |
| GS |  | How regularly do you do these activities now? | As above | July - August 2020; February 2021 | |  |  |
|  |  | - Meet with family members face-to-face |  |  |  |  |  |
|  |  | - Meet with friends face-to-face |  |  |  |  |  |
|  |  | - Call family members |  |  |  |  |  |
|  |  | - Call friends |  |  |  |  |  |
|  |  | - Video call with family members (e.g., Skype, Facetime) |  |  |  |  |  |
|  |  | - Video call with friends (e.g., Skype, Facetime) |  |  |  |  |  |
|  |  | - Text or instant message (e.g., WhatsApp, Facebook Messenger) with family members |  |  |  |  |  |
|  |  | - Text or instant message (e.g., WhatsApp, Facebook Messenger) with friends |  |  |  |  |  |

### 5. Measures of Self-Assessed Health

|  | *Covid Surveys* | | | *Pre-Covid Surveys* | | |
| --- | --- | --- | --- | --- | --- | --- |
| **Study** |  | **Possible Answer categories/range:** | **When assessed:** |  | **Possible Answer categories/range:** | **When assessed (if retrospective, say when answers refer to):** |
| **NCDS** | In general, would you say your health is... | 1.Excellent | March-May 2020; | In general, would you say your health is... | 1.Excellent | (MCS: 2018/2019) (NS: 2015) (BCS70: 2016);(NCDS:2013) |
| **BCS70** |  | 2.Very good | Aug-Sept 2020 |  | 2.Very good |  |
| **NS** |  | 3.Good |  |  | 3.Good |  |
| **MCS** |  | 4.Fair |  |  | 4.Fair |  |
|  |  | 5.Poor |  |  | 5.Poor |  |
| ELSA | In the past month would you say your health was…? | 1.Excellent | June-July 2020; | Would you say your health is... | 1.Excellent | 2018/19 |
|  |  | 2.Very good | Nov-Dec 2020 |  | 2.Very good |  |
|  |  | 3.Good |  |  | 3.Good |  |
|  |  | 4.Fair |  |  | 4.Fair |  |
|  |  | 5.Poor |  |  | 5.Poor |  |
| GS | In general, would you say your health is... | 1.Excellent | April-June 2020 |  |  |  |
|  |  | 2.Very good |  |  |  |  |
|  |  | 3.Good |  |  |  |  |
|  |  | 4.Fair |  |  |  |  |
|  |  | 5.Poor |  |  |  |  |
| ALSPAC | PRISMA-7;In general, do you have health problems that require you to limit your activities? | Yes/No to each question (can sum to 0-5 for PRISMA-7 analog) | April-May 2020 | Previous adjustment for pre-pandemic SRH was derived as "prepandemic history obese/diabetic/asthmatic" - can derive something similar for this, but nothing bespoke comes to mind! |  |  |
|  | Do you need someone to help you on a regular basis? |  |  |  |  |  |
|  | In general, do you have any health problems that require you to stay at home? |  |  |  |  |  |
|  | If you need help, can you count on someone close to you? |  |  |  |  |  |
|  | Do you regularly use a stick, walker or wheelchair to move about? |  |  |  |  |  |
| USOC | In general, would you say your health is... | 1.Excellent | Nov 2020-Jan 2021 | same | same | wave 10 (2018-2019)+annually before that |
|  |  | 2.Very good |  |  |  |  |
|  |  | 3.Good |  |  |  |  |
|  |  | 4.Fair |  |  |  |  |
|  |  | 5.Poor |  |  |  |  |
| TwinsUK | PRIMSA-7: In general, did you have any health problems that require you to limit your activities? | Yes/No | April 2020 - ? | same | same | Retrospective ('Before COVID-19 pandemic') but also have asked previously in waves since ~2015 |
|  | Did you need someone to help you on a regular basis? | Yes/No |  |  |  |  |
|  | In general, did you have any health problems that require you to stay at home? | Yes/No |  |  |  |  |
|  | If you needed, could you count on someone close to you? | Yes/No |  |  |  |  |
|  | Did you regularly use a stick, walker or wheelchair to move about? | Yes/No |  |  |  |  |
| TwinsUK | **In general would you say your health was:** | 1.Excellent | April 2020 - ? | same | same | Retrospective ('Before COVID-19 pandemic') but also have asked previously in waves since ~2015 |
|  |  | 2.Very good |  |  |  |  |
|  |  | 3.Good |  |  |  |  |
|  |  | 4.Fair |  |  |  |  |
|  |  | 5.Poor |  |  |  |  |
| GS | In general, would you say your health is... | 1.Excellent | April - May 2020; February 2021 | None |  |  |
|  |  | 2.Very good |  |  |  |  |
|  |  | 3.Good |  |  |  |  |
|  |  | 4.Fair |  |  |  |  |
|  |  | 5.Poor |  |  |  |  |

### 6. Measures of employment

| ***Study*** | ***Question (exact wording)*** | **Possible Answers** | **Recoding** |
| --- | --- | --- | --- |
| **CHANGE in EMPLOYMENT STATUS** | |  |  |
|  | **1='Stable employed'; 2='Furloughed'; 3='Became employed'; 4='No longer employed'; 5='Stable unemployed'; 6='Stable in other category'** | | |
| **MCS** | Q1: Which of these best describes what you were doing just before the Coronavirus outbreak in March? If you were doing more than one activity, please choose the activity that you spent most time doing. | Q1. 1=Employed; 2=Self-employed; 3=In unpaid/ voluntary work; 4=Apprenticeship; 5=Unemployed; 6=Permanently sick or disabled; 7=Looking after home or family; 8=In education at school/college/university; 9=Retired; 10=Doing something else. | 1= if (Q1 = 1) & (Q2 = 1 OR 6) |
| **NS** | Q2: Which of these would you say best describes your situation now? | Q2. 1=Employed and currently working (or on annual leave/holiday); 2=Employed but on paid leave (including furlough); 3=Employed and on unpaid leave; 4=Apprenticeship; 5=In unpaid/voluntary work; 6=Self-employed and currently working (or on holiday); 7=Self-employed but not currently working; 8=Unemployed; 9=Permanently sick or disabled; 10=Looking after home or family; 11=In education at school/college/university; 12=Retired; 13=Doing something else. | 2= if (Q1 = 1) & (Q2 = 2 OR 7) |
| **BCS 70** |  | Q3. 1=There was no interruption to learning activities 2. I took a break from learning activities 3. I was studying at home with online resources provided by my learning establishment 4. I was studying at home with no online resources provided by my learning establishment 5. My course finished earlier than planned 6. I dropped out from learning activities | 3= if (Q1 != 1 OR 5) & (Q2 = (1 OR 6) |
| **NCDS** |  |  | 4= if (Q1 = 1) & (Q2 != 1 OR 6) |
|  |  |  | 5= if (Q1 = 5) & (Q2 = 8) |
|  |  |  | 6= if (Q1 = != 1 OR 5) & (Q2 != 1 OR 2 OR 6 OR 7 OR 8) |
| **ALSPAC** | Q1. Just before the lockdown on the 23rd March 2020, were you? | Q1. 1=In full time paid work (30 or more hours a week); 2=In part-time paid work (less than 30 | 1=if(Q1=1/3) & (Q2= 1/3 OR 8) |
|  | Q2. Which of these would you say best describes your current situation now? | hours a week); 3=In irregular or occasional work; 4=Doing a modern apprenticeship or other government supported training/work-experience scheme; 5=Unemployed and looking for work; 6=Unable to work through sickness/disability; 7=In full-time education; 8=In part-time education; 9=Doing voluntary work; 10=Self-employed; 11=A full/part time carer; 12=Retired; 13=Other. | 2=if(Q1=1/3) & (Q2=4) |
|  |  | Q2. 1=Employed and working same number of hours as pre-lockdown; 2=Employed and working | 3=if(Q1!=1/3) & (Q2 = 1/3 OR 8) |
|  |  | reduced number of hours; 3=Employed and working more hours than before; 4=Employed but on paid leave (including furlough); 5=Employed and on unpaid leave; 6=Apprenticeship; 7=In unpaid/voluntary work; 8=Self-employed and currently working; 9=self-employed but not currently working; 10=Unemployed; 11=Permanently Sick/Disabled; 12=looking after home or family;13=In education at school/college/university | 4=if(Q1=1/3) & (Q2 = 5/7 OR 9/13) |
|  |  |  | 5=if(Q1=5) & (Q2 = 10) |
|  |  |  | 6=if(Q1!=1/3) & (Q2 = 5/7 OR 9/13) |
| **USOC** | Q1: Thinking back to earlier this year, before the outbreak of the coronavirus pandemic. Were you in paid work or self-employment at any time in January or February 2020? | Q1: 1. Yes, employed; 2. Yes, self-employed; 3. Yes, both employed and self-employed; 4. No. | 1=if (Q1<=3) & (Q2<=3) |
|  | Q2: Thinking about your situation now. Even if you did not do any paid work last week, are you currently employed or self-employed? | Q2: 1. Yes, employed; 2. Yes, self-employed; 3. Yes, both employed and self-employed; 4. No. | 2=if (Q1<=3) & Q3=1 |
|  | Q3: Have you received a written letter or email from your employer to confirm that you have been furloughed under the Coronavirus Job Retention Scheme? | Q3: 1. Yes; 2. No. | 3=if (Q1=4) & (Q2<=3) |
|  | Q4: (asked pre-pandemic) Which of these best describes your current employment situation? | Q4: 1. Self employed; 2. Paid employment; 3. Unemployed; 4. Retired; 5. On maternity leave; 6. Family care or home; 7. Full-time student; 8. LT sick or disabled; 9. Govt training scheme; 10. Unpaid, family business; 11. On apprenticeship. 97. Doing something else. | 4=if (Q1<=3) & (Q2=4) |
|  |  |  | 5=if (Q1=4) & (Q2=4) & (Q4=3) |
|  |  |  | 6=if (Q1=4) & (Q2=4) & (Q4!=3) |
| **ELSA** | Q1: Which of these best describes what you were doing just before the coronavirus outbreak? | Q1. 1=Retired; 2=Employed; 3=Self-employed; 4=Unemployed; 5=Permanently sick or disabled; 6=Looking after home or family | 1= if (Q1=2 OR 3) & (Q2=2 OR 4) |
|  | Q2: And which of these would you say best describes your current situation? | Q2. 1=Retired; 2=Employed; 3=Paid/unpaid leave from employment (including furlough); 4=Self-employed and currently working; 5=Self-employed but not currently working; 6=Unemployed; 7=Permanently sick or disabled; 8=Looking after home or family. | 2= if (Q1=1 OR 2 OR 3) & (Q2=3 OR 5) |
|  |  |  | 3= if (Q1=1 OR >=4) & (Q2=2 OR =4) |
|  |  |  | 4= if (Q1=2 OR 3) & (Q2>=6 OR Q2==1) |
|  |  |  | 5= if Q1=4 & Q2=6 |
|  |  |  | 6= if (Q1=1 & Q2=1) OR (Q1>=4 & (Q2=1 OR 3 OR 5 OR 7 OR 8)) OR (Q1=1 & Q2>=7) |
| **GS** | Q1.What was your employment status just before the COVID-19 measures were introduced (i.e. January 2020)? | Q1 and Q2. 1=Self-employed employing others; 2=Self-employed not employing others; 3=Paid employee supervising others; 4=Paid employee not supervising others; 5=In unpaid employment; 6=Homemaker; 7=Looking after children; 8=Looking after other dependents; 9=Retired; 10=Still in school/studying full-time; 11=Unemployed as sick or disabled; 12=Unemployed; 13=Other | 1= if Q1<=4 & Q2<=4 |
|  | Q2. What is your employment status now? | Q3. 1=Yes; 0=NA | 2= if Q3=1 |
|  | Q3. Have any of the following happened to you due to COVID-19 measures? |  | 3= if Q1>=5 & Q2<=4 |
|  |  |  | 4= if Q1<=4 & Q2 >= 5 |
|  |  |  | 5= if Q1=12 & Q2=12 |
|  |  |  | 6= if (Q1>=5 & Q1<=11\|Q1==13) & ((Q2>=5 & Q2<=11)\|Q2==13) |

### 7. Covariates

| ***Variables*** | ***Study*** | ***Options*** | ***Recoding if needed*** |
| --- | --- | --- | --- |
| *** Sex * 0=Male; 1=Female** |  |  |  |
|  | **All** | 0=Male; 1=Female |  |
| *** Ethnicity * 0=White; 1=Ethnic Minority** | | | |
|  | **MCS** | 1=White; 2=Mixed; 3=Indian; 4=Pakistani; 5=Bangladeshi; 6=Other Asian; 7=Black Caribbean; 8=Black African; 9=Other Black; 10=Chinese; 11=Other ethnic group | 1=0; 2/11=1 |
|  | **NS** | 1=White; 2=Mixed; 3=Indian; 4=Pakistani; 5=Bangladeshi; 6=Black Caribbean; 7=Black African; 8=Other | 1=0; 2/8=1 |
|  | **BCS70** | Not Available |  |
|  | **NCDS** | Not Available |  |
|  | **ALSPAC** | G0 (Parents) 1=White; 2=Black carribean; 3=Black african; 4=Other black; 5=Indian; 6=Pakistani; 7=Bangladeshi; 8=Chinese; 9=Other | 1=0; 2/9=1 |
|  |  | G1 (Children) 1=White; 2=Mixed/Multiple Ethnic group; 3=Asian; 4=Black/African/Caribbean/Black British; 5=Arab or Other |  |
|  | **USOC** | 1=White British; 2=Irish (White); 3=Gypsy or Irish Traveller (white); 4=Any other white background; 5=White and black caribbean (mixed); 6=White and black african (mixed); 7=White and Asian (mixed); 8=Any other mixed background; 9=Indian (Asian or Asian British); 10=Pakistani (Asian or Asian British); 11=Bangladeshi (Asian or Asian British); 12=Chinese (Asian or Asian British); 13=Any other Asian background (Asian or Asian British); 14=Caribbean (Black or Black British); 15=African (Black or Black British); 16=Any other Black background (Black or Black British); 17=Arab (other Ethnic group); 97=Any other ethnic group | 1-4=0; 5-97=1 |
|  | **ELSA** | 1.White; 2=Mixed ethnic group; 3=Black; 4=Black British; 5=Asian; 6=Asian British | 1=0; 2/6=1 |
|  | **GS** | 1=White Scottish; 2=White English; 3=White Welsh; 4=White N. Irish; 5=White Irish; 6=White Gypsy/Irish traveller; 7=White Polish; 8=Any other white; 9=Asian/British Asian - Indian; 10=Asian/British Asian - Pakistani; 11=Asian/British Asian - Bangladeshi; 12=Asian/British Asian - Chinese; 13=Any other Asian background; 14=Black or Black British - African; 15=Black or Black British - Carribean; 16=Any other Black/African/Caribbean background; 17=Arab or Arab British; 18=Mixed - White and Black Caribbean; 19=Mixed - White and Black African; 20=Mixed - White and Asian; 21=Any other Mixed/Multiple ethnic background; 22=Any other ethnic group | 1-8=0; 9-22=1 |
| *** Education * 0=No Degree; 1=Degree** | | | |
|  | **MCS** | 0=None; 1=Nvq1; 2=Nvq2; 3=Nvq3; 4=Nvq4; 5=Nvq5 | 1/=0; 4/5=1 |
|  | **NS** | *parent's education for MCS |  |
|  | **BCS 70** |  |  |
|  | **NCDS** |  |  |
|  | **ALSPAC** | 1=Degree; 2=A levels/AS levels or equivalent; 3=O levels; 4=Vocational; 5=CSE | 2/5=0 |
|  |  | *parent's education for G1 (Children) |  |
|  | **USOC** | 1=Degree or equivalent; 2=A-level or equivalent; 3=GSCE or equivalent; 4=No qualifications | 2/4=0 |
|  | **ELSA** | 1=Nvq4/nvq5/degree or equivalent; 2=Higher Education below degree; 3=Nvq3/GCE A level equivalent; 4=Nvq2/GCE O level equivalent; 5=Nvq1/CSE other grade equivalent; 6=Foreign/other; 7=No qualification | 2/7=0 |
|  | **GS** | 1=No qualifications; 2=Other (please specify); 3=School leavers certificate; 4=CSEs or equivalent; 5=Standard grade, National 4 or 5, O levels, GCSEs or equivalent; 6=Higher grade, A levels, AS levels or equivalent; 7=NVQ or HND or HNC or equivalent; 8=Other professional or technical qualification; 9=Undergraduate degree; 10=Postgraduate degree | 1/8=0; 9 OR 10=1 |
| *** Living Arrangement * 1=Alone; 2=With partner/spouse only; 3=With partner/spouse and child(ren); 4=With child(ren), without partner/spouse; 5=Any other living arrangement** | | | |
| **OR * Partnership Status * 1=Married/Partnered; 0=Not married/partnered** | | | |
| *** Pre-Pandemic Self-Assessed Health * 1=Good/Very Good/Excellent; 0=Fair/Poor** | | | |
|  | **MCS** | In general, in the 3 months before the Coronavirus outbreak would you say your health was … 1=Excellent; 2=Very Good; 3=Good; 4=Fair; 5=Poor | 1/3=1; 4/5=0 |
|  | **NS** |  |  |
|  | **BCS 70** |  |  |
|  | **NCDS** |  |  |
|  | **ALSPAC** | Are you or do you currently have any of the following: If yes, please tell us exactly what you have: | If (2, 5 or 10 = TRUE)=1, else =0 |
|  |  | 1=Organ transplant recipient, 2=Diabetes (Type I or II), 3=Heart disease or heart problems, 4=Hypertension (high blood pressure), 5=Overweight, 6=A recent stroke, 7= Kidney disease, 8=Liver disease, 9= Anaemia, 10= Asthma, 11= Other lung condition such as COPD, bronchitis or emphysema, 12=Cancer, 13=Condition affecting the brain and nerves (e.g. Dementia, Parkinson’s, Multiple Sclerosis), 14=A weakened immune system/reduced ability to deal with infections (as a result of a disease or treatment), 15=Depression, 16=Anxiety, 17=Psychiatric disorder |  |
|  | **USOC** | In general, would you say your health is… 1=Excellent; 2=Very Good; 3=Good; 4=Fair; 5=Poor | 1/3=1; 4/5=0 |
|  | **ELSA** | Would you say your health is… 1=Excellent; 2=Very Good; 3=Good; 4=Fair; 5=Poor | 1/3=1; 4/5=0 |
|  | **GS** | Not available |  |
| *** Pre-Pandemic Mental Health * 1=High Psychological Distress; 0=No Physchological Distress** | | | |
|  | **MCS** | ~2 years prior. Kessler K6 measure of psychological distress [Kessler, R. C., Andrews, G., Colpe, L. J., Hiripi, E., Mroczek, D. K., Normand, S. L., ... & Zaslavsky, A. M. (2002). Short screening scales to monitor population prevalences and trends in non-specific psychological distress. Psychological medicine, 32(6), 959-976.] | 13+ = 1 |
|  | **NS** | ~5 years prior. GHQ-12 [Goldberg DP, Gater R, Sartorius N,et al.The validity of two versions of the GHQ in the WHO study of mental illness in general health care. Psychol Med 1997;27:191–7.] | =1 if 4 or more |
|  | **BCS 70** | ~4 years prior. Malaise Inventory. [Rodgers, B., Pickles, A., Power, C., Collishaw, S., & Maughan, B. (1999). Validity of the Malaise Inventory in general population samples. Social psychiatry and psychiatric epidemiology, 34(6), 333-341.] | =1 if 4 or more |
|  | **NCDS** | ~4 years prior. Malaise Inventory. [Rodgers, B., Pickles, A., Power, C., Collishaw, S., & Maughan, B. (1999). Validity of the Malaise Inventory in general population samples. Social psychiatry and psychiatric epidemiology, 34(6), 333-341.] | =1 if 4 or more |
|  | **ALSPAC** | G0 (parents), EPDS [Levis et al. 2020] | =1 if more than 11 |
|  |  | G1 (children), SMFQ [Angold, A., Costello, E.J., Messer, S.C., Pickles, A., Winder, F., & Silver, D. (1995).] |  |
|  | **USOC** | ~1 years prior. GHQ-12 [Goldberg DP, Gater R, Sartorius N,et al.The validity of two versions of the GHQ in the WHO study of mental illness in general health care. Psychol Med 1997;27:191–7.] | =1 if 4+ |
|  | **ELSA** | Eight-item version of the original CES-D (felt depressed; felt everything was an effort; restless sleep; not happy; felt lonely; not enjoyed life; felt sad; could not get going) | =1 if 4 or more symptoms reported |
|  | **GS** | GHQ-28 (score of >24 =1) |  |
