## Supplementary File 3 for "Mental and social wellbeing and the UK Coronavirus Job Retention Scheme: evidence from nine longitudinal studies"

**Supplementary File 3 - Model Estimates**

### 1. Unadjusted

| **Adjustment** | **Outcome** | **Exposure** | **Study** | **Coefficient** | **lower_ci** | **upper_ci** | **%Weight** | **%I2** | **Reason for missing** | **Method** | 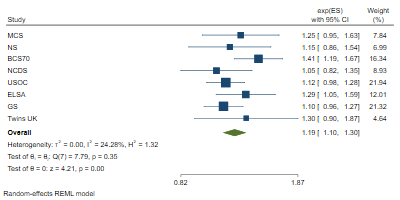 |  |
| --- | --- | --- | --- | --- | --- | --- | --- | --- | --- | --- | --- | --- |
| Unadjusted | low life satisfaction | Furloughed | MCS | 1.25 | 0.95 | 1.63 | 7.84 |  |  | REML |  |  |
| Unadjusted | low life satisfaction | Furloughed | ALSPAC(G1) |  |  |  |  |  | no measure |  |  |  |
| Unadjusted | low life satisfaction | Furloughed | NS | 1.15 | 0.86 | 1.54 | 6.99 |  |  |  |  |  |
| Unadjusted | low life satisfaction | Furloughed | BCS70 | 1.41 | 1.19 | 1.67 | 16.34 |  |  |  |  |  |
| Unadjusted | low life satisfaction | Furloughed | NCDS | 1.05 | 0.82 | 1.35 | 8.93 |  |  |  |  |  |
| Unadjusted | low life satisfaction | Furloughed | USOC | 1.12 | 0.98 | 1.28 | 21.94 |  |  |  |  |  |
| Unadjusted | low life satisfaction | Furloughed | ELSA | 1.29 | 1.05 | 1.59 | 12.01 |  |  |  |  |  |
| Unadjusted | low life satisfaction | Furloughed | GS | 1.10 | 0.96 | 1.27 | 21.32 |  |  |  |  |  |
| Unadjusted | low life satisfaction | Furloughed | ALSPAC(G0) |  |  |  |  |  | no measure |  |  |  |
| Unadjusted | low life satisfaction | Furloughed | Twins UK | 1.30 | 0.90 | 1.87 | 4.64 |  |  |  |  |  |
| Unadjusted | low life satisfaction | Furloughed | Overall | 1.19 | 1.10 | 1.30 |  | 24.28 |  |  |  |  |
|  |  |  |  |  |  |  |  |  |  |  | 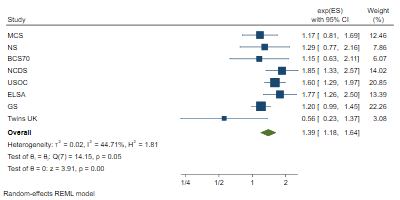 |  |
| Unadjusted | low life satisfaction | No longer employed | MCS | 1.17 | 0.81 | 1.69 | 12.46 |  |  | REML |  |  |
| Unadjusted | low life satisfaction | No longer employed | ALSPAC(G1) |  |  |  |  |  | no measure |  |  |  |
| Unadjusted | low life satisfaction | No longer employed | NS | 1.29 | 0.77 | 2.16 | 7.86 |  |  |  |  |  |
| Unadjusted | low life satisfaction | No longer employed | BCS70 | 1.15 | 0.63 | 2.11 | 6.07 |  |  |  |  |  |
| Unadjusted | low life satisfaction | No longer employed | NCDS | 1.85 | 1.33 | 2.57 | 14.02 |  |  |  |  |  |
| Unadjusted | low life satisfaction | No longer employed | USOC | 1.60 | 1.29 | 1.97 | 20.85 |  |  |  |  |  |
| Unadjusted | low life satisfaction | No longer employed | ELSA | 1.77 | 1.26 | 2.50 | 13.39 |  |  |  |  |  |
| Unadjusted | low life satisfaction | No longer employed | GS | 1.20 | 0.99 | 1.45 | 22.26 |  |  |  |  |  |
| Unadjusted | low life satisfaction | No longer employed | ALSPAC(G0) |  |  |  |  |  | no measure |  |  |  |
| Unadjusted | low life satisfaction | No longer employed | Twins UK | 0.56 | 0.23 | 1.37 | 3.08 |  |  |  |  |  |
| Unadjusted | low life satisfaction | No longer employed | Overall | 1.39 | 1.18 | 1.64 |  | 44.71 |  |  |  |  |
|  |  |  |  |  |  |  |  |  |  |  | 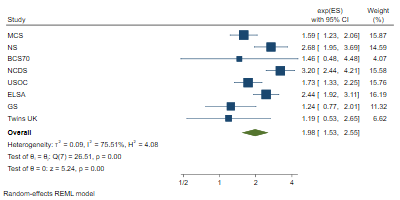 |  |
| Unadjusted | low life satisfaction | Stable unemployed | MCS | 1.59 | 1.23 | 2.06 | 15.87 |  |  | REML |  |  |
| Unadjusted | low life satisfaction | Stable unemployed | ALSPAC(G1) |  |  |  |  |  | no measure |  |  |  |
| Unadjusted | low life satisfaction | Stable unemployed | NS | 2.68 | 1.95 | 3.69 | 14.59 |  |  |  |  |  |
| Unadjusted | low life satisfaction | Stable unemployed | BCS70 | 1.46 | 0.48 | 4.48 | 4.07 |  |  |  |  |  |
| Unadjusted | low life satisfaction | Stable unemployed | NCDS | 3.20 | 2.44 | 4.21 | 15.58 |  |  |  |  |  |
| Unadjusted | low life satisfaction | Stable unemployed | USOC | 1.73 | 1.33 | 2.25 | 15.76 |  |  |  |  |  |
| Unadjusted | low life satisfaction | Stable unemployed | ELSA | 2.44 | 1.92 | 3.11 | 16.19 |  |  |  |  |  |
| Unadjusted | low life satisfaction | Stable unemployed | GS | 1.24 | 0.77 | 2.01 | 11.32 |  |  |  |  |  |
| Unadjusted | low life satisfaction | Stable unemployed | ALSPAC(G0) |  |  |  |  |  | no measure |  |  |  |
| Unadjusted | low life satisfaction | Stable unemployed | Twins UK | 1.19 | 0.53 | 2.65 | 6.62 |  |  |  |  |  |
| Unadjusted | low life satisfaction | Stable unemployed | Overall | 1.98 | 1.53 | 2.55 |  | 75.51 |  |  |  |  |
|  |  |  |  |  |  |  |  |  |  |  | 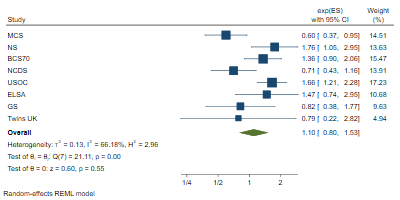 |  |
| Unadjusted | often lonely | Furloughed | MCS | 0.60 | 0.37 | 0.95 | 14.51 |  |  | REML |  |  |
| Unadjusted | often lonely | Furloughed | ALSPAC(G1) |  |  |  |  |  | no measure |  |  |  |
| Unadjusted | often lonely | Furloughed | NS | 1.76 | 1.05 | 2.95 | 13.63 |  |  |  |  |  |
| Unadjusted | often lonely | Furloughed | BCS70 | 1.36 | 0.90 | 2.06 | 15.47 |  |  |  |  |  |
| Unadjusted | often lonely | Furloughed | NCDS | 0.71 | 0.43 | 1.16 | 13.91 |  |  |  |  |  |
| Unadjusted | often lonely | Furloughed | USOC | 1.66 | 1.21 | 2.28 | 17.23 |  |  |  |  |  |
| Unadjusted | often lonely | Furloughed | ELSA | 1.47 | 0.74 | 2.95 | 10.68 |  |  |  |  |  |
| Unadjusted | often lonely | Furloughed | GS | 0.82 | 0.38 | 1.77 | 9.63 |  |  |  |  |  |
| Unadjusted | often lonely | Furloughed | ALSPAC(G0) |  |  |  |  |  | no measure |  |  |  |
| Unadjusted | often lonely | Furloughed | Twins UK | 0.79 | 0.22 | 2.82 | 4.94 |  |  |  |  |  |
| Unadjusted | often lonely | Furloughed | Overall | 1.10 | 0.80 | 1.53 |  | 66.18 |  |  |  |  |
|  |  |  |  |  |  |  |  |  |  |  | 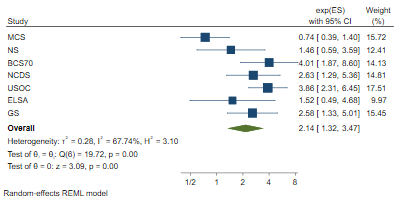 |  |
| Unadjusted | often lonely | No longer employed | MCS | 0.74 | 0.39 | 1.40 | 15.72 |  |  | REML |  |  |
| Unadjusted | often lonely | No longer employed | ALSPAC(G1) |  |  |  |  |  | no measure |  |  |  |
| Unadjusted | often lonely | No longer employed | NS | 1.46 | 0.59 | 3.59 | 12.41 |  |  |  |  |  |
| Unadjusted | often lonely | No longer employed | BCS70 | 4.01 | 1.87 | 8.60 | 14.13 |  |  |  |  |  |
| Unadjusted | often lonely | No longer employed | NCDS | 2.63 | 1.29 | 5.36 | 14.81 |  |  |  |  |  |
| Unadjusted | often lonely | No longer employed | USOC | 3.86 | 2.31 | 6.45 | 17.51 |  |  |  |  |  |
| Unadjusted | often lonely | No longer employed | ELSA | 1.52 | 0.49 | 4.68 | 9.97 |  |  |  |  |  |
| Unadjusted | often lonely | No longer employed | GS | 2.58 | 1.33 | 5.01 | 15.45 |  |  |  |  |  |
| Unadjusted | often lonely | No longer employed | ALSPAC(G0) |  |  |  |  |  | no measure |  |  |  |
| Unadjusted | often lonely | No longer employed | Twins UK |  |  |  |  |  | low counts |  |  |  |
| Unadjusted | often lonely | No longer employed | Overall | 2.14 | 1.32 | 3.47 |  | 67.74 |  |  |  |  |
|  |  |  |  |  |  |  |  |  |  |  | 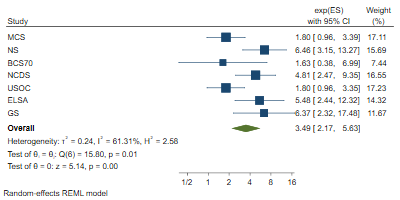 |  |
| Unadjusted | often lonely | Stable unemployed | MCS | 1.80 | 0.96 | 3.39 | 17.11 |  |  | REML |  |  |
| Unadjusted | often lonely | Stable unemployed | ALSPAC(G1) |  |  |  |  |  | no measure |  |  |  |
| Unadjusted | often lonely | Stable unemployed | NS | 6.46 | 3.15 | 13.27 | 15.69 |  |  |  |  |  |
| Unadjusted | often lonely | Stable unemployed | BCS70 | 1.63 | 0.38 | 6.99 | 7.44 |  |  |  |  |  |
| Unadjusted | often lonely | Stable unemployed | NCDS | 4.81 | 2.47 | 9.35 | 16.55 |  |  |  |  |  |
| Unadjusted | often lonely | Stable unemployed | USOC | 1.80 | 0.96 | 3.35 | 17.23 |  |  |  |  |  |
| Unadjusted | often lonely | Stable unemployed | ELSA | 5.48 | 2.44 | 12.32 | 14.32 |  |  |  |  |  |
| Unadjusted | often lonely | Stable unemployed | GS | 6.37 | 2.32 | 17.48 | 11.67 |  |  |  |  |  |
| Unadjusted | often lonely | Stable unemployed | ALSPAC(G0) |  |  |  |  |  | no measure |  |  |  |
| Unadjusted | often lonely | Stable unemployed | Twins UK |  |  |  |  |  | low counts |  |  |  |
| Unadjusted | often lonely | Stable unemployed | Overall | 3.49 | 2.17 | 5.63 |  | 61.31 |  |  |  |  |
|  |  |  |  |  |  |  |  |  |  |  | 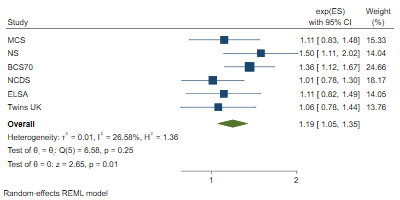 |  |
| Unadjusted | high loneliness | Furloughed | MCS | 1.11 | 0.83 | 1.48 | 15.33 |  |  | REML |  |  |
| Unadjusted | high loneliness | Furloughed | ALSPAC(G1) |  |  |  |  |  | no measure |  |  |  |
| Unadjusted | high loneliness | Furloughed | NS | 1.50 | 1.11 | 2.02 | 14.04 |  |  |  |  |  |
| Unadjusted | high loneliness | Furloughed | BCS70 | 1.36 | 1.12 | 1.67 | 24.66 |  |  |  |  |  |
| Unadjusted | high loneliness | Furloughed | NCDS | 1.01 | 0.78 | 1.30 | 18.17 |  |  |  |  |  |
| Unadjusted | high loneliness | Furloughed | USOC |  |  |  |  |  | no measure |  |  |  |
| Unadjusted | high loneliness | Furloughed | ELSA | 1.11 | 0.82 | 1.49 | 14.05 |  |  |  |  |  |
| Unadjusted | high loneliness | Furloughed | GS |  |  |  |  |  | no measure |  |  |  |
| Unadjusted | high loneliness | Furloughed | ALSPAC(G0) |  |  |  |  |  | no measure |  |  |  |
| Unadjusted | high loneliness | Furloughed | Twins UK | 1.06 | 0.78 | 1.44 | 13.76 |  |  |  |  |  |
| Unadjusted | high loneliness | Furloughed | Overall | 1.19 | 1.05 | 1.35 |  | 26.58 |  |  |  |  |
|  |  |  |  |  |  |  |  |  |  |  | 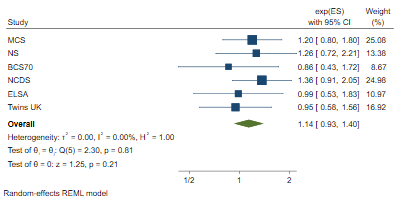 |  |
| Unadjusted | high loneliness | No longer employed | MCS | 1.20 | 0.80 | 1.80 | 25.08 |  |  | REML |  |  |
| Unadjusted | high loneliness | No longer employed | ALSPAC(G1) |  |  |  |  |  | no measure |  |  |  |
| Unadjusted | high loneliness | No longer employed | NS | 1.26 | 0.72 | 2.21 | 13.38 |  |  |  |  |  |
| Unadjusted | high loneliness | No longer employed | BCS70 | 0.86 | 0.43 | 1.72 | 8.67 |  |  |  |  |  |
| Unadjusted | high loneliness | No longer employed | NCDS | 1.36 | 0.91 | 2.05 | 24.98 |  |  |  |  |  |
| Unadjusted | high loneliness | No longer employed | USOC |  |  |  |  |  | no measure |  |  |  |
| Unadjusted | high loneliness | No longer employed | ELSA | 0.99 | 0.53 | 1.83 | 10.97 |  |  |  |  |  |
| Unadjusted | high loneliness | No longer employed | GS |  |  |  |  |  | no measure |  |  |  |
| Unadjusted | high loneliness | No longer employed | ALSPAC(G0) |  |  |  |  |  | no measure |  |  |  |
| Unadjusted | high loneliness | No longer employed | Twins UK | 0.95 | 0.58 | 1.56 | 16.92 |  |  |  |  |  |
| Unadjusted | high loneliness | No longer employed | Overall | 1.14 | 0.93 | 1.40 |  | 0.00 |  |  |  |  |
|  |  |  |  |  |  |  |  |  |  |  | 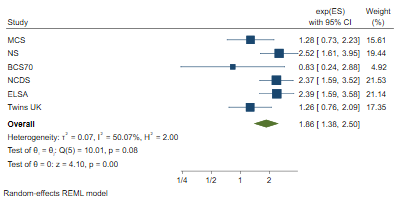 |  |
| Unadjusted | high loneliness | Stable unemployed | MCS | 1.28 | 0.73 | 2.23 | 15.61 |  |  | REML |  |  |
| Unadjusted | high loneliness | Stable unemployed | ALSPAC(G1) |  |  |  |  |  | no measure |  |  |  |
| Unadjusted | high loneliness | Stable unemployed | NS | 2.52 | 1.61 | 3.95 | 19.44 |  |  |  |  |  |
| Unadjusted | high loneliness | Stable unemployed | BCS70 | 0.83 | 0.24 | 2.88 | 4.92 |  |  |  |  |  |
| Unadjusted | high loneliness | Stable unemployed | NCDS | 2.37 | 1.59 | 3.52 | 21.53 |  |  |  |  |  |
| Unadjusted | high loneliness | Stable unemployed | USOC |  |  |  |  |  | no measure |  |  |  |
| Unadjusted | high loneliness | Stable unemployed | ELSA | 2.39 | 1.59 | 3.58 | 21.14 |  |  |  |  |  |
| Unadjusted | high loneliness | Stable unemployed | GS |  |  |  |  |  | no measure |  |  |  |
| Unadjusted | high loneliness | Stable unemployed | ALSPAC(G0) |  |  |  |  |  | no measure |  |  |  |
| Unadjusted | high loneliness | Stable unemployed | Twins UK | 1.26 | 0.76 | 2.09 | 17.35 |  |  |  |  |  |
| Unadjusted | high loneliness | Stable unemployed | Overall | 1.86 | 1.38 | 2.50 |  | 50.07 |  |  |  |  |
|  |  |  |  |  |  |  |  |  |  |  | 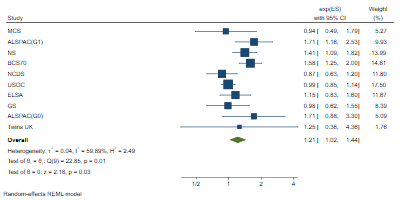 |  |
| Unadjusted | distressed (bin.) | Furloughed | MCS | 0.94 | 0.49 | 1.79 | 5.27 |  |  | REML |  |  |
| Unadjusted | distressed (bin.) | Furloughed | ALSPAC(G1) | 1.71 | 1.16 | 2.53 | 9.93 |  |  |  |  |  |
| Unadjusted | distressed (bin.) | Furloughed | NS | 1.41 | 1.09 | 1.82 | 13.99 |  |  |  |  |  |
| Unadjusted | distressed (bin.) | Furloughed | BCS70 | 1.58 | 1.25 | 2.00 | 14.61 |  |  |  |  |  |
| Unadjusted | distressed (bin.) | Furloughed | NCDS | 0.87 | 0.63 | 1.20 | 11.80 |  |  |  |  |  |
| Unadjusted | distressed (bin.) | Furloughed | USOC | 0.99 | 0.85 | 1.14 | 17.50 |  |  |  |  |  |
| Unadjusted | distressed (bin.) | Furloughed | ELSA | 1.15 | 0.83 | 1.60 | 11.67 |  |  |  |  |  |
| Unadjusted | distressed (bin.) | Furloughed | GS | 0.98 | 0.62 | 1.55 | 8.39 |  |  |  |  |  |
| Unadjusted | distressed (bin.) | Furloughed | ALSPAC(G0) | 1.71 | 0.88 | 3.30 | 5.09 |  |  |  |  |  |
| Unadjusted | distressed (bin.) | Furloughed | Twins UK | 1.25 | 0.36 | 4.36 | 1.76 |  |  |  |  |  |
| Unadjusted | distressed (bin.) | Furloughed | Overall | 1.21 | 1.02 | 1.44 |  | 59.89 |  |  |  |  |
|  |  |  |  |  |  |  |  |  |  |  | 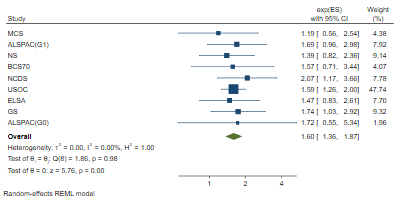 |  |
| Unadjusted | distressed (bin.) | No longer employed | MCS | 1.19 | 0.56 | 2.54 | 4.38 |  |  | REML |  |  |
| Unadjusted | distressed (bin.) | No longer employed | ALSPAC(G1) | 1.69 | 0.96 | 2.98 | 7.92 |  |  |  |  |  |
| Unadjusted | distressed (bin.) | No longer employed | NS | 1.39 | 0.82 | 2.36 | 9.14 |  |  |  |  |  |
| Unadjusted | distressed (bin.) | No longer employed | BCS70 | 1.57 | 0.71 | 3.44 | 4.07 |  |  |  |  |  |
| Unadjusted | distressed (bin.) | No longer employed | NCDS | 2.07 | 1.17 | 3.66 | 7.78 |  |  |  |  |  |
| Unadjusted | distressed (bin.) | No longer employed | USOC | 1.59 | 1.26 | 2.00 | 47.74 |  |  |  |  |  |
| Unadjusted | distressed (bin.) | No longer employed | ELSA | 1.47 | 0.83 | 2.61 | 7.70 |  |  |  |  |  |
| Unadjusted | distressed (bin.) | No longer employed | GS | 1.74 | 1.03 | 2.92 | 9.32 |  |  |  |  |  |
| Unadjusted | distressed (bin.) | No longer employed | ALSPAC(G0) | 1.72 | 0.55 | 5.34 | 1.96 |  |  |  |  |  |
| Unadjusted | distressed (bin.) | No longer employed | Twins UK |  |  |  |  |  | low counts |  |  |  |
| Unadjusted | distressed (bin.) | No longer employed | Overall | 1.60 | 1.36 | 1.87 |  | 0.00 |  |  |  |  |
|  |  |  |  |  |  |  |  |  |  |  | 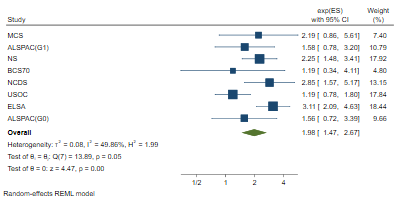 |  |
| Unadjusted | distressed (bin.) | Stable unemployed | MCS | 2.19 | 0.86 | 5.61 | 7.40 |  |  | REML |  |  |
| Unadjusted | distressed (bin.) | Stable unemployed | ALSPAC(G1) | 1.58 | 0.78 | 3.20 | 10.79 |  |  |  |  |  |
| Unadjusted | distressed (bin.) | Stable unemployed | NS | 2.25 | 1.48 | 3.41 | 17.92 |  |  |  |  |  |
| Unadjusted | distressed (bin.) | Stable unemployed | BCS70 | 1.19 | 0.34 | 4.11 | 4.80 |  |  |  |  |  |
| Unadjusted | distressed (bin.) | Stable unemployed | NCDS | 2.85 | 1.57 | 5.17 | 13.15 |  |  |  |  |  |
| Unadjusted | distressed (bin.) | Stable unemployed | USOC | 1.19 | 0.78 | 1.80 | 17.84 |  |  |  |  |  |
| Unadjusted | distressed (bin.) | Stable unemployed | ELSA | 3.11 | 2.09 | 4.63 | 18.44 |  |  |  |  |  |
| Unadjusted | distressed (bin.) | Stable unemployed | GS |  |  |  |  |  | low counts |  |  |  |
| Unadjusted | distressed (bin.) | Stable unemployed | ALSPAC(G0) | 1.56 | 0.72 | 3.39 | 9.66 |  |  |  |  |  |
| Unadjusted | distressed (bin.) | Stable unemployed | Twins UK |  |  |  |  |  | low counts |  |  |  |
| Unadjusted | distressed (bin.) | Stable unemployed | Overall | 1.98 | 1.47 | 2.67 |  | 49.86 |  |  |  |  |
|  |  |  |  |  |  |  |  |  |  |  | 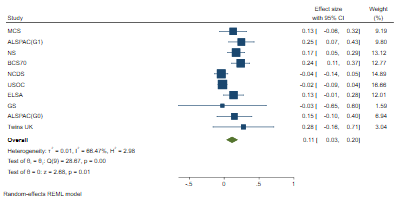 |  |
| Unadjusted | distressed (cont.) | Furloughed | MCS | 0.13 | -0.06 | 0.32 | 9.19 |  |  | REML |  |  |
| Unadjusted | distressed (cont.) | Furloughed | ALSPAC(G1) | 0.25 | 0.07 | 0.43 | 9.80 |  |  |  |  |  |
| Unadjusted | distressed (cont.) | Furloughed | NS | 0.17 | 0.05 | 0.29 | 13.12 |  |  |  |  |  |
| Unadjusted | distressed (cont.) | Furloughed | BCS70 | 0.24 | 0.11 | 0.37 | 12.77 |  |  |  |  |  |
| Unadjusted | distressed (cont.) | Furloughed | NCDS | -0.04 | -0.14 | 0.05 | 14.89 |  |  |  |  |  |
| Unadjusted | distressed (cont.) | Furloughed | USOC | -0.02 | -0.09 | 0.04 | 16.66 |  |  |  |  |  |
| Unadjusted | distressed (cont.) | Furloughed | ELSA | 0.13 | -0.01 | 0.28 | 12.01 |  |  |  |  |  |
| Unadjusted | distressed (cont.) | Furloughed | GS | -0.03 | -0.65 | 0.60 | 1.59 |  |  |  |  |  |
| Unadjusted | distressed (cont.) | Furloughed | ALSPAC(G0) | 0.15 | -0.10 | 0.40 | 6.94 |  |  |  |  |  |
| Unadjusted | distressed (cont.) | Furloughed | Twins UK | 0.28 | -0.16 | 0.71 | 3.04 |  |  |  |  |  |
| Unadjusted | distressed (cont.) | Furloughed | Overall | 0.11 | 0.03 | 0.20 |  | 66.47 |  |  |  |  |
|  |  |  |  |  |  |  |  |  |  |  | 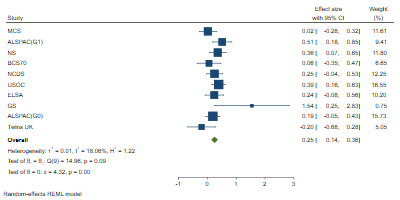 |  |
| Unadjusted | distressed (cont.) | No longer employed | MCS | 0.02 | -0.28 | 0.32 | 11.61 |  |  | REML |  |  |
| Unadjusted | distressed (cont.) | No longer employed | ALSPAC(G1) | 0.51 | 0.18 | 0.85 | 9.41 |  |  |  |  |  |
| Unadjusted | distressed (cont.) | No longer employed | NS | 0.36 | 0.07 | 0.65 | 11.80 |  |  |  |  |  |
| Unadjusted | distressed (cont.) | No longer employed | BCS70 | 0.06 | -0.35 | 0.47 | 6.65 |  |  |  |  |  |
| Unadjusted | distressed (cont.) | No longer employed | NCDS | 0.25 | -0.04 | 0.53 | 12.25 |  |  |  |  |  |
| Unadjusted | distressed (cont.) | No longer employed | USOC | 0.39 | 0.16 | 0.63 | 16.55 |  |  |  |  |  |
| Unadjusted | distressed (cont.) | No longer employed | ELSA | 0.24 | -0.08 | 0.56 | 10.20 |  |  |  |  |  |
| Unadjusted | distressed (cont.) | No longer employed | GS | 1.54 | 0.25 | 2.83 | 0.75 |  |  |  |  |  |
| Unadjusted | distressed (cont.) | No longer employed | ALSPAC(G0) | 0.19 | -0.05 | 0.43 | 15.73 |  |  |  |  |  |
| Unadjusted | distressed (cont.) | No longer employed | Twins UK | -0.20 | -0.68 | 0.28 | 5.05 |  |  |  |  |  |
| Unadjusted | distressed (cont.) | No longer employed | Overall | 0.25 | 0.14 | 0.36 |  | 18.06 |  |  |  |  |
|  |  |  |  |  |  |  |  |  |  |  | 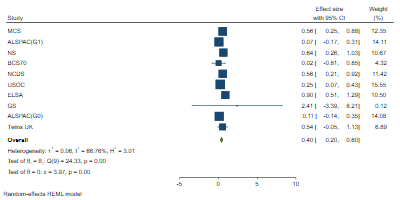 |  |
| Unadjusted | distressed (cont.) | Stable unemployed | MCS | 0.56 | 0.25 | 0.88 | 12.35 |  |  | REML |  |  |
| Unadjusted | distressed (cont.) | Stable unemployed | ALSPAC(G1) | 0.07 | -0.17 | 0.31 | 14.11 |  |  |  |  |  |
| Unadjusted | distressed (cont.) | Stable unemployed | NS | 0.64 | 0.26 | 1.03 | 10.67 |  |  |  |  |  |
| Unadjusted | distressed (cont.) | Stable unemployed | BCS70 | 0.02 | -0.81 | 0.85 | 4.32 |  |  |  |  |  |
| Unadjusted | distressed (cont.) | Stable unemployed | NCDS | 0.56 | 0.21 | 0.92 | 11.42 |  |  |  |  |  |
| Unadjusted | distressed (cont.) | Stable unemployed | USOC | 0.25 | 0.07 | 0.43 | 15.55 |  |  |  |  |  |
| Unadjusted | distressed (cont.) | Stable unemployed | ELSA | 0.90 | 0.51 | 1.29 | 10.50 |  |  |  |  |  |
| Unadjusted | distressed (cont.) | Stable unemployed | GS | 2.41 | -3.39 | 8.21 | 0.12 |  |  |  |  |  |
| Unadjusted | distressed (cont.) | Stable unemployed | ALSPAC(G0) | 0.11 | -0.14 | 0.35 | 14.08 |  |  |  |  |  |
| Unadjusted | distressed (cont.) | Stable unemployed | Twins UK | 0.54 | -0.05 | 1.13 | 6.89 |  |  |  |  |  |
| Unadjusted | distressed (cont.) | Stable unemployed | Overall | 0.40 | 0.20 | 0.60 |  | 66.76 |  |  |  |  |
|  |  |  |  |  |  |  |  |  |  |  | 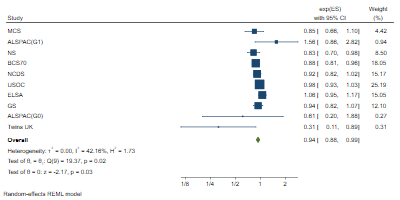 |  |
| Unadjusted | less than daily contact | Furloughed | MCS | 0.85 | 0.66 | 1.10 | 4.42 |  |  | REML |  |  |
| Unadjusted | less than daily contact | Furloughed | ALSPAC(G1) | 1.56 | 0.86 | 2.82 | 0.94 |  |  |  |  |  |
| Unadjusted | less than daily contact | Furloughed | NS | 0.83 | 0.70 | 0.98 | 8.50 |  |  |  |  |  |
| Unadjusted | less than daily contact | Furloughed | BCS70 | 0.88 | 0.81 | 0.96 | 18.05 |  |  |  |  |  |
| Unadjusted | less than daily contact | Furloughed | NCDS | 0.92 | 0.82 | 1.02 | 15.17 |  |  |  |  |  |
| Unadjusted | less than daily contact | Furloughed | USOC | 0.98 | 0.93 | 1.03 | 25.19 |  |  |  |  |  |
| Unadjusted | less than daily contact | Furloughed | ELSA | 1.06 | 0.95 | 1.17 | 15.05 |  |  |  |  |  |
| Unadjusted | less than daily contact | Furloughed | GS | 0.94 | 0.82 | 1.07 | 12.10 |  |  |  |  |  |
| Unadjusted | less than daily contact | Furloughed | ALSPAC(G0) | 0.61 | 0.20 | 1.88 | 0.27 |  |  |  |  |  |
| Unadjusted | less than daily contact | Furloughed | Twins UK | 0.31 | 0.11 | 0.89 | 0.31 |  |  |  |  |  |
| Unadjusted | less than daily contact | Furloughed | Overall | 0.94 | 0.88 | 0.99 |  | 42.16 |  |  |  |  |
|  |  |  |  |  |  |  |  |  |  |  | 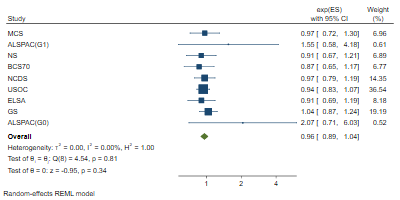 |  |
| Unadjusted | less than daily contact | No longer employed | MCS | 0.97 | 0.72 | 1.30 | 6.96 |  |  | REML |  |  |
| Unadjusted | less than daily contact | No longer employed | ALSPAC(G1) | 1.55 | 0.58 | 4.18 | 0.61 |  |  |  |  |  |
| Unadjusted | less than daily contact | No longer employed | NS | 0.91 | 0.67 | 1.21 | 6.89 |  |  |  |  |  |
| Unadjusted | less than daily contact | No longer employed | BCS70 | 0.87 | 0.65 | 1.17 | 6.77 |  |  |  |  |  |
| Unadjusted | less than daily contact | No longer employed | NCDS | 0.97 | 0.79 | 1.19 | 14.35 |  |  |  |  |  |
| Unadjusted | less than daily contact | No longer employed | USOC | 0.94 | 0.83 | 1.07 | 36.54 |  |  |  |  |  |
| Unadjusted | less than daily contact | No longer employed | ELSA | 0.91 | 0.69 | 1.19 | 8.18 |  |  |  |  |  |
| Unadjusted | less than daily contact | No longer employed | GS | 1.04 | 0.87 | 1.24 | 19.19 |  |  |  |  |  |
| Unadjusted | less than daily contact | No longer employed | ALSPAC(G0) | 2.07 | 0.71 | 6.03 | 0.52 |  |  |  |  |  |
| Unadjusted | less than daily contact | No longer employed | Twins UK |  |  |  |  |  | low counts |  |  |  |
| Unadjusted | less than daily contact | No longer employed | Overall | 0.96 | 0.89 | 1.04 |  | 0.00 |  |  |  |  |
|  |  |  |  |  |  |  |  |  |  |  | 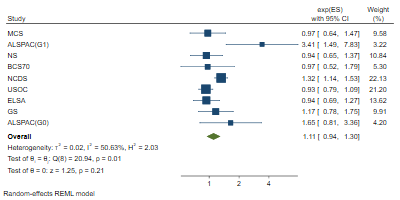 |  |
| Unadjusted | less than daily contact | Stable unemployed | MCS | 0.97 | 0.64 | 1.47 | 9.58 |  |  | REML |  |  |
| Unadjusted | less than daily contact | Stable unemployed | ALSPAC(G1) | 3.41 | 1.49 | 7.83 | 3.22 |  |  |  |  |  |
| Unadjusted | less than daily contact | Stable unemployed | NS | 0.94 | 0.65 | 1.37 | 10.84 |  |  |  |  |  |
| Unadjusted | less than daily contact | Stable unemployed | BCS70 | 0.97 | 0.52 | 1.79 | 5.30 |  |  |  |  |  |
| Unadjusted | less than daily contact | Stable unemployed | NCDS | 1.32 | 1.14 | 1.53 | 22.13 |  |  |  |  |  |
| Unadjusted | less than daily contact | Stable unemployed | USOC | 0.93 | 0.79 | 1.09 | 21.20 |  |  |  |  |  |
| Unadjusted | less than daily contact | Stable unemployed | ELSA | 0.94 | 0.69 | 1.27 | 13.62 |  |  |  |  |  |
| Unadjusted | less than daily contact | Stable unemployed | GS | 1.17 | 0.78 | 1.75 | 9.91 |  |  |  |  |  |
| Unadjusted | less than daily contact | Stable unemployed | ALSPAC(G0) | 1.65 | 0.81 | 3.36 | 4.20 |  |  |  |  |  |
| Unadjusted | less than daily contact | Stable unemployed | Twins UK |  |  |  |  |  | low counts |  |  |  |
| Unadjusted | less than daily contact | Stable unemployed | Overall | 1.11 | 0.94 | 1.30 |  | 50.63 |  |  |  |  |
| Unadjusted | fair or poor self-rated health | Furloughed | MCS | 1.35 | 0.62 | 2.94 | 5.39 |  |  | MLE | 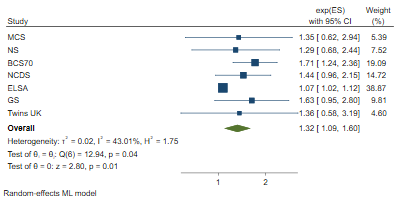 |  |
| Unadjusted | fair or poor self-rated health | Furloughed | ALSPAC(G1) |  |  |  |  |  | no measure |  |  |  |
| Unadjusted | fair or poor self-rated health | Furloughed | NS | 1.29 | 0.68 | 2.44 | 7.52 |  |  |  |  |  |
| Unadjusted | fair or poor self-rated health | Furloughed | BCS70 | 1.71 | 1.24 | 2.36 | 19.09 |  |  |  |  |  |
| Unadjusted | fair or poor self-rated health | Furloughed | NCDS | 1.44 | 0.96 | 2.15 | 14.72 |  |  |  |  |  |
| Unadjusted | fair or poor self-rated health | Furloughed | USOC |  |  |  |  |  | no measure |  |  |  |
| Unadjusted | fair or poor self-rated health | Furloughed | ELSA | 1.07 | 1.02 | 1.12 | 38.87 |  |  |  |  |  |
| Unadjusted | fair or poor self-rated health | Furloughed | GS | 1.63 | 0.95 | 2.80 | 9.81 |  |  |  |  |  |
| Unadjusted | fair or poor self-rated health | Furloughed | ALSPAC(G0) |  |  |  |  |  | no measure |  |  |  |
| Unadjusted | fair or poor self-rated health | Furloughed | Twins UK | 1.36 | 0.58 | 3.19 | 4.60 |  |  |  |  |  |
| Unadjusted | fair or poor self-rated health | Furloughed | Overall | 1.32 | 1.09 | 1.60 |  | 43.01 |  |  |  |  |
| Unadjusted | fair or poor self-rated health | No longer employed | MCS |  |  |  |  |  | low counts | REML |  |  |
| Unadjusted | fair or poor self-rated health | No longer employed | ALSPAC(G1) |  |  |  |  |  | no measure |  |  |  |
| Unadjusted | fair or poor self-rated health | No longer employed | NS | 3.55 | 1.21 | 10.44 | 10.13 |  |  |  |  | 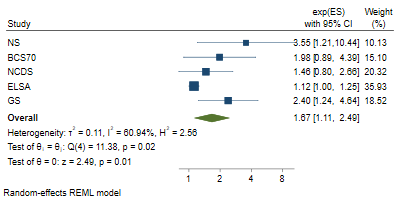 |
| Unadjusted | fair or poor self-rated health | No longer employed | BCS70 | 1.98 | 0.89 | 4.39 | 15.10 |  |  |  |  |  |
| Unadjusted | fair or poor self-rated health | No longer employed | NCDS | 1.46 | 0.80 | 2.66 | 20.32 |  |  |  |  |  |
| Unadjusted | fair or poor self-rated health | No longer employed | USOC |  |  |  |  |  | no measure |  |  |  |
| Unadjusted | fair or poor self-rated health | No longer employed | ELSA | 1.12 | 1.00 | 1.25 | 35.93 |  |  |  |  |  |
| Unadjusted | fair or poor self-rated health | No longer employed | GS | 2.40 | 1.24 | 4.64 | 18.52 |  |  |  |  |  |
| Unadjusted | fair or poor self-rated health | No longer employed | ALSPAC(G0) |  |  |  |  |  | no measure |  |  |  |
| Unadjusted | fair or poor self-rated health | No longer employed | Twins UK |  |  |  |  |  | low counts |  |  |  |
| Unadjusted | fair or poor self-rated health | No longer employed | Overall | 1.67 | 1.11 | 2.49 |  | 60.94 |  |  |  |  |
| Unadjusted | fair or poor self-rated health | Stable unemployed | MCS | 6.24 | 2.58 | 15.11 | 14.76 |  |  | REML | 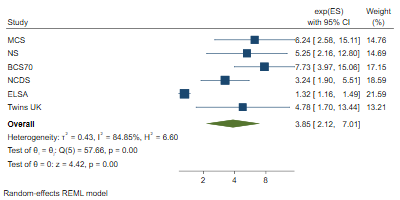 |  |
| Unadjusted | fair or poor self-rated health | Stable unemployed | ALSPAC(G1) |  |  |  |  |  | no measure |  |  |  |
| Unadjusted | fair or poor self-rated health | Stable unemployed | NS | 5.25 | 2.16 | 12.80 | 14.69 |  |  |  |  |  |
| Unadjusted | fair or poor self-rated health | Stable unemployed | BCS70 | 7.73 | 3.97 | 15.06 | 17.15 |  |  |  |  |  |
| Unadjusted | fair or poor self-rated health | Stable unemployed | NCDS | 3.24 | 1.90 | 5.51 | 18.59 |  |  |  |  |  |
| Unadjusted | fair or poor self-rated health | Stable unemployed | USOC |  |  |  |  |  | no measure |  |  |  |
| Unadjusted | fair or poor self-rated health | Stable unemployed | ELSA | 1.32 | 1.16 | 1.49 | 21.59 |  |  |  |  |  |
| Unadjusted | fair or poor self-rated health | Stable unemployed | GS |  |  |  |  |  | low counts |  |  |  |
| Unadjusted | fair or poor self-rated health | Stable unemployed | ALSPAC(G0) |  |  |  |  |  | no measure |  |  |  |
| Unadjusted | fair or poor self-rated health | Stable unemployed | Twins UK | 4.78 | 1.70 | 13.44 | 13.21 |  |  |  |  |  |
| Unadjusted | fair or poor self-rated health | Stable unemployed | Overall | 3.85 | 2.12 | 7.01 |  | 84.85 |  |  |  |  |

### 2. Basic

| **Adjustment** | **Outcome** | **Exposure** | **Study** | **Coefficient** | **lower_ci** | **upper_ci** | **%Weight** | **%I2** | **Reason for missing** | 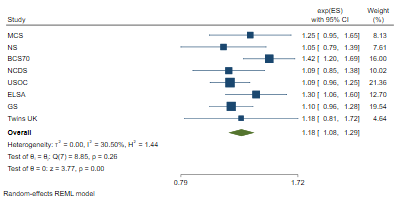 **Method** |
| --- | --- | --- | --- | --- | --- | --- | --- | --- | --- | --- |
| Basic | low life satisfaction | Furloughed | MCS | 1.25 | 0.95 | 1.65 | 8.13 |  |  | REML |
| Basic | low life satisfaction | Furloughed | ALSPAC(G1) | |  |  |  |  | no measure |  |
| Basic | low life satisfaction | Furloughed | NS | 1.05 | 0.79 | 1.39 | 7.61 |  |  |  |
| Basic | low life satisfaction | Furloughed | BCS70 | 1.42 | 1.20 | 1.69 | 16.00 |  |  |  |
| Basic | low life satisfaction | Furloughed | NCDS | 1.09 | 0.85 | 1.38 | 10.02 |  |  |  |
| Basic | low life satisfaction | Furloughed | USOC | 1.09 | 0.96 | 1.25 | 21.36 |  |  |  |
| Basic | low life satisfaction | Furloughed | ELSA | 1.30 | 1.06 | 1.60 | 12.70 |  |  |  |
| Basic | low life satisfaction | Furloughed | GS | 1.10 | 0.96 | 1.28 | 19.54 |  |  |  |
| Basic | low life satisfaction | Furloughed | ALSPAC(G0) | |  |  |  |  | no measure |  |
| Basic | low life satisfaction | Furloughed | Twins UK | 1.18 | 0.81 | 1.72 | 4.64 |  |  |  |
| Basic | low life satisfaction | Furloughed | Overall | 1.18 | 1.08 | 1.29 |  | 30.50 |  |  |
|  |  |  |  |  |  |  |  |  |  | 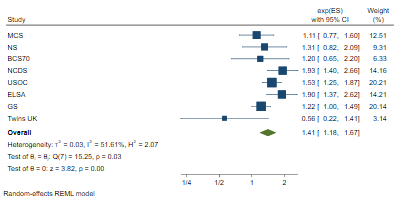 |
| Basic | low life satisfaction | No longer employed | MCS | 1.11 | 0.77 | 1.60 | 12.51 |  |  | REML |
| Basic | low life satisfaction | No longer employed | ALSPAC(G1) | |  |  |  |  | no measure |  |
| Basic | low life satisfaction | No longer employed | NS | 1.31 | 0.82 | 2.09 | 9.31 |  |  |  |
| Basic | low life satisfaction | No longer employed | BCS70 | 1.20 | 0.65 | 2.20 | 6.33 |  |  |  |
| Basic | low life satisfaction | No longer employed | NCDS | 1.93 | 1.40 | 2.66 | 14.16 |  |  |  |
| Basic | low life satisfaction | No longer employed | USOC | 1.53 | 1.25 | 1.87 | 20.21 |  |  |  |
| Basic | low life satisfaction | No longer employed | ELSA | 1.90 | 1.37 | 2.62 | 14.21 |  |  |  |
| Basic | low life satisfaction | No longer employed | GS | 1.22 | 1.00 | 1.49 | 20.14 |  |  |  |
| Basic | low life satisfaction | No longer employed | ALSPAC(G0) | |  |  |  |  | no measure |  |
| Basic | low life satisfaction | No longer employed | Twins UK | 0.56 | 0.22 | 1.41 | 3.14 |  |  |  |
| Basic | low life satisfaction | No longer employed | Overall | 1.41 | 1.18 | 1.67 |  | 51.61 |  |  |
|  |  |  |  |  |  |  |  |  |  | 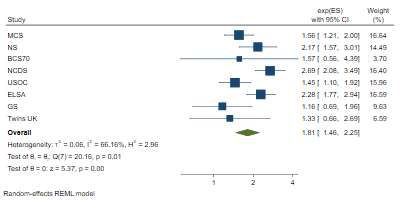 |
| Basic | low life satisfaction | Stable unemployed | MCS | 1.56 | 1.21 | 2.00 | 16.64 |  |  | REML |
| Basic | low life satisfaction | Stable unemployed | ALSPAC(G1) | |  |  |  |  | no measure |  |
| Basic | low life satisfaction | Stable unemployed | NS | 2.17 | 1.57 | 3.01 | 14.49 |  |  |  |
| Basic | low life satisfaction | Stable unemployed | BCS70 | 1.57 | 0.56 | 4.39 | 3.70 |  |  |  |
| Basic | low life satisfaction | Stable unemployed | NCDS | 2.69 | 2.08 | 3.49 | 16.40 |  |  |  |
| Basic | low life satisfaction | Stable unemployed | USOC | 1.45 | 1.10 | 1.92 | 15.96 |  |  |  |
| Basic | low life satisfaction | Stable unemployed | ELSA | 2.28 | 1.77 | 2.94 | 16.59 |  |  |  |
| Basic | low life satisfaction | Stable unemployed | GS | 1.16 | 0.69 | 1.96 | 9.63 |  |  |  |
| Basic | low life satisfaction | Stable unemployed | ALSPAC(G0) | |  |  |  |  | no measure |  |
| Basic | low life satisfaction | Stable unemployed | Twins UK | 1.33 | 0.66 | 2.69 | 6.59 |  |  |  |
| Basic | low life satisfaction | Stable unemployed | Overall | 1.81 | 1.46 | 2.25 |  | 66.16 |  |  |
|  |  |  |  |  |  |  |  |  |  | 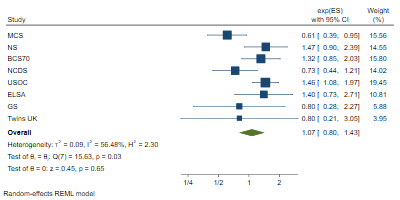 |
| Basic | often lonely | Furloughed | MCS | 0.61 | 0.39 | 0.95 | 15.56 |  |  | REML |
| Basic | often lonely | Furloughed | ALSPAC(G1) | |  |  |  |  | no measure |  |
| Basic | often lonely | Furloughed | NS | 1.47 | 0.90 | 2.39 | 14.55 |  |  |  |
| Basic | often lonely | Furloughed | BCS70 | 1.32 | 0.85 | 2.03 | 15.80 |  |  |  |
| Basic | often lonely | Furloughed | NCDS | 0.73 | 0.44 | 1.21 | 14.02 |  |  |  |
| Basic | often lonely | Furloughed | USOC | 1.46 | 1.08 | 1.97 | 19.45 |  |  |  |
| Basic | often lonely | Furloughed | ELSA | 1.40 | 0.73 | 2.71 | 10.81 |  |  |  |
| Basic | often lonely | Furloughed | GS | 0.80 | 0.28 | 2.27 | 5.88 |  |  |  |
| Basic | often lonely | Furloughed | ALSPAC(G0) | |  |  |  |  | no measure |  |
| Basic | often lonely | Furloughed | Twins UK | 0.80 | 0.21 | 3.05 | 3.95 |  |  |  |
| Basic | often lonely | Furloughed | Overall | 1.07 | 0.80 | 1.43 |  | 56.48 |  |  |
|  |  |  |  |  |  |  |  |  |  | 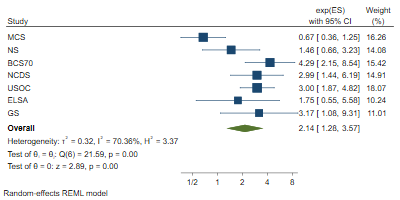 |
| Basic | often lonely | No longer employed | MCS | 0.67 | 0.36 | 1.25 | 16.26 |  |  | REML |
| Basic | often lonely | No longer employed | ALSPAC(G1) | |  |  |  |  | no measure |  |
| Basic | often lonely | No longer employed | NS | 1.46 | 0.66 | 3.23 | 14.08 |  |  |  |
| Basic | often lonely | No longer employed | BCS70 | 4.29 | 2.15 | 8.54 | 15.42 |  |  |  |
| Basic | often lonely | No longer employed | NCDS | 2.99 | 1.44 | 6.19 | 14.91 |  |  |  |
| Basic | often lonely | No longer employed | USOC | 3.00 | 1.87 | 4.82 | 18.07 |  |  |  |
| Basic | often lonely | No longer employed | ELSA | 1.75 | 0.55 | 5.58 | 10.24 |  |  |  |
| Basic | often lonely | No longer employed | GS | 3.17 | 1.08 | 9.31 | 11.01 |  |  |  |
| Basic | often lonely | No longer employed | ALSPAC(G0) | |  |  |  |  | no measure |  |
| Basic | often lonely | No longer employed | Twins UK |  |  |  |  |  | low counts |  |
| Basic | often lonely | No longer employed | Overall | 2.14 | 1.28 | 3.57 |  | 70.36 |  |  |
|  |  |  |  |  |  |  |  |  |  | 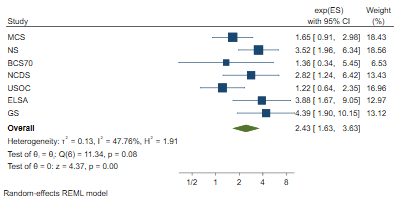 |
| Basic | often lonely | Stable unemployed | MCS | 1.65 | 0.91 | 2.98 | 18.43 |  |  | REML |
| Basic | often lonely | Stable unemployed | ALSPAC(G1) | |  |  |  |  | no measure |  |
| Basic | often lonely | Stable unemployed | NS | 3.52 | 1.96 | 6.34 | 18.56 |  |  |  |
| Basic | often lonely | Stable unemployed | BCS70 | 1.36 | 0.34 | 5.45 | 6.53 |  |  |  |
| Basic | often lonely | Stable unemployed | NCDS | 2.82 | 1.24 | 6.42 | 13.43 |  |  |  |
| Basic | often lonely | Stable unemployed | USOC | 1.22 | 0.64 | 2.35 | 16.96 |  |  |  |
| Basic | often lonely | Stable unemployed | ELSA | 3.88 | 1.67 | 9.05 | 12.97 |  |  |  |
| Basic | often lonely | Stable unemployed | GS | 4.39 | 1.90 | 10.15 | 13.12 |  |  |  |
| Basic | often lonely | Stable unemployed | ALSPAC(G0) | |  |  |  |  | no measure |  |
| Basic | often lonely | Stable unemployed | Twins UK |  |  |  |  |  | low counts |  |
| Basic | often lonely | Stable unemployed | Overall | 2.43 | 1.63 | 3.63 |  | 47.76 |  |  |
|  |  |  |  |  |  |  |  |  |  | 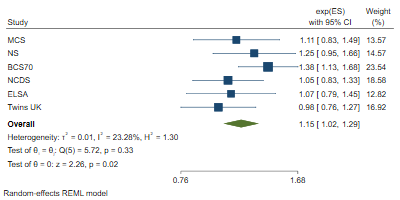 |
| Basic | high loneliness | Furloughed | MCS | 1.11 | 0.83 | 1.49 | 13.57 |  |  | REML |
| Basic | high loneliness | Furloughed | ALSPAC(G1) | |  |  |  |  | no measure |  |
| Basic | high loneliness | Furloughed | NS | 1.25 | 0.95 | 1.66 | 14.57 |  |  |  |
| Basic | high loneliness | Furloughed | BCS70 | 1.38 | 1.13 | 1.68 | 23.54 |  |  |  |
| Basic | high loneliness | Furloughed | NCDS | 1.05 | 0.83 | 1.33 | 18.58 |  |  |  |
| Basic | high loneliness | Furloughed | USOC |  |  |  |  |  | no measure |  |
| Basic | high loneliness | Furloughed | ELSA | 1.07 | 0.79 | 1.45 | 12.82 |  |  |  |
| Basic | high loneliness | Furloughed | GS |  |  |  |  |  | no measure |  |
| Basic | high loneliness | Furloughed | ALSPAC(G0) | |  |  |  |  | no measure |  |
| Basic | high loneliness | Furloughed | Twins UK | 0.98 | 0.76 | 1.27 | 16.92 |  |  |  |
| Basic | high loneliness | Furloughed | Overall | 1.15 | 1.02 | 1.29 |  | 23.28 |  |  |
|  |  |  |  |  |  |  |  |  |  | 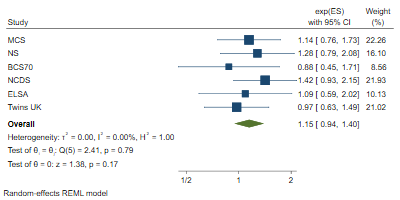 |
| Basic | high loneliness | No longer employed | MCS | 1.14 | 0.76 | 1.73 | 22.26 |  |  | REML |
| Basic | high loneliness | No longer employed | ALSPAC(G1) | |  |  |  |  | no measure |  |
| Basic | high loneliness | No longer employed | NS | 1.28 | 0.79 | 2.08 | 16.10 |  |  |  |
| Basic | high loneliness | No longer employed | BCS70 | 0.88 | 0.45 | 1.71 | 8.56 |  |  |  |
| Basic | high loneliness | No longer employed | NCDS | 1.42 | 0.93 | 2.15 | 21.93 |  |  |  |
| Basic | high loneliness | No longer employed | USOC |  |  |  |  |  | no measure |  |
| Basic | high loneliness | No longer employed | ELSA | 1.09 | 0.59 | 2.02 | 10.13 |  |  |  |
| Basic | high loneliness | No longer employed | GS |  |  |  |  |  | no measure |  |
| Basic | high loneliness | No longer employed | ALSPAC(G0) | |  |  |  |  | no measure |  |
| Basic | high loneliness | No longer employed | Twins UK | 0.97 | 0.63 | 1.49 | 21.02 |  |  |  |
| Basic | high loneliness | No longer employed | Overall | 1.15 | 0.94 | 1.40 |  | 0.00 |  |  |
|  |  |  |  |  |  |  |  |  |  | 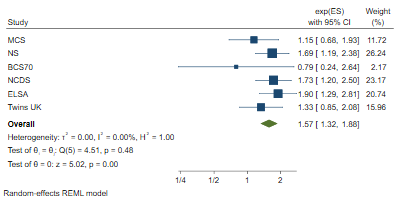 |
| Basic | high loneliness | Stable unemployed | MCS | 1.15 | 0.68 | 1.93 | 11.72 |  |  | REML |
| Basic | high loneliness | Stable unemployed | ALSPAC(G1) | |  |  |  |  | no measure |  |
| Basic | high loneliness | Stable unemployed | NS | 1.69 | 1.19 | 2.38 | 26.24 |  |  |  |
| Basic | high loneliness | Stable unemployed | BCS70 | 0.79 | 0.24 | 2.64 | 2.17 |  |  |  |
| Basic | high loneliness | Stable unemployed | NCDS | 1.73 | 1.20 | 2.50 | 23.17 |  |  |  |
| Basic | high loneliness | Stable unemployed | USOC |  |  |  |  |  | no measure |  |
| Basic | high loneliness | Stable unemployed | ELSA | 1.90 | 1.29 | 2.81 | 20.74 |  |  |  |
| Basic | high loneliness | Stable unemployed | GS |  |  |  |  |  | no measure |  |
| Basic | high loneliness | Stable unemployed | ALSPAC(G0) | |  |  |  |  | no measure |  |
| Basic | high loneliness | Stable unemployed | Twins UK | 1.33 | 0.85 | 2.08 | 15.96 |  |  |  |
| Basic | high loneliness | Stable unemployed | Overall | 1.57 | 1.32 | 1.88 |  | 0.00 |  |  |
|  |  |  |  |  |  |  |  |  |  | 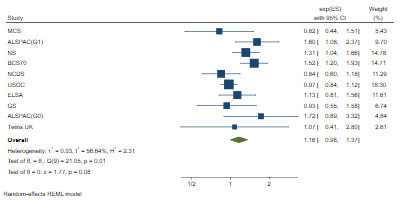 |
| Basic | distressed (bin.) | Furloughed | MCS | 0.82 | 0.44 | 1.51 | 5.43 |  |  | REML |
| Basic | distressed (bin.) | Furloughed | ALSPAC(G1) | 1.60 | 1.08 | 2.37 | 9.70 |  |  |  |
| Basic | distressed (bin.) | Furloughed | NS | 1.31 | 1.04 | 1.66 | 14.78 |  |  |  |
| Basic | distressed (bin.) | Furloughed | BCS70 | 1.52 | 1.20 | 1.93 | 14.71 |  |  |  |
| Basic | distressed (bin.) | Furloughed | NCDS | 0.84 | 0.60 | 1.18 | 11.29 |  |  |  |
| Basic | distressed (bin.) | Furloughed | USOC | 0.97 | 0.84 | 1.12 | 18.30 |  |  |  |
| Basic | distressed (bin.) | Furloughed | ELSA | 1.13 | 0.81 | 1.56 | 11.61 |  |  |  |
| Basic | distressed (bin.) | Furloughed | GS | 0.93 | 0.55 | 1.58 | 6.74 |  |  |  |
| Basic | distressed (bin.) | Furloughed | ALSPAC(G0) | 1.72 | 0.89 | 3.32 | 4.84 |  |  |  |
| Basic | distressed (bin.) | Furloughed | Twins UK | 1.07 | 0.41 | 2.80 | 2.61 |  |  |  |
| Basic | distressed (bin.) | Furloughed | Overall | 1.16 | 0.98 | 1.37 |  | 56.64 |  |  |
|  |  |  |  |  |  |  |  |  |  | 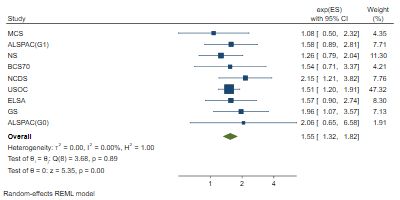 |
| Basic | distressed (bin.) | No longer employed | MCS | 1.08 | 0.50 | 2.32 | 4.35 |  |  | REML |
| Basic | distressed (bin.) | No longer employed | ALSPAC(G1) | 1.58 | 0.89 | 2.81 | 7.71 |  |  |  |
| Basic | distressed (bin.) | No longer employed | NS | 1.26 | 0.79 | 2.04 | 11.30 |  |  |  |
| Basic | distressed (bin.) | No longer employed | BCS70 | 1.54 | 0.71 | 3.37 | 4.21 |  |  |  |
| Basic | distressed (bin.) | No longer employed | NCDS | 2.15 | 1.21 | 3.82 | 7.76 |  |  |  |
| Basic | distressed (bin.) | No longer employed | USOC | 1.51 | 1.20 | 1.91 | 47.32 |  |  |  |
| Basic | distressed (bin.) | No longer employed | ELSA | 1.57 | 0.90 | 2.74 | 8.30 |  |  |  |
| Basic | distressed (bin.) | No longer employed | GS | 1.96 | 1.07 | 3.57 | 7.13 |  |  |  |
| Basic | distressed (bin.) | No longer employed | ALSPAC(G0) | 2.06 | 0.65 | 6.58 | 1.91 |  |  |  |
| Basic | distressed (bin.) | No longer employed | Twins UK |  |  |  |  |  | low counts |  |
| Basic | distressed (bin.) | No longer employed | Overall | 1.55 | 1.32 | 1.82 |  | 0.00 |  |  |
|  |  |  |  |  |  |  |  |  |  | 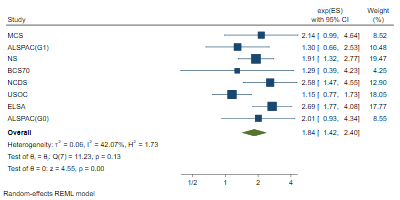 |
| Basic | distressed (bin.) | Stable unemployed | MCS | 2.14 | 0.99 | 4.64 | 8.52 |  |  | REML |
| Basic | distressed (bin.) | Stable unemployed | ALSPAC(G1) | 1.30 | 0.66 | 2.53 | 10.48 |  |  |  |
| Basic | distressed (bin.) | Stable unemployed | NS | 1.91 | 1.32 | 2.77 | 19.47 |  |  |  |
| Basic | distressed (bin.) | Stable unemployed | BCS70 | 1.29 | 0.39 | 4.23 | 4.25 |  |  |  |
| Basic | distressed (bin.) | Stable unemployed | NCDS | 2.58 | 1.47 | 4.55 | 12.90 |  |  |  |
| Basic | distressed (bin.) | Stable unemployed | USOC | 1.15 | 0.77 | 1.73 | 18.05 |  |  |  |
| Basic | distressed (bin.) | Stable unemployed | ELSA | 2.69 | 1.77 | 4.08 | 17.77 |  |  |  |
| Basic | distressed (bin.) | Stable unemployed | GS |  |  |  |  |  | low counts |  |
| Basic | distressed (bin.) | Stable unemployed | ALSPAC(G0) | 2.01 | 0.93 | 4.34 | 8.55 |  |  |  |
| Basic | distressed (bin.) | Stable unemployed | Twins UK |  |  |  |  |  | low counts |  |
| Basic | distressed (bin.) | Stable unemployed | Overall | 1.84 | 1.42 | 2.40 |  | 42.07 |  |  |
|  |  |  |  |  |  |  |  |  |  | 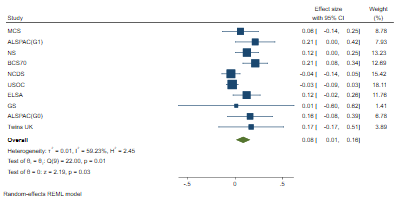 |
| Basic | distressed (cont.) | Furloughed | MCS | 0.06 | -0.14 | 0.25 | 8.78 |  |  | REML |
| Basic | distressed (cont.) | Furloughed | ALSPAC(G1) | 0.21 | 0.00 | 0.42 | 7.93 |  |  |  |
| Basic | distressed (cont.) | Furloughed | NS | 0.12 | 0.00 | 0.25 | 13.23 |  |  |  |
| Basic | distressed (cont.) | Furloughed | BCS70 | 0.21 | 0.08 | 0.34 | 12.69 |  |  |  |
| Basic | distressed (cont.) | Furloughed | NCDS | -0.04 | -0.14 | 0.05 | 15.42 |  |  |  |
| Basic | distressed (cont.) | Furloughed | USOC | -0.03 | -0.09 | 0.03 | 18.11 |  |  |  |
| Basic | distressed (cont.) | Furloughed | ELSA | 0.12 | -0.02 | 0.26 | 11.76 |  |  |  |
| Basic | distressed (cont.) | Furloughed | GS | 0.01 | -0.60 | 0.62 | 1.41 |  |  |  |
| Basic | distressed (cont.) | Furloughed | ALSPAC(G0) | 0.16 | -0.08 | 0.39 | 6.78 |  |  |  |
| Basic | distressed (cont.) | Furloughed | Twins UK | 0.17 | -0.17 | 0.51 | 3.89 |  |  |  |
| Basic | distressed (cont.) | Furloughed | Overall | 0.08 | 0.01 | 0.16 |  | 59.23 |  |  |
|  |  |  |  |  |  |  |  |  |  | 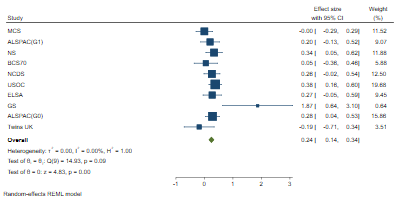 |
| Basic | distressed (cont.) | No longer employed | MCS | 0.00 | -0.29 | 0.29 | 11.52 |  |  | REML |
| Basic | distressed (cont.) | No longer employed | ALSPAC(G1) | 0.20 | -0.13 | 0.52 | 9.07 |  |  |  |
| Basic | distressed (cont.) | No longer employed | NS | 0.34 | 0.05 | 0.62 | 11.88 |  |  |  |
| Basic | distressed (cont.) | No longer employed | BCS70 | 0.05 | -0.36 | 0.46 | 5.88 |  |  |  |
| Basic | distressed (cont.) | No longer employed | NCDS | 0.26 | -0.02 | 0.54 | 12.50 |  |  |  |
| Basic | distressed (cont.) | No longer employed | USOC | 0.38 | 0.16 | 0.60 | 19.68 |  |  |  |
| Basic | distressed (cont.) | No longer employed | ELSA | 0.27 | -0.05 | 0.59 | 9.45 |  |  |  |
| Basic | distressed (cont.) | No longer employed | GS | 1.87 | 0.64 | 3.10 | 0.64 |  |  |  |
| Basic | distressed (cont.) | No longer employed | ALSPAC(G0) | 0.28 | 0.04 | 0.53 | 15.86 |  |  |  |
| Basic | distressed (cont.) | No longer employed | Twins UK | -0.19 | -0.71 | 0.34 | 3.51 |  |  |  |
| Basic | distressed (cont.) | No longer employed | Overall | 0.24 | 0.14 | 0.34 |  | 0.00 |  |  |
|  |  |  |  |  |  |  |  |  |  | 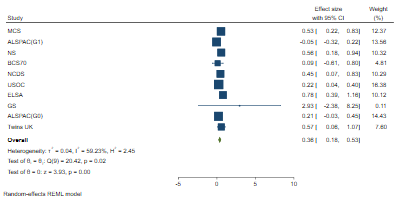 |
| Basic | distressed (cont.) | Stable unemployed | MCS | 0.53 | 0.22 | 0.83 | 12.37 |  |  | REML |
| Basic | distressed (cont.) | Stable unemployed | ALSPAC(G1) | -0.05 | -0.32 | 0.22 | 13.56 |  |  |  |
| Basic | distressed (cont.) | Stable unemployed | NS | 0.56 | 0.18 | 0.94 | 10.32 |  |  |  |
| Basic | distressed (cont.) | Stable unemployed | BCS70 | 0.09 | -0.61 | 0.80 | 4.81 |  |  |  |
| Basic | distressed (cont.) | Stable unemployed | NCDS | 0.45 | 0.07 | 0.83 | 10.29 |  |  |  |
| Basic | distressed (cont.) | Stable unemployed | USOC | 0.22 | 0.04 | 0.40 | 16.38 |  |  |  |
| Basic | distressed (cont.) | Stable unemployed | ELSA | 0.78 | 0.39 | 1.16 | 10.12 |  |  |  |
| Basic | distressed (cont.) | Stable unemployed | GS | 2.93 | -2.38 | 8.25 | 0.11 |  |  |  |
| Basic | distressed (cont.) | Stable unemployed | ALSPAC(G0) | 0.21 | -0.03 | 0.45 | 14.43 |  |  |  |
| Basic | distressed (cont.) | Stable unemployed | Twins UK | 0.57 | 0.06 | 1.07 | 7.60 |  |  |  |
| Basic | distressed (cont.) | Stable unemployed | Overall | 0.36 | 0.18 | 0.53 |  | 59.23 |  |  |
|  |  |  |  |  |  |  |  |  |  | 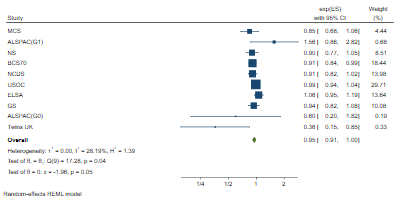 |
| Basic | less than daily contact | Furloughed | MCS | 0.85 | 0.68 | 1.06 | 4.44 |  |  | REML |
| Basic | less than daily contact | Furloughed | ALSPAC(G1) | 1.56 | 0.86 | 2.82 | 0.68 |  |  |  |
| Basic | less than daily contact | Furloughed | NS | 0.90 | 0.77 | 1.05 | 8.51 |  |  |  |
| Basic | less than daily contact | Furloughed | BCS70 | 0.91 | 0.84 | 0.99 | 18.44 |  |  |  |
| Basic | less than daily contact | Furloughed | NCDS | 0.91 | 0.82 | 1.02 | 13.98 |  |  |  |
| Basic | less than daily contact | Furloughed | USOC | 0.99 | 0.94 | 1.04 | 29.71 |  |  |  |
| Basic | less than daily contact | Furloughed | ELSA | 1.06 | 0.95 | 1.19 | 13.64 |  |  |  |
| Basic | less than daily contact | Furloughed | GS | 0.94 | 0.82 | 1.08 | 10.08 |  |  |  |
| Basic | less than daily contact | Furloughed | ALSPAC(G0) | 0.60 | 0.20 | 1.82 | 0.19 |  |  |  |
| Basic | less than daily contact | Furloughed | Twins UK | 0.36 | 0.15 | 0.85 | 0.33 |  |  |  |
| Basic | less than daily contact | Furloughed | Overall | 0.95 | 0.91 | 1.00 |  | 28.19 |  |  |
|  |  |  |  |  |  |  |  |  |  | 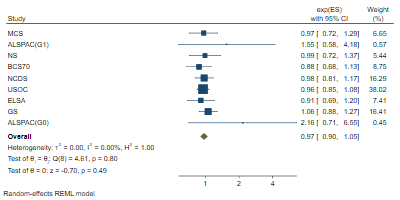 |
| Basic | less than daily contact | No longer employed | MCS | 0.97 | 0.72 | 1.29 | 6.65 |  |  | REML |
| Basic | less than daily contact | No longer employed | ALSPAC(G1) | 1.55 | 0.58 | 4.18 | 0.57 |  |  |  |
| Basic | less than daily contact | No longer employed | NS | 0.99 | 0.72 | 1.37 | 5.44 |  |  |  |
| Basic | less than daily contact | No longer employed | BCS70 | 0.88 | 0.68 | 1.13 | 8.75 |  |  |  |
| Basic | less than daily contact | No longer employed | NCDS | 0.98 | 0.81 | 1.17 | 16.29 |  |  |  |
| Basic | less than daily contact | No longer employed | USOC | 0.96 | 0.85 | 1.08 | 38.02 |  |  |  |
| Basic | less than daily contact | No longer employed | ELSA | 0.91 | 0.69 | 1.20 | 7.41 |  |  |  |
| Basic | less than daily contact | No longer employed | GS | 1.06 | 0.88 | 1.27 | 16.41 |  |  |  |
| Basic | less than daily contact | No longer employed | ALSPAC(G0) | 2.16 | 0.71 | 6.55 | 0.45 |  |  |  |
| Basic | less than daily contact | No longer employed | Twins UK |  |  |  |  |  | low counts |  |
| Basic | less than daily contact | No longer employed | Overall | 0.97 | 0.90 | 1.05 |  | 0.00 |  |  |
|  |  |  |  |  |  |  |  |  |  | 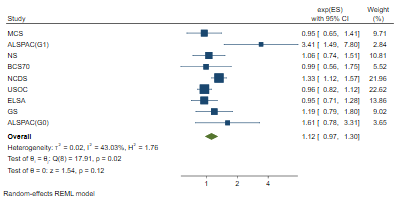 |
| Basic | less than daily contact | Stable unemployed | MCS | 0.95 | 0.65 | 1.41 | 9.71 |  |  | REML |
| Basic | less than daily contact | Stable unemployed | ALSPAC(G1) | 3.41 | 1.49 | 7.80 | 2.84 |  |  |  |
| Basic | less than daily contact | Stable unemployed | NS | 1.06 | 0.74 | 1.51 | 10.81 |  |  |  |
| Basic | less than daily contact | Stable unemployed | BCS70 | 0.99 | 0.56 | 1.75 | 5.52 |  |  |  |
| Basic | less than daily contact | Stable unemployed | NCDS | 1.33 | 1.12 | 1.57 | 21.96 |  |  |  |
| Basic | less than daily contact | Stable unemployed | USOC | 0.96 | 0.82 | 1.12 | 22.62 |  |  |  |
| Basic | less than daily contact | Stable unemployed | ELSA | 0.95 | 0.71 | 1.28 | 13.86 |  |  |  |
| Basic | less than daily contact | Stable unemployed | GS | 1.19 | 0.79 | 1.80 | 9.02 |  |  |  |
| Basic | less than daily contact | Stable unemployed | ALSPAC(G0) | 1.61 | 0.78 | 3.31 | 3.65 |  |  |  |
| Basic | less than daily contact | Stable unemployed | Twins UK |  |  |  |  |  | low counts |  |
| Basic | less than daily contact | Stable unemployed | Overall | 1.12 | 0.97 | 1.30 |  | 43.03 |  |  |
| Basic | fair or poor self-rated health | Furloughed | MCS | 1.35 | 0.63 | 2.91 | 4.75 |  |  | 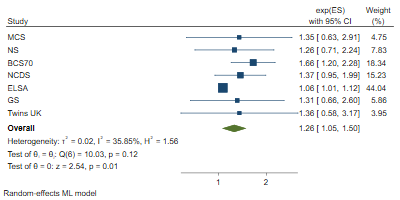 MLE |
| Basic | fair or poor self-rated health | Furloughed | ALSPAC(G1) | |  |  |  |  | no measure |  |
| Basic | fair or poor self-rated health | Furloughed | NS | 1.26 | 0.71 | 2.24 | 7.83 |  |  |  |
| Basic | fair or poor self-rated health | Furloughed | BCS70 | 1.66 | 1.20 | 2.28 | 18.34 |  |  |  |
| Basic | fair or poor self-rated health | Furloughed | NCDS | 1.37 | 0.95 | 1.99 | 15.23 |  |  |  |
| Basic | fair or poor self-rated health | Furloughed | USOC |  |  |  |  |  | no measure |  |
| Basic | fair or poor self-rated health | Furloughed | ELSA | 1.06 | 1.01 | 1.12 | 44.04 |  |  |  |
| Basic | fair or poor self-rated health | Furloughed | GS | 1.31 | 0.66 | 2.60 | 5.86 |  |  |  |
| Basic | fair or poor self-rated health | Furloughed | ALSPAC(G0) | |  |  |  |  | no measure |  |
| Basic | fair or poor self-rated health | Furloughed | Twins UK | 1.36 | 0.58 | 3.17 | 3.95 |  |  |  |
| Basic | fair or poor self-rated health | Furloughed | Overall | 1.26 | 1.05 | 1.50 |  | 35.85 |  |  |
| Basic | fair or poor self-rated health | No longer employed | MCS |  |  |  |  |  | low counts | 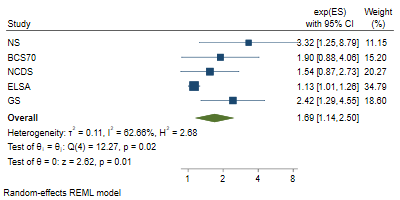 REML |
| Basic | fair or poor self-rated health | No longer employed | ALSPAC(G1) | |  |  |  |  | no measure |  |
| Basic | fair or poor self-rated health | No longer employed | NS | 3.32 | 1.25 | 8.79 | 11.15 |  |  |  |
| Basic | fair or poor self-rated health | No longer employed | BCS70 | 1.90 | 0.88 | 4.06 | 15.20 |  |  |  |
| Basic | fair or poor self-rated health | No longer employed | NCDS | 1.54 | 0.87 | 2.73 | 20.27 |  |  |  |
| Basic | fair or poor self-rated health | No longer employed | USOC |  |  |  |  |  | no measure |  |
| Basic | fair or poor self-rated health | No longer employed | ELSA | 1.13 | 1.01 | 1.26 | 34.79 |  |  |  |
| Basic | fair or poor self-rated health | No longer employed | GS | 2.42 | 1.29 | 4.55 | 18.60 |  |  |  |
| Basic | fair or poor self-rated health | No longer employed | ALSPAC(G0) | |  |  |  |  | no measure |  |
| Basic | fair or poor self-rated health | No longer employed | Twins UK |  |  |  |  |  | low counts |  |
| Basic | fair or poor self-rated health | No longer employed | Overall | 1.69 | 1.14 | 2.50 |  | 62.66 |  |  |
| Basic | fair or poor self-rated health | Stable unemployed | MCS | 5.52 | 2.18 | 13.98 | 13.50 |  |  | 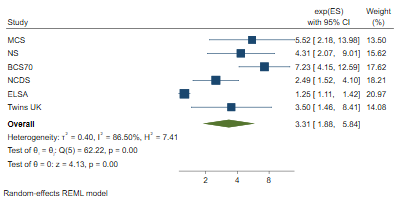 REML |
| Basic | fair or poor self-rated health | Stable unemployed | ALSPAC(G1) | |  |  |  |  | no measure |  |
| Basic | fair or poor self-rated health | Stable unemployed | NS | 4.31 | 2.07 | 9.01 | 15.62 |  |  |  |
| Basic | fair or poor self-rated health | Stable unemployed | BCS70 | 7.23 | 4.15 | 12.59 | 17.62 |  |  |  |
| Basic | fair or poor self-rated health | Stable unemployed | NCDS | 2.49 | 1.52 | 4.10 | 18.21 |  |  |  |
| Basic | fair or poor self-rated health | Stable unemployed | USOC |  |  |  |  |  | no measure |  |
| Basic | fair or poor self-rated health | Stable unemployed | ELSA | 1.25 | 1.11 | 1.42 | 20.97 |  |  |  |
| Basic | fair or poor self-rated health | Stable unemployed | GS |  |  |  |  |  | low counts |  |
| Basic | fair or poor self-rated health | Stable unemployed | ALSPAC(G0) | |  |  |  |  | no measure |  |
| Basic | fair or poor self-rated health | Stable unemployed | Twins UK | 3.50 | 1.46 | 8.41 | 14.08 |  |  |  |
| Basic | fair or poor self-rated health | Stable unemployed | Overall | 3.31 | 1.88 | 5.84 |  | 86.50 |  |  |

### 3. Full

| **Adjustment** | **Outcome** | **Exposure** | **Study** | **Coefficient** | **lower_ci** | **upper_ci** | **%Weight** | **%I2** | **Reason for missing** | **Method** |  |  |
| --- | --- | --- | --- | --- | --- | --- | --- | --- | --- | --- | --- | --- |
| Full | low life satisfaction | Furloughed | MCS | 1.23 | 0.96 | 1.58 | 7.40 |  |  | REML |  |  |
| Full | low life satisfaction | Furloughed | ALSPAC(G1) | |  |  |  |  | no measure |  |  |  |
| Full | low life satisfaction | Furloughed | NS | 1.01 | 0.76 | 1.35 | 5.53 |  |  |  |  |  |
| Full | low life satisfaction | Furloughed | BCS70 | 1.31 | 1.11 | 1.53 | 16.77 |  |  |  |  |  |
| Full | low life satisfaction | Furloughed | NCDS | 1.16 | 0.93 | 1.43 | 9.76 |  |  |  |  | 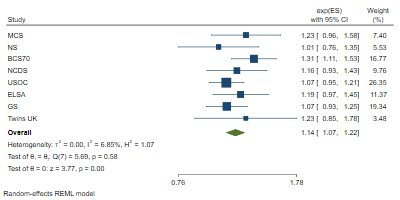 |
| Full | low life satisfaction | Furloughed | USOC | 1.07 | 0.95 | 1.21 | 26.35 |  |  |  |  |  |
| Full | low life satisfaction | Furloughed | ELSA | 1.19 | 0.97 | 1.45 | 11.37 |  |  |  |  |  |
| Full | low life satisfaction | Furloughed | GS | 1.07 | 0.93 | 1.25 | 19.34 |  |  |  |  |  |
| Full | low life satisfaction | Furloughed | ALSPAC(G0) | |  |  |  |  | no measure |  |  |  |
| Full | low life satisfaction | Furloughed | Twins UK | 1.23 | 0.85 | 1.78 | 3.48 |  |  |  |  |  |
| Full | low life satisfaction | Furloughed | Overall | 1.14 | 1.07 | 1.22 |  | 6.85 |  |  |  |  |
|  |  |  |  |  |  |  |  |  |  | 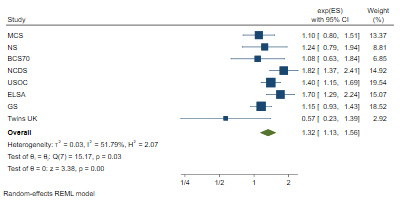 |  |  |
| Full | low life satisfaction | No longer employed | MCS | 1.10 | 0.80 | 1.51 | 13.37 |  |  | REML |  |  |
| Full | low life satisfaction | No longer employed | ALSPAC(G1) | |  |  |  |  | no measure |  |  |  |
| Full | low life satisfaction | No longer employed | NS | 1.24 | 0.79 | 1.94 | 8.81 |  |  |  |  |  |
| Full | low life satisfaction | No longer employed | BCS70 | 1.08 | 0.63 | 1.84 | 6.85 |  |  |  |  |  |
| Full | low life satisfaction | No longer employed | NCDS | 1.82 | 1.37 | 2.41 | 14.92 |  |  |  |  |  |
| Full | low life satisfaction | No longer employed | USOC | 1.40 | 1.15 | 1.69 | 19.54 |  |  |  |  |  |
| Full | low life satisfaction | No longer employed | ELSA | 1.70 | 1.29 | 2.24 | 15.07 |  |  |  |  |  |
| Full | low life satisfaction | No longer employed | GS | 1.15 | 0.93 | 1.43 | 18.52 |  |  |  |  |  |
| Full | low life satisfaction | No longer employed | ALSPAC(G0) | |  |  |  |  | no measure |  |  |  |
| Full | low life satisfaction | No longer employed | Twins UK | 0.57 | 0.23 | 1.39 | 2.92 |  |  |  |  |  |
| Full | low life satisfaction | No longer employed | Overall | 1.32 | 1.13 | 1.56 |  | 51.79 |  |  |  |  |
|  |  |  |  |  |  |  |  |  |  | 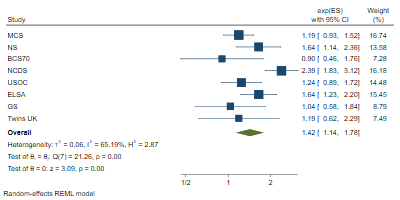 |  |  |
| Full | low life satisfaction | Stable unemployed | MCS | 1.19 | 0.93 | 1.52 | 16.74 |  |  | REML |  |  |
| Full | low life satisfaction | Stable unemployed | ALSPAC(G1) | |  |  |  |  | no measure |  |  |  |
| Full | low life satisfaction | Stable unemployed | NS | 1.64 | 1.14 | 2.36 | 13.58 |  |  |  |  |  |
| Full | low life satisfaction | Stable unemployed | BCS70 | 0.90 | 0.46 | 1.76 | 7.28 |  |  |  |  |  |
| Full | low life satisfaction | Stable unemployed | NCDS | 2.39 | 1.83 | 3.12 | 16.18 |  |  |  |  |  |
| Full | low life satisfaction | Stable unemployed | USOC | 1.24 | 0.89 | 1.72 | 14.48 |  |  |  |  |  |
| Full | low life satisfaction | Stable unemployed | ELSA | 1.64 | 1.23 | 2.20 | 15.45 |  |  |  |  |  |
| Full | low life satisfaction | Stable unemployed | GS | 1.04 | 0.58 | 1.84 | 8.79 |  |  |  |  |  |
| Full | low life satisfaction | Stable unemployed | ALSPAC(G0) | |  |  |  |  | no measure |  |  |  |
| Full | low life satisfaction | Stable unemployed | Twins UK | 1.19 | 0.62 | 2.29 | 7.49 |  |  |  |  |  |
| Full | low life satisfaction | Stable unemployed | Overall | 1.42 | 1.14 | 1.78 |  | 65.19 |  |  |  |  |
|  |  |  |  |  |  |  |  |  |  | 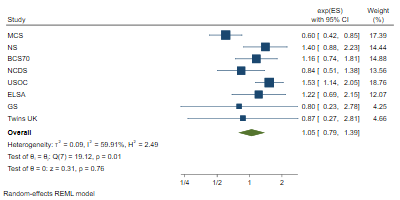 |  |  |
| Full | often lonely | Furloughed | MCS | 0.60 | 0.42 | 0.85 | 17.39 |  |  | REML |  |  |
| Full | often lonely | Furloughed | ALSPAC(G1) | |  |  |  |  | no measure |  |  |  |
| Full | often lonely | Furloughed | NS | 1.40 | 0.88 | 2.23 | 14.44 |  |  |  |  |  |
| Full | often lonely | Furloughed | BCS70 | 1.16 | 0.74 | 1.81 | 14.88 |  |  |  |  |  |
| Full | often lonely | Furloughed | NCDS | 0.84 | 0.51 | 1.38 | 13.56 |  |  |  |  |  |
| Full | often lonely | Furloughed | USOC | 1.53 | 1.14 | 2.05 | 18.76 |  |  |  |  |  |
| Full | often lonely | Furloughed | ELSA | 1.22 | 0.69 | 2.15 | 12.07 |  |  |  |  |  |
| Full | often lonely | Furloughed | GS | 0.80 | 0.23 | 2.78 | 4.25 |  |  |  |  |  |
| Full | often lonely | Furloughed | ALSPAC(G0) | |  |  |  |  | no measure |  |  |  |
| Full | often lonely | Furloughed | Twins UK | 0.87 | 0.27 | 2.81 | 4.66 |  |  |  |  |  |
| Full | often lonely | Furloughed | Overall | 1.05 | 0.79 | 1.39 |  | 59.91 |  |  |  |  |
|  |  |  |  |  |  |  |  |  |  | 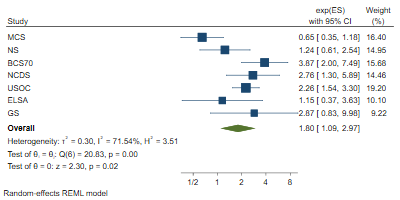 |  |  |
| Full | often lonely | No longer employed | MCS | 0.65 | 0.35 | 1.18 | 16.40 |  |  | REML |  |  |
| Full | often lonely | No longer employed | ALSPAC(G1) | |  |  |  |  | no measure |  |  |  |
| Full | often lonely | No longer employed | NS | 1.24 | 0.61 | 2.54 | 14.95 |  |  |  |  |  |
| Full | often lonely | No longer employed | BCS70 | 3.87 | 2.00 | 7.49 | 15.68 |  |  |  |  |  |
| Full | often lonely | No longer employed | NCDS | 2.76 | 1.30 | 5.89 | 14.46 |  |  |  |  |  |
| Full | often lonely | No longer employed | USOC | 2.26 | 1.54 | 3.30 | 19.20 |  |  |  |  |  |
| Full | often lonely | No longer employed | ELSA | 1.15 | 0.37 | 3.63 | 10.10 |  |  |  |  |  |
| Full | often lonely | No longer employed | GS | 2.87 | 0.83 | 9.98 | 9.22 |  |  |  |  |  |
| Full | often lonely | No longer employed | ALSPAC(G0) | |  |  |  |  | no measure |  |  |  |
| Full | often lonely | No longer employed | Twins UK |  |  |  |  |  | low counts |  |  |  |
| Full | often lonely | No longer employed | Overall | 1.80 | 1.09 | 2.97 |  | 71.54 |  |  |  |  |
|  |  |  |  |  |  |  |  |  |  | 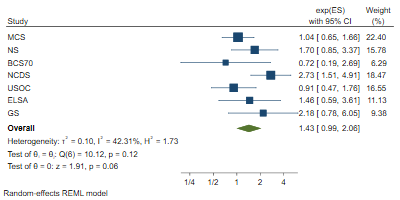 |  |  |
| Full | often lonely | Stable unemployed | MCS | 1.04 | 0.65 | 1.66 | 22.40 |  |  | REML |  |  |
| Full | often lonely | Stable unemployed | ALSPAC(G1) | |  |  |  |  | no measure |  |  |  |
| Full | often lonely | Stable unemployed | NS | 1.70 | 0.85 | 3.37 | 15.78 |  |  |  |  |  |
| Full | often lonely | Stable unemployed | BCS70 | 0.72 | 0.19 | 2.69 | 6.29 |  |  |  |  |  |
| Full | often lonely | Stable unemployed | NCDS | 2.73 | 1.51 | 4.91 | 18.47 |  |  |  |  |  |
| Full | often lonely | Stable unemployed | USOC | 0.91 | 0.47 | 1.76 | 16.55 |  |  |  |  |  |
| Full | often lonely | Stable unemployed | ELSA | 1.46 | 0.59 | 3.61 | 11.13 |  |  |  |  |  |
| Full | often lonely | Stable unemployed | GS | 2.18 | 0.78 | 6.05 | 9.38 |  |  |  |  |  |
| Full | often lonely | Stable unemployed | ALSPAC(G0) | |  |  |  |  | no measure |  |  |  |
| Full | often lonely | Stable unemployed | Twins UK |  |  |  |  |  | low counts |  |  |  |
| Full | often lonely | Stable unemployed | Overall | 1.43 | 0.99 | 2.06 |  | 42.31 |  |  |  |  |
|  |  |  |  |  |  |  |  |  |  | 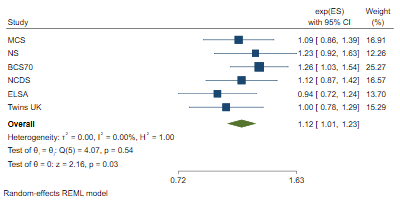 |  |  |
| Full | high loneliness | Furloughed | MCS | 1.09 | 0.86 | 1.39 | 16.91 |  |  | REML |  |  |
| Full | high loneliness | Furloughed | ALSPAC(G1) | |  |  |  |  | no measure |  |  |  |
| Full | high loneliness | Furloughed | NS | 1.23 | 0.92 | 1.63 | 12.26 |  |  |  |  |  |
| Full | high loneliness | Furloughed | BCS70 | 1.26 | 1.03 | 1.54 | 25.27 |  |  |  |  |  |
| Full | high loneliness | Furloughed | NCDS | 1.12 | 0.87 | 1.42 | 16.57 |  |  |  |  |  |
| Full | high loneliness | Furloughed | USOC |  |  |  |  |  | no measure |  |  |  |
| Full | high loneliness | Furloughed | ELSA | 0.94 | 0.72 | 1.24 | 13.70 |  |  |  |  |  |
| Full | high loneliness | Furloughed | GS |  |  |  |  |  | no measure |  |  |  |
| Full | high loneliness | Furloughed | ALSPAC(G0) | |  |  |  |  | no measure |  |  |  |
| Full | high loneliness | Furloughed | Twins UK | 1.00 | 0.78 | 1.29 | 15.29 |  |  |  |  |  |
| Full | high loneliness | Furloughed | Overall | 1.12 | 1.01 | 1.23 |  | 0.00 |  |  |  |  |
|  |  |  |  |  |  |  |  |  |  | 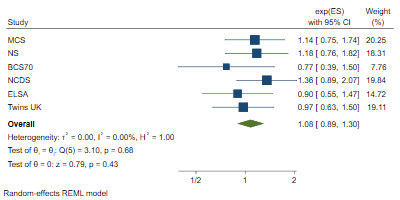 |  |  |
| Full | high loneliness | No longer employed | MCS | 1.14 | 0.75 | 1.74 | 20.25 |  |  | REML |  |  |
| Full | high loneliness | No longer employed | ALSPAC(G1) | |  |  |  |  | no measure |  |  |  |
| Full | high loneliness | No longer employed | NS | 1.18 | 0.76 | 1.82 | 18.31 |  |  |  |  |  |
| Full | high loneliness | No longer employed | BCS70 | 0.77 | 0.39 | 1.50 | 7.76 |  |  |  |  |  |
| Full | high loneliness | No longer employed | NCDS | 1.36 | 0.89 | 2.07 | 19.84 |  |  |  |  |  |
| Full | high loneliness | No longer employed | USOC |  |  |  |  |  | no measure |  |  |  |
| Full | high loneliness | No longer employed | ELSA | 0.90 | 0.55 | 1.47 | 14.72 |  |  |  |  |  |
| Full | high loneliness | No longer employed | GS |  |  |  |  |  | no measure |  |  |  |
| Full | high loneliness | No longer employed | ALSPAC(G0) | |  |  |  |  | no measure |  |  |  |
| Full | high loneliness | No longer employed | Twins UK | 0.97 | 0.63 | 1.50 | 19.11 |  |  |  |  |  |
| Full | high loneliness | No longer employed | Overall | 1.08 | 0.89 | 1.30 |  | 0.00 |  |  |  |  |
|  |  |  |  |  |  |  |  |  |  | 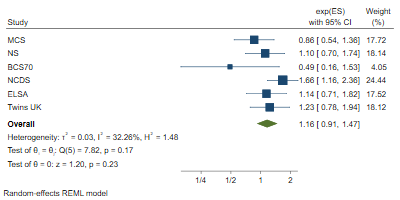 |  |  |
| Full | high loneliness | Stable unemployed | MCS | 0.86 | 0.54 | 1.36 | 17.72 |  |  | REML |  |  |
| Full | high loneliness | Stable unemployed | ALSPAC(G1) | |  |  |  |  | no measure |  |  |  |
| Full | high loneliness | Stable unemployed | NS | 1.10 | 0.70 | 1.74 | 18.14 |  |  |  |  |  |
| Full | high loneliness | Stable unemployed | BCS70 | 0.49 | 0.16 | 1.53 | 4.05 |  |  |  |  |  |
| Full | high loneliness | Stable unemployed | NCDS | 1.66 | 1.16 | 2.36 | 24.44 |  |  |  |  |  |
| Full | high loneliness | Stable unemployed | USOC |  |  |  |  |  | no measure |  |  |  |
| Full | high loneliness | Stable unemployed | ELSA | 1.14 | 0.71 | 1.82 | 17.52 |  |  |  |  |  |
| Full | high loneliness | Stable unemployed | GS |  |  |  |  |  | no measure |  |  |  |
| Full | high loneliness | Stable unemployed | ALSPAC(G0) | |  |  |  |  | no measure |  |  |  |
| Full | high loneliness | Stable unemployed | Twins UK | 1.23 | 0.78 | 1.94 | 18.12 |  |  |  |  |  |
| Full | high loneliness | Stable unemployed | Overall | 1.16 | 0.91 | 1.47 |  | 32.26 |  |  |  |  |
|  |  |  |  |  |  |  |  |  |  | 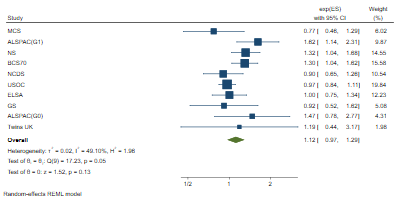 |  |  |
| Full | distressed (bin.) | Furloughed | MCS | 0.77 | 0.46 | 1.29 | 6.02 |  |  | REML |  |  |
| Full | distressed (bin.) | Furloughed | ALSPAC(G1) | 1.62 | 1.14 | 2.31 | 9.87 |  |  |  |  |  |
| Full | distressed (bin.) | Furloughed | NS | 1.32 | 1.04 | 1.68 | 14.55 |  |  |  |  |  |
| Full | distressed (bin.) | Furloughed | BCS70 | 1.30 | 1.04 | 1.62 | 15.58 |  |  |  |  |  |
| Full | distressed (bin.) | Furloughed | NCDS | 0.90 | 0.65 | 1.26 | 10.54 |  |  |  |  |  |
| Full | distressed (bin.) | Furloughed | USOC | 0.97 | 0.84 | 1.11 | 19.84 |  |  |  |  |  |
| Full | distressed (bin.) | Furloughed | ELSA | 1.00 | 0.75 | 1.34 | 12.23 |  |  |  |  |  |
| Full | distressed (bin.) | Furloughed | GS | 0.92 | 0.52 | 1.62 | 5.08 |  |  |  |  |  |
| Full | distressed (bin.) | Furloughed | ALSPAC(G0) | 1.47 | 0.78 | 2.77 | 4.31 |  |  |  |  |  |
| Full | distressed (bin.) | Furloughed | Twins UK | 1.19 | 0.44 | 3.17 | 1.98 |  |  |  |  |  |
| Full | distressed (bin.) | Furloughed | Overall | 1.12 | 0.97 | 1.29 |  | 49.10 |  |  |  |  |
|  |  |  |  |  |  |  |  |  |  | 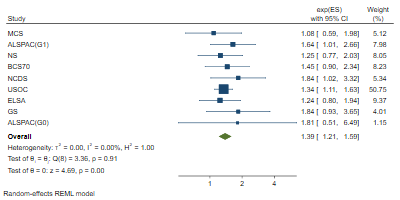 |  |  |
| Full | distressed (bin.) | No longer employed | MCS | 1.08 | 0.59 | 1.98 | 5.12 |  |  | REML |  |  |
| Full | distressed (bin.) | No longer employed | ALSPAC(G1) | 1.64 | 1.01 | 2.66 | 7.98 |  |  |  |  |  |
| Full | distressed (bin.) | No longer employed | NS | 1.25 | 0.77 | 2.03 | 8.05 |  |  |  |  |  |
| Full | distressed (bin.) | No longer employed | BCS70 | 1.45 | 0.90 | 2.34 | 8.23 |  |  |  |  |  |
| Full | distressed (bin.) | No longer employed | NCDS | 1.84 | 1.02 | 3.32 | 5.34 |  |  |  |  |  |
| Full | distressed (bin.) | No longer employed | USOC | 1.34 | 1.11 | 1.63 | 50.75 |  |  |  |  |  |
| Full | distressed (bin.) | No longer employed | ELSA | 1.24 | 0.80 | 1.94 | 9.37 |  |  |  |  |  |
| Full | distressed (bin.) | No longer employed | GS | 1.84 | 0.93 | 3.65 | 4.01 |  |  |  |  |  |
| Full | distressed (bin.) | No longer employed | ALSPAC(G0) | 1.81 | 0.51 | 6.49 | 1.15 |  |  |  |  |  |
| Full | distressed (bin.) | No longer employed | Twins UK |  |  |  |  |  | low counts |  |  |  |
| Full | distressed (bin.) | No longer employed | Overall | 1.39 | 1.21 | 1.59 |  | 0.00 |  |  |  |  |
|  |  |  |  |  |  |  |  |  |  | 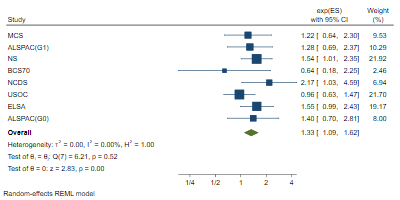 |  |  |
| Full | distressed (bin.) | Stable unemployed | MCS | 1.22 | 0.64 | 2.30 | 9.53 |  |  | REML |  |  |
| Full | distressed (bin.) | Stable unemployed | ALSPAC(G1) | 1.28 | 0.69 | 2.37 | 10.29 |  |  |  |  |  |
| Full | distressed (bin.) | Stable unemployed | NS | 1.54 | 1.01 | 2.35 | 21.92 |  |  |  |  |  |
| Full | distressed (bin.) | Stable unemployed | BCS70 | 0.64 | 0.18 | 2.25 | 2.46 |  |  |  |  |  |
| Full | distressed (bin.) | Stable unemployed | NCDS | 2.17 | 1.03 | 4.59 | 6.94 |  |  |  |  |  |
| Full | distressed (bin.) | Stable unemployed | USOC | 0.96 | 0.63 | 1.47 | 21.70 |  |  |  |  |  |
| Full | distressed (bin.) | Stable unemployed | ELSA | 1.55 | 0.99 | 2.43 | 19.17 |  |  |  |  |  |
| Full | distressed (bin.) | Stable unemployed | GS |  |  |  |  |  | low counts |  |  |  |
| Full | distressed (bin.) | Stable unemployed | ALSPAC(G0) | 1.40 | 0.70 | 2.81 | 8.00 |  |  |  |  |  |
| Full | distressed (bin.) | Stable unemployed | Twins UK |  |  |  |  |  | low counts |  |  |  |
| Full | distressed (bin.) | Stable unemployed | Overall | 1.33 | 1.09 | 1.62 |  | 0.00 |  |  |  |  |
|  |  |  |  |  |  |  |  |  |  | 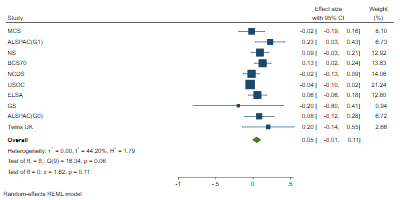 |  |  |
| Full | distressed (cont.) | Furloughed | MCS | -0.02 | -0.19 | 0.16 | 8.10 |  |  | REML |  |  |
| Full | distressed (cont.) | Furloughed | ALSPAC(G1) | 0.23 | 0.03 | 0.43 | 6.73 |  |  |  |  |  |
| Full | distressed (cont.) | Furloughed | NS | 0.09 | -0.03 | 0.21 | 12.92 |  |  |  |  |  |
| Full | distressed (cont.) | Furloughed | BCS70 | 0.13 | 0.02 | 0.24 | 13.83 |  |  |  |  |  |
| Full | distressed (cont.) | Furloughed | NCDS | -0.02 | -0.13 | 0.09 | 14.06 |  |  |  |  |  |
| Full | distressed (cont.) | Furloughed | USOC | -0.04 | -0.10 | 0.02 | 21.24 |  |  |  |  |  |
| Full | distressed (cont.) | Furloughed | ELSA | 0.06 | -0.06 | 0.18 | 12.80 |  |  |  |  |  |
| Full | distressed (cont.) | Furloughed | GS | -0.20 | -0.80 | 0.41 | 0.94 |  |  |  |  |  |
| Full | distressed (cont.) | Furloughed | ALSPAC(G0) | 0.08 | -0.12 | 0.28 | 6.72 |  |  |  |  |  |
| Full | distressed (cont.) | Furloughed | Twins UK | 0.20 | -0.14 | 0.55 | 2.66 |  |  |  |  |  |
| Full | distressed (cont.) | Furloughed | Overall | 0.05 | -0.01 | 0.11 |  | 44.20 |  |  |  |  |
|  |  |  |  |  |  |  |  |  |  | 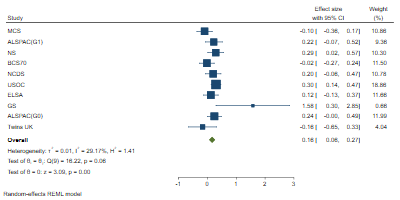 |  |  |
| Full | distressed (cont.) | No longer employed | MCS | -0.10 | -0.36 | 0.17 | 10.86 |  |  | REML |  |  |
| Full | distressed (cont.) | No longer employed | ALSPAC(G1) | 0.22 | -0.07 | 0.52 | 9.36 |  |  |  |  |  |
| Full | distressed (cont.) | No longer employed | NS | 0.29 | 0.02 | 0.57 | 10.30 |  |  |  |  |  |
| Full | distressed (cont.) | No longer employed | BCS70 | -0.02 | -0.27 | 0.24 | 11.50 |  |  |  |  |  |
| Full | distressed (cont.) | No longer employed | NCDS | 0.20 | -0.06 | 0.47 | 10.78 |  |  |  |  |  |
| Full | distressed (cont.) | No longer employed | USOC | 0.30 | 0.14 | 0.47 | 18.86 |  |  |  |  |  |
| Full | distressed (cont.) | No longer employed | ELSA | 0.12 | -0.13 | 0.37 | 11.66 |  |  |  |  |  |
| Full | distressed (cont.) | No longer employed | GS | 1.58 | 0.30 | 2.85 | 0.66 |  |  |  |  |  |
| Full | distressed (cont.) | No longer employed | ALSPAC(G0) | 0.24 | 0.00 | 0.49 | 11.99 |  |  |  |  |  |
| Full | distressed (cont.) | No longer employed | Twins UK | -0.16 | -0.65 | 0.33 | 4.04 |  |  |  |  |  |
| Full | distressed (cont.) | No longer employed | Overall | 0.16 | 0.06 | 0.27 |  | 29.17 |  |  |  |  |
|  |  |  |  |  |  |  |  |  |  | 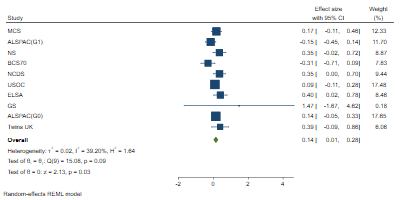 |  |  |
| Full | distressed (cont.) | Stable unemployed | MCS | 0.17 | -0.11 | 0.46 | 12.33 |  |  | REML |  |  |
| Full | distressed (cont.) | Stable unemployed | ALSPAC(G1) | -0.15 | -0.45 | 0.14 | 11.70 |  |  |  |  |  |
| Full | distressed (cont.) | Stable unemployed | NS | 0.35 | -0.02 | 0.72 | 8.87 |  |  |  |  |  |
| Full | distressed (cont.) | Stable unemployed | BCS70 | -0.31 | -0.71 | 0.09 | 7.83 |  |  |  |  |  |
| Full | distressed (cont.) | Stable unemployed | NCDS | 0.35 | 0.00 | 0.70 | 9.44 |  |  |  |  |  |
| Full | distressed (cont.) | Stable unemployed | USOC | 0.09 | -0.11 | 0.28 | 17.48 |  |  |  |  |  |
| Full | distressed (cont.) | Stable unemployed | ELSA | 0.40 | 0.02 | 0.78 | 8.46 |  |  |  |  |  |
| Full | distressed (cont.) | Stable unemployed | GS | 1.47 | -1.67 | 4.62 | 0.18 |  |  |  |  |  |
| Full | distressed (cont.) | Stable unemployed | ALSPAC(G0) | 0.14 | -0.05 | 0.33 | 17.65 |  |  |  |  |  |
| Full | distressed (cont.) | Stable unemployed | Twins UK | 0.39 | -0.09 | 0.86 | 6.06 |  |  |  |  |  |
| Full | distressed (cont.) | Stable unemployed | Overall | 0.14 | 0.01 | 0.28 |  | 39.20 |  |  |  |  |
| Full | less than daily contact | Furloughed | MCS | 0.85 | 0.68 | 1.05 | 4.82 |  |  | REML |  |  |
| Full | less than daily contact | Furloughed | ALSPAC(G1) | 1.53 | 0.86 | 2.74 | 0.77 |  |  |  |  |  |
| Full | less than daily contact | Furloughed | NS | 0.89 | 0.76 | 1.03 | 8.91 |  |  |  |  |  |
| Full | less than daily contact | Furloughed | BCS70 | 0.91 | 0.84 | 0.99 | 18.30 |  |  |  |  | 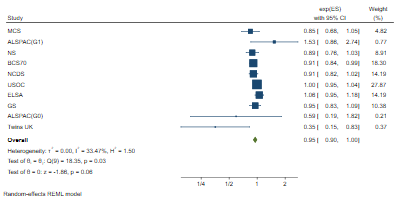 |
| Full | less than daily contact | Furloughed | NCDS | 0.91 | 0.82 | 1.02 | 14.19 |  |  |  |  |  |
| Full | less than daily contact | Furloughed | USOC | 1.00 | 0.95 | 1.04 | 27.87 |  |  |  |  |  |
| Full | less than daily contact | Furloughed | ELSA | 1.06 | 0.95 | 1.18 | 14.19 |  |  |  |  |  |
| Full | less than daily contact | Furloughed | GS | 0.95 | 0.83 | 1.09 | 10.38 |  |  |  |  |  |
| Full | less than daily contact | Furloughed | ALSPAC(G0) | 0.59 | 0.19 | 1.82 | 0.21 |  |  |  |  |  |
| Full | less than daily contact | Furloughed | Twins UK | 0.35 | 0.15 | 0.83 | 0.37 |  |  |  |  |  |
| Full | less than daily contact | Furloughed | Overall | 0.95 | 0.90 | 1.00 |  | 33.47 |  |  |  |  |
|  |  |  |  |  |  |  |  |  |  | 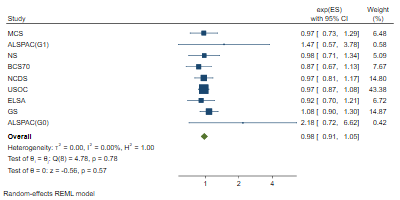 |  |  |
| Full | less than daily contact | No longer employed | MCS | 0.97 | 0.73 | 1.29 | 6.48 |  |  | REML |  |  |
| Full | less than daily contact | No longer employed | ALSPAC(G1) | 1.47 | 0.57 | 3.78 | 0.58 |  |  |  |  |  |
| Full | less than daily contact | No longer employed | NS | 0.98 | 0.71 | 1.34 | 5.09 |  |  |  |  |  |
| Full | less than daily contact | No longer employed | BCS70 | 0.87 | 0.67 | 1.13 | 7.67 |  |  |  |  |  |
| Full | less than daily contact | No longer employed | NCDS | 0.97 | 0.81 | 1.17 | 14.80 |  |  |  |  |  |
| Full | less than daily contact | No longer employed | USOC | 0.97 | 0.87 | 1.08 | 43.38 |  |  |  |  |  |
| Full | less than daily contact | No longer employed | ELSA | 0.92 | 0.70 | 1.21 | 6.72 |  |  |  |  |  |
| Full | less than daily contact | No longer employed | GS | 1.08 | 0.90 | 1.30 | 14.87 |  |  |  |  |  |
| Full | less than daily contact | No longer employed | ALSPAC(G0) | 2.18 | 0.72 | 6.62 | 0.42 |  |  |  |  |  |
| Full | less than daily contact | No longer employed | Twins UK |  |  |  |  |  | low counts |  |  |  |
| Full | less than daily contact | No longer employed | Overall | 0.98 | 0.91 | 1.05 |  | 0.00 |  |  |  |  |
|  |  |  |  |  |  |  |  |  |  | 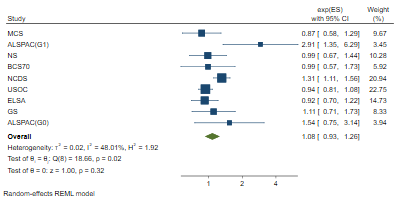 |  |  |
| Full | less than daily contact | Stable unemployed | MCS | 0.87 | 0.58 | 1.29 | 9.67 |  |  | REML |  |  |
| Full | less than daily contact | Stable unemployed | ALSPAC(G1) | 2.91 | 1.35 | 6.29 | 3.45 |  |  |  |  |  |
| Full | less than daily contact | Stable unemployed | NS | 0.99 | 0.67 | 1.44 | 10.28 |  |  |  |  |  |
| Full | less than daily contact | Stable unemployed | BCS70 | 0.99 | 0.57 | 1.73 | 5.92 |  |  |  |  |  |
| Full | less than daily contact | Stable unemployed | NCDS | 1.31 | 1.11 | 1.56 | 20.94 |  |  |  |  |  |
| Full | less than daily contact | Stable unemployed | USOC | 0.94 | 0.81 | 1.08 | 22.75 |  |  |  |  |  |
| Full | less than daily contact | Stable unemployed | ELSA | 0.92 | 0.70 | 1.22 | 14.73 |  |  |  |  |  |
| Full | less than daily contact | Stable unemployed | GS | 1.11 | 0.71 | 1.73 | 8.33 |  |  |  |  |  |
| Full | less than daily contact | Stable unemployed | ALSPAC(G0) | 1.54 | 0.75 | 3.14 | 3.94 |  |  |  |  |  |
| Full | less than daily contact | Stable unemployed | Twins UK |  |  |  |  |  | low counts |  |  |  |
| Full | less than daily contact | Stable unemployed | Overall | 1.08 | 0.93 | 1.26 |  | 48.01 |  |  |  |  |
| Full | fair or poor self-rated health | Furloughed | MCS | 1.35 | 0.60 | 3.05 | 4.17 |  |  | 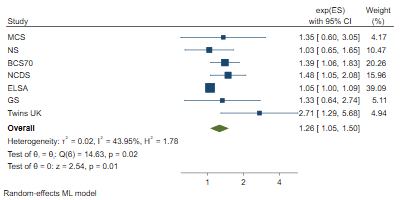 MLE |  |  |
| Full | fair or poor self-rated health | Furloughed | ALSPAC(G1) | |  |  |  |  | no measure |  |  |  |
| Full | fair or poor self-rated health | Furloughed | NS | 1.03 | 0.65 | 1.65 | 10.47 |  |  |  |  |  |
| Full | fair or poor self-rated health | Furloughed | BCS70 | 1.39 | 1.06 | 1.83 | 20.26 |  |  |  |  |  |
| Full | fair or poor self-rated health | Furloughed | NCDS | 1.48 | 1.05 | 2.08 | 15.96 |  |  |  |  |  |
| Full | fair or poor self-rated health | Furloughed | USOC |  |  |  |  |  | no measure |  |  |  |
| Full | fair or poor self-rated health | Furloughed | ELSA | 1.05 | 1.00 | 1.09 | 39.09 |  |  |  |  |  |
| Full | fair or poor self-rated health | Furloughed | GS | 1.33 | 0.64 | 2.74 | 5.11 |  |  |  |  |  |
| Full | fair or poor self-rated health | Furloughed | ALSPAC(G0) | |  |  |  |  | no measure |  |  |  |
| Full | fair or poor self-rated health | Furloughed | Twins UK | 2.71 | 1.29 | 5.68 | 4.94 |  |  |  |  |  |
| Full | fair or poor self-rated health | Furloughed | Overall | 1.26 | 1.05 | 1.50 |  | 43.95 |  |  |  |  |
| Full | fair or poor self-rated health | No longer employed | MCS |  |  |  |  |  | low counts | REML |  |  |
| Full | fair or poor self-rated health | No longer employed | ALSPAC(G1) | |  |  |  |  | no measure |  |  |  |
| Full | fair or poor self-rated health | No longer employed | NS | 3.39 | 1.28 | 9.00 | 10.26 |  |  |  |  |  |
| Full | fair or poor self-rated health | No longer employed | BCS70 | 1.50 | 0.82 | 2.76 | 18.49 |  |  |  |  |  |
| Full | fair or poor self-rated health | No longer employed | NCDS | 1.44 | 0.81 | 2.57 | 19.44 |  |  |  |  |  |
| Full | fair or poor self-rated health | No longer employed | USOC |  |  |  |  |  | no measure |  |  |  |
| Full | fair or poor self-rated health | No longer employed | ELSA | 1.06 | 0.96 | 1.17 | 36.81 |  |  |  |  | 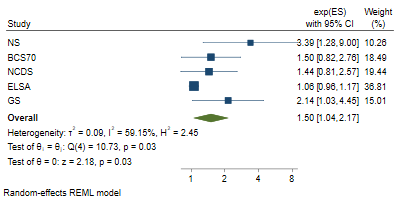 |
| Full | fair or poor self-rated health | No longer employed | GS | 2.14 | 1.03 | 4.45 | 15.01 |  |  |  |  |  |
| Full | fair or poor self-rated health | No longer employed | ALSPAC(G0) | |  |  |  |  | no measure |  |  |  |
| Full | fair or poor self-rated health | No longer employed | Twins UK |  |  |  |  |  | low counts |  |  |  |
| Full | fair or poor self-rated health | No longer employed | Overall | 1.50 | 1.04 | 2.17 |  | 59.15 |  |  |  |  |
| Full | fair or poor self-rated health | Stable unemployed | MCS | 3.39 | 1.25 | 9.22 | 9.82 |  |  | 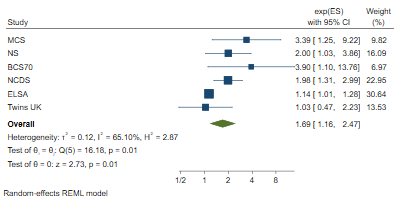 REML |  |  |
| Full | fair or poor self-rated health | Stable unemployed | ALSPAC(G1) | |  |  |  |  | no measure |  |  |  |
| Full | fair or poor self-rated health | Stable unemployed | NS | 2.00 | 1.03 | 3.86 | 16.09 |  |  |  |  |  |
| Full | fair or poor self-rated health | Stable unemployed | BCS70 | 3.90 | 1.10 | 13.76 | 6.97 |  |  |  |  |  |
| Full | fair or poor self-rated health | Stable unemployed | NCDS | 1.98 | 1.31 | 2.99 | 22.95 |  |  |  |  |  |
| Full | fair or poor self-rated health | Stable unemployed | USOC |  |  |  |  |  | no measure |  |  |  |
| Full | fair or poor self-rated health | Stable unemployed | ELSA | 1.14 | 1.01 | 1.28 | 30.64 |  |  |  |  |  |
| Full | fair or poor self-rated health | Stable unemployed | GS |  |  |  |  |  | low counts |  |  |  |
| Full | fair or poor self-rated health | Stable unemployed | ALSPAC(G0) | |  |  |  |  | no measure |  |  |  |
| Full | fair or poor self-rated health | Stable unemployed | Twins UK | 1.03 | 0.47 | 2.23 | 13.53 |  |  |  |  |  |
| Full | fair or poor self-rated health | Stable unemployed | Overall | 1.69 | 1.16 | 2.47 |  | 65.10 |  |  |  |  |

### 4. Subgroup differences

| **Outcome** | **Exposure** | **By gender** | | **By education** | | **By Age** | | **By HH** | |
| --- | --- | --- | --- | --- | --- | --- | --- | --- | --- |
|  |  | chi2 | p-vaue | chi2 | p-vaue | chi2 | p-vaue | chi2 | p-vaue |
| LifeSatBin | Furlough | 0.42 | 0.516 | 1.01 | 0.314 | 0.83 | 0.662 | 0.37 | 0.832 |
|  | No longer employed | 0.00 | 0.968 | 0.11 | 0.738 | 2.73 | 0.255 | 0.67 | 0.714 |
|  | Stable unemployed | 0.92 | 0.336 | 0.43 | 0.510 | 2.88 | 0.237 | 2.03 | 0.362 |
| LonBin | Furlough | 0.11 | 0.746 | 0.03 | 0.860 | 1.00 | 0.605 | 1.20 | 0.548 |
|  | No longer employed | 3.81 | 0.051 | 1.10 | 0.294 | 3.90 | 0.142 | 0.87 | 0.649 |
|  | Stable unemployed | 0.05 | 0.827 | 0.01 | 0.916 | 3.06 | 0.217 | 17.59 | 0.000 |
| LonUCLA | Furlough | 0.19 | 0.666 | 0.79 | 0.375 | 0.80 | 0.670 | 1.07 | 0.586 |
|  | No longer employed | 0.25 | 0.616 | 0.14 | 0.706 | 0.50 | 0.777 | 3.39 | 0.184 |
|  | Stable unemployed | 0.40 | 0.526 | 0.32 | 0.573 | 1.20 | 0.548 | 0.20 | 0.906 |
| PsyDisBin | Furlough | 0.02 | 0.894 | 0.24 | 0.621 | 0.02 | 0.992 | 3.34 | 0.188 |
|  | No longer employed | 1.12 | 0.289 | 0.55 | 0.459 | 1.20 | 0.548 | 3.73 | 0.155 |
|  | Stable unemployed | 1.68 | 0.195 | 1.63 | 0.202 | 0.18 | 0.914 | 2.95 | 0.228 |
| PsyDisCon | Furlough | 0.79 | 0.375 | 0.08 | 0.774 | 0.16 | 0.687 | 1.06 | 0.304 |
|  | No longer employed | 0.08 | 0.779 | 0.22 | 0.642 | 0.12 | 0.732 | 5.17 | 0.075 |
|  | Stable unemployed | 0.41 | 0.520 | 2.59 | 0.107 | 0.14 | 0.707 | 0.85 | 0.355 |
| SCon1 | Furlough | 0.85 | 0.358 | 2.04 | 0.154 | 0.57 | 0.750 | 0.91 | 0.634 |
|  | No longer employed | 1.13 | 0.288 | 0.11 | 0.744 | 0.08 | 0.963 | 1.30 | 0.521 |
|  | Stable unemployed | 1.07 | 0.300 | 0.02 | 0.900 | 0.39 | 0.823 | 1.78 | 0.411 |
| SRHBin | Furlough | 6.81 | 0.009 | 0.00 | 0.977 | 0.13 | 0.936 | 5.03 | 0.081 |
|  | No longer employed | 0.69 | 0.406 | 0.13 | 0.720 | 5.51 | 0.019 | 0.09 | 0.956 |
|  | Stable unemployed | 0.33 | 0.564 | 0.14 | 0.706 | 2.15 | 0.342 | 5.68 | 0.059 |

### 5. Female

| **Adjustment** | **Outcome** | **Exposure** | **Study** | **Coefficient** | **lower_ci** | **upper_ci** | **%Weight** | **%I2** | **Reason for missing** | **Method** | 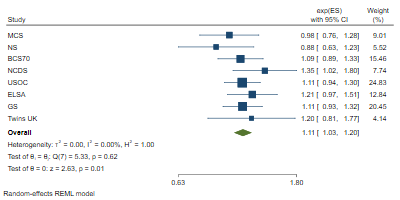 |  |
| --- | --- | --- | --- | --- | --- | --- | --- | --- | --- | --- | --- | --- |
| Female | low life satisfaction | Furloughed | MCS | 0.98 | 0.76 | 1.28 | 9.01 |  |  | REML |  |  |
| Female | low life satisfaction | Furloughed | ALSPAC(G1) | |  |  |  |  | no measure |  |  |  |
| Female | low life satisfaction | Furloughed | NS | 0.88 | 0.63 | 1.23 | 5.52 |  |  |  |  |  |
| Female | low life satisfaction | Furloughed | BCS70 | 1.09 | 0.89 | 1.33 | 15.46 |  |  |  |  |  |
| Female | low life satisfaction | Furloughed | NCDS | 1.35 | 1.02 | 1.80 | 7.74 |  |  |  |  |  |
| Female | low life satisfaction | Furloughed | USOC | 1.11 | 0.94 | 1.30 | 24.83 |  |  |  |  |  |
| Female | low life satisfaction | Furloughed | ELSA | 1.21 | 0.97 | 1.51 | 12.84 |  |  |  |  |  |
| Female | low life satisfaction | Furloughed | GS | 1.11 | 0.93 | 1.32 | 20.45 |  |  |  |  |  |
| Female | low life satisfaction | Furloughed | ALSPAC(G0) | |  |  |  |  | no measure |  |  |  |
| Female | low life satisfaction | Furloughed | Twins UK | 1.20 | 0.81 | 1.77 | 4.14 |  |  |  |  |  |
| Female | low life satisfaction | Furloughed | Overall | 1.11 | 1.03 | 1.20 |  | 0.00 |  |  |  |  |
|  |  |  |  |  |  |  |  |  |  |  | 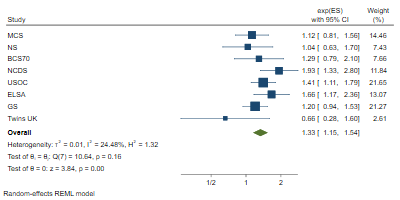 |  |
| Female | low life satisfaction | No longer employed | MCS | 1.12 | 0.81 | 1.56 | 14.46 |  |  | REML |  |  |
| Female | low life satisfaction | No longer employed | ALSPAC(G1) | |  |  |  |  | no measure |  |  |  |
| Female | low life satisfaction | No longer employed | NS | 1.04 | 0.63 | 1.70 | 7.43 |  |  |  |  |  |
| Female | low life satisfaction | No longer employed | BCS70 | 1.29 | 0.79 | 2.10 | 7.66 |  |  |  |  |  |
| Female | low life satisfaction | No longer employed | NCDS | 1.93 | 1.33 | 2.80 | 11.84 |  |  |  |  |  |
| Female | low life satisfaction | No longer employed | USOC | 1.41 | 1.11 | 1.79 | 21.65 |  |  |  |  |  |
| Female | low life satisfaction | No longer employed | ELSA | 1.66 | 1.17 | 2.36 | 13.07 |  |  |  |  |  |
| Female | low life satisfaction | No longer employed | GS | 1.20 | 0.94 | 1.53 | 21.27 |  |  |  |  |  |
| Female | low life satisfaction | No longer employed | ALSPAC(G0) | |  |  |  |  | no measure |  |  |  |
| Female | low life satisfaction | No longer employed | Twins UK | 0.66 | 0.28 | 1.60 | 2.61 |  |  |  |  |  |
| Female | low life satisfaction | No longer employed | Overall | 1.33 | 1.15 | 1.54 |  | 24.48 |  |  |  |  |
|  |  |  |  |  |  |  |  |  |  |  | 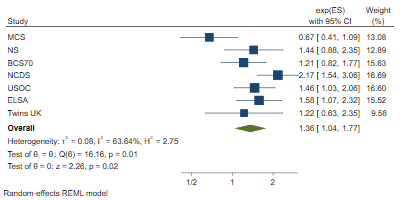 |  |
| Female | low life satisfaction | Stable unemployed | MCS | 0.67 | 0.41 | 1.09 | 13.08 |  |  | REML |  |  |
| Female | low life satisfaction | Stable unemployed | ALSPAC(G1) | |  |  |  |  | no measure |  |  |  |
| Female | low life satisfaction | Stable unemployed | NS | 1.44 | 0.88 | 2.35 | 12.89 |  |  |  |  |  |
| Female | low life satisfaction | Stable unemployed | BCS70 | 1.21 | 0.82 | 1.77 | 15.63 |  |  |  |  |  |
| Female | low life satisfaction | Stable unemployed | NCDS | 2.17 | 1.54 | 3.06 | 16.69 |  |  |  |  |  |
| Female | low life satisfaction | Stable unemployed | USOC | 1.46 | 1.03 | 2.06 | 16.60 |  |  |  |  |  |
| Female | low life satisfaction | Stable unemployed | ELSA | 1.58 | 1.07 | 2.32 | 15.52 |  |  |  |  |  |
| Female | low life satisfaction | Stable unemployed | GS |  |  |  |  |  | low counts |  |  |  |
| Female | low life satisfaction | Stable unemployed | ALSPAC(G0) | |  |  |  |  | no measure |  |  |  |
| Female | low life satisfaction | Stable unemployed | Twins UK | 1.22 | 0.63 | 2.35 | 9.58 |  |  |  |  |  |
| Female | low life satisfaction | Stable unemployed | Overall | 1.36 | 1.04 | 1.77 |  | 63.64 |  |  |  |  |
|  |  |  |  |  |  |  |  |  |  |  | 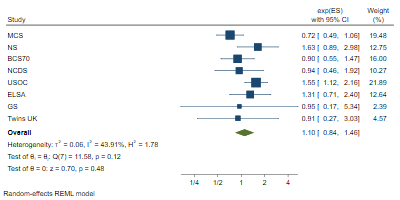 |  |
| Female | often lonely | Furloughed | MCS | 0.72 | 0.49 | 1.06 | 19.48 |  |  | REML |  |  |
| Female | often lonely | Furloughed | ALSPAC(G1) | |  |  |  |  | no measure |  |  |  |
| Female | often lonely | Furloughed | NS | 1.63 | 0.89 | 2.98 | 12.75 |  |  |  |  |  |
| Female | often lonely | Furloughed | BCS70 | 0.90 | 0.55 | 1.47 | 16.00 |  |  |  |  |  |
| Female | often lonely | Furloughed | NCDS | 0.94 | 0.46 | 1.92 | 10.27 |  |  |  |  |  |
| Female | often lonely | Furloughed | USOC | 1.55 | 1.12 | 2.16 | 21.89 |  |  |  |  |  |
| Female | often lonely | Furloughed | ELSA | 1.31 | 0.71 | 2.40 | 12.64 |  |  |  |  |  |
| Female | often lonely | Furloughed | GS | 0.95 | 0.17 | 5.34 | 2.39 |  |  |  |  |  |
| Female | often lonely | Furloughed | ALSPAC(G0) | |  |  |  |  | no measure |  |  |  |
| Female | often lonely | Furloughed | Twins UK | 0.91 | 0.27 | 3.03 | 4.57 |  |  |  |  |  |
| Female | often lonely | Furloughed | Overall | 1.10 | 0.84 | 1.46 |  | 43.91 |  |  |  |  |
|  |  |  |  |  |  |  |  |  |  |  | 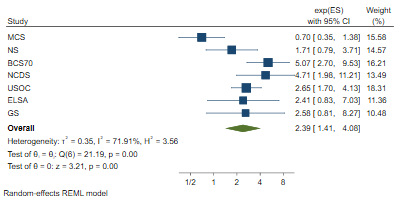 |  |
| Female | often lonely | No longer employed | MCS | 0.70 | 0.35 | 1.38 | 15.58 |  |  | REML |  |  |
| Female | often lonely | No longer employed | ALSPAC(G1) | |  |  |  |  | no measure |  |  |  |
| Female | often lonely | No longer employed | NS | 1.71 | 0.79 | 3.71 | 14.57 |  |  |  |  |  |
| Female | often lonely | No longer employed | BCS70 | 5.07 | 2.70 | 9.53 | 16.21 |  |  |  |  |  |
| Female | often lonely | No longer employed | NCDS | 4.71 | 1.98 | 11.21 | 13.49 |  |  |  |  |  |
| Female | often lonely | No longer employed | USOC | 2.65 | 1.70 | 4.13 | 18.31 |  |  |  |  |  |
| Female | often lonely | No longer employed | ELSA | 2.41 | 0.83 | 7.03 | 11.36 |  |  |  |  |  |
| Female | often lonely | No longer employed | GS | 2.58 | 0.81 | 8.27 | 10.48 |  |  |  |  |  |
| Female | often lonely | No longer employed | ALSPAC(G0) | |  |  |  |  | no measure |  |  |  |
| Female | often lonely | No longer employed | Twins UK |  |  |  |  |  | low counts |  |  |  |
| Female | often lonely | No longer employed | Overall | 2.39 | 1.41 | 4.08 |  | 71.91 |  |  |  |  |
|  |  |  |  |  |  |  |  |  |  |  | 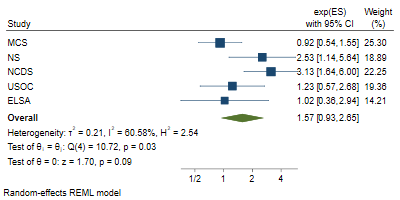 |  |
| Female | often lonely | Stable unemployed | MCS | 0.92 | 0.54 | 1.55 | 25.30 |  |  | REML |  |  |
| Female | often lonely | Stable unemployed | ALSPAC(G1) | |  |  |  |  | no measure |  |  |  |
| Female | often lonely | Stable unemployed | NS | 2.53 | 1.14 | 5.64 | 18.89 |  |  |  |  |  |
| Female | often lonely | Stable unemployed | BCS70 |  |  |  |  |  | low counts |  |  |  |
| Female | often lonely | Stable unemployed | NCDS | 3.13 | 1.64 | 6.00 | 22.25 |  |  |  |  |  |
| Female | often lonely | Stable unemployed | USOC | 1.23 | 0.57 | 2.68 | 19.36 |  |  |  |  |  |
| Female | often lonely | Stable unemployed | ELSA | 1.02 | 0.36 | 2.94 | 14.21 |  |  |  |  |  |
| Female | often lonely | Stable unemployed | GS |  |  |  |  |  | low counts |  |  |  |
| Female | often lonely | Stable unemployed | ALSPAC(G0) | |  |  |  |  | no measure |  |  |  |
| Female | often lonely | Stable unemployed | Twins UK |  |  |  |  |  | low counts |  |  |  |
| Female | often lonely | Stable unemployed | Overall | 1.57 | 0.93 | 2.65 |  | 60.58 |  |  |  |  |
|  |  |  |  |  |  |  |  |  |  |  | 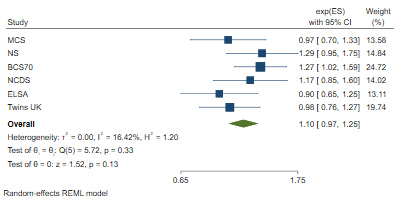 |  |
| Female | high loneliness | Furloughed | MCS | 0.97 | 0.70 | 1.33 | 13.58 |  |  | REML |  |  |
| Female | high loneliness | Furloughed | ALSPAC(G1) | |  |  |  |  | no measure |  |  |  |
| Female | high loneliness | Furloughed | NS | 1.29 | 0.95 | 1.75 | 14.84 |  |  |  |  |  |
| Female | high loneliness | Furloughed | BCS70 | 1.27 | 1.02 | 1.59 | 24.72 |  |  |  |  |  |
| Female | high loneliness | Furloughed | NCDS | 1.17 | 0.85 | 1.60 | 14.02 |  |  |  |  |  |
| Female | high loneliness | Furloughed | USOC |  |  |  |  |  | no measure |  |  |  |
| Female | high loneliness | Furloughed | ELSA | 0.90 | 0.65 | 1.25 | 13.11 |  |  |  |  |  |
| Female | high loneliness | Furloughed | GS |  |  |  |  |  | no measure |  |  |  |
| Female | high loneliness | Furloughed | ALSPAC(G0) | |  |  |  |  | no measure |  |  |  |
| Female | high loneliness | Furloughed | Twins UK | 0.98 | 0.76 | 1.27 | 19.74 |  |  |  |  |  |
| Female | high loneliness | Furloughed | Overall | 1.10 | 0.97 | 1.25 |  | 16.42 |  |  |  |  |
|  |  |  |  |  |  |  |  |  |  |  | 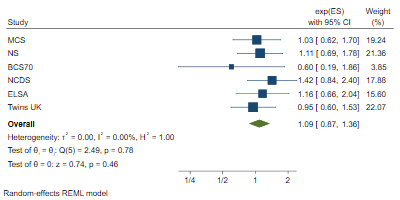 |  |
| Female | high loneliness | No longer employed | MCS | 1.03 | 0.62 | 1.70 | 19.24 |  |  | REML |  |  |
| Female | high loneliness | No longer employed | ALSPAC(G1) | |  |  |  |  | no measure |  |  |  |
| Female | high loneliness | No longer employed | NS | 1.11 | 0.69 | 1.78 | 21.36 |  |  |  |  |  |
| Female | high loneliness | No longer employed | BCS70 | 0.60 | 0.19 | 1.86 | 3.85 |  |  |  |  |  |
| Female | high loneliness | No longer employed | NCDS | 1.42 | 0.84 | 2.40 | 17.88 |  |  |  |  |  |
| Female | high loneliness | No longer employed | USOC |  |  |  |  |  | no measure |  |  |  |
| Female | high loneliness | No longer employed | ELSA | 1.16 | 0.66 | 2.04 | 15.60 |  |  |  |  |  |
| Female | high loneliness | No longer employed | GS |  |  |  |  |  | no measure |  |  |  |
| Female | high loneliness | No longer employed | ALSPAC(G0) | |  |  |  |  | no measure |  |  |  |
| Female | high loneliness | No longer employed | Twins UK | 0.95 | 0.60 | 1.53 | 22.07 |  |  |  |  |  |
| Female | high loneliness | No longer employed | Overall | 1.09 | 0.87 | 1.36 |  | 0.00 |  |  |  |  |
|  |  |  |  |  |  |  |  |  |  |  | 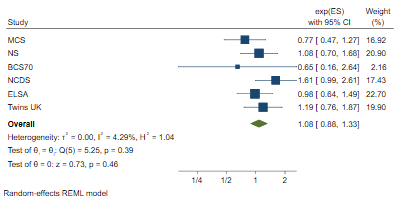 |  |
| Female | high loneliness | Stable unemployed | MCS | 0.77 | 0.47 | 1.27 | 16.92 |  |  | REML |  |  |
| Female | high loneliness | Stable unemployed | ALSPAC(G1) | |  |  |  |  | no measure |  |  |  |
| Female | high loneliness | Stable unemployed | NS | 1.08 | 0.70 | 1.68 | 20.90 |  |  |  |  |  |
| Female | high loneliness | Stable unemployed | BCS70 | 0.65 | 0.16 | 2.64 | 2.16 |  |  |  |  |  |
| Female | high loneliness | Stable unemployed | NCDS | 1.61 | 0.99 | 2.61 | 17.43 |  |  |  |  |  |
| Female | high loneliness | Stable unemployed | USOC |  |  |  |  |  | no measure |  |  |  |
| Female | high loneliness | Stable unemployed | ELSA | 0.98 | 0.64 | 1.49 | 22.70 |  |  |  |  |  |
| Female | high loneliness | Stable unemployed | GS |  |  |  |  |  | no measure |  |  |  |
| Female | high loneliness | Stable unemployed | ALSPAC(G0) | |  |  |  |  | no measure |  |  |  |
| Female | high loneliness | Stable unemployed | Twins UK | 1.19 | 0.76 | 1.87 | 19.90 |  |  |  |  |  |
| Female | high loneliness | Stable unemployed | Overall | 1.08 | 0.88 | 1.33 |  | 4.29 |  |  |  |  |
|  |  |  |  |  |  |  |  |  |  |  | 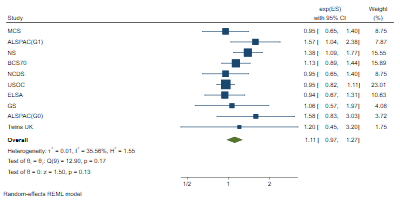 |  |
| Female | distressed (bin.) | Furloughed | MCS | 0.95 | 0.65 | 1.40 | 8.75 |  |  | REML |  |  |
| Female | distressed (bin.) | Furloughed | ALSPAC(G1) | 1.57 | 1.04 | 2.38 | 7.87 |  |  |  |  |  |
| Female | distressed (bin.) | Furloughed | NS | 1.38 | 1.09 | 1.77 | 15.55 |  |  |  |  |  |
| Female | distressed (bin.) | Furloughed | BCS70 | 1.13 | 0.89 | 1.44 | 15.89 |  |  |  |  |  |
| Female | distressed (bin.) | Furloughed | NCDS | 0.95 | 0.65 | 1.40 | 8.75 |  |  |  |  |  |
| Female | distressed (bin.) | Furloughed | USOC | 0.95 | 0.82 | 1.11 | 23.01 |  |  |  |  |  |
| Female | distressed (bin.) | Furloughed | ELSA | 0.94 | 0.67 | 1.31 | 10.63 |  |  |  |  |  |
| Female | distressed (bin.) | Furloughed | GS | 1.06 | 0.57 | 1.97 | 4.08 |  |  |  |  |  |
| Female | distressed (bin.) | Furloughed | ALSPAC(G0) | 1.58 | 0.83 | 3.03 | 3.72 |  |  |  |  |  |
| Female | distressed (bin.) | Furloughed | Twins UK | 1.20 | 0.45 | 3.20 | 1.75 |  |  |  |  |  |
| Female | distressed (bin.) | Furloughed | Overall | 1.11 | 0.97 | 1.27 |  | 35.56 |  |  |  |  |
|  |  |  |  |  |  |  |  |  |  |  | 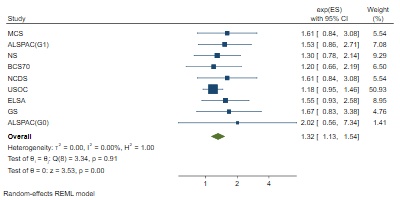 |  |
| Female | distressed (bin.) | No longer employed | MCS | 1.61 | 0.84 | 3.08 | 5.54 |  |  | REML |  |  |
| Female | distressed (bin.) | No longer employed | ALSPAC(G1) | 1.53 | 0.86 | 2.71 | 7.08 |  |  |  |  |  |
| Female | distressed (bin.) | No longer employed | NS | 1.30 | 0.78 | 2.14 | 9.29 |  |  |  |  |  |
| Female | distressed (bin.) | No longer employed | BCS70 | 1.20 | 0.66 | 2.19 | 6.50 |  |  |  |  |  |
| Female | distressed (bin.) | No longer employed | NCDS | 1.61 | 0.84 | 3.08 | 5.54 |  |  |  |  |  |
| Female | distressed (bin.) | No longer employed | USOC | 1.18 | 0.95 | 1.46 | 50.93 |  |  |  |  |  |
| Female | distressed (bin.) | No longer employed | ELSA | 1.55 | 0.93 | 2.58 | 8.95 |  |  |  |  |  |
| Female | distressed (bin.) | No longer employed | GS | 1.67 | 0.83 | 3.38 | 4.76 |  |  |  |  |  |
| Female | distressed (bin.) | No longer employed | ALSPAC(G0) | 2.02 | 0.56 | 7.34 | 1.41 |  |  |  |  |  |
| Female | distressed (bin.) | No longer employed | Twins UK |  |  |  |  |  | low counts |  |  |  |
| Female | distressed (bin.) | No longer employed | Overall | 1.32 | 1.13 | 1.54 |  | 0.00 |  |  |  |  |
| Female | distressed (bin.) | Stable unemployed | MCS | 2.31 | 0.86 | 6.17 | 3.79 |  |  | REML |  |  |
| Female | distressed (bin.) | Stable unemployed | ALSPAC(G1) | 1.60 | 0.84 | 3.04 | 8.87 |  |  |  |  | 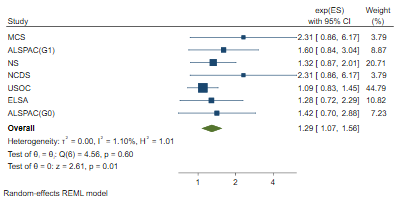 |
| Female | distressed (bin.) | Stable unemployed | NS | 1.32 | 0.87 | 2.01 | 20.71 |  |  |  |  |  |
| Female | distressed (bin.) | Stable unemployed | BCS70 |  |  |  |  |  | low counts |  |  |  |
| Female | distressed (bin.) | Stable unemployed | NCDS | 2.31 | 0.86 | 6.17 | 3.79 |  |  |  |  |  |
| Female | distressed (bin.) | Stable unemployed | USOC | 1.09 | 0.83 | 1.45 | 44.79 |  |  |  |  |  |
| Female | distressed (bin.) | Stable unemployed | ELSA | 1.28 | 0.72 | 2.29 | 10.82 |  |  |  |  |  |
| Female | distressed (bin.) | Stable unemployed | GS |  |  |  |  |  | low counts |  |  |  |
| Female | distressed (bin.) | Stable unemployed | ALSPAC(G0) | 1.42 | 0.70 | 2.88 | 7.23 |  |  |  |  |  |
| Female | distressed (bin.) | Stable unemployed | Twins UK |  |  |  |  |  | low counts |  |  |  |
| Female | distressed (bin.) | Stable unemployed | Overall | 1.29 | 1.07 | 1.56 |  | 1.10 |  |  |  |  |
|  |  |  |  |  |  |  |  |  |  |  | 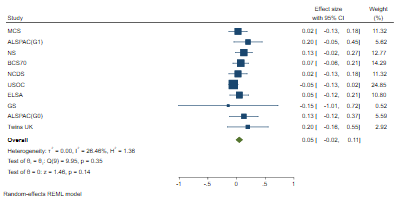 |  |
| Female | distressed (cont.) | Furloughed | MCS | 0.02 | -0.13 | 0.18 | 11.32 |  |  | REML |  |  |
| Female | distressed (cont.) | Furloughed | ALSPAC(G1) | 0.20 | -0.05 | 0.45 | 5.62 |  |  |  |  |  |
| Female | distressed (cont.) | Furloughed | NS | 0.13 | -0.02 | 0.27 | 12.77 |  |  |  |  |  |
| Female | distressed (cont.) | Furloughed | BCS70 | 0.07 | -0.06 | 0.21 | 14.29 |  |  |  |  |  |
| Female | distressed (cont.) | Furloughed | NCDS | 0.02 | -0.13 | 0.18 | 11.32 |  |  |  |  |  |
| Female | distressed (cont.) | Furloughed | USOC | -0.05 | -0.13 | 0.02 | 24.85 |  |  |  |  |  |
| Female | distressed (cont.) | Furloughed | ELSA | 0.05 | -0.12 | 0.21 | 10.80 |  |  |  |  |  |
| Female | distressed (cont.) | Furloughed | GS | -0.15 | -1.01 | 0.72 | 0.52 |  |  |  |  |  |
| Female | distressed (cont.) | Furloughed | ALSPAC(G0) | 0.13 | -0.12 | 0.37 | 5.59 |  |  |  |  |  |
| Female | distressed (cont.) | Furloughed | Twins UK | 0.20 | -0.16 | 0.55 | 2.92 |  |  |  |  |  |
| Female | distressed (cont.) | Furloughed | Overall | 0.05 | -0.02 | 0.11 |  | 26.46 |  |  |  |  |
|  |  |  |  |  |  |  |  |  |  |  | 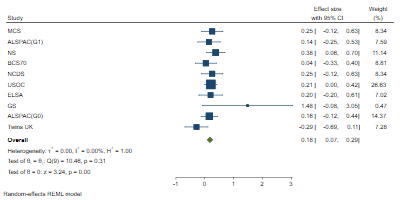 |  |
| Female | distressed (cont.) | No longer employed | MCS | 0.25 | -0.12 | 0.63 | 8.34 |  |  | REML |  |  |
| Female | distressed (cont.) | No longer employed | ALSPAC(G1) | 0.14 | -0.25 | 0.53 | 7.59 |  |  |  |  |  |
| Female | distressed (cont.) | No longer employed | NS | 0.38 | 0.06 | 0.70 | 11.14 |  |  |  |  |  |
| Female | distressed (cont.) | No longer employed | BCS70 | 0.04 | -0.33 | 0.40 | 8.81 |  |  |  |  |  |
| Female | distressed (cont.) | No longer employed | NCDS | 0.25 | -0.12 | 0.63 | 8.34 |  |  |  |  |  |
| Female | distressed (cont.) | No longer employed | USOC | 0.21 | 0.00 | 0.42 | 26.63 |  |  |  |  |  |
| Female | distressed (cont.) | No longer employed | ELSA | 0.20 | -0.20 | 0.61 | 7.02 |  |  |  |  |  |
| Female | distressed (cont.) | No longer employed | GS | 1.48 | -0.08 | 3.05 | 0.47 |  |  |  |  |  |
| Female | distressed (cont.) | No longer employed | ALSPAC(G0) | 0.16 | -0.12 | 0.44 | 14.37 |  |  |  |  |  |
| Female | distressed (cont.) | No longer employed | Twins UK | -0.29 | -0.69 | 0.11 | 7.28 |  |  |  |  |  |
| Female | distressed (cont.) | No longer employed | Overall | 0.18 | 0.07 | 0.29 |  | 0.00 |  |  |  |  |
|  |  |  |  |  |  |  |  |  |  |  | 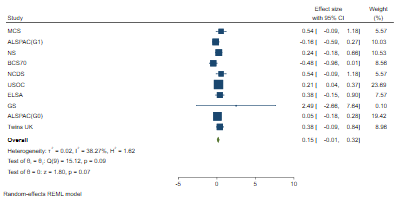 |  |
| Female | distressed (cont.) | Stable unemployed | MCS | 0.54 | -0.09 | 1.18 | 5.57 |  |  | REML |  |  |
| Female | distressed (cont.) | Stable unemployed | ALSPAC(G1) | -0.16 | -0.59 | 0.27 | 10.03 |  |  |  |  |  |
| Female | distressed (cont.) | Stable unemployed | NS | 0.24 | -0.18 | 0.66 | 10.53 |  |  |  |  |  |
| Female | distressed (cont.) | Stable unemployed | BCS70 | -0.48 | -0.96 | 0.01 | 8.56 |  |  |  |  |  |
| Female | distressed (cont.) | Stable unemployed | NCDS | 0.54 | -0.09 | 1.18 | 5.57 |  |  |  |  |  |
| Female | distressed (cont.) | Stable unemployed | USOC | 0.21 | 0.04 | 0.37 | 23.69 |  |  |  |  |  |
| Female | distressed (cont.) | Stable unemployed | ELSA | 0.38 | -0.15 | 0.90 | 7.57 |  |  |  |  |  |
| Female | distressed (cont.) | Stable unemployed | GS | 2.49 | -2.66 | 7.64 | 0.10 |  |  |  |  |  |
| Female | distressed (cont.) | Stable unemployed | ALSPAC(G0) | 0.05 | -0.18 | 0.28 | 19.42 |  |  |  |  |  |
| Female | distressed (cont.) | Stable unemployed | Twins UK | 0.38 | -0.09 | 0.84 | 8.96 |  |  |  |  |  |
| Female | distressed (cont.) | Stable unemployed | Overall | 0.15 | -0.01 | 0.32 |  | 38.27 |  |  |  |  |
|  |  |  |  |  |  |  |  |  |  |  | 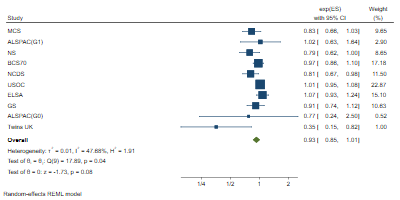 |  |
| Female | less than daily contact | Furloughed | MCS | 0.83 | 0.66 | 1.03 | 9.65 |  |  | REML |  |  |
| Female | less than daily contact | Furloughed | ALSPAC(G1) | 1.02 | 0.63 | 1.64 | 2.90 |  |  |  |  |  |
| Female | less than daily contact | Furloughed | NS | 0.79 | 0.62 | 1.00 | 8.65 |  |  |  |  |  |
| Female | less than daily contact | Furloughed | BCS70 | 0.97 | 0.86 | 1.10 | 17.18 |  |  |  |  |  |
| Female | less than daily contact | Furloughed | NCDS | 0.81 | 0.67 | 0.98 | 11.50 |  |  |  |  |  |
| Female | less than daily contact | Furloughed | USOC | 1.01 | 0.95 | 1.08 | 22.87 |  |  |  |  |  |
| Female | less than daily contact | Furloughed | ELSA | 1.07 | 0.93 | 1.24 | 15.10 |  |  |  |  |  |
| Female | less than daily contact | Furloughed | GS | 0.91 | 0.74 | 1.12 | 10.63 |  |  |  |  |  |
| Female | less than daily contact | Furloughed | ALSPAC(G0) | 0.77 | 0.24 | 2.50 | 0.52 |  |  |  |  |  |
| Female | less than daily contact | Furloughed | Twins UK | 0.35 | 0.15 | 0.82 | 1.00 |  |  |  |  |  |
| Female | less than daily contact | Furloughed | Overall | 0.93 | 0.85 | 1.01 |  | 47.68 |  |  |  |  |
|  |  |  |  |  |  |  |  |  |  |  | 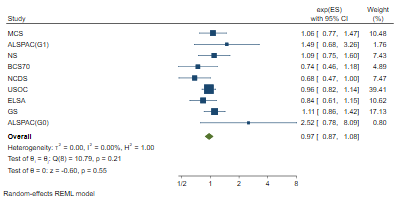 |  |
| Female | less than daily contact | No longer employed | MCS | 1.06 | 0.77 | 1.47 | 10.48 |  |  | REML |  |  |
| Female | less than daily contact | No longer employed | ALSPAC(G1) | 1.49 | 0.68 | 3.26 | 1.76 |  |  |  |  |  |
| Female | less than daily contact | No longer employed | NS | 1.09 | 0.75 | 1.60 | 7.43 |  |  |  |  |  |
| Female | less than daily contact | No longer employed | BCS70 | 0.74 | 0.46 | 1.18 | 4.89 |  |  |  |  |  |
| Female | less than daily contact | No longer employed | NCDS | 0.68 | 0.47 | 1.00 | 7.47 |  |  |  |  |  |
| Female | less than daily contact | No longer employed | USOC | 0.96 | 0.82 | 1.14 | 39.41 |  |  |  |  |  |
| Female | less than daily contact | No longer employed | ELSA | 0.84 | 0.61 | 1.15 | 10.62 |  |  |  |  |  |
| Female | less than daily contact | No longer employed | GS | 1.11 | 0.86 | 1.42 | 17.13 |  |  |  |  |  |
| Female | less than daily contact | No longer employed | ALSPAC(G0) | 2.52 | 0.78 | 8.09 | 0.80 |  |  |  |  |  |
| Female | less than daily contact | No longer employed | Twins UK |  |  |  |  |  | low counts |  |  |  |
| Female | less than daily contact | No longer employed | Overall | 0.97 | 0.87 | 1.08 |  | 0.00 |  |  |  |  |
|  |  |  |  |  |  |  |  |  |  |  | 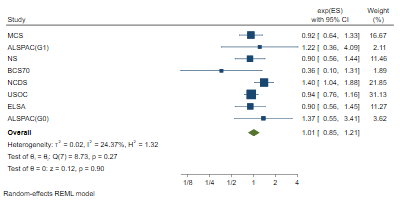 |  |
| Female | less than daily contact | Stable unemployed | MCS | 0.92 | 0.64 | 1.33 | 16.67 |  |  | REML |  |  |
| Female | less than daily contact | Stable unemployed | ALSPAC(G1) | 1.22 | 0.36 | 4.09 | 2.11 |  |  |  |  |  |
| Female | less than daily contact | Stable unemployed | NS | 0.90 | 0.56 | 1.44 | 11.46 |  |  |  |  |  |
| Female | less than daily contact | Stable unemployed | BCS70 | 0.36 | 0.10 | 1.31 | 1.89 |  |  |  |  |  |
| Female | less than daily contact | Stable unemployed | NCDS | 1.40 | 1.04 | 1.88 | 21.85 |  |  |  |  |  |
| Female | less than daily contact | Stable unemployed | USOC | 0.94 | 0.76 | 1.16 | 31.13 |  |  |  |  |  |
| Female | less than daily contact | Stable unemployed | ELSA | 0.90 | 0.56 | 1.45 | 11.27 |  |  |  |  |  |
| Female | less than daily contact | Stable unemployed | GS |  |  |  |  |  | low counts |  |  |  |
| Female | less than daily contact | Stable unemployed | ALSPAC(G0) | 1.37 | 0.55 | 3.41 | 3.62 |  |  |  |  |  |
| Female | less than daily contact | Stable unemployed | Twins UK |  |  |  |  |  | low counts |  |  |  |
| Female | less than daily contact | Stable unemployed | Overall | 1.01 | 0.85 | 1.21 |  | 24.37 |  |  |  |  |
| Female | fair or poor self-rated health | Furloughed | MCS | 2.39 | 0.94 | 6.09 | 5.55 |  |  | MLE | 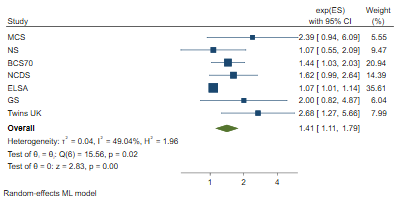 |  |
| Female | fair or poor self-rated health | Furloughed | ALSPAC(G1) | |  |  |  |  | no measure |  |  |  |
| Female | fair or poor self-rated health | Furloughed | NS | 1.07 | 0.55 | 2.09 | 9.47 |  |  |  |  |  |
| Female | fair or poor self-rated health | Furloughed | BCS70 | 1.44 | 1.03 | 2.03 | 20.94 |  |  |  |  |  |
| Female | fair or poor self-rated health | Furloughed | NCDS | 1.62 | 0.99 | 2.64 | 14.39 |  |  |  |  |  |
| Female | fair or poor self-rated health | Furloughed | USOC |  |  |  |  |  | no measure |  |  |  |
| Female | fair or poor self-rated health | Furloughed | ELSA | 1.07 | 1.01 | 1.14 | 35.61 |  |  |  |  |  |
| Female | fair or poor self-rated health | Furloughed | GS | 2.00 | 0.82 | 4.87 | 6.04 |  |  |  |  |  |
| Female | fair or poor self-rated health | Furloughed | ALSPAC(G0) | |  |  |  |  | no measure |  |  |  |
| Female | fair or poor self-rated health | Furloughed | Twins UK | 2.68 | 1.27 | 5.66 | 7.99 |  |  |  |  |  |
| Female | fair or poor self-rated health | Furloughed | Overall | 1.41 | 1.11 | 1.79 |  | 49.04 |  |  |  |  |
| Female | fair or poor self-rated health | No longer employed | MCS |  |  |  |  |  | low counts | REML |  |  |
| Female | fair or poor self-rated health | No longer employed | ALSPAC(G1) | |  |  |  |  | no measure |  |  |  |
| Female | fair or poor self-rated health | No longer employed | NS | 4.37 | 1.65 | 11.57 | 15.48 |  |  |  |  |  |
| Female | fair or poor self-rated health | No longer employed | BCS70 | 1.23 | 0.54 | 2.82 | 18.06 |  |  |  |  |  |
| Female | fair or poor self-rated health | No longer employed | NCDS | 0.90 | 0.41 | 1.96 | 18.93 |  |  |  |  |  |
| Female | fair or poor self-rated health | No longer employed | USOC |  |  |  |  |  | no measure |  |  |  |
| Female | fair or poor self-rated health | No longer employed | ELSA | 1.00 | 0.87 | 1.15 | 31.25 |  |  |  |  | 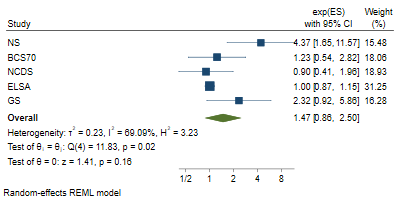 |
| Female | fair or poor self-rated health | No longer employed | GS | 2.32 | 0.92 | 5.86 | 16.28 |  |  |  |  |  |
| Female | fair or poor self-rated health | No longer employed | ALSPAC(G0) | |  |  |  |  | no measure |  |  |  |
| Female | fair or poor self-rated health | No longer employed | Twins UK |  |  |  |  |  | low counts |  |  |  |
| Female | fair or poor self-rated health | No longer employed | Overall | 1.47 | 0.86 | 2.50 |  | 69.09 |  |  |  |  |
| Female | fair or poor self-rated health | Stable unemployed | MCS | 1.64 | 0.60 | 4.48 | 10.45 |  |  | REML | 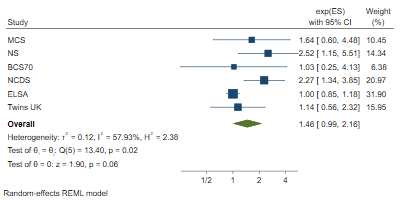 |  |
| Female | fair or poor self-rated health | Stable unemployed | ALSPAC(G1) | |  |  |  |  | no measure |  |  |  |
| Female | fair or poor self-rated health | Stable unemployed | NS | 2.52 | 1.15 | 5.51 | 14.34 |  |  |  |  |  |
| Female | fair or poor self-rated health | Stable unemployed | BCS70 | 1.03 | 0.25 | 4.13 | 6.38 |  |  |  |  |  |
| Female | fair or poor self-rated health | Stable unemployed | NCDS | 2.27 | 1.34 | 3.85 | 20.97 |  |  |  |  |  |
| Female | fair or poor self-rated health | Stable unemployed | USOC |  |  |  |  |  | no measure |  |  |  |
| Female | fair or poor self-rated health | Stable unemployed | ELSA | 1.00 | 0.85 | 1.18 | 31.90 |  |  |  |  |  |
| Female | fair or poor self-rated health | Stable unemployed | GS |  |  |  |  |  | low counts |  |  |  |
| Female | fair or poor self-rated health | Stable unemployed | ALSPAC(G0) | |  |  |  |  | no measure |  |  |  |
| Female | fair or poor self-rated health | Stable unemployed | Twins UK | 1.14 | 0.56 | 2.32 | 15.95 |  |  |  |  |  |
| Female | fair or poor self-rated health | Stable unemployed | Overall | 1.46 | 0.99 | 2.16 |  | 57.93 |  |  |  |  |

### 6. Male

| **Adjustment** | **Outcome** | **Exposure** | **Study** | **Coefficient** | **lower_ci** | **upper_ci** | **%Weight** | **%I2** | **Reason for missing** | **Method** | 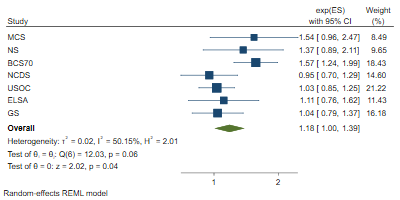 |
| --- | --- | --- | --- | --- | --- | --- | --- | --- | --- | --- | --- |
| Male | low life satisfaction | Furloughed | MCS | 1.54 | 0.96 | 2.47 | 8.49 |  |  | REML |  |
| Male | low life satisfaction | Furloughed | ALSPAC(G1) | |  |  |  |  | no measure |  |  |
| Male | low life satisfaction | Furloughed | NS | 1.37 | 0.89 | 2.11 | 9.65 |  |  |  |  |
| Male | low life satisfaction | Furloughed | BCS70 | 1.57 | 1.24 | 1.99 | 18.43 |  |  |  |  |
| Male | low life satisfaction | Furloughed | NCDS | 0.95 | 0.70 | 1.29 | 14.60 |  |  |  |  |
| Male | low life satisfaction | Furloughed | USOC | 1.03 | 0.85 | 1.25 | 21.22 |  |  |  |  |
| Male | low life satisfaction | Furloughed | ELSA | 1.11 | 0.76 | 1.62 | 11.43 |  |  |  |  |
| Male | low life satisfaction | Furloughed | GS | 1.04 | 0.79 | 1.37 | 16.18 |  |  |  |  |
| Male | low life satisfaction | Furloughed | ALSPAC(G0) | |  |  |  |  | no measure |  |  |
| Male | low life satisfaction | Furloughed | Twins UK |  |  |  |  |  | low counts |  |  |
| Male | low life satisfaction | Furloughed | Overall | 1.18 | 1.00 | 1.39 |  | 50.15 |  |  |  |
|  |  |  |  |  |  |  |  |  |  |  | 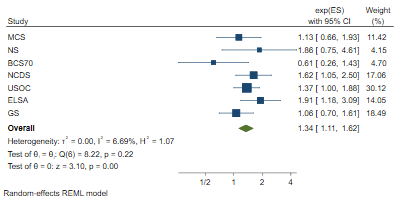 |
| Male | low life satisfaction | No longer employed | MCS | 1.13 | 0.66 | 1.93 | 11.42 |  |  | REML |  |
| Male | low life satisfaction | No longer employed | ALSPAC(G1) | |  |  |  |  | no measure |  |  |
| Male | low life satisfaction | No longer employed | NS | 1.86 | 0.75 | 4.61 | 4.15 |  |  |  |  |
| Male | low life satisfaction | No longer employed | BCS70 | 0.61 | 0.26 | 1.43 | 4.70 |  |  |  |  |
| Male | low life satisfaction | No longer employed | NCDS | 1.62 | 1.05 | 2.50 | 17.06 |  |  |  |  |
| Male | low life satisfaction | No longer employed | USOC | 1.37 | 1.00 | 1.88 | 30.12 |  |  |  |  |
| Male | low life satisfaction | No longer employed | ELSA | 1.91 | 1.18 | 3.09 | 14.05 |  |  |  |  |
| Male | low life satisfaction | No longer employed | GS | 1.06 | 0.70 | 1.61 | 18.49 |  |  |  |  |
| Male | low life satisfaction | No longer employed | ALSPAC(G0) | |  |  |  |  | no measure |  |  |
| Male | low life satisfaction | No longer employed | Twins UK |  |  |  |  |  | low counts |  |  |
| Male | low life satisfaction | No longer employed | Overall | 1.34 | 1.11 | 1.62 |  | 6.69 |  |  |  |
|  |  |  |  |  |  |  |  |  |  |  | 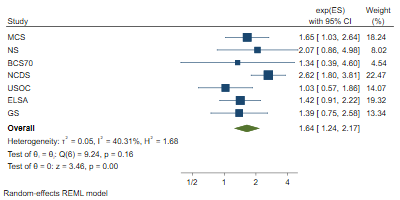 |
| Male | low life satisfaction | Stable unemployed | MCS | 1.65 | 1.03 | 2.64 | 18.24 |  |  | REML |  |
| Male | low life satisfaction | Stable unemployed | ALSPAC(G1) | |  |  |  |  | no measure |  |  |
| Male | low life satisfaction | Stable unemployed | NS | 2.07 | 0.86 | 4.98 | 8.02 |  |  |  |  |
| Male | low life satisfaction | Stable unemployed | BCS70 | 1.34 | 0.39 | 4.60 | 4.54 |  |  |  |  |
| Male | low life satisfaction | Stable unemployed | NCDS | 2.62 | 1.80 | 3.81 | 22.47 |  |  |  |  |
| Male | low life satisfaction | Stable unemployed | USOC | 1.03 | 0.57 | 1.86 | 14.07 |  |  |  |  |
| Male | low life satisfaction | Stable unemployed | ELSA | 1.42 | 0.91 | 2.22 | 19.32 |  |  |  |  |
| Male | low life satisfaction | Stable unemployed | GS | 1.39 | 0.75 | 2.58 | 13.34 |  |  |  |  |
| Male | low life satisfaction | Stable unemployed | ALSPAC(G0) | |  |  |  |  | no measure |  |  |
| Male | low life satisfaction | Stable unemployed | Twins UK |  |  |  |  |  | dont know |  |  |
| Male | low life satisfaction | Stable unemployed | Overall | 1.64 | 1.24 | 2.17 |  | 40.31 |  |  |  |
|  |  |  |  |  |  |  |  |  |  |  | 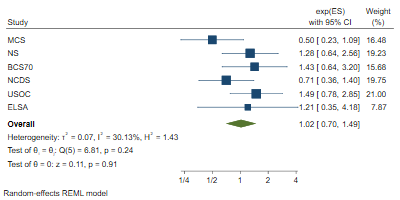 |
| Male | often lonely | Furloughed | MCS | 0.50 | 0.23 | 1.09 | 16.48 |  |  | REML |  |
| Male | often lonely | Furloughed | ALSPAC(G1) | |  |  |  |  | no measure |  |  |
| Male | often lonely | Furloughed | NS | 1.28 | 0.64 | 2.56 | 19.23 |  |  |  |  |
| Male | often lonely | Furloughed | BCS70 | 1.43 | 0.64 | 3.20 | 15.68 |  |  |  |  |
| Male | often lonely | Furloughed | NCDS | 0.71 | 0.36 | 1.40 | 19.75 |  |  |  |  |
| Male | often lonely | Furloughed | USOC | 1.49 | 0.78 | 2.85 | 21.00 |  |  |  |  |
| Male | often lonely | Furloughed | ELSA | 1.21 | 0.35 | 4.18 | 7.87 |  |  |  |  |
| Male | often lonely | Furloughed | GS |  |  |  |  |  | low counts |  |  |
| Male | often lonely | Furloughed | ALSPAC(G0) | |  |  |  |  | no measure |  |  |
| Male | often lonely | Furloughed | Twins UK |  |  |  |  |  | low counts |  |  |
| Male | often lonely | Furloughed | Overall | 1.02 | 0.70 | 1.49 |  | 30.13 |  |  |  |
|  |  |  |  |  |  |  |  |  |  |  | 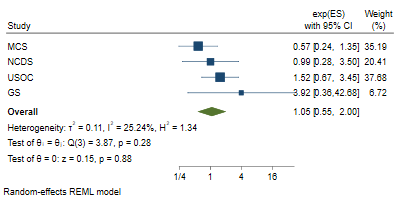 |
| Male | often lonely | No longer employed | MCS | 0.57 | 0.24 | 1.35 | 35.19 |  |  | REML |  |
| Male | often lonely | No longer employed | ALSPAC(G1) | |  |  |  |  | no measure |  |  |
| Male | often lonely | No longer employed | NS |  |  |  |  |  | low counts |  |  |
| Male | often lonely | No longer employed | BCS70 |  |  |  |  |  | low counts |  |  |
| Male | often lonely | No longer employed | NCDS | 0.99 | 0.28 | 3.50 | 20.41 |  |  |  |  |
| Male | often lonely | No longer employed | USOC | 1.52 | 0.67 | 3.45 | 37.68 |  |  |  |  |
| Male | often lonely | No longer employed | ELSA |  |  |  |  |  | low counts |  |  |
| Male | often lonely | No longer employed | GS | 3.92 | 0.36 | 42.68 | 6.72 |  |  |  |  |
| Male | often lonely | No longer employed | ALSPAC(G0) | |  |  |  |  | no measure |  |  |
| Male | often lonely | No longer employed | Twins UK |  |  |  |  |  | low counts |  |  |
| Male | often lonely | No longer employed | Overall | 1.05 | 0.55 | 2.00 |  | 25.24 |  |  |  |
|  |  |  |  |  |  |  |  |  |  |  | 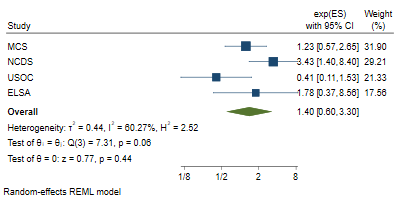 |
| Male | often lonely | Stable unemployed | MCS | 1.23 | 0.57 | 2.65 | 31.90 |  |  | REML |  |
| Male | often lonely | Stable unemployed | ALSPAC(G1) | |  |  |  |  | no measure |  |  |
| Male | often lonely | Stable unemployed | NS |  |  |  |  |  | low counts |  |  |
| Male | often lonely | Stable unemployed | BCS70 |  |  |  |  |  | low counts |  |  |
| Male | often lonely | Stable unemployed | NCDS | 3.43 | 1.40 | 8.40 | 29.21 |  |  |  |  |
| Male | often lonely | Stable unemployed | USOC | 0.41 | 0.11 | 1.53 | 21.33 |  |  |  |  |
| Male | often lonely | Stable unemployed | ELSA | 1.78 | 0.37 | 8.56 | 17.56 |  |  |  |  |
| Male | often lonely | Stable unemployed | GS |  |  |  |  |  | low counts |  |  |
| Male | often lonely | Stable unemployed | ALSPAC(G0) | |  |  |  |  | no measure |  |  |
| Male | often lonely | Stable unemployed | Twins UK |  |  |  |  |  | dont know |  |  |
| Male | often lonely | Stable unemployed | Overall | 1.40 | 0.60 | 3.30 |  | 60.27 |  |  |  |
|  |  |  |  |  |  |  |  |  |  |  | 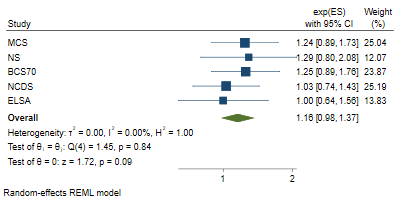 |
| Male | high loneliness | Furloughed | MCS | 1.24 | 0.89 | 1.73 | 25.04 |  |  | REML |  |
| Male | high loneliness | Furloughed | ALSPAC(G1) | |  |  |  |  | no measure |  |  |
| Male | high loneliness | Furloughed | NS | 1.29 | 0.80 | 2.08 | 12.07 |  |  |  |  |
| Male | high loneliness | Furloughed | BCS70 | 1.25 | 0.89 | 1.76 | 23.87 |  |  |  |  |
| Male | high loneliness | Furloughed | NCDS | 1.03 | 0.74 | 1.43 | 25.19 |  |  |  |  |
| Male | high loneliness | Furloughed | USOC |  |  |  |  |  | no measure |  |  |
| Male | high loneliness | Furloughed | ELSA | 1.00 | 0.64 | 1.56 | 13.83 |  |  |  |  |
| Male | high loneliness | Furloughed | GS |  |  |  |  |  | no measure |  |  |
| Male | high loneliness | Furloughed | ALSPAC(G0) | |  |  |  |  | no measure |  |  |
| Male | high loneliness | Furloughed | Twins UK |  |  |  |  |  | low counts |  |  |
| Male | high loneliness | Furloughed | Overall | 1.16 | 0.98 | 1.37 |  | 0.00 |  |  |  |
|  |  |  |  |  |  |  |  |  |  |  | 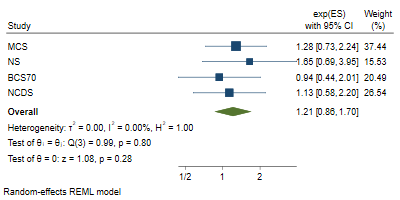 |
| Male | high loneliness | No longer employed | MCS | 1.28 | 0.73 | 2.24 | 37.44 |  |  | REML |  |
| Male | high loneliness | No longer employed | ALSPAC(G1) | |  |  |  |  | no measure |  |  |
| Male | high loneliness | No longer employed | NS | 1.65 | 0.69 | 3.95 | 15.53 |  |  |  |  |
| Male | high loneliness | No longer employed | BCS70 | 0.94 | 0.44 | 2.01 | 20.49 |  |  |  |  |
| Male | high loneliness | No longer employed | NCDS | 1.13 | 0.58 | 2.20 | 26.54 |  |  |  |  |
| Male | high loneliness | No longer employed | USOC |  |  |  |  |  | no measure |  |  |
| Male | high loneliness | No longer employed | ELSA |  |  |  |  |  | low counts |  |  |
| Male | high loneliness | No longer employed | GS |  |  |  |  |  | no measure |  |  |
| Male | high loneliness | No longer employed | ALSPAC(G0) | |  |  |  |  | no measure |  |  |
| Male | high loneliness | No longer employed | Twins UK |  |  |  |  |  | low counts |  |  |
| Male | high loneliness | No longer employed | Overall | 1.21 | 0.86 | 1.70 |  | 0.00 |  |  |  |
|  |  |  |  |  |  |  |  |  |  |  | 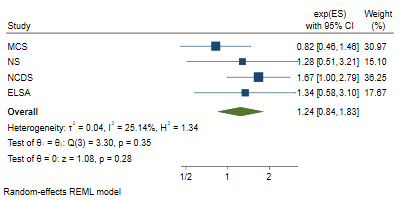 |
| Male | high loneliness | Stable unemployed | MCS | 0.82 | 0.46 | 1.46 | 30.97 |  |  | REML |  |
| Male | high loneliness | Stable unemployed | ALSPAC(G1) | |  |  |  |  | no measure |  |  |
| Male | high loneliness | Stable unemployed | NS | 1.28 | 0.51 | 3.21 | 15.10 |  |  |  |  |
| Male | high loneliness | Stable unemployed | BCS70 |  |  |  |  |  | no measure |  |  |
| Male | high loneliness | Stable unemployed | NCDS | 1.67 | 1.00 | 2.79 | 36.25 |  |  |  |  |
| Male | high loneliness | Stable unemployed | USOC |  |  |  |  |  | no measure |  |  |
| Male | high loneliness | Stable unemployed | ELSA | 1.34 | 0.58 | 3.10 | 17.67 |  |  |  |  |
| Male | high loneliness | Stable unemployed | GS |  |  |  |  |  | no measure |  |  |
| Male | high loneliness | Stable unemployed | ALSPAC(G0) | |  |  |  |  | no measure |  |  |
| Male | high loneliness | Stable unemployed | Twins UK |  |  |  |  |  | dont know |  |  |
| Male | high loneliness | Stable unemployed | Overall | 1.24 | 0.84 | 1.83 |  | 25.14 |  |  |  |
|  |  |  |  |  |  |  |  |  |  |  | 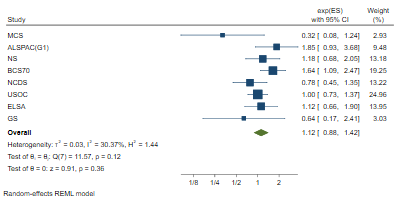 |
| Male | distressed (bin.) | Furloughed | MCS | 0.32 | 0.08 | 1.24 | 2.93 |  |  | REML |  |
| Male | distressed (bin.) | Furloughed | ALSPAC(G1) | 1.85 | 0.93 | 3.68 | 9.48 |  |  |  |  |
| Male | distressed (bin.) | Furloughed | NS | 1.18 | 0.68 | 2.05 | 13.18 |  |  |  |  |
| Male | distressed (bin.) | Furloughed | BCS70 | 1.64 | 1.09 | 2.47 | 19.25 |  |  |  |  |
| Male | distressed (bin.) | Furloughed | NCDS | 0.78 | 0.45 | 1.35 | 13.22 |  |  |  |  |
| Male | distressed (bin.) | Furloughed | USOC | 1.00 | 0.73 | 1.37 | 24.96 |  |  |  |  |
| Male | distressed (bin.) | Furloughed | ELSA | 1.12 | 0.66 | 1.90 | 13.95 |  |  |  |  |
| Male | distressed (bin.) | Furloughed | GS | 0.64 | 0.17 | 2.41 | 3.03 |  |  |  |  |
| Male | distressed (bin.) | Furloughed | ALSPAC(G0) | |  |  |  |  | low counts |  |  |
| Male | distressed (bin.) | Furloughed | Twins UK |  |  |  |  |  | low counts |  |  |
| Male | distressed (bin.) | Furloughed | Overall | 1.12 | 0.88 | 1.42 |  | 30.37 |  |  |  |
|  |  |  |  |  |  |  |  |  |  |  | 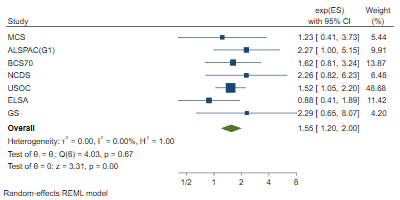 |
| Male | distressed (bin.) | No longer employed | MCS | 1.23 | 0.41 | 3.73 | 5.44 |  |  | REML |  |
| Male | distressed (bin.) | No longer employed | ALSPAC(G1) | 2.27 | 1.00 | 5.15 | 9.91 |  |  |  |  |
| Male | distressed (bin.) | No longer employed | NS |  |  |  |  |  | low counts |  |  |
| Male | distressed (bin.) | No longer employed | BCS70 | 1.62 | 0.81 | 3.24 | 13.87 |  |  |  |  |
| Male | distressed (bin.) | No longer employed | NCDS | 2.26 | 0.82 | 6.23 | 6.48 |  |  |  |  |
| Male | distressed (bin.) | No longer employed | USOC | 1.52 | 1.05 | 2.20 | 48.68 |  |  |  |  |
| Male | distressed (bin.) | No longer employed | ELSA | 0.88 | 0.41 | 1.89 | 11.42 |  |  |  |  |
| Male | distressed (bin.) | No longer employed | GS | 2.29 | 0.65 | 8.07 | 4.20 |  |  |  |  |
| Male | distressed (bin.) | No longer employed | ALSPAC(G0) | |  |  |  |  | low counts |  |  |
| Male | distressed (bin.) | No longer employed | Twins UK |  |  |  |  |  | low counts |  |  |
| Male | distressed (bin.) | No longer employed | Overall | 1.55 | 1.20 | 2.00 |  | 0.00 |  |  |  |
|  |  |  |  |  |  |  |  |  |  |  | 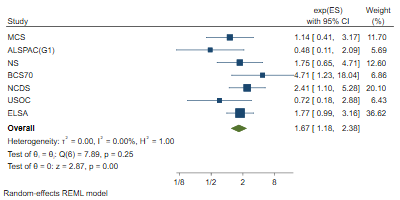 |
| Male | distressed (bin.) | Stable unemployed | MCS | 1.14 | 0.41 | 3.17 | 11.70 |  |  | REML |  |
| Male | distressed (bin.) | Stable unemployed | ALSPAC(G1) | 0.48 | 0.11 | 2.09 | 5.69 |  |  |  |  |
| Male | distressed (bin.) | Stable unemployed | NS | 1.75 | 0.65 | 4.71 | 12.60 |  |  |  |  |
| Male | distressed (bin.) | Stable unemployed | BCS70 | 4.71 | 1.23 | 18.04 | 6.86 |  |  |  |  |
| Male | distressed (bin.) | Stable unemployed | NCDS | 2.41 | 1.10 | 5.28 | 20.10 |  |  |  |  |
| Male | distressed (bin.) | Stable unemployed | USOC | 0.72 | 0.18 | 2.88 | 6.43 |  |  |  |  |
| Male | distressed (bin.) | Stable unemployed | ELSA | 1.77 | 0.99 | 3.16 | 36.62 |  |  |  |  |
| Male | distressed (bin.) | Stable unemployed | GS |  |  |  |  |  | low counts |  |  |
| Male | distressed (bin.) | Stable unemployed | ALSPAC(G0) | |  |  |  |  | low counts |  |  |
| Male | distressed (bin.) | Stable unemployed | Twins UK |  |  |  |  |  | dont know |  |  |
| Male | distressed (bin.) | Stable unemployed | Overall | 1.67 | 1.18 | 2.38 |  | 0.00 |  |  |  |
|  |  |  |  |  |  |  |  |  |  |  | 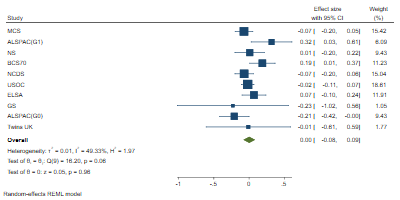 |
| Male | distressed (cont.) | Furloughed | MCS | -0.07 | -0.20 | 0.05 | 15.42 |  |  | REML |  |
| Male | distressed (cont.) | Furloughed | ALSPAC(G1) | 0.32 | 0.03 | 0.61 | 6.09 |  |  |  |  |
| Male | distressed (cont.) | Furloughed | NS | 0.01 | -0.20 | 0.22 | 9.43 |  |  |  |  |
| Male | distressed (cont.) | Furloughed | BCS70 | 0.19 | 0.01 | 0.37 | 11.23 |  |  |  |  |
| Male | distressed (cont.) | Furloughed | NCDS | -0.07 | -0.20 | 0.06 | 15.04 |  |  |  |  |
| Male | distressed (cont.) | Furloughed | USOC | -0.02 | -0.11 | 0.07 | 18.61 |  |  |  |  |
| Male | distressed (cont.) | Furloughed | ELSA | 0.07 | -0.10 | 0.24 | 11.91 |  |  |  |  |
| Male | distressed (cont.) | Furloughed | GS | -0.23 | -1.02 | 0.56 | 1.05 |  |  |  |  |
| Male | distressed (cont.) | Furloughed | ALSPAC(G0) | -0.21 | -0.42 | 0.00 | 9.43 |  |  |  |  |
| Male | distressed (cont.) | Furloughed | Twins UK | -0.01 | -0.61 | 0.59 | 1.77 |  |  |  |  |
| Male | distressed (cont.) | Furloughed | Overall | 0.00 | -0.08 | 0.09 |  | 49.33 |  |  |  |
|  |  |  |  |  |  |  |  |  |  |  | 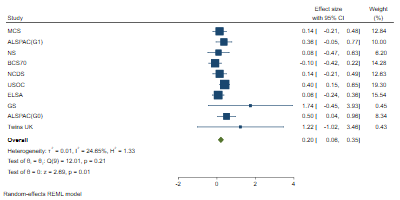 |
| Male | distressed (cont.) | No longer employed | MCS | 0.14 | -0.21 | 0.48 | 12.84 |  |  | REML |  |
| Male | distressed (cont.) | No longer employed | ALSPAC(G1) | 0.36 | -0.05 | 0.77 | 10.00 |  |  |  |  |
| Male | distressed (cont.) | No longer employed | NS | 0.08 | -0.47 | 0.63 | 6.20 |  |  |  |  |
| Male | distressed (cont.) | No longer employed | BCS70 | -0.10 | -0.42 | 0.22 | 14.28 |  |  |  |  |
| Male | distressed (cont.) | No longer employed | NCDS | 0.14 | -0.21 | 0.49 | 12.63 |  |  |  |  |
| Male | distressed (cont.) | No longer employed | USOC | 0.40 | 0.15 | 0.65 | 19.30 |  |  |  |  |
| Male | distressed (cont.) | No longer employed | ELSA | 0.06 | -0.24 | 0.36 | 15.54 |  |  |  |  |
| Male | distressed (cont.) | No longer employed | GS | 1.74 | -0.45 | 3.93 | 0.45 |  |  |  |  |
| Male | distressed (cont.) | No longer employed | ALSPAC(G0) | 0.50 | 0.04 | 0.96 | 8.34 |  |  |  |  |
| Male | distressed (cont.) | No longer employed | Twins UK | 1.22 | -1.02 | 3.46 | 0.43 |  |  |  |  |
| Male | distressed (cont.) | No longer employed | Overall | 0.20 | 0.06 | 0.35 |  | 24.65 |  |  |  |
|  |  |  |  |  |  |  |  |  |  |  | 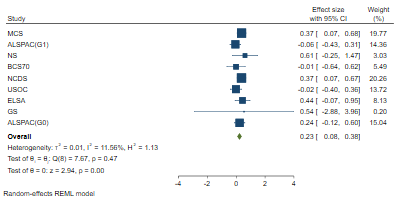 |
| Male | distressed (cont.) | Stable unemployed | MCS | 0.37 | 0.07 | 0.68 | 19.77 |  |  | REML |  |
| Male | distressed (cont.) | Stable unemployed | ALSPAC(G1) | -0.06 | -0.43 | 0.31 | 14.36 |  |  |  |  |
| Male | distressed (cont.) | Stable unemployed | NS | 0.61 | -0.25 | 1.47 | 3.03 |  |  |  |  |
| Male | distressed (cont.) | Stable unemployed | BCS70 | -0.01 | -0.64 | 0.62 | 5.49 |  |  |  |  |
| Male | distressed (cont.) | Stable unemployed | NCDS | 0.37 | 0.07 | 0.67 | 20.26 |  |  |  |  |
| Male | distressed (cont.) | Stable unemployed | USOC | -0.02 | -0.40 | 0.36 | 13.72 |  |  |  |  |
| Male | distressed (cont.) | Stable unemployed | ELSA | 0.44 | -0.07 | 0.95 | 8.13 |  |  |  |  |
| Male | distressed (cont.) | Stable unemployed | GS | 0.54 | -2.88 | 3.96 | 0.20 |  |  |  |  |
| Male | distressed (cont.) | Stable unemployed | ALSPAC(G0) | 0.24 | -0.12 | 0.60 | 15.04 |  |  |  |  |
| Male | distressed (cont.) | Stable unemployed | Twins UK |  |  |  |  |  | dont know |  |  |
| Male | distressed (cont.) | Stable unemployed | Overall | 0.23 | 0.08 | 0.38 |  | 11.56 |  |  |  |
|  |  |  |  |  |  |  |  |  |  |  | 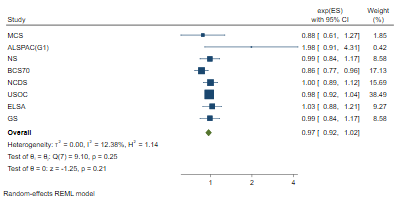 |
| Male | less than daily contact | Furloughed | MCS | 0.88 | 0.61 | 1.27 | 1.85 |  |  | REML |  |
| Male | less than daily contact | Furloughed | ALSPAC(G1) | 1.98 | 0.91 | 4.31 | 0.42 |  |  |  |  |
| Male | less than daily contact | Furloughed | NS | 0.99 | 0.84 | 1.17 | 8.58 |  |  |  |  |
| Male | less than daily contact | Furloughed | BCS70 | 0.86 | 0.77 | 0.96 | 17.13 |  |  |  |  |
| Male | less than daily contact | Furloughed | NCDS | 1.00 | 0.89 | 1.12 | 15.69 |  |  |  |  |
| Male | less than daily contact | Furloughed | USOC | 0.98 | 0.92 | 1.04 | 38.49 |  |  |  |  |
| Male | less than daily contact | Furloughed | ELSA | 1.03 | 0.88 | 1.21 | 9.27 |  |  |  |  |
| Male | less than daily contact | Furloughed | GS | 0.99 | 0.84 | 1.17 | 8.58 |  |  |  |  |
| Male | less than daily contact | Furloughed | ALSPAC(G0) | |  |  |  |  | low counts |  |  |
| Male | less than daily contact | Furloughed | Twins UK |  |  |  |  |  | low counts |  |  |
| Male | less than daily contact | Furloughed | Overall | 0.97 | 0.92 | 1.02 |  | 12.38 |  |  |  |
| Male | less than daily contact | No longer employed | MCS | 0.95 | 0.58 | 1.56 | 3.94 |  |  | REML |  |
| Male | less than daily contact | No longer employed | ALSPAC(G1) | |  |  |  |  | low counts |  |  |
| Male | less than daily contact | No longer employed | NS | 0.83 | 0.53 | 1.30 | 4.72 |  |  |  |  |
| Male | less than daily contact | No longer employed | BCS70 | 0.98 | 0.76 | 1.26 | 13.18 |  |  |  |  |
| Male | less than daily contact | No longer employed | NCDS | 1.23 | 1.04 | 1.45 | 25.30 |  |  |  |  |
| Male | less than daily contact | No longer employed | USOC | 1.01 | 0.88 | 1.16 | 32.91 |  |  |  |  |
| Male | less than daily contact | No longer employed | ELSA | 0.95 | 0.62 | 1.46 | 5.19 |  |  |  |  |
| Male | less than daily contact | No longer employed | GS | 1.04 | 0.82 | 1.32 | 14.76 |  |  |  |  |
| Male | less than daily contact | No longer employed | ALSPAC(G0) | |  |  |  |  | low counts |  |  |
| Male | less than daily contact | No longer employed | Twins UK |  |  |  |  |  | low counts |  |  |
| Male | less than daily contact | No longer employed | Overall | 1.05 | 0.95 | 1.16 |  | 16.34 |  |  |  |
| Male | less than daily contact | Stable unemployed | MCS | 0.91 | 0.51 | 1.62 | 4.31 |  |  | REML |  |
| Male | less than daily contact | Stable unemployed | ALSPAC(G1) | 2.26 | 0.88 | 5.80 | 1.71 |  |  |  |  |
| Male | less than daily contact | Stable unemployed | NS | 0.98 | 0.63 | 1.52 | 6.99 |  |  |  |  |
| Male | less than daily contact | Stable unemployed | BCS70 | 1.38 | 1.03 | 1.85 | 13.62 |  |  |  |  |
| Male | less than daily contact | Stable unemployed | NCDS | 1.25 | 1.05 | 1.49 | 26.07 |  |  |  |  |
| Male | less than daily contact | Stable unemployed | USOC | 0.98 | 0.81 | 1.19 | 23.75 |  |  |  |  |
| Male | less than daily contact | Stable unemployed | ELSA | 0.95 | 0.68 | 1.33 | 11.10 |  |  |  |  |
| Male | less than daily contact | Stable unemployed | GS | 1.20 | 0.86 | 1.67 | 11.16 |  |  |  |  |
| Male | less than daily contact | Stable unemployed | ALSPAC(G0) | 2.02 | 0.68 | 6.00 | 1.29 |  |  |  |  |
| Male | less than daily contact | Stable unemployed | Twins UK |  |  |  |  |  | dont know |  |  |
| Male | less than daily contact | Stable unemployed | Overall | 1.14 | 1.00 | 1.29 |  | 22.72 |  |  |  |
| Male | fair or poor self-rated health | Furloughed | MCS | 0.42 | 0.08 | 2.20 | 0.09 |  |  | MLE |  |
| Male | fair or poor self-rated health | Furloughed | ALSPAC(G1) | |  |  |  |  | no measure |  |  |
| Male | fair or poor self-rated health | Furloughed | NS | 0.80 | 0.41 | 1.56 | 0.56 |  |  |  |  |
| Male | fair or poor self-rated health | Furloughed | BCS70 | 1.28 | 0.86 | 1.91 | 1.57 |  |  |  |  |
| Male | fair or poor self-rated health | Furloughed | NCDS | 1.36 | 0.86 | 2.15 | 1.18 |  |  |  |  |
| Male | fair or poor self-rated health | Furloughed | USOC |  |  |  |  |  | no measure |  |  |
| Male | fair or poor self-rated health | Furloughed | ELSA | 1.01 | 0.96 | 1.06 | 96.48 |  |  |  |  |
| Male | fair or poor self-rated health | Furloughed | GS | 0.61 | 0.14 | 2.66 | 0.11 |  |  |  |  |
| Male | fair or poor self-rated health | Furloughed | ALSPAC(G0) | |  |  |  |  | no measure |  |  |
| Male | fair or poor self-rated health | Furloughed | Twins UK |  |  |  |  |  | low counts |  |  |
| Male | fair or poor self-rated health | Furloughed | Overall | 1.01 | 0.97 | 1.07 |  | 0.00 |  |  |  |
| Male | fair or poor self-rated health | No longer employed | MCS |  |  |  |  |  | low counts | REML |  |
| Male | fair or poor self-rated health | No longer employed | ALSPAC(G1) | |  |  |  |  | no measure |  |  |
| Male | fair or poor self-rated health | No longer employed | NS |  |  |  |  |  | low counts |  |  |
| Male | fair or poor self-rated health | No longer employed | BCS70 | 1.69 | 0.78 | 3.66 | 16.39 |  |  |  |  |
| Male | fair or poor self-rated health | No longer employed | NCDS | 1.91 | 0.88 | 4.15 | 16.33 |  |  |  |  |
| Male | fair or poor self-rated health | No longer employed | USOC |  |  |  |  |  | no measure |  |  |
| Male | fair or poor self-rated health | No longer employed | ELSA | 1.12 | 0.98 | 1.28 | 59.31 |  |  |  |  |
| Male | fair or poor self-rated health | No longer employed | GS | 2.13 | 0.64 | 7.09 | 7.96 |  |  |  |  |
| Male | fair or poor self-rated health | No longer employed | ALSPAC(G0) | |  |  |  |  | no measure |  |  |
| Male | fair or poor self-rated health | No longer employed | Twins UK |  |  |  |  |  | low counts |  |  |
| Male | fair or poor self-rated health | No longer employed | Overall | 1.38 | 0.96 | 1.98 |  | 34.29 |  |  |  |
| Male | fair or poor self-rated health | Stable unemployed | MCS | 3.13 | 1.01 | 9.70 | 2.15 |  |  | REML |  |
| Male | fair or poor self-rated health | Stable unemployed | ALSPAC(G1) | |  |  |  |  | no measure |  |  |
| Male | fair or poor self-rated health | Stable unemployed | NS |  |  |  |  |  | low counts |  |  |
| Male | fair or poor self-rated health | Stable unemployed | BCS70 |  |  |  |  |  | low counts |  |  |
| Male | fair or poor self-rated health | Stable unemployed | NCDS | 1.29 | 0.64 | 2.60 | 5.61 |  |  |  |  |
| Male | fair or poor self-rated health | Stable unemployed | USOC |  |  |  |  |  | no measure |  |  |
| Male | fair or poor self-rated health | Stable unemployed | ELSA | 1.26 | 1.06 | 1.50 | 92.24 |  |  |  |  |
| Male | fair or poor self-rated health | Stable unemployed | GS |  |  |  |  |  | low counts |  |  |
| Male | fair or poor self-rated health | Stable unemployed | ALSPAC(G0) | |  |  |  |  | no measure |  |  |
| Male | fair or poor self-rated health | Stable unemployed | Twins UK |  |  |  |  |  | low counts |  |  |
| Male | fair or poor self-rated health | Stable unemployed | Overall | 1.29 | 1.09 | 1.52 |  | 0.00 |  |  |  |

### 7. No degree

| **Adjustment** | **Outcome** | **Exposure** | **Study** | **Coefficient** | **lower_ci** | **upper_ci** | **%Weight** | **%I2** | **Reason for missing** | **Method** |
| --- | --- | --- | --- | --- | --- | --- | --- | --- | --- | --- |
| No degree | low life satisfaction | Furloughed | MCS | 1.22 | 0.92 | 1.62 | 8.86 |  |  | REML |
| No degree | low life satisfaction | Furloughed | ALSPAC(G1) | |  |  |  |  | no measure |  |
| No degree | low life satisfaction | Furloughed | NS | 0.94 | 0.63 | 1.41 | 4.31 |  |  |  |
| No degree | low life satisfaction | Furloughed | BCS70 | 1.34 | 1.06 | 1.69 | 12.91 |  |  |  |
| No degree | low life satisfaction | Furloughed | NCDS | 1.07 | 0.80 | 1.43 | 8.47 |  |  |  |
| No degree | low life satisfaction | Furloughed | USOC | 1.03 | 0.89 | 1.20 | 30.69 |  |  |  |
| No degree | low life satisfaction | Furloughed | ELSA | 1.23 | 0.97 | 1.58 | 11.84 |  |  |  |
| No degree | low life satisfaction | Furloughed | GS | 1.13 | 0.94 | 1.36 | 21.12 |  |  |  |
| No degree | low life satisfaction | Furloughed | ALSPAC(G0) | |  |  |  |  | no measure |  |
| No degree | low life satisfaction | Furloughed | Twins UK | 1.12 | 0.60 | 2.09 | 1.81 |  |  |  |
| No degree | low life satisfaction | Furloughed | Overall | 1.13 | 1.04 | 1.23 |  | 0.00 |  |  |
| No degree | low life satisfaction | No longer employed | MCS | 1.12 | 0.72 | 1.73 | 12.71 |  |  | REML |
| No degree | low life satisfaction | No longer employed | ALSPAC(G1) | |  |  |  |  | no measure |  |
| No degree | low life satisfaction | No longer employed | NS | 0.98 | 0.46 | 2.08 | 5.23 |  |  |  |
| No degree | low life satisfaction | No longer employed | BCS70 | 1.16 | 0.54 | 2.48 | 5.20 |  |  |  |
| No degree | low life satisfaction | No longer employed | NCDS | 1.62 | 1.07 | 2.47 | 13.67 |  |  |  |
| No degree | low life satisfaction | No longer employed | USOC | 1.42 | 1.13 | 1.79 | 26.99 |  |  |  |
| No degree | low life satisfaction | No longer employed | ELSA | 1.73 | 1.25 | 2.41 | 18.84 |  |  |  |
| No degree | low life satisfaction | No longer employed | GS | 0.98 | 0.69 | 1.39 | 17.37 |  |  |  |
| No degree | low life satisfaction | No longer employed | ALSPAC(G0) | |  |  |  |  | no measure |  |
| No degree | low life satisfaction | No longer employed | Twins UK |  |  |  |  |  | low counts |  |
| No degree | low life satisfaction | No longer employed | Overall | 1.32 | 1.10 | 1.59 |  | 32.03 |  |  |
| No degree | low life satisfaction | Stable unemployed | MCS | 1.21 | 0.89 | 1.66 | 20.85 |  |  | REML |
| No degree | low life satisfaction | Stable unemployed | ALSPAC(G1) | |  |  |  |  | no measure |  |
| No degree | low life satisfaction | Stable unemployed | NS | 1.68 | 1.12 | 2.52 | 16.01 |  |  |  |
| No degree | low life satisfaction | Stable unemployed | BCS70 | 0.63 | 0.17 | 2.28 | 2.55 |  |  |  |
| No degree | low life satisfaction | Stable unemployed | NCDS | 2.19 | 1.54 | 3.11 | 18.69 |  |  |  |
| No degree | low life satisfaction | Stable unemployed | USOC | 1.25 | 0.87 | 1.81 | 17.67 |  |  |  |
| No degree | low life satisfaction | Stable unemployed | ELSA | 1.54 | 1.07 | 2.20 | 18.18 |  |  |  |
| No degree | low life satisfaction | Stable unemployed | GS | 0.97 | 0.44 | 2.13 | 6.04 |  |  |  |
| No degree | low life satisfaction | Stable unemployed | ALSPAC(G0) | |  |  |  |  | no measure |  |
| No degree | low life satisfaction | Stable unemployed | Twins UK |  |  |  |  |  | low counts |  |
| No degree | low life satisfaction | Stable unemployed | Overall | 1.45 | 1.18 | 1.80 |  | 40.28 |  |  |
| No degree | often lonely | Furloughed | MCS | 0.70 | 0.45 | 1.09 | 18.71 |  |  | REML |
| No degree | often lonely | Furloughed | ALSPAC(G1) | |  |  |  |  | no measure |  |
| No degree | often lonely | Furloughed | NS | 1.77 | 0.96 | 3.28 | 13.93 |  |  |  |
| No degree | often lonely | Furloughed | BCS70 | 0.94 | 0.53 | 1.67 | 14.95 |  |  |  |
| No degree | often lonely | Furloughed | NCDS | 0.66 | 0.34 | 1.28 | 12.77 |  |  |  |
| No degree | often lonely | Furloughed | USOC | 1.52 | 1.05 | 2.20 | 21.09 |  |  |  |
| No degree | often lonely | Furloughed | ELSA | 1.35 | 0.72 | 2.53 | 13.68 |  |  |  |
| No degree | often lonely | Furloughed | GS | 1.32 | 0.36 | 4.87 | 4.87 |  |  |  |
| No degree | often lonely | Furloughed | ALSPAC(G0) | |  |  |  |  | no measure |  |
| No degree | often lonely | Furloughed | Twins UK |  |  |  |  |  | low counts |  |
| No degree | often lonely | Furloughed | Overall | 1.10 | 0.80 | 1.51 |  | 51.17 |  |  |
| No degree | often lonely | No longer employed | MCS | 0.33 | 0.12 | 0.91 | 14.56 |  |  | REML |
| No degree | often lonely | No longer employed | ALSPAC(G1) | |  |  |  |  | no measure |  |
| No degree | often lonely | No longer employed | NS | 2.03 | 0.62 | 6.62 | 12.89 |  |  |  |
| No degree | often lonely | No longer employed | BCS70 | 4.86 | 2.32 | 10.21 | 16.99 |  |  |  |
| No degree | often lonely | No longer employed | NCDS | 3.24 | 1.33 | 7.92 | 15.53 |  |  |  |
| No degree | often lonely | No longer employed | USOC | 2.10 | 1.31 | 3.37 | 19.36 |  |  |  |
| No degree | often lonely | No longer employed | ELSA | 1.29 | 0.35 | 4.73 | 11.88 |  |  |  |
| No degree | often lonely | No longer employed | GS | 2.60 | 0.46 | 14.83 | 8.78 |  |  |  |
| No degree | often lonely | No longer employed | ALSPAC(G0) | |  |  |  |  | no measure |  |
| No degree | often lonely | No longer employed | Twins UK |  |  |  |  |  | low counts |  |
| No degree | often lonely | No longer employed | Overall | 1.90 | 0.97 | 3.72 |  | 72.57 |  |  |
| No degree | often lonely | Stable unemployed | MCS | 1.12 | 0.66 | 1.90 | 22.57 |  |  | REML |
| No degree | often lonely | Stable unemployed | ALSPAC(G1) | |  |  |  |  | no measure |  |
| No degree | often lonely | Stable unemployed | NS | 2.94 | 1.24 | 6.97 | 15.36 |  |  |  |
| No degree | often lonely | Stable unemployed | BCS70 |  |  |  |  |  | low counts |  |
| No degree | often lonely | Stable unemployed | NCDS | 2.29 | 1.20 | 4.38 | 19.75 |  |  |  |
| No degree | often lonely | Stable unemployed | USOC | 0.77 | 0.36 | 1.63 | 17.55 |  |  |  |
| No degree | often lonely | Stable unemployed | ELSA | 1.68 | 0.65 | 4.37 | 13.77 |  |  |  |
| No degree | often lonely | Stable unemployed | GS | 4.04 | 1.28 | 12.72 | 11.00 |  |  |  |
| No degree | often lonely | Stable unemployed | ALSPAC(G0) | |  |  |  |  | no measure |  |
| No degree | often lonely | Stable unemployed | Twins UK |  |  |  |  |  | low counts |  |
| No degree | often lonely | Stable unemployed | Overall | 1.71 | 1.07 | 2.74 |  | 54.92 |  |  |
| No degree | high loneliness | Furloughed | MCS | 1.23 | 0.92 | 1.64 | 20.00 |  |  | REML |
| No degree | high loneliness | Furloughed | ALSPAC(G1) | |  |  |  |  | no measure |  |
| No degree | high loneliness | Furloughed | NS | 1.23 | 0.87 | 1.76 | 16.56 |  |  |  |
| No degree | high loneliness | Furloughed | BCS70 | 1.30 | 0.98 | 1.73 | 20.27 |  |  |  |
| No degree | high loneliness | Furloughed | NCDS | 1.08 | 0.79 | 1.49 | 18.19 |  |  |  |
| No degree | high loneliness | Furloughed | USOC |  |  |  |  |  | no measure |  |
| No degree | high loneliness | Furloughed | ELSA | 0.84 | 0.60 | 1.16 | 17.78 |  |  |  |
| No degree | high loneliness | Furloughed | GS |  |  |  |  |  | no measure |  |
| No degree | high loneliness | Furloughed | ALSPAC(G0) | |  |  |  |  | no measure |  |
| No degree | high loneliness | Furloughed | Twins UK | 0.46 | 0.24 | 0.89 | 7.19 |  |  |  |
| No degree | high loneliness | Furloughed | Overall | 1.06 | 0.87 | 1.29 |  | 50.89 |  |  |
| No degree | high loneliness | No longer employed | MCS | 1.58 | 0.93 | 2.69 | 25.47 |  |  | REML |
| No degree | high loneliness | No longer employed | ALSPAC(G1) | |  |  |  |  | no measure |  |
| No degree | high loneliness | No longer employed | NS | 0.97 | 0.51 | 1.87 | 17.13 |  |  |  |
| No degree | high loneliness | No longer employed | BCS70 | 0.95 | 0.40 | 2.26 | 9.70 |  |  |  |
| No degree | high loneliness | No longer employed | NCDS | 1.25 | 0.69 | 2.27 | 20.62 |  |  |  |
| No degree | high loneliness | No longer employed | USOC |  |  |  |  |  | no measure |  |
| No degree | high loneliness | No longer employed | ELSA | 1.01 | 0.60 | 1.69 | 27.09 |  |  |  |
| No degree | high loneliness | No longer employed | GS |  |  |  |  |  | no measure |  |
| No degree | high loneliness | No longer employed | ALSPAC(G0) | |  |  |  |  | no measure |  |
| No degree | high loneliness | No longer employed | Twins UK |  |  |  |  |  | low counts |  |
| No degree | high loneliness | No longer employed | Overall | 1.17 | 0.89 | 1.53 |  | 0.00 |  |  |
| No degree | high loneliness | Stable unemployed | MCS | 0.88 | 0.51 | 1.51 | 20.54 |  |  | REML |
| No degree | high loneliness | Stable unemployed | ALSPAC(G1) | |  |  |  |  | no measure |  |
| No degree | high loneliness | Stable unemployed | NS | 1.24 | 0.73 | 2.10 | 21.79 |  |  |  |
| No degree | high loneliness | Stable unemployed | BCS70 |  |  |  |  |  | low counts |  |
| No degree | high loneliness | Stable unemployed | NCDS | 1.52 | 0.98 | 2.35 | 31.26 |  |  |  |
| No degree | high loneliness | Stable unemployed | USOC |  |  |  |  |  | no measure |  |
| No degree | high loneliness | Stable unemployed | ELSA | 1.15 | 0.71 | 1.86 | 26.40 |  |  |  |
| No degree | high loneliness | Stable unemployed | GS |  |  |  |  |  | no measure |  |
| No degree | high loneliness | Stable unemployed | ALSPAC(G0) | |  |  |  |  | no measure |  |
| No degree | high loneliness | Stable unemployed | Twins UK |  |  |  |  |  | low counts |  |
| No degree | high loneliness | Stable unemployed | Overall | 1.21 | 0.94 | 1.54 |  | 0.00 |  |  |
| No degree | distressed (bin.) | Furloughed | MCS | 1.03 | 0.58 | 1.83 | 5.60 |  |  | REML |
| No degree | distressed (bin.) | Furloughed | ALSPAC(G1) | 1.28 | 0.73 | 2.24 | 5.85 |  |  |  |
| No degree | distressed (bin.) | Furloughed | NS | 1.44 | 1.03 | 2.01 | 13.57 |  |  |  |
| No degree | distressed (bin.) | Furloughed | BCS70 | 1.24 | 0.93 | 1.65 | 16.44 |  |  |  |
| No degree | distressed (bin.) | Furloughed | NCDS | 0.81 | 0.52 | 1.24 | 9.12 |  |  |  |
| No degree | distressed (bin.) | Furloughed | USOC | 0.96 | 0.80 | 1.14 | 27.76 |  |  |  |
| No degree | distressed (bin.) | Furloughed | ELSA | 1.12 | 0.80 | 1.56 | 13.36 |  |  |  |
| No degree | distressed (bin.) | Furloughed | GS | 1.12 | 0.56 | 2.26 | 3.89 |  |  |  |
| No degree | distressed (bin.) | Furloughed | ALSPAC(G0) | 1.59 | 0.83 | 3.06 | 4.41 |  |  |  |
| No degree | distressed (bin.) | Furloughed | Twins UK |  |  |  |  |  | low counts |  |
| No degree | distressed (bin.) | Furloughed | Overall | 1.11 | 0.97 | 1.29 |  | 24.27 |  |  |
| No degree | distressed (bin.) | No longer employed | MCS | 1.36 | 0.69 | 2.70 | 5.62 |  |  | REML |
| No degree | distressed (bin.) | No longer employed | ALSPAC(G1) | 1.43 | 0.62 | 3.33 | 3.69 |  |  |  |
| No degree | distressed (bin.) | No longer employed | NS | 1.62 | 0.92 | 2.87 | 8.07 |  |  |  |
| No degree | distressed (bin.) | No longer employed | BCS70 | 1.70 | 1.07 | 2.69 | 12.34 |  |  |  |
| No degree | distressed (bin.) | No longer employed | NCDS | 1.92 | 0.90 | 4.12 | 4.52 |  |  |  |
| No degree | distressed (bin.) | No longer employed | USOC | 1.40 | 1.11 | 1.77 | 48.84 |  |  |  |
| No degree | distressed (bin.) | No longer employed | ELSA | 1.44 | 0.88 | 2.35 | 10.95 |  |  |  |
| No degree | distressed (bin.) | No longer employed | GS | 2.01 | 0.92 | 4.42 | 4.25 |  |  |  |
| No degree | distressed (bin.) | No longer employed | ALSPAC(G0) | 2.30 | 0.67 | 7.93 | 1.71 |  |  |  |
| No degree | distressed (bin.) | No longer employed | Twins UK |  |  |  |  |  | low counts |  |
| No degree | distressed (bin.) | No longer employed | Overall | 1.51 | 1.28 | 1.78 |  | 0.00 |  |  |
| No degree | distressed (bin.) | Stable unemployed | MCS | 1.18 | 0.57 | 2.46 | 9.66 |  |  | REML |
| No degree | distressed (bin.) | Stable unemployed | ALSPAC(G1) | 0.89 | 0.41 | 1.92 | 8.69 |  |  |  |
| No degree | distressed (bin.) | Stable unemployed | NS | 1.59 | 0.90 | 2.82 | 15.85 |  |  |  |
| No degree | distressed (bin.) | Stable unemployed | BCS70 |  |  |  |  |  | low counts |  |
| No degree | distressed (bin.) | Stable unemployed | NCDS | 2.28 | 0.99 | 5.23 | 7.56 |  |  |  |
| No degree | distressed (bin.) | Stable unemployed | USOC | 1.03 | 0.64 | 1.64 | 23.86 |  |  |  |
| No degree | distressed (bin.) | Stable unemployed | ELSA | 1.42 | 0.89 | 2.27 | 23.58 |  |  |  |
| No degree | distressed (bin.) | Stable unemployed | GS |  |  |  |  |  | low counts |  |
| No degree | distressed (bin.) | Stable unemployed | ALSPAC(G0) | 1.67 | 0.83 | 3.34 | 10.80 |  |  |  |
| No degree | distressed (bin.) | Stable unemployed | Twins UK |  |  |  |  |  | low counts |  |
| No degree | distressed (bin.) | Stable unemployed | Overall | 1.33 | 1.06 | 1.67 |  | 0.00 |  |  |
| No degree | distressed (cont.) | Furloughed | MCS | 0.05 | -0.17 | 0.27 | 7.87 |  |  | REML |
| No degree | distressed (cont.) | Furloughed | ALSPAC(G1) | 0.08 | -0.24 | 0.41 | 4.15 |  |  |  |
| No degree | distressed (cont.) | Furloughed | NS | 0.22 | 0.06 | 0.39 | 11.64 |  |  |  |
| No degree | distressed (cont.) | Furloughed | BCS70 | 0.06 | -0.08 | 0.21 | 13.88 |  |  |  |
| No degree | distressed (cont.) | Furloughed | NCDS | -0.03 | -0.18 | 0.11 | 14.02 |  |  |  |
| No degree | distressed (cont.) | Furloughed | USOC | -0.05 | -0.12 | 0.02 | 24.43 |  |  |  |
| No degree | distressed (cont.) | Furloughed | ELSA | 0.07 | -0.07 | 0.22 | 14.18 |  |  |  |
| No degree | distressed (cont.) | Furloughed | GS | 0.10 | -0.65 | 0.85 | 0.88 |  |  |  |
| No degree | distressed (cont.) | Furloughed | ALSPAC(G0) | 0.09 | -0.14 | 0.32 | 7.34 |  |  |  |
| No degree | distressed (cont.) | Furloughed | Twins UK | -0.13 | -0.68 | 0.42 | 1.61 |  |  |  |
| No degree | distressed (cont.) | Furloughed | Overall | 0.04 | -0.03 | 0.11 |  | 35.30 |  |  |
| No degree | distressed (cont.) | No longer employed | MCS | -0.25 | -0.66 | 0.16 | 9.78 |  |  | REML |
| No degree | distressed (cont.) | No longer employed | ALSPAC(G1) | 0.18 | -0.50 | 0.86 | 4.60 |  |  |  |
| No degree | distressed (cont.) | No longer employed | NS | 0.52 | 0.07 | 0.96 | 8.81 |  |  |  |
| No degree | distressed (cont.) | No longer employed | BCS70 | 0.01 | -0.31 | 0.33 | 12.92 |  |  |  |
| No degree | distressed (cont.) | No longer employed | NCDS | 0.19 | -0.16 | 0.54 | 11.95 |  |  |  |
| No degree | distressed (cont.) | No longer employed | USOC | 0.34 | 0.15 | 0.54 | 19.39 |  |  |  |
| No degree | distressed (cont.) | No longer employed | ELSA | 0.16 | -0.13 | 0.45 | 14.38 |  |  |  |
| No degree | distressed (cont.) | No longer employed | GS | 1.98 | -0.17 | 4.14 | 0.54 |  |  |  |
| No degree | distressed (cont.) | No longer employed | ALSPAC(G0) | 0.30 | -0.04 | 0.64 | 12.19 |  |  |  |
| No degree | distressed (cont.) | No longer employed | Twins UK | -0.49 | -1.10 | 0.12 | 5.44 |  |  |  |
| No degree | distressed (cont.) | No longer employed | Overall | 0.16 | 0.00 | 0.32 |  | 41.31 |  |  |
| No degree | distressed (cont.) | Stable unemployed | MCS | 0.05 | -0.30 | 0.39 | 11.67 |  |  | REML |
| No degree | distressed (cont.) | Stable unemployed | ALSPAC(G1) | -0.60 | -0.98 | -0.23 | 11.10 |  |  |  |
| No degree | distressed (cont.) | Stable unemployed | NS | 0.58 | 0.09 | 1.06 | 9.18 |  |  |  |
| No degree | distressed (cont.) | Stable unemployed | BCS70 | -0.39 | -0.73 | -0.05 | 11.69 |  |  |  |
| No degree | distressed (cont.) | Stable unemployed | NCDS | 0.36 | -0.12 | 0.84 | 9.26 |  |  |  |
| No degree | distressed (cont.) | Stable unemployed | USOC | 0.08 | -0.14 | 0.30 | 13.83 |  |  |  |
| No degree | distressed (cont.) | Stable unemployed | ELSA | 0.26 | -0.14 | 0.66 | 10.66 |  |  |  |
| No degree | distressed (cont.) | Stable unemployed | GS | 0.59 | -3.31 | 4.50 | 0.33 |  |  |  |
| No degree | distressed (cont.) | Stable unemployed | ALSPAC(G0) | 0.15 | -0.11 | 0.41 | 13.21 |  |  |  |
| No degree | distressed (cont.) | Stable unemployed | Twins UK | 0.02 | -0.47 | 0.51 | 9.08 |  |  |  |
| No degree | distressed (cont.) | Stable unemployed | Overall | 0.04 | -0.18 | 0.27 |  | 70.46 |  |  |
| No degree | less than daily contact | Furloughed | MCS | 0.86 | 0.65 | 1.14 | 4.05 |  |  | REML |
| No degree | less than daily contact | Furloughed | ALSPAC(G1) | 1.34 | 0.55 | 3.27 | 0.44 |  |  |  |
| No degree | less than daily contact | Furloughed | NS | 0.87 | 0.72 | 1.05 | 8.44 |  |  |  |
| No degree | less than daily contact | Furloughed | BCS70 | 0.86 | 0.76 | 0.97 | 16.47 |  |  |  |
| No degree | less than daily contact | Furloughed | NCDS | 0.89 | 0.77 | 1.02 | 13.25 |  |  |  |
| No degree | less than daily contact | Furloughed | USOC | 0.99 | 0.93 | 1.06 | 31.99 |  |  |  |
| No degree | less than daily contact | Furloughed | ELSA | 1.01 | 0.89 | 1.15 | 14.99 |  |  |  |
| No degree | less than daily contact | Furloughed | GS | 0.97 | 0.82 | 1.15 | 10.09 |  |  |  |
| No degree | less than daily contact | Furloughed | ALSPAC(G0) | 0.66 | 0.21 | 2.10 | 0.26 |  |  |  |
| No degree | less than daily contact | Furloughed | Twins UK |  |  |  |  |  | low counts |  |
| No degree | less than daily contact | Furloughed | Overall | 0.94 | 0.89 | 1.00 |  | 23.80 |  |  |
| No degree | less than daily contact | No longer employed | MCS | 1.17 | 0.77 | 1.78 | 9.73 |  |  | REML |
| No degree | less than daily contact | No longer employed | ALSPAC(G1) | 1.32 | 0.24 | 7.36 | 0.87 |  |  |  |
| No degree | less than daily contact | No longer employed | NS | 1.10 | 0.69 | 1.75 | 8.53 |  |  |  |
| No degree | less than daily contact | No longer employed | BCS70 | 0.75 | 0.49 | 1.16 | 9.40 |  |  |  |
| No degree | less than daily contact | No longer employed | NCDS | 1.00 | 0.78 | 1.28 | 16.92 |  |  |  |
| No degree | less than daily contact | No longer employed | USOC | 0.93 | 0.79 | 1.09 | 22.03 |  |  |  |
| No degree | less than daily contact | No longer employed | ELSA | 0.83 | 0.59 | 1.18 | 12.31 |  |  |  |
| No degree | less than daily contact | No longer employed | GS | 1.40 | 1.14 | 1.72 | 19.22 |  |  |  |
| No degree | less than daily contact | No longer employed | ALSPAC(G0) | 1.83 | 0.36 | 9.21 | 0.98 |  |  |  |
| No degree | less than daily contact | No longer employed | Twins UK |  |  |  |  |  | low counts |  |
| No degree | less than daily contact | No longer employed | Overall | 1.03 | 0.88 | 1.21 |  | 48.27 |  |  |
| No degree | less than daily contact | Stable unemployed | MCS | 0.98 | 0.58 | 1.64 | 7.81 |  |  | REML |
| No degree | less than daily contact | Stable unemployed | ALSPAC(G1) | 2.79 | 1.07 | 7.26 | 2.79 |  |  |  |
| No degree | less than daily contact | Stable unemployed | NS | 0.94 | 0.55 | 1.63 | 7.17 |  |  |  |
| No degree | less than daily contact | Stable unemployed | BCS70 | 1.22 | 0.82 | 1.82 | 11.22 |  |  |  |
| No degree | less than daily contact | Stable unemployed | NCDS | 1.36 | 1.08 | 1.70 | 19.76 |  |  |  |
| No degree | less than daily contact | Stable unemployed | USOC | 0.89 | 0.75 | 1.05 | 23.57 |  |  |  |
| No degree | less than daily contact | Stable unemployed | ELSA | 0.91 | 0.68 | 1.21 | 16.16 |  |  |  |
| No degree | less than daily contact | Stable unemployed | GS | 1.23 | 0.75 | 2.01 | 8.37 |  |  |  |
| No degree | less than daily contact | Stable unemployed | ALSPAC(G0) | 1.28 | 0.52 | 3.14 | 3.13 |  |  |  |
| No degree | less than daily contact | Stable unemployed | Twins UK |  |  |  |  |  | low counts |  |
| No degree | less than daily contact | Stable unemployed | Overall | 1.09 | 0.92 | 1.29 |  | 42.65 |  |  |
| No degree | fair or poor self-rated health | Furloughed | MCS | 1.31 | 0.51 | 3.33 | 3.40 |  |  | MLE |
| No degree | fair or poor self-rated health | Furloughed | ALSPAC(G1) | |  |  |  |  | no measure |  |
| No degree | fair or poor self-rated health | Furloughed | NS | 1.43 | 0.73 | 2.81 | 6.17 |  |  |  |
| No degree | fair or poor self-rated health | Furloughed | BCS70 | 1.47 | 1.01 | 2.14 | 16.06 |  |  |  |
| No degree | fair or poor self-rated health | Furloughed | NCDS | 1.41 | 0.94 | 2.13 | 14.11 |  |  |  |
| No degree | fair or poor self-rated health | Furloughed | USOC |  |  |  |  |  | no measure |  |
| No degree | fair or poor self-rated health | Furloughed | ELSA | 1.04 | 0.99 | 1.10 | 54.85 |  |  |  |
| No degree | fair or poor self-rated health | Furloughed | GS | 1.51 | 0.73 | 3.12 | 5.41 |  |  |  |
| No degree | fair or poor self-rated health | Furloughed | ALSPAC(G0) | |  |  |  |  | no measure |  |
| No degree | fair or poor self-rated health | Furloughed | Twins UK |  |  |  |  |  | low counts |  |
| No degree | fair or poor self-rated health | Furloughed | Overall | 1.20 | 1.01 | 1.44 |  | 27.97 |  |  |
| No degree | fair or poor self-rated health | No longer employed | MCS |  |  |  |  |  | low counts | REML |
| No degree | fair or poor self-rated health | No longer employed | ALSPAC(G1) | |  |  |  |  | no measure |  |
| No degree | fair or poor self-rated health | No longer employed | NS |  |  |  |  |  | low counts |  |
| No degree | fair or poor self-rated health | No longer employed | BCS70 | 1.45 | 0.54 | 3.85 | 12.01 |  |  |  |
| No degree | fair or poor self-rated health | No longer employed | NCDS | 1.41 | 0.68 | 2.91 | 18.37 |  |  |  |
| No degree | fair or poor self-rated health | No longer employed | USOC |  |  |  |  |  | no measure |  |
| No degree | fair or poor self-rated health | No longer employed | ELSA | 1.06 | 0.95 | 1.19 | 49.78 |  |  |  |
| No degree | fair or poor self-rated health | No longer employed | GS | 2.30 | 1.16 | 4.56 | 19.84 |  |  |  |
| No degree | fair or poor self-rated health | No longer employed | ALSPAC(G0) | |  |  |  |  | no measure |  |
| No degree | fair or poor self-rated health | No longer employed | Twins UK |  |  |  |  |  | low counts |  |
| No degree | fair or poor self-rated health | No longer employed | Overall | 1.35 | 0.92 | 1.99 |  | 48.24 |  |  |
| No degree | fair or poor self-rated health | Stable unemployed | MCS | 3.16 | 1.13 | 8.85 | 15.68 |  |  | REML |
| No degree | fair or poor self-rated health | Stable unemployed | ALSPAC(G1) | |  |  |  |  | no measure |  |
| No degree | fair or poor self-rated health | Stable unemployed | NS | 2.35 | 1.04 | 5.31 | 18.51 |  |  |  |
| No degree | fair or poor self-rated health | Stable unemployed | BCS70 | 8.35 | 3.07 | 22.68 | 16.03 |  |  |  |
| No degree | fair or poor self-rated health | Stable unemployed | NCDS | 2.08 | 1.33 | 3.25 | 23.48 |  |  |  |
| No degree | fair or poor self-rated health | Stable unemployed | USOC |  |  |  |  |  | no measure |  |
| No degree | fair or poor self-rated health | Stable unemployed | ELSA | 1.12 | 0.99 | 1.28 | 26.30 |  |  |  |
| No degree | fair or poor self-rated health | Stable unemployed | GS |  |  |  |  |  | low counts |  |
| No degree | fair or poor self-rated health | Stable unemployed | ALSPAC(G0) | |  |  |  |  | no measure |  |
| No degree | fair or poor self-rated health | Stable unemployed | Twins UK |  |  |  |  |  | low counts |  |
| No degree | fair or poor self-rated health | Stable unemployed | Overall | 2.41 | 1.28 | 4.56 |  | 85.43 |  |  |

`

### 8. Degreee

| Adjustment | Outcome | Exposure | Study | Coefficient | lower_ci | upper_ci | %Weight | %I2 | Reason for missing | Method |
| --- | --- | --- | --- | --- | --- | --- | --- | --- | --- | --- |
| Degree | low life satisfaction | Furloughed | MCS | 1.25 | 0.82 | 1.92 | 5.23 |  |  | REML |
| Degree | low life satisfaction | Furloughed | ALSPAC(G1) | |  |  |  |  | no measure |  |
| Degree | low life satisfaction | Furloughed | NS | 1.11 | 0.76 | 1.62 | 6.55 |  |  |  |
| Degree | low life satisfaction | Furloughed | BCS70 | 1.28 | 1.05 | 1.56 | 23.88 |  |  |  |
| Degree | low life satisfaction | Furloughed | NCDS | 1.37 | 1.06 | 1.75 | 15.12 |  |  |  |
| Degree | low life satisfaction | Furloughed | USOC | 1.16 | 0.94 | 1.41 | 23.16 |  |  |  |
| Degree | low life satisfaction | Furloughed | ELSA | 1.26 | 0.91 | 1.76 | 8.70 |  |  |  |
| Degree | low life satisfaction | Furloughed | GS | 0.98 | 0.75 | 1.29 | 12.54 |  |  |  |
| Degree | low life satisfaction | Furloughed | ALSPAC(G0) | |  |  |  |  | no measure |  |
| Degree | low life satisfaction | Furloughed | Twins UK | 1.28 | 0.82 | 1.99 | 4.81 |  |  |  |
| Degree | low life satisfaction | Furloughed | Overall | 1.21 | 1.10 | 1.33 |  | 0.00 |  |  |
| Degree | low life satisfaction | No longer employed | MCS | 1.08 | 0.66 | 1.77 | 11.76 |  |  | REML |
| Degree | low life satisfaction | No longer employed | ALSPAC(G1) | |  |  |  |  | no measure |  |
| Degree | low life satisfaction | No longer employed | NS | 1.41 | 0.83 | 2.39 | 10.82 |  |  |  |
| Degree | low life satisfaction | No longer employed | BCS70 | 1.01 | 0.54 | 1.87 | 8.68 |  |  |  |
| Degree | low life satisfaction | No longer employed | NCDS | 2.22 | 1.61 | 3.05 | 18.00 |  |  |  |
| Degree | low life satisfaction | No longer employed | USOC | 1.36 | 0.96 | 1.93 | 16.70 |  |  |  |
| Degree | low life satisfaction | No longer employed | ELSA | 1.78 | 1.01 | 3.14 | 9.83 |  |  |  |
| Degree | low life satisfaction | No longer employed | GS | 1.29 | 0.99 | 1.69 | 20.35 |  |  |  |
| Degree | low life satisfaction | No longer employed | ALSPAC(G0) | |  |  |  |  | no measure |  |
| Degree | low life satisfaction | No longer employed | Twins UK | 0.59 | 0.21 | 1.66 | 3.86 |  |  |  |
| Degree | low life satisfaction | No longer employed | Overall | 1.39 | 1.12 | 1.73 |  | 46.96 |  |  |
| Degree | low life satisfaction | Stable unemployed | MCS | 1.35 | 0.78 | 2.36 | 12.55 |  |  | REML |
| Degree | low life satisfaction | Stable unemployed | ALSPAC(G1) | |  |  |  |  | no measure |  |
| Degree | low life satisfaction | Stable unemployed | NS | 2.02 | 1.07 | 3.82 | 10.72 |  |  |  |
| Degree | low life satisfaction | Stable unemployed | BCS70 | 1.18 | 0.91 | 1.52 | 21.37 |  |  |  |
| Degree | low life satisfaction | Stable unemployed | NCDS | 2.68 | 1.83 | 3.92 | 17.29 |  |  |  |
| Degree | low life satisfaction | Stable unemployed | USOC | 1.27 | 0.75 | 2.13 | 13.38 |  |  |  |
| Degree | low life satisfaction | Stable unemployed | ELSA | 2.08 | 1.36 | 3.18 | 15.94 |  |  |  |
| Degree | low life satisfaction | Stable unemployed | GS | 1.30 | 0.62 | 2.75 | 8.76 |  |  |  |
| Degree | low life satisfaction | Stable unemployed | ALSPAC(G0) | |  |  |  |  | no measure |  |
| Degree | low life satisfaction | Stable unemployed | Twins UK |  |  |  |  |  | low counts |  |
| Degree | low life satisfaction | Stable unemployed | Overall | 1.63 | 1.24 | 2.14 |  | 58.33 |  |  |
| Degree | often lonely | Furloughed | MCS | 0.45 | 0.24 | 0.82 | 20.17 |  |  | REML |
| Degree | often lonely | Furloughed | ALSPAC(G1) | |  |  |  |  | no measure |  |
| Degree | often lonely | Furloughed | NS | 1.28 | 0.65 | 2.53 | 18.82 |  |  |  |
| Degree | often lonely | Furloughed | BCS70 | 1.46 | 0.74 | 2.88 | 18.81 |  |  |  |
| Degree | often lonely | Furloughed | NCDS | 1.51 | 0.79 | 2.89 | 19.38 |  |  |  |
| Degree | often lonely | Furloughed | USOC | 1.62 | 0.99 | 2.66 | 22.82 |  |  |  |
| Degree | often lonely | Furloughed | ELSA |  |  |  |  |  | low counts |  |
| Degree | often lonely | Furloughed | GS |  |  |  |  |  | low counts |  |
| Degree | often lonely | Furloughed | ALSPAC(G0) | |  |  |  |  | no measure |  |
| Degree | often lonely | Furloughed | Twins UK |  |  |  |  |  | low counts |  |
| Degree | often lonely | Furloughed | Overall | 1.16 | 0.72 | 1.87 |  | 66.87 |  |  |
| Degree | often lonely | No longer employed | MCS | 0.82 | 0.42 | 1.62 | 30.13 |  |  | REML |
| Degree | often lonely | No longer employed | ALSPAC(G1) | |  |  |  |  | no measure |  |
| Degree | often lonely | No longer employed | NS | 0.55 | 0.21 | 1.47 | 21.45 |  |  |  |
| Degree | often lonely | No longer employed | BCS70 |  |  |  |  |  | low counts |  |
| Degree | often lonely | No longer employed | NCDS | 1.32 | 0.39 | 4.44 | 16.57 |  |  |  |
| Degree | often lonely | No longer employed | USOC | 2.48 | 1.03 | 5.96 | 24.19 |  |  |  |
| Degree | often lonely | No longer employed | ELSA |  |  |  |  |  | low counts |  |
| Degree | often lonely | No longer employed | GS | 2.76 | 0.36 | 20.96 | 7.65 |  |  |  |
| Degree | often lonely | No longer employed | ALSPAC(G0) | |  |  |  |  | no measure |  |
| Degree | often lonely | No longer employed | Twins UK |  |  |  |  |  | low counts |  |
| Degree | often lonely | No longer employed | Overall | 1.17 | 0.63 | 2.16 |  | 43.66 |  |  |
| Degree | often lonely | Stable unemployed | MCS | 0.87 | 0.42 | 1.81 | 27.73 |  |  | REML |
| Degree | often lonely | Stable unemployed | ALSPAC(G1) | |  |  |  |  | no measure |  |
| Degree | often lonely | Stable unemployed | NS | 2.10 | 0.94 | 4.69 | 25.37 |  |  |  |
| Degree | often lonely | Stable unemployed | BCS70 | 1.55 | 0.30 | 7.93 | 10.23 |  |  |  |
| Degree | often lonely | Stable unemployed | NCDS | 3.49 | 1.47 | 8.32 | 23.58 |  |  |  |
| Degree | often lonely | Stable unemployed | USOC | 1.04 | 0.26 | 4.18 | 13.09 |  |  |  |
| Degree | often lonely | Stable unemployed | ELSA |  |  |  |  |  | low counts |  |
| Degree | often lonely | Stable unemployed | GS |  |  |  |  |  | low counts |  |
| Degree | often lonely | Stable unemployed | ALSPAC(G0) | |  |  |  |  | no measure |  |
| Degree | often lonely | Stable unemployed | Twins UK |  |  |  |  |  | low counts |  |
| Degree | often lonely | Stable unemployed | Overall | 1.64 | 0.91 | 2.95 |  | 42.94 |  |  |
| Degree | high loneliness | Furloughed | MCS | 0.84 | 0.61 | 1.14 | 16.88 |  |  | REML |
| Degree | high loneliness | Furloughed | ALSPAC(G1) | |  |  |  |  | no measure |  |
| Degree | high loneliness | Furloughed | NS | 1.27 | 0.81 | 2.00 | 8.88 |  |  |  |
| Degree | high loneliness | Furloughed | BCS70 | 1.23 | 0.96 | 1.58 | 23.66 |  |  |  |
| Degree | high loneliness | Furloughed | NCDS | 1.23 | 0.92 | 1.63 | 19.27 |  |  |  |
| Degree | high loneliness | Furloughed | USOC |  |  |  |  |  | no measure |  |
| Degree | high loneliness | Furloughed | ELSA | 1.49 | 0.95 | 2.36 | 8.80 |  |  |  |
| Degree | high loneliness | Furloughed | GS |  |  |  |  |  | no measure |  |
| Degree | high loneliness | Furloughed | ALSPAC(G0) | |  |  |  |  | no measure |  |
| Degree | high loneliness | Furloughed | Twins UK | 1.27 | 0.98 | 1.65 | 22.52 |  |  |  |
| Degree | high loneliness | Furloughed | Overall | 1.18 | 1.03 | 1.36 |  | 18.59 |  |  |
| Degree | high loneliness | No longer employed | MCS | 0.74 | 0.44 | 1.23 | 24.27 |  |  | REML |
| Degree | high loneliness | No longer employed | ALSPAC(G1) | |  |  |  |  | no measure |  |
| Degree | high loneliness | No longer employed | NS | 1.17 | 0.68 | 2.02 | 22.05 |  |  |  |
| Degree | high loneliness | No longer employed | BCS70 |  |  |  |  |  | low counts |  |
| Degree | high loneliness | No longer employed | NCDS | 1.55 | 0.96 | 2.51 | 26.22 |  |  |  |
| Degree | high loneliness | No longer employed | USOC |  |  |  |  |  | no measure |  |
| Degree | high loneliness | No longer employed | ELSA |  |  |  |  |  | low counts |  |
| Degree | high loneliness | No longer employed | GS |  |  |  |  |  | no measure |  |
| Degree | high loneliness | No longer employed | ALSPAC(G0) | |  |  |  |  | no measure |  |
| Degree | high loneliness | No longer employed | Twins UK | 1.00 | 0.63 | 1.59 | 27.45 |  |  |  |
| Degree | high loneliness | No longer employed | Overall | 1.08 | 0.80 | 1.47 |  | 32.99 |  |  |
| Degree | high loneliness | Stable unemployed | MCS | 0.98 | 0.63 | 1.54 | 27.04 |  |  | REML |
| Degree | high loneliness | Stable unemployed | ALSPAC(G1) | |  |  |  |  | no measure |  |
| Degree | high loneliness | Stable unemployed | NS | 1.44 | 0.71 | 2.90 | 16.20 |  |  |  |
| Degree | high loneliness | Stable unemployed | BCS70 | 0.51 | 0.11 | 2.38 | 4.47 |  |  |  |
| Degree | high loneliness | Stable unemployed | NCDS | 2.15 | 1.33 | 3.46 | 25.64 |  |  |  |
| Degree | high loneliness | Stable unemployed | USOC |  |  |  |  |  | no measure |  |
| Degree | high loneliness | Stable unemployed | ELSA | 1.22 | 0.28 | 5.26 | 4.89 |  |  |  |
| Degree | high loneliness | Stable unemployed | GS |  |  |  |  |  | no measure |  |
| Degree | high loneliness | Stable unemployed | ALSPAC(G0) | |  |  |  |  | no measure |  |
| Degree | high loneliness | Stable unemployed | Twins UK | 1.44 | 0.83 | 2.51 | 21.75 |  |  |  |
| Degree | high loneliness | Stable unemployed | Overall | 1.36 | 0.97 | 1.92 |  | 34.58 |  |  |
| Degree | distressed (bin.) | Furloughed | MCS | 1.21 | 0.80 | 1.82 | 12.21 |  |  | REML |
| Degree | distressed (bin.) | Furloughed | ALSPAC(G1) | 1.79 | 1.19 | 2.68 | 12.29 |  |  |  |
| Degree | distressed (bin.) | Furloughed | NS | 1.17 | 0.80 | 1.72 | 13.22 |  |  |  |
| Degree | distressed (bin.) | Furloughed | BCS70 | 1.47 | 1.06 | 2.03 | 15.58 |  |  |  |
| Degree | distressed (bin.) | Furloughed | NCDS | 1.21 | 0.80 | 1.82 | 12.21 |  |  |  |
| Degree | distressed (bin.) | Furloughed | USOC | 1.01 | 0.82 | 1.24 | 21.31 |  |  |  |
| Degree | distressed (bin.) | Furloughed | ELSA | 0.64 | 0.35 | 1.18 | 7.21 |  |  |  |
| Degree | distressed (bin.) | Furloughed | GS | 0.50 | 0.19 | 1.33 | 3.30 |  |  |  |
| Degree | distressed (bin.) | Furloughed | ALSPAC(G0) | |  |  |  |  | low counts |  |
| Degree | distressed (bin.) | Furloughed | Twins UK | 2.34 | 0.78 | 7.03 | 2.66 |  |  |  |
| Degree | distressed (bin.) | Furloughed | Overall | 1.18 | 0.98 | 1.43 |  | 42.93 |  |  |
| Degree | distressed (bin.) | No longer employed | MCS | 1.63 | 0.80 | 3.32 | 10.96 |  |  | REML |
| Degree | distressed (bin.) | No longer employed | ALSPAC(G1) | 1.71 | 1.01 | 2.90 | 19.88 |  |  |  |
| Degree | distressed (bin.) | No longer employed | NS | 0.93 | 0.45 | 1.93 | 10.35 |  |  |  |
| Degree | distressed (bin.) | No longer employed | BCS70 |  |  |  |  |  | low counts |  |
| Degree | distressed (bin.) | No longer employed | NCDS | 1.63 | 0.80 | 3.32 | 10.96 |  |  |  |
| Degree | distressed (bin.) | No longer employed | USOC | 1.21 | 0.85 | 1.72 | 43.56 |  |  |  |
| Degree | distressed (bin.) | No longer employed | ELSA |  |  |  |  |  | low counts |  |
| Degree | distressed (bin.) | No longer employed | GS | 1.47 | 0.47 | 4.57 | 4.30 |  |  |  |
| Degree | distressed (bin.) | No longer employed | ALSPAC(G0) | |  |  |  |  | low counts |  |
| Degree | distressed (bin.) | No longer employed | Twins UK |  |  |  |  |  | low counts |  |
| Degree | distressed (bin.) | No longer employed | Overall | 1.36 | 1.07 | 1.72 |  | 0.00 |  |  |
| Degree | distressed (bin.) | Stable unemployed | MCS | 2.65 | 1.21 | 5.82 | 15.90 |  |  | REML |
| Degree | distressed (bin.) | Stable unemployed | ALSPAC(G1) | 3.12 | 1.28 | 7.57 | 14.33 |  |  |  |
| Degree | distressed (bin.) | Stable unemployed | NS | 1.35 | 0.67 | 2.72 | 17.23 |  |  |  |
| Degree | distressed (bin.) | Stable unemployed | BCS70 | 0.49 | 0.07 | 3.65 | 4.98 |  |  |  |
| Degree | distressed (bin.) | Stable unemployed | NCDS | 2.65 | 1.21 | 5.82 | 15.90 |  |  |  |
| Degree | distressed (bin.) | Stable unemployed | USOC | 0.74 | 0.35 | 1.56 | 16.50 |  |  |  |
| Degree | distressed (bin.) | Stable unemployed | ELSA | 3.84 | 1.67 | 8.82 | 15.17 |  |  |  |
| Degree | distressed (bin.) | Stable unemployed | GS |  |  |  |  |  | low counts |  |
| Degree | distressed (bin.) | Stable unemployed | ALSPAC(G0) | |  |  |  |  | low counts |  |
| Degree | distressed (bin.) | Stable unemployed | Twins UK |  |  |  |  |  | low counts |  |
| Degree | distressed (bin.) | Stable unemployed | Overall | 1.90 | 1.16 | 3.13 |  | 56.79 |  |  |
| Degree | distressed (cont.) | Furloughed | MCS | -0.14 | -0.44 | 0.17 | 6.13 |  |  | REML |
| Degree | distressed (cont.) | Furloughed | ALSPAC(G1) | 0.28 | 0.05 | 0.51 | 8.76 |  |  |  |
| Degree | distressed (cont.) | Furloughed | NS | -0.03 | -0.20 | 0.14 | 12.45 |  |  |  |
| Degree | distressed (cont.) | Furloughed | BCS70 | 0.25 | 0.08 | 0.42 | 12.49 |  |  |  |
| Degree | distressed (cont.) | Furloughed | NCDS | 0.04 | -0.07 | 0.15 | 16.90 |  |  |  |
| Degree | distressed (cont.) | Furloughed | USOC | 0.00 | -0.10 | 0.09 | 18.99 |  |  |  |
| Degree | distressed (cont.) | Furloughed | ELSA | 0.07 | -0.14 | 0.27 | 10.24 |  |  |  |
| Degree | distressed (cont.) | Furloughed | GS | -0.60 | -1.64 | 0.43 | 0.68 |  |  |  |
| Degree | distressed (cont.) | Furloughed | ALSPAC(G0) | -0.09 | -0.31 | 0.13 | 9.58 |  |  |  |
| Degree | distressed (cont.) | Furloughed | Twins UK | 0.35 | -0.06 | 0.76 | 3.78 |  |  |  |
| Degree | distressed (cont.) | Furloughed | Overall | 0.06 | -0.03 | 0.14 |  | 49.02 |  |  |
| Degree | distressed (cont.) | No longer employed | MCS | -0.03 | -0.40 | 0.35 | 8.89 |  |  | REML |
| Degree | distressed (cont.) | No longer employed | ALSPAC(G1) | 0.24 | -0.02 | 0.49 | 18.63 |  |  |  |
| Degree | distressed (cont.) | No longer employed | NS | 0.13 | -0.22 | 0.49 | 9.97 |  |  |  |
| Degree | distressed (cont.) | No longer employed | BCS70 | -0.08 | -0.47 | 0.31 | 7.94 |  |  |  |
| Degree | distressed (cont.) | No longer employed | NCDS | 0.22 | -0.16 | 0.60 | 8.41 |  |  |  |
| Degree | distressed (cont.) | No longer employed | USOC | 0.21 | -0.05 | 0.48 | 17.80 |  |  |  |
| Degree | distressed (cont.) | No longer employed | ELSA | 0.01 | -0.43 | 0.45 | 6.28 |  |  |  |
| Degree | distressed (cont.) | No longer employed | GS | 1.09 | -0.38 | 2.56 | 0.57 |  |  |  |
| Degree | distressed (cont.) | No longer employed | ALSPAC(G0) | 0.02 | -0.24 | 0.29 | 17.98 |  |  |  |
| Degree | distressed (cont.) | No longer employed | Twins UK | 0.03 | -0.56 | 0.62 | 3.53 |  |  |  |
| Degree | distressed (cont.) | No longer employed | Overall | 0.12 | 0.01 | 0.23 |  | 0.00 |  |  |
| Degree | distressed (cont.) | Stable unemployed | MCS | 0.56 | 0.02 | 1.10 | 9.27 |  |  | REML |
| Degree | distressed (cont.) | Stable unemployed | ALSPAC(G1) | 0.48 | 0.19 | 0.76 | 16.09 |  |  |  |
| Degree | distressed (cont.) | Stable unemployed | NS | 0.04 | -0.57 | 0.64 | 8.08 |  |  |  |
| Degree | distressed (cont.) | Stable unemployed | BCS70 | -0.07 | -0.99 | 0.85 | 4.37 |  |  |  |
| Degree | distressed (cont.) | Stable unemployed | NCDS | 0.33 | 0.01 | 0.66 | 14.89 |  |  |  |
| Degree | distressed (cont.) | Stable unemployed | USOC | 0.08 | -0.27 | 0.44 | 13.85 |  |  |  |
| Degree | distressed (cont.) | Stable unemployed | ELSA | 1.03 | 0.38 | 1.68 | 7.34 |  |  |  |
| Degree | distressed (cont.) | Stable unemployed | GS | 2.88 | -1.13 | 6.90 | 0.28 |  |  |  |
| Degree | distressed (cont.) | Stable unemployed | ALSPAC(G0) | -0.05 | -0.25 | 0.15 | 18.78 |  |  |  |
| Degree | distressed (cont.) | Stable unemployed | Twins UK | 0.50 | -0.17 | 1.17 | 7.05 |  |  |  |
| Degree | distressed (cont.) | Stable unemployed | Overall | 0.30 | 0.08 | 0.51 |  | 53.87 |  |  |
| Degree | less than daily contact | Furloughed | MCS | 0.81 | 0.65 | 1.02 | 3.74 |  |  | REML |
| Degree | less than daily contact | Furloughed | ALSPAC(G1) | 1.57 | 0.72 | 3.41 | 0.32 |  |  |  |
| Degree | less than daily contact | Furloughed | NS | 0.91 | 0.72 | 1.14 | 3.56 |  |  |  |
| Degree | less than daily contact | Furloughed | BCS70 | 1.00 | 0.90 | 1.11 | 17.59 |  |  |  |
| Degree | less than daily contact | Furloughed | NCDS | 0.95 | 0.84 | 1.08 | 12.70 |  |  |  |
| Degree | less than daily contact | Furloughed | USOC | 1.00 | 0.94 | 1.06 | 52.42 |  |  |  |
| Degree | less than daily contact | Furloughed | ELSA | 1.20 | 1.01 | 1.43 | 6.34 |  |  |  |
| Degree | less than daily contact | Furloughed | GS | 0.87 | 0.69 | 1.11 | 3.33 |  |  |  |
| Degree | less than daily contact | Furloughed | ALSPAC(G0) | |  |  |  |  | low counts |  |
| Degree | less than daily contact | Furloughed | Twins UK |  |  |  |  |  | low counts |  |
| Degree | less than daily contact | Furloughed | Overall | 0.99 | 0.95 | 1.04 |  | 0.00 |  |  |
| Degree | less than daily contact | No longer employed | MCS | 0.73 | 0.53 | 0.99 | 9.39 |  |  | REML |
| Degree | less than daily contact | No longer employed | ALSPAC(G1) | 1.38 | 0.48 | 3.94 | 1.00 |  |  |  |
| Degree | less than daily contact | No longer employed | NS | 0.94 | 0.61 | 1.44 | 5.44 |  |  |  |
| Degree | less than daily contact | No longer employed | BCS70 | 1.06 | 0.84 | 1.34 | 14.83 |  |  |  |
| Degree | less than daily contact | No longer employed | NCDS | 0.92 | 0.73 | 1.17 | 14.07 |  |  |  |
| Degree | less than daily contact | No longer employed | USOC | 1.02 | 0.92 | 1.14 | 33.91 |  |  |  |
| Degree | less than daily contact | No longer employed | ELSA | 1.28 | 0.98 | 1.68 | 11.71 |  |  |  |
| Degree | less than daily contact | No longer employed | GS | 0.84 | 0.61 | 1.16 | 8.89 |  |  |  |
| Degree | less than daily contact | No longer employed | ALSPAC(G0) | 2.95 | 0.88 | 9.84 | 0.76 |  |  |  |
| Degree | less than daily contact | No longer employed | Twins UK |  |  |  |  |  | low counts |  |
| Degree | less than daily contact | No longer employed | Overall | 1.00 | 0.90 | 1.11 |  | 22.72 |  |  |
| Degree | less than daily contact | Stable unemployed | MCS | 0.58 | 0.31 | 1.07 | 2.17 |  |  | REML |
| Degree | less than daily contact | Stable unemployed | ALSPAC(G1) | 2.29 | 0.75 | 7.05 | 0.66 |  |  |  |
| Degree | less than daily contact | Stable unemployed | NS | 1.15 | 0.91 | 1.45 | 15.19 |  |  |  |
| Degree | less than daily contact | Stable unemployed | BCS70 | 0.51 | 0.16 | 1.58 | 0.65 |  |  |  |
| Degree | less than daily contact | Stable unemployed | NCDS | 1.19 | 0.99 | 1.44 | 24.72 |  |  |  |
| Degree | less than daily contact | Stable unemployed | USOC | 1.08 | 0.96 | 1.22 | 54.76 |  |  |  |
| Degree | less than daily contact | Stable unemployed | ELSA | 0.97 | 0.43 | 2.16 | 1.28 |  |  |  |
| Degree | less than daily contact | Stable unemployed | GS |  |  |  |  |  | low counts |  |
| Degree | less than daily contact | Stable unemployed | ALSPAC(G0) | 1.98 | 0.59 | 6.59 | 0.58 |  |  |  |
| Degree | less than daily contact | Stable unemployed | Twins UK |  |  |  |  |  | low counts |  |
| Degree | less than daily contact | Stable unemployed | Overall | 1.11 | 1.01 | 1.21 |  | 0.00 |  |  |
| Degree | fair or poor self-rated health | Furloughed | MCS | 1.07 | 0.22 | 5.19 | 1.65 |  |  | MLE |
| Degree | fair or poor self-rated health | Furloughed | ALSPAC(G1) | |  |  |  |  | no measure |  |
| Degree | fair or poor self-rated health | Furloughed | NS | 0.58 | 0.28 | 1.20 | 6.81 |  |  |  |
| Degree | fair or poor self-rated health | Furloughed | BCS70 | 1.29 | 0.89 | 1.86 | 20.12 |  |  |  |
| Degree | fair or poor self-rated health | Furloughed | NCDS | 1.61 | 1.02 | 2.53 | 15.01 |  |  |  |
| Degree | fair or poor self-rated health | Furloughed | USOC |  |  |  |  |  | no measure |  |
| Degree | fair or poor self-rated health | Furloughed | ELSA | 1.07 | 0.99 | 1.16 | 51.28 |  |  |  |
| Degree | fair or poor self-rated health | Furloughed | GS |  |  |  |  |  | low counts |  |
| Degree | fair or poor self-rated health | Furloughed | ALSPAC(G0) | |  |  |  |  | no measure |  |
| Degree | fair or poor self-rated health | Furloughed | Twins UK | 3.35 | 1.41 | 7.94 | 5.14 |  |  |  |
| Degree | fair or poor self-rated health | Furloughed | Overall | 1.20 | 0.98 | 1.47 |  | 31.13 |  |  |
| Degree | fair or poor self-rated health | No longer employed | MCS |  |  |  |  |  | low counts | REML |
| Degree | fair or poor self-rated health | No longer employed | ALSPAC(G1) | |  |  |  |  | no measure |  |
| Degree | fair or poor self-rated health | No longer employed | NS | 4.12 | 1.24 | 13.67 | 8.39 |  |  |  |
| Degree | fair or poor self-rated health | No longer employed | BCS70 | 1.29 | 0.60 | 2.78 | 17.01 |  |  |  |
| Degree | fair or poor self-rated health | No longer employed | NCDS | 1.55 | 0.70 | 3.45 | 16.06 |  |  |  |
| Degree | fair or poor self-rated health | No longer employed | USOC |  |  |  |  |  | no measure |  |
| Degree | fair or poor self-rated health | No longer employed | ELSA | 1.08 | 0.91 | 1.27 | 52.36 |  |  |  |
| Degree | fair or poor self-rated health | No longer employed | GS | 1.74 | 0.42 | 7.27 | 6.17 |  |  |  |
| Degree | fair or poor self-rated health | No longer employed | ALSPAC(G0) | |  |  |  |  | no measure |  |
| Degree | fair or poor self-rated health | No longer employed | Twins UK |  |  |  |  |  | low counts |  |
| Degree | fair or poor self-rated health | No longer employed | Overall | 1.36 | 0.93 | 1.98 |  | 33.32 |  |  |
| Degree | fair or poor self-rated health | Stable unemployed | MCS | 3.05 | 0.62 | 14.97 | 38.10 |  |  | REML |
| Degree | fair or poor self-rated health | Stable unemployed | ALSPAC(G1) | |  |  |  |  | no measure |  |
| Degree | fair or poor self-rated health | Stable unemployed | NS |  |  |  |  |  | low counts |  |
| Degree | fair or poor self-rated health | Stable unemployed | BCS70 |  |  |  |  |  | low counts |  |
| Degree | fair or poor self-rated health | Stable unemployed | NCDS | 1.40 | 0.40 | 4.88 | 61.90 |  |  |  |
| Degree | fair or poor self-rated health | Stable unemployed | USOC |  |  |  |  |  | no measure |  |
| Degree | fair or poor self-rated health | Stable unemployed | ELSA |  |  |  |  |  | low counts |  |
| Degree | fair or poor self-rated health | Stable unemployed | GS |  |  |  |  |  | low counts |  |
| Degree | fair or poor self-rated health | Stable unemployed | ALSPAC(G0) | |  |  |  |  | no measure |  |
| Degree | fair or poor self-rated health | Stable unemployed | Twins UK |  |  |  |  |  | low counts |  |
| Degree | fair or poor self-rated health | Stable unemployed | Overall | 1.88 | 0.70 | 5.03 |  | 0.00 |  |  |

# 9. 16-29

| **Adjustment** | **Outcome** | **Exposure** | **Study** | **Coefficient** | **lower_ci** | **upper_ci** | **%Weight** | **%I2** | **Reason for missing** | **Method** |
| --- | --- | --- | --- | --- | --- | --- | --- | --- | --- | --- |
| 16-29 | low life satisfaction | Furloughed | MCS | 1.23 | 0.96 | 1.58 | 59.55 |  |  | REML |
| 16-29 | low life satisfaction | Furloughed | ALSPAC(G1) | |  |  |  |  | no measure |  |
| 16-29 | low life satisfaction | Furloughed | NS |  |  |  |  |  | no representation |  |
| 16-29 | low life satisfaction | Furloughed | BCS70 |  |  |  |  |  | no representation |  |
| 16-29 | low life satisfaction | Furloughed | NCDS |  |  |  |  |  | no representation |  |
| 16-29 | low life satisfaction | Furloughed | USOC | 1.05 | 0.78 | 1.42 | 40.45 |  |  |  |
| 16-29 | low life satisfaction | Furloughed | ELSA |  |  |  |  |  | no representation |  |
| 16-29 | low life satisfaction | Furloughed | GS |  |  |  |  |  | no representation |  |
| 16-29 | low life satisfaction | Furloughed | ALSPAC(G0) | |  |  |  |  | no measure |  |
| 16-29 | low life satisfaction | Furloughed | Twins UK |  |  |  |  |  | no representation |  |
| 16-29 | low life satisfaction | Furloughed | Overall | 1.16 | 0.95 | 1.40 |  | 0.00 |  |  |
| 16-29 | low life satisfaction | No longer employed | MCS | 1.10 | 0.80 | 1.51 | 51.99 |  |  | REML |
| 16-29 | low life satisfaction | No longer employed | ALSPAC(G1) | |  |  |  |  | no measure |  |
| 16-29 | low life satisfaction | No longer employed | NS |  |  |  |  |  | no representation |  |
| 16-29 | low life satisfaction | No longer employed | BCS70 |  |  |  |  |  | no representation |  |
| 16-29 | low life satisfaction | No longer employed | NCDS |  |  |  |  |  | no representation |  |
| 16-29 | low life satisfaction | No longer employed | USOC | 1.65 | 1.16 | 2.35 | 48.01 |  |  |  |
| 16-29 | low life satisfaction | No longer employed | ELSA |  |  |  |  |  | no representation |  |
| 16-29 | low life satisfaction | No longer employed | GS |  |  |  |  |  | no representation |  |
| 16-29 | low life satisfaction | No longer employed | ALSPAC(G0) | |  |  |  |  | no measure |  |
| 16-29 | low life satisfaction | No longer employed | Twins UK |  |  |  |  |  | no representation |  |
| 16-29 | low life satisfaction | No longer employed | Overall | 1.34 | 0.90 | 1.99 |  | 64.59 |  |  |
| 16-29 | low life satisfaction | Stable unemployed | MCS | 1.19 | 0.93 | 1.52 | 91.08 |  |  | REML |
| 16-29 | low life satisfaction | Stable unemployed | ALSPAC(G1) | |  |  |  |  | no measure |  |
| 16-29 | low life satisfaction | Stable unemployed | NS |  |  |  |  |  | no representation |  |
| 16-29 | low life satisfaction | Stable unemployed | BCS70 |  |  |  |  |  | no representation |  |
| 16-29 | low life satisfaction | Stable unemployed | NCDS |  |  |  |  |  | no representation |  |
| 16-29 | low life satisfaction | Stable unemployed | USOC | 1.21 | 0.55 | 2.67 | 8.92 |  |  |  |
| 16-29 | low life satisfaction | Stable unemployed | ELSA |  |  |  |  |  | no representation |  |
| 16-29 | low life satisfaction | Stable unemployed | GS |  |  |  |  |  | no representation |  |
| 16-29 | low life satisfaction | Stable unemployed | ALSPAC(G0) | |  |  |  |  | no measure |  |
| 16-29 | low life satisfaction | Stable unemployed | Twins UK |  |  |  |  |  | no representation |  |
| 16-29 | low life satisfaction | Stable unemployed | Overall | 1.19 | 0.94 | 1.50 |  | 0.00 |  |  |
| 16-29 | often lonely | Furloughed | MCS | 0.60 | 0.42 | 0.85 | 52.31 |  |  | REML |
| 16-29 | often lonely | Furloughed | ALSPAC(G1) | |  |  |  |  | no measure |  |
| 16-29 | often lonely | Furloughed | NS |  |  |  |  |  | no representation |  |
| 16-29 | often lonely | Furloughed | BCS70 |  |  |  |  |  | no representation |  |
| 16-29 | often lonely | Furloughed | NCDS |  |  |  |  |  | no representation |  |
| 16-29 | often lonely | Furloughed | USOC | 1.34 | 0.82 | 2.17 | 47.69 |  |  |  |
| 16-29 | often lonely | Furloughed | ELSA |  |  |  |  |  | no representation |  |
| 16-29 | often lonely | Furloughed | GS |  |  |  |  |  | no representation |  |
| 16-29 | often lonely | Furloughed | ALSPAC(G0) | |  |  |  |  | no measure |  |
| 16-29 | often lonely | Furloughed | Twins UK |  |  |  |  |  | no representation |  |
| 16-29 | often lonely | Furloughed | Overall | 0.88 | 0.40 | 1.93 |  | 85.60 |  |  |
| 16-29 | often lonely | No longer employed | MCS | 0.65 | 0.35 | 1.18 | 49.64 |  |  | REML |
| 16-29 | often lonely | No longer employed | ALSPAC(G1) | |  |  |  |  | no measure |  |
| 16-29 | often lonely | No longer employed | NS |  |  |  |  |  | no representation |  |
| 16-29 | often lonely | No longer employed | BCS70 |  |  |  |  |  | no representation |  |
| 16-29 | often lonely | No longer employed | NCDS |  |  |  |  |  | no representation |  |
| 16-29 | often lonely | No longer employed | USOC | 2.56 | 1.46 | 4.48 | 50.36 |  |  |  |
| 16-29 | often lonely | No longer employed | ELSA |  |  |  |  |  | no representation |  |
| 16-29 | often lonely | No longer employed | GS |  |  |  |  |  | no representation |  |
| 16-29 | often lonely | No longer employed | ALSPAC(G0) | |  |  |  |  | no measure |  |
| 16-29 | often lonely | No longer employed | Twins UK |  |  |  |  |  | no representation |  |
| 16-29 | often lonely | No longer employed | Overall | 1.29 | 0.34 | 4.98 |  | 90.64 |  |  |
| 16-29 | often lonely | Stable unemployed | MCS | 1.04 | 0.65 | 1.66 | 88.31 |  |  | REML |
| 16-29 | often lonely | Stable unemployed | ALSPAC(G1) | |  |  |  |  | no measure |  |
| 16-29 | often lonely | Stable unemployed | NS |  |  |  |  |  | no representation |  |
| 16-29 | often lonely | Stable unemployed | BCS70 |  |  |  |  |  | no representation |  |
| 16-29 | often lonely | Stable unemployed | NCDS |  |  |  |  |  | no representation |  |
| 16-29 | often lonely | Stable unemployed | USOC | 0.53 | 0.15 | 1.94 | 11.69 |  |  |  |
| 16-29 | often lonely | Stable unemployed | ELSA |  |  |  |  |  | no representation |  |
| 16-29 | often lonely | Stable unemployed | GS |  |  |  |  |  | no representation |  |
| 16-29 | often lonely | Stable unemployed | ALSPAC(G0) | |  |  |  |  | no measure |  |
| 16-29 | often lonely | Stable unemployed | Twins UK |  |  |  |  |  | no representation |  |
| 16-29 | often lonely | Stable unemployed | Overall | 0.96 | 0.62 | 1.49 |  | 0.00 |  |  |
| 16-29 | high loneliness | Furloughed | MCS | 1.09 | 0.86 | 1.39 | 100.00 |  |  | REML |
| 16-29 | high loneliness | Furloughed | ALSPAC(G1) | |  |  |  |  | no measure |  |
| 16-29 | high loneliness | Furloughed | NS |  |  |  |  |  | no representation |  |
| 16-29 | high loneliness | Furloughed | BCS70 |  |  |  |  |  | no representation |  |
| 16-29 | high loneliness | Furloughed | NCDS |  |  |  |  |  | no representation |  |
| 16-29 | high loneliness | Furloughed | USOC |  |  |  |  |  | no measure |  |
| 16-29 | high loneliness | Furloughed | ELSA |  |  |  |  |  | no representation |  |
| 16-29 | high loneliness | Furloughed | GS |  |  |  |  |  | no representation |  |
| 16-29 | high loneliness | Furloughed | ALSPAC(G0) | |  |  |  |  | no measure |  |
| 16-29 | high loneliness | Furloughed | Twins UK |  |  |  |  |  | no representation |  |
| 16-29 | high loneliness | Furloughed | Overall | 1.09 | 0.86 | 1.39 |  |  |  |  |
| 16-29 | high loneliness | No longer employed | MCS | 1.14 | 0.75 | 1.74 | 100.00 |  |  | REML |
| 16-29 | high loneliness | No longer employed | ALSPAC(G1) | |  |  |  |  | no measure |  |
| 16-29 | high loneliness | No longer employed | NS |  |  |  |  |  | no representation |  |
| 16-29 | high loneliness | No longer employed | BCS70 |  |  |  |  |  | no representation |  |
| 16-29 | high loneliness | No longer employed | NCDS |  |  |  |  |  | no representation |  |
| 16-29 | high loneliness | No longer employed | USOC |  |  |  |  |  | no measure |  |
| 16-29 | high loneliness | No longer employed | ELSA |  |  |  |  |  | no representation |  |
| 16-29 | high loneliness | No longer employed | GS |  |  |  |  |  | no representation |  |
| 16-29 | high loneliness | No longer employed | ALSPAC(G0) | |  |  |  |  | no measure |  |
| 16-29 | high loneliness | No longer employed | Twins UK |  |  |  |  |  | no representation |  |
| 16-29 | high loneliness | No longer employed | Overall | 1.14 | 0.75 | 1.74 |  |  |  |  |
| 16-29 | high loneliness | Stable unemployed | MCS | 0.86 | 0.54 | 1.36 | 100.00 |  |  | REML |
| 16-29 | high loneliness | Stable unemployed | ALSPAC(G1) | |  |  |  |  | no measure |  |
| 16-29 | high loneliness | Stable unemployed | NS |  |  |  |  |  | no representation |  |
| 16-29 | high loneliness | Stable unemployed | BCS70 |  |  |  |  |  | no representation |  |
| 16-29 | high loneliness | Stable unemployed | NCDS |  |  |  |  |  | no representation |  |
| 16-29 | high loneliness | Stable unemployed | USOC |  |  |  |  |  | no measure |  |
| 16-29 | high loneliness | Stable unemployed | ELSA |  |  |  |  |  | no representation |  |
| 16-29 | high loneliness | Stable unemployed | GS |  |  |  |  |  | no representation |  |
| 16-29 | high loneliness | Stable unemployed | ALSPAC(G0) | |  |  |  |  | no measure |  |
| 16-29 | high loneliness | Stable unemployed | Twins UK |  |  |  |  |  | no representation |  |
| 16-29 | high loneliness | Stable unemployed | Overall | 0.86 | 0.54 | 1.36 |  |  |  |  |
| 16-29 | distressed (bin.) | Furloughed | MCS | 0.77 | 0.46 | 1.29 | 27.76 |  |  | REML |
| 16-29 | distressed (bin.) | Furloughed | ALSPAC(G1) | 1.62 | 1.14 | 2.31 | 34.46 |  |  |  |
| 16-29 | distressed (bin.) | Furloughed | NS |  |  |  |  |  | no representation |  |
| 16-29 | distressed (bin.) | Furloughed | BCS70 |  |  |  |  |  | no representation |  |
| 16-29 | distressed (bin.) | Furloughed | NCDS |  |  |  |  |  | no representation |  |
| 16-29 | distressed (bin.) | Furloughed | USOC | 0.89 | 0.68 | 1.18 | 37.77 |  |  |  |
| 16-29 | distressed (bin.) | Furloughed | ELSA |  |  |  |  |  | no representation |  |
| 16-29 | distressed (bin.) | Furloughed | GS |  |  |  |  |  | no representation |  |
| 16-29 | distressed (bin.) | Furloughed | ALSPAC(G0) | |  |  |  |  | no representation |  |
| 16-29 | distressed (bin.) | Furloughed | Twins UK |  |  |  |  |  | no representation |  |
| 16-29 | distressed (bin.) | Furloughed | Overall | 1.05 | 0.68 | 1.64 |  | 76.50 |  |  |
| 16-29 | distressed (bin.) | No longer employed | MCS | 1.08 | 0.59 | 1.98 | 15.68 |  |  | REML |
| 16-29 | distressed (bin.) | No longer employed | ALSPAC(G1) | 1.64 | 1.01 | 2.66 | 24.44 |  |  |  |
| 16-29 | distressed (bin.) | No longer employed | NS |  |  |  |  |  | no representation |  |
| 16-29 | distressed (bin.) | No longer employed | BCS70 |  |  |  |  |  | no representation |  |
| 16-29 | distressed (bin.) | No longer employed | NCDS |  |  |  |  |  | no representation |  |
| 16-29 | distressed (bin.) | No longer employed | USOC | 1.48 | 1.09 | 2.02 | 59.88 |  |  |  |
| 16-29 | distressed (bin.) | No longer employed | ELSA |  |  |  |  |  | no representation |  |
| 16-29 | distressed (bin.) | No longer employed | GS |  |  |  |  |  | no representation |  |
| 16-29 | distressed (bin.) | No longer employed | ALSPAC(G0) | |  |  |  |  | no representation |  |
| 16-29 | distressed (bin.) | No longer employed | Twins UK |  |  |  |  |  | no representation |  |
| 16-29 | distressed (bin.) | No longer employed | Overall | 1.45 | 1.14 | 1.84 |  | 0.00 |  |  |
| 16-29 | distressed (bin.) | Stable unemployed | MCS | 1.22 | 0.64 | 2.30 | 37.70 |  |  | REML |
| 16-29 | distressed (bin.) | Stable unemployed | ALSPAC(G1) | 1.28 | 0.69 | 2.37 | 40.74 |  |  |  |
| 16-29 | distressed (bin.) | Stable unemployed | NS |  |  |  |  |  | no representation |  |
| 16-29 | distressed (bin.) | Stable unemployed | BCS70 |  |  |  |  |  | no representation |  |
| 16-29 | distressed (bin.) | Stable unemployed | NCDS |  |  |  |  |  | no representation |  |
| 16-29 | distressed (bin.) | Stable unemployed | USOC | 1.07 | 0.46 | 2.49 | 21.57 |  |  |  |
| 16-29 | distressed (bin.) | Stable unemployed | ELSA |  |  |  |  |  | no representation |  |
| 16-29 | distressed (bin.) | Stable unemployed | GS |  |  |  |  |  | no representation |  |
| 16-29 | distressed (bin.) | Stable unemployed | ALSPAC(G0) | |  |  |  |  | no representation |  |
| 16-29 | distressed (bin.) | Stable unemployed | Twins UK |  |  |  |  |  | no representation |  |
| 16-29 | distressed (bin.) | Stable unemployed | Overall | 1.21 | 0.82 | 1.79 |  | 0.00 |  |  |
| 16-29 | distressed (cont.) | Furloughed | MCS | -0.02 | -0.19 | 0.16 | 32.38 |  |  | REML |
| 16-29 | distressed (cont.) | Furloughed | ALSPAC(G1) | 0.23 | 0.03 | 0.43 | 28.91 |  |  |  |
| 16-29 | distressed (cont.) | Furloughed | NS |  |  |  |  |  | no representation |  |
| 16-29 | distressed (cont.) | Furloughed | BCS70 |  |  |  |  |  | no representation |  |
| 16-29 | distressed (cont.) | Furloughed | NCDS |  |  |  |  |  | no representation |  |
| 16-29 | distressed (cont.) | Furloughed | USOC | -0.03 | -0.16 | 0.11 | 38.71 |  |  |  |
| 16-29 | distressed (cont.) | Furloughed | ELSA |  |  |  |  |  | no representation |  |
| 16-29 | distressed (cont.) | Furloughed | GS |  |  |  |  |  | no representation |  |
| 16-29 | distressed (cont.) | Furloughed | ALSPAC(G0) | |  |  |  |  | no representation |  |
| 16-29 | distressed (cont.) | Furloughed | Twins UK |  |  |  |  |  | no representation |  |
| 16-29 | distressed (cont.) | Furloughed | Overall | 0.05 | -0.11 | 0.20 |  | 60.71 |  |  |
| 16-29 | distressed (cont.) | No longer employed | MCS | -0.10 | -0.36 | 0.17 | 34.00 |  |  | REML |
| 16-29 | distressed (cont.) | No longer employed | ALSPAC(G1) | 0.22 | -0.07 | 0.52 | 32.42 |  |  |  |
| 16-29 | distressed (cont.) | No longer employed | NS |  |  |  |  |  | no representation |  |
| 16-29 | distressed (cont.) | No longer employed | BCS70 |  |  |  |  |  | no representation |  |
| 16-29 | distressed (cont.) | No longer employed | NCDS |  |  |  |  |  | no representation |  |
| 16-29 | distressed (cont.) | No longer employed | USOC | 0.49 | 0.22 | 0.76 | 33.58 |  |  |  |
| 16-29 | distressed (cont.) | No longer employed | ELSA |  |  |  |  |  | no representation |  |
| 16-29 | distressed (cont.) | No longer employed | GS |  |  |  |  |  | no representation |  |
| 16-29 | distressed (cont.) | No longer employed | ALSPAC(G0) | |  |  |  |  | no representation |  |
| 16-29 | distressed (cont.) | No longer employed | Twins UK |  |  |  |  |  | no representation |  |
| 16-29 | distressed (cont.) | No longer employed | Overall | 0.20 | -0.13 | 0.54 |  | 77.38 |  |  |
| 16-29 | distressed (cont.) | Stable unemployed | MCS | 0.17 | -0.11 | 0.46 | 38.60 |  |  | REML |
| 16-29 | distressed (cont.) | Stable unemployed | ALSPAC(G1) | -0.15 | -0.45 | 0.14 | 36.93 |  |  |  |
| 16-29 | distressed (cont.) | Stable unemployed | NS |  |  |  |  |  | no representation |  |
| 16-29 | distressed (cont.) | Stable unemployed | BCS70 |  |  |  |  |  | no representation |  |
| 16-29 | distressed (cont.) | Stable unemployed | NCDS |  |  |  |  |  | no representation |  |
| 16-29 | distressed (cont.) | Stable unemployed | USOC | 0.28 | -0.14 | 0.70 | 24.47 |  |  |  |
| 16-29 | distressed (cont.) | Stable unemployed | ELSA |  |  |  |  |  | no representation |  |
| 16-29 | distressed (cont.) | Stable unemployed | GS |  |  |  |  |  | no representation |  |
| 16-29 | distressed (cont.) | Stable unemployed | ALSPAC(G0) | |  |  |  |  | no representation |  |
| 16-29 | distressed (cont.) | Stable unemployed | Twins UK |  |  |  |  |  | no representation |  |
| 16-29 | distressed (cont.) | Stable unemployed | Overall | 0.08 | -0.17 | 0.34 |  | 45.61 |  |  |
| 16-29 | less than daily contact | Furloughed | MCS | 0.85 | 0.68 | 1.05 | 33.82 |  |  | REML |
| 16-29 | less than daily contact | Furloughed | ALSPAC(G1) | 1.53 | 0.86 | 2.74 | 7.24 |  |  |  |
| 16-29 | less than daily contact | Furloughed | NS |  |  |  |  |  | no representation |  |
| 16-29 | less than daily contact | Furloughed | BCS70 |  |  |  |  |  | no representation |  |
| 16-29 | less than daily contact | Furloughed | NCDS |  |  |  |  |  | no representation |  |
| 16-29 | less than daily contact | Furloughed | USOC | 1.01 | 0.89 | 1.13 | 58.95 |  |  |  |
| 16-29 | less than daily contact | Furloughed | ELSA |  |  |  |  |  | no representation |  |
| 16-29 | less than daily contact | Furloughed | GS |  |  |  |  |  | no representation |  |
| 16-29 | less than daily contact | Furloughed | ALSPAC(G0) | |  |  |  |  | no representation |  |
| 16-29 | less than daily contact | Furloughed | Twins UK |  |  |  |  |  | no representation |  |
| 16-29 | less than daily contact | Furloughed | Overall | 0.98 | 0.83 | 1.15 |  | 36.46 |  |  |
| 16-29 | less than daily contact | No longer employed | MCS | 0.97 | 0.73 | 1.29 | 37.03 |  |  | REML |
| 16-29 | less than daily contact | No longer employed | ALSPAC(G1) | 1.47 | 0.57 | 3.78 | 3.32 |  |  |  |
| 16-29 | less than daily contact | No longer employed | NS |  |  |  |  |  | no representation |  |
| 16-29 | less than daily contact | No longer employed | BCS70 |  |  |  |  |  | no representation |  |
| 16-29 | less than daily contact | No longer employed | NCDS |  |  |  |  |  | no representation |  |
| 16-29 | less than daily contact | No longer employed | USOC | 0.95 | 0.76 | 1.19 | 59.65 |  |  |  |
| 16-29 | less than daily contact | No longer employed | ELSA |  |  |  |  |  | no representation |  |
| 16-29 | less than daily contact | No longer employed | GS |  |  |  |  |  | no representation |  |
| 16-29 | less than daily contact | No longer employed | ALSPAC(G0) | |  |  |  |  | no representation |  |
| 16-29 | less than daily contact | No longer employed | Twins UK |  |  |  |  |  | no representation |  |
| 16-29 | less than daily contact | No longer employed | Overall | 0.97 | 0.82 | 1.16 |  | 0.00 |  |  |
| 16-29 | less than daily contact | Stable unemployed | MCS | 0.87 | 0.58 | 1.29 | 35.17 |  |  | REML |
| 16-29 | less than daily contact | Stable unemployed | ALSPAC(G1) | 2.91 | 1.35 | 6.29 | 25.65 |  |  |  |
| 16-29 | less than daily contact | Stable unemployed | NS |  |  |  |  |  | no representation |  |
| 16-29 | less than daily contact | Stable unemployed | BCS70 |  |  |  |  |  | no representation |  |
| 16-29 | less than daily contact | Stable unemployed | NCDS |  |  |  |  |  | no representation |  |
| 16-29 | less than daily contact | Stable unemployed | USOC | 1.02 | 0.83 | 1.24 | 39.18 |  |  |  |
| 16-29 | less than daily contact | Stable unemployed | ELSA |  |  |  |  |  | no representation |  |
| 16-29 | less than daily contact | Stable unemployed | GS |  |  |  |  |  | no representation |  |
| 16-29 | less than daily contact | Stable unemployed | ALSPAC(G0) | |  |  |  |  | no representation |  |
| 16-29 | less than daily contact | Stable unemployed | Twins UK |  |  |  |  |  | no representation |  |
| 16-29 | less than daily contact | Stable unemployed | Overall | 1.26 | 0.66 | 2.39 |  | 86.66 |  |  |
| 16-29 | fair or poor self-rated health | Furloughed | MCS | 1.35 | 0.60 | 3.05 | 100.00 |  |  | MLE |
| 16-29 | fair or poor self-rated health | Furloughed | ALSPAC(G1) | |  |  |  |  | no measure |  |
| 16-29 | fair or poor self-rated health | Furloughed | NS |  |  |  |  |  | no representation |  |
| 16-29 | fair or poor self-rated health | Furloughed | BCS70 |  |  |  |  |  | no representation |  |
| 16-29 | fair or poor self-rated health | Furloughed | NCDS |  |  |  |  |  | no representation |  |
| 16-29 | fair or poor self-rated health | Furloughed | USOC |  |  |  |  |  | no measure |  |
| 16-29 | fair or poor self-rated health | Furloughed | ELSA |  |  |  |  |  | no representation |  |
| 16-29 | fair or poor self-rated health | Furloughed | GS |  |  |  |  |  | no representation |  |
| 16-29 | fair or poor self-rated health | Furloughed | ALSPAC(G0) | |  |  |  |  | no measure |  |
| 16-29 | fair or poor self-rated health | Furloughed | Twins UK |  |  |  |  |  | no representation |  |
| 16-29 | fair or poor self-rated health | Furloughed | Overall | 1.35 | 0.60 | 3.05 |  |  |  |  |

# 10. 30-49

| **Adjustment** | **Outcome** | **Exposure** | **Study** | **Coefficient** | **lower_ci** | **upper_ci** | **%Weight** | **%I2** | **Reason for missing** | **Method** |
| --- | --- | --- | --- | --- | --- | --- | --- | --- | --- | --- |
| 30-49 | low life satisfaction | Furloughed | MCS |  |  |  |  |  | no representation | REML |
| 30-49 | low life satisfaction | Furloughed | ALSPAC(G1) | |  |  |  |  | no representation |  |
| 30-49 | low life satisfaction | Furloughed | NS | 1.01 | 0.76 | 1.35 | 20.80 |  |  |  |
| 30-49 | low life satisfaction | Furloughed | BCS70 |  |  |  |  |  | no representation |  |
| 30-49 | low life satisfaction | Furloughed | NCDS |  |  |  |  |  | no representation |  |
| 30-49 | low life satisfaction | Furloughed | USOC | 0.98 | 0.81 | 1.19 | 39.13 |  |  |  |
| 30-49 | low life satisfaction | Furloughed | ELSA |  |  |  |  |  | no representation |  |
| 30-49 | low life satisfaction | Furloughed | GS | 1.12 | 0.90 | 1.40 | 32.20 |  |  |  |
| 30-49 | low life satisfaction | Furloughed | ALSPAC(G0) | |  |  |  |  | no representation |  |
| 30-49 | low life satisfaction | Furloughed | Twins UK | 1.90 | 1.15 | 3.14 | 7.87 |  |  |  |
| 30-49 | low life satisfaction | Furloughed | Overall | 1.09 | 0.94 | 1.26 |  | 20.35 |  |  |
| 30-49 | low life satisfaction | No longer employed | MCS |  |  |  |  |  | no representation | REML |
| 30-49 | low life satisfaction | No longer employed | ALSPAC(G1) | |  |  |  |  | no representation |  |
| 30-49 | low life satisfaction | No longer employed | NS | 1.24 | 0.79 | 1.94 | 22.01 |  |  |  |
| 30-49 | low life satisfaction | No longer employed | BCS70 |  |  |  |  |  | no representation |  |
| 30-49 | low life satisfaction | No longer employed | NCDS |  |  |  |  |  | no representation |  |
| 30-49 | low life satisfaction | No longer employed | USOC | 1.12 | 0.84 | 1.48 | 54.90 |  |  |  |
| 30-49 | low life satisfaction | No longer employed | ELSA |  |  |  |  |  | no representation |  |
| 30-49 | low life satisfaction | No longer employed | GS | 1.04 | 0.67 | 1.62 | 23.09 |  |  |  |
| 30-49 | low life satisfaction | No longer employed | ALSPAC(G0) | |  |  |  |  | no representation |  |
| 30-49 | low life satisfaction | No longer employed | Twins UK |  |  |  |  |  | low counts |  |
| 30-49 | low life satisfaction | No longer employed | Overall | 1.12 | 0.91 | 1.39 |  | 0.00 |  |  |
| 30-49 | low life satisfaction | Stable unemployed | MCS |  |  |  |  |  | no representation | REML |
| 30-49 | low life satisfaction | Stable unemployed | ALSPAC(G1) | |  |  |  |  | no representation |  |
| 30-49 | low life satisfaction | Stable unemployed | NS | 1.64 | 1.14 | 2.36 | 40.86 |  |  |  |
| 30-49 | low life satisfaction | Stable unemployed | BCS70 |  |  |  |  |  | no representation |  |
| 30-49 | low life satisfaction | Stable unemployed | NCDS |  |  |  |  |  | no representation |  |
| 30-49 | low life satisfaction | Stable unemployed | USOC | 1.54 | 1.14 | 2.08 | 59.14 |  |  |  |
| 30-49 | low life satisfaction | Stable unemployed | ELSA |  |  |  |  |  | no representation |  |
| 30-49 | low life satisfaction | Stable unemployed | GS |  |  |  |  |  |  |  |
| 30-49 | low life satisfaction | Stable unemployed | ALSPAC(G0) | |  |  |  |  | no representation |  |
| 30-49 | low life satisfaction | Stable unemployed | Twins UK |  |  |  |  |  | low counts |  |
| 30-49 | low life satisfaction | Stable unemployed | Overall | 1.58 | 1.25 | 1.99 |  | 0.00 |  |  |
| 30-49 | often lonely | Furloughed | MCS |  |  |  |  |  | no representation | REML |
| 30-49 | often lonely | Furloughed | ALSPAC(G1) | |  |  |  |  | no representation |  |
| 30-49 | often lonely | Furloughed | NS | 1.40 | 0.88 | 2.23 | 49.61 |  |  |  |
| 30-49 | often lonely | Furloughed | BCS70 |  |  |  |  |  | no representation |  |
| 30-49 | often lonely | Furloughed | NCDS |  |  |  |  |  | no representation |  |
| 30-49 | often lonely | Furloughed | USOC | 1.32 | 0.83 | 2.09 | 50.39 |  |  |  |
| 30-49 | often lonely | Furloughed | ELSA |  |  |  |  |  | no representation |  |
| 30-49 | often lonely | Furloughed | GS |  |  |  |  |  |  |  |
| 30-49 | often lonely | Furloughed | ALSPAC(G0) | |  |  |  |  | no representation |  |
| 30-49 | often lonely | Furloughed | Twins UK |  |  |  |  |  | low counts |  |
| 30-49 | often lonely | Furloughed | Overall | 1.36 | 0.98 | 1.88 |  | 0.00 |  |  |
| 30-49 | often lonely | No longer employed | MCS |  |  |  |  |  | no representation | REML |
| 30-49 | often lonely | No longer employed | ALSPAC(G1) | |  |  |  |  | no representation |  |
| 30-49 | often lonely | No longer employed | NS | 1.24 | 0.61 | 2.54 | 36.68 |  |  |  |
| 30-49 | often lonely | No longer employed | BCS70 |  |  |  |  |  | no representation |  |
| 30-49 | often lonely | No longer employed | NCDS |  |  |  |  |  | no representation |  |
| 30-49 | often lonely | No longer employed | USOC | 1.44 | 0.80 | 2.63 | 52.87 |  |  |  |
| 30-49 | often lonely | No longer employed | ELSA |  |  |  |  |  | no representation |  |
| 30-49 | often lonely | No longer employed | GS | 4.57 | 1.19 | 17.48 | 10.46 |  |  |  |
| 30-49 | often lonely | No longer employed | ALSPAC(G0) | |  |  |  |  | no representation |  |
| 30-49 | often lonely | No longer employed | Twins UK |  |  |  |  |  | low counts |  |
| 30-49 | often lonely | No longer employed | Overall | 1.54 | 1.00 | 2.38 |  | 0.00 |  |  |
| 30-49 | often lonely | Stable unemployed | MCS |  |  |  |  |  | no representation | REML |
| 30-49 | often lonely | Stable unemployed | ALSPAC(G1) | |  |  |  |  | no representation |  |
| 30-49 | often lonely | Stable unemployed | NS | 1.70 | 0.85 | 3.37 | 74.05 |  |  |  |
| 30-49 | often lonely | Stable unemployed | BCS70 |  |  |  |  |  | no representation |  |
| 30-49 | often lonely | Stable unemployed | NCDS |  |  |  |  |  | no representation |  |
| 30-49 | often lonely | Stable unemployed | USOC | 1.15 | 0.36 | 3.68 | 25.95 |  |  |  |
| 30-49 | often lonely | Stable unemployed | ELSA |  |  |  |  |  | no representation |  |
| 30-49 | often lonely | Stable unemployed | GS |  |  |  |  |  | low counts |  |
| 30-49 | often lonely | Stable unemployed | ALSPAC(G0) | |  |  |  |  | no representation |  |
| 30-49 | often lonely | Stable unemployed | Twins UK |  |  |  |  |  | low counts |  |
| 30-49 | often lonely | Stable unemployed | Overall | 1.53 | 0.85 | 2.77 |  | 0.00 |  |  |
| 30-49 | high loneliness | Furloughed | MCS |  |  |  |  |  | no representation | REML |
| 30-49 | high loneliness | Furloughed | ALSPAC(G1) | |  |  |  |  | no representation |  |
| 30-49 | high loneliness | Furloughed | NS | 1.23 | 0.92 | 1.63 | 76.49 |  |  |  |
| 30-49 | high loneliness | Furloughed | BCS70 |  |  |  |  |  | no representation |  |
| 30-49 | high loneliness | Furloughed | NCDS |  |  |  |  |  | no representation |  |
| 30-49 | high loneliness | Furloughed | USOC |  |  |  |  |  |  |  |
| 30-49 | high loneliness | Furloughed | ELSA |  |  |  |  |  | no representation |  |
| 30-49 | high loneliness | Furloughed | GS |  |  |  |  |  | no measure |  |
| 30-49 | high loneliness | Furloughed | ALSPAC(G0) | |  |  |  |  | no representation |  |
| 30-49 | high loneliness | Furloughed | Twins UK | 1.08 | 0.65 | 1.80 | 23.51 |  |  |  |
| 30-49 | high loneliness | Furloughed | Overall | 1.19 | 0.93 | 1.53 |  | 0.00 |  |  |
| 30-49 | high loneliness | No longer employed | MCS |  |  |  |  |  | no representation | REML |
| 30-49 | high loneliness | No longer employed | ALSPAC(G1) | |  |  |  |  | no representation |  |
| 30-49 | high loneliness | No longer employed | NS | 1.18 | 0.76 | 1.82 | 64.33 |  |  |  |
| 30-49 | high loneliness | No longer employed | BCS70 |  |  |  |  |  | no representation |  |
| 30-49 | high loneliness | No longer employed | NCDS |  |  |  |  |  | no representation |  |
| 30-49 | high loneliness | No longer employed | USOC |  |  |  |  |  |  |  |
| 30-49 | high loneliness | No longer employed | ELSA |  |  |  |  |  | no representation |  |
| 30-49 | high loneliness | No longer employed | GS |  |  |  |  |  | no measure |  |
| 30-49 | high loneliness | No longer employed | ALSPAC(G0) | |  |  |  |  | no representation |  |
| 30-49 | high loneliness | No longer employed | Twins UK | 1.17 | 0.65 | 2.11 | 35.67 |  |  |  |
| 30-49 | high loneliness | No longer employed | Overall | 1.17 | 0.83 | 1.67 |  | 0.00 |  |  |
| 30-49 | high loneliness | Stable unemployed | MCS |  |  |  |  |  | no representation | REML |
| 30-49 | high loneliness | Stable unemployed | ALSPAC(G1) | |  |  |  |  | no representation |  |
| 30-49 | high loneliness | Stable unemployed | NS | 1.10 | 0.70 | 1.74 | 100.00 |  |  |  |
| 30-49 | high loneliness | Stable unemployed | BCS70 |  |  |  |  |  | no representation |  |
| 30-49 | high loneliness | Stable unemployed | NCDS |  |  |  |  |  | no representation |  |
| 30-49 | high loneliness | Stable unemployed | USOC |  |  |  |  |  |  |  |
| 30-49 | high loneliness | Stable unemployed | ELSA |  |  |  |  |  | no representation |  |
| 30-49 | high loneliness | Stable unemployed | GS |  |  |  |  |  | no measure |  |
| 30-49 | high loneliness | Stable unemployed | ALSPAC(G0) | |  |  |  |  | no representation |  |
| 30-49 | high loneliness | Stable unemployed | Twins UK |  |  |  |  |  | low counts |  |
| 30-49 | high loneliness | Stable unemployed | Overall | 1.10 | 0.70 | 1.74 |  |  |  |  |
| 30-49 | distressed (bin.) | Furloughed | MCS |  |  |  |  |  | no representation | REML |
| 30-49 | distressed (bin.) | Furloughed | ALSPAC(G1) | |  |  |  |  | no representation |  |
| 30-49 | distressed (bin.) | Furloughed | NS | 1.32 | 1.04 | 1.68 | 43.20 |  |  |  |
| 30-49 | distressed (bin.) | Furloughed | BCS70 |  |  |  |  |  | no representation |  |
| 30-49 | distressed (bin.) | Furloughed | NCDS |  |  |  |  |  | no representation |  |
| 30-49 | distressed (bin.) | Furloughed | USOC | 0.96 | 0.78 | 1.19 | 46.47 |  |  |  |
| 30-49 | distressed (bin.) | Furloughed | ELSA |  |  |  |  |  | no representation |  |
| 30-49 | distressed (bin.) | Furloughed | GS | 0.85 | 0.40 | 1.82 | 10.33 |  |  |  |
| 30-49 | distressed (bin.) | Furloughed | ALSPAC(G0) | |  |  |  |  | no representation |  |
| 30-49 | distressed (bin.) | Furloughed | Twins UK |  |  |  |  |  | low counts |  |
| 30-49 | distressed (bin.) | Furloughed | Overall | 1.09 | 0.83 | 1.42 |  | 53.92 |  |  |
| 30-49 | distressed (bin.) | No longer employed | MCS |  |  |  |  |  | no representation | REML |
| 30-49 | distressed (bin.) | No longer employed | ALSPAC(G1) | |  |  |  |  | no representation |  |
| 30-49 | distressed (bin.) | No longer employed | NS | 1.25 | 0.77 | 2.03 | 28.67 |  |  |  |
| 30-49 | distressed (bin.) | No longer employed | BCS70 |  |  |  |  |  | no representation |  |
| 30-49 | distressed (bin.) | No longer employed | NCDS |  |  |  |  |  | no representation |  |
| 30-49 | distressed (bin.) | No longer employed | USOC | 1.13 | 0.82 | 1.55 | 65.41 |  |  |  |
| 30-49 | distressed (bin.) | No longer employed | ELSA |  |  |  |  |  | no representation |  |
| 30-49 | distressed (bin.) | No longer employed | GS | 1.68 | 0.58 | 4.86 | 5.92 |  |  |  |
| 30-49 | distressed (bin.) | No longer employed | ALSPAC(G0) | |  |  |  |  | no representation |  |
| 30-49 | distressed (bin.) | No longer employed | Twins UK |  |  |  |  |  | low counts |  |
| 30-49 | distressed (bin.) | No longer employed | Overall | 1.19 | 0.92 | 1.54 |  | 0.00 |  |  |
| 30-49 | distressed (bin.) | Stable unemployed | MCS |  |  |  |  |  | no representation | REML |
| 30-49 | distressed (bin.) | Stable unemployed | ALSPAC(G1) | |  |  |  |  | no representation |  |
| 30-49 | distressed (bin.) | Stable unemployed | NS | 1.54 | 1.01 | 2.35 | 59.54 |  |  |  |
| 30-49 | distressed (bin.) | Stable unemployed | BCS70 |  |  |  |  |  | no representation |  |
| 30-49 | distressed (bin.) | Stable unemployed | NCDS |  |  |  |  |  | no representation |  |
| 30-49 | distressed (bin.) | Stable unemployed | USOC | 0.71 | 0.33 | 1.56 | 40.46 |  |  |  |
| 30-49 | distressed (bin.) | Stable unemployed | ELSA |  |  |  |  |  | no representation |  |
| 30-49 | distressed (bin.) | Stable unemployed | GS |  |  |  |  |  |  |  |
| 30-49 | distressed (bin.) | Stable unemployed | ALSPAC(G0) | |  |  |  |  | no representation |  |
| 30-49 | distressed (bin.) | Stable unemployed | Twins UK |  |  |  |  |  | low counts |  |
| 30-49 | distressed (bin.) | Stable unemployed | Overall | 1.13 | 0.54 | 2.37 |  | 65.38 |  |  |
| 30-49 | distressed (cont.) | Furloughed | MCS |  |  |  |  |  | no representation | REML |
| 30-49 | distressed (cont.) | Furloughed | ALSPAC(G1) | |  |  |  |  | no representation |  |
| 30-49 | distressed (cont.) | Furloughed | NS | 0.09 | -0.03 | 0.21 | 42.54 |  |  |  |
| 30-49 | distressed (cont.) | Furloughed | BCS70 |  |  |  |  |  | no representation |  |
| 30-49 | distressed (cont.) | Furloughed | NCDS |  |  |  |  |  | no representation |  |
| 30-49 | distressed (cont.) | Furloughed | USOC | -0.04 | -0.13 | 0.05 | 49.22 |  |  |  |
| 30-49 | distressed (cont.) | Furloughed | ELSA |  |  |  |  |  | no representation |  |
| 30-49 | distressed (cont.) | Furloughed | GS | -0.26 | -1.39 | 0.88 | 1.36 |  |  |  |
| 30-49 | distressed (cont.) | Furloughed | ALSPAC(G0) | |  |  |  |  | no representation |  |
| 30-49 | distressed (cont.) | Furloughed | Twins UK | 0.39 | -0.10 | 0.87 | 6.88 |  |  |  |
| 30-49 | distressed (cont.) | Furloughed | Overall | 0.04 | -0.09 | 0.18 |  | 47.85 |  |  |
| 30-49 | distressed (cont.) | No longer employed | MCS |  |  |  |  |  | no representation | REML |
| 30-49 | distressed (cont.) | No longer employed | ALSPAC(G1) | |  |  |  |  | no representation |  |
| 30-49 | distressed (cont.) | No longer employed | NS | 0.29 | 0.02 | 0.57 | 58.31 |  |  |  |
| 30-49 | distressed (cont.) | No longer employed | BCS70 |  |  |  |  |  | no representation |  |
| 30-49 | distressed (cont.) | No longer employed | NCDS |  |  |  |  |  | no representation |  |
| 30-49 | distressed (cont.) | No longer employed | USOC | 0.24 | -0.11 | 0.58 | 37.75 |  |  |  |
| 30-49 | distressed (cont.) | No longer employed | ELSA |  |  |  |  |  | no representation |  |
| 30-49 | distressed (cont.) | No longer employed | GS | 1.84 | -1.12 | 4.80 | 0.50 |  |  |  |
| 30-49 | distressed (cont.) | No longer employed | ALSPAC(G0) | |  |  |  |  | no representation |  |
| 30-49 | distressed (cont.) | No longer employed | Twins UK | 0.35 | -0.78 | 1.48 | 3.44 |  |  |  |
| 30-49 | distressed (cont.) | No longer employed | Overall | 0.28 | 0.07 | 0.49 |  | 0.00 |  |  |
| 30-49 | distressed (cont.) | Stable unemployed | MCS |  |  |  |  |  | no representation | REML |
| 30-49 | distressed (cont.) | Stable unemployed | ALSPAC(G1) | |  |  |  |  | no representation |  |
| 30-49 | distressed (cont.) | Stable unemployed | NS | 0.35 | -0.02 | 0.72 | 32.81 |  |  |  |
| 30-49 | distressed (cont.) | Stable unemployed | BCS70 |  |  |  |  |  | no representation |  |
| 30-49 | distressed (cont.) | Stable unemployed | NCDS |  |  |  |  |  | no representation |  |
| 30-49 | distressed (cont.) | Stable unemployed | USOC | -0.08 | -0.35 | 0.18 | 37.17 |  |  |  |
| 30-49 | distressed (cont.) | Stable unemployed | ELSA |  |  |  |  |  | no representation |  |
| 30-49 | distressed (cont.) | Stable unemployed | GS |  |  |  |  |  |  |  |
| 30-49 | distressed (cont.) | Stable unemployed | ALSPAC(G0) | |  |  |  |  | no representation |  |
| 30-49 | distressed (cont.) | Stable unemployed | Twins UK | 0.65 | 0.22 | 1.09 | 30.02 |  |  |  |
| 30-49 | distressed (cont.) | Stable unemployed | Overall | 0.28 | -0.15 | 0.71 |  | 77.39 |  |  |
| 30-49 | less than daily contact | Furloughed | MCS |  |  |  |  |  | no representation | REML |
| 30-49 | less than daily contact | Furloughed | ALSPAC(G1) | |  |  |  |  | no representation |  |
| 30-49 | less than daily contact | Furloughed | NS | 0.89 | 0.76 | 1.03 | 22.21 |  |  |  |
| 30-49 | less than daily contact | Furloughed | BCS70 |  |  |  |  |  | no representation |  |
| 30-49 | less than daily contact | Furloughed | NCDS |  |  |  |  |  | no representation |  |
| 30-49 | less than daily contact | Furloughed | USOC | 1.01 | 0.94 | 1.07 | 63.46 |  |  |  |
| 30-49 | less than daily contact | Furloughed | ELSA |  |  |  |  |  | no representation |  |
| 30-49 | less than daily contact | Furloughed | GS | 1.07 | 0.88 | 1.31 | 14.33 |  |  |  |
| 30-49 | less than daily contact | Furloughed | ALSPAC(G0) | |  |  |  |  | no representation |  |
| 30-49 | less than daily contact | Furloughed | Twins UK |  |  |  |  |  | low counts |  |
| 30-49 | less than daily contact | Furloughed | Overall | 0.99 | 0.91 | 1.07 |  | 25.43 |  |  |
| 30-49 | less than daily contact | No longer employed | MCS |  |  |  |  |  | no representation | REML |
| 30-49 | less than daily contact | No longer employed | ALSPAC(G1) | |  |  |  |  | no representation |  |
| 30-49 | less than daily contact | No longer employed | NS | 0.98 | 0.71 | 1.34 | 17.19 |  |  |  |
| 30-49 | less than daily contact | No longer employed | BCS70 |  |  |  |  |  | no representation |  |
| 30-49 | less than daily contact | No longer employed | NCDS |  |  |  |  |  | no representation |  |
| 30-49 | less than daily contact | No longer employed | USOC | 0.99 | 0.85 | 1.16 | 72.23 |  |  |  |
| 30-49 | less than daily contact | No longer employed | ELSA |  |  |  |  |  | no representation |  |
| 30-49 | less than daily contact | No longer employed | GS | 0.95 | 0.63 | 1.42 | 10.58 |  |  |  |
| 30-49 | less than daily contact | No longer employed | ALSPAC(G0) | |  |  |  |  | no representation |  |
| 30-49 | less than daily contact | No longer employed | Twins UK |  |  |  |  |  | low counts |  |
| 30-49 | less than daily contact | No longer employed | Overall | 0.98 | 0.86 | 1.12 |  | 0.00 |  |  |
| 30-49 | less than daily contact | Stable unemployed | MCS |  |  |  |  |  | no representation | REML |
| 30-49 | less than daily contact | Stable unemployed | ALSPAC(G1) | |  |  |  |  | no representation |  |
| 30-49 | less than daily contact | Stable unemployed | NS | 0.99 | 0.67 | 1.44 | 30.08 |  |  |  |
| 30-49 | less than daily contact | Stable unemployed | BCS70 |  |  |  |  |  | no representation |  |
| 30-49 | less than daily contact | Stable unemployed | NCDS |  |  |  |  |  | no representation |  |
| 30-49 | less than daily contact | Stable unemployed | USOC | 1.03 | 0.80 | 1.32 | 69.92 |  |  |  |
| 30-49 | less than daily contact | Stable unemployed | ELSA |  |  |  |  |  | no representation |  |
| 30-49 | less than daily contact | Stable unemployed | GS |  |  |  |  |  | low counts |  |
| 30-49 | less than daily contact | Stable unemployed | ALSPAC(G0) | |  |  |  |  | no representation |  |
| 30-49 | less than daily contact | Stable unemployed | Twins UK |  |  |  |  |  | low counts |  |
| 30-49 | less than daily contact | Stable unemployed | Overall | 1.02 | 0.82 | 1.25 |  | 0.00 |  |  |
| 30-49 | fair or poor self-rated health | Furloughed | MCS |  |  |  |  |  | no representation | MLE |
| 30-49 | fair or poor self-rated health | Furloughed | ALSPAC(G1) | |  |  |  |  | no representation |  |
| 30-49 | fair or poor self-rated health | Furloughed | NS | 1.03 | 0.65 | 1.65 | 81.64 |  |  |  |
| 30-49 | fair or poor self-rated health | Furloughed | BCS70 |  |  |  |  |  | no representation |  |
| 30-49 | fair or poor self-rated health | Furloughed | NCDS |  |  |  |  |  | no representation |  |
| 30-49 | fair or poor self-rated health | Furloughed | USOC |  |  |  |  |  | no measure |  |
| 30-49 | fair or poor self-rated health | Furloughed | ELSA |  |  |  |  |  | no representation |  |
| 30-49 | fair or poor self-rated health | Furloughed | GS | 2.06 | 0.77 | 5.51 | 18.36 |  |  |  |
| 30-49 | fair or poor self-rated health | Furloughed | ALSPAC(G0) | |  |  |  |  | no representation |  |
| 30-49 | fair or poor self-rated health | Furloughed | Twins UK |  |  |  |  |  | low counts |  |
| 30-49 | fair or poor self-rated health | Furloughed | Overall | 1.17 | 0.77 | 1.79 |  | 0.00 |  |  |
| 30-49 | fair or poor self-rated health | No longer employed | MCS |  |  |  |  |  | no representation | REML |
| 30-49 | fair or poor self-rated health | No longer employed | ALSPAC(G1) | |  |  |  |  | no representation |  |
| 30-49 | fair or poor self-rated health | No longer employed | NS | 3.39 | 1.28 | 9.00 | 67.37 |  |  |  |
| 30-49 | fair or poor self-rated health | No longer employed | BCS70 |  |  |  |  |  | no representation |  |
| 30-49 | fair or poor self-rated health | No longer employed | NCDS |  |  |  |  |  | no representation |  |
| 30-49 | fair or poor self-rated health | No longer employed | USOC |  |  |  |  |  | no measure |  |
| 30-49 | fair or poor self-rated health | No longer employed | ELSA |  |  |  |  |  | no representation |  |
| 30-49 | fair or poor self-rated health | No longer employed | GS | 2.00 | 0.49 | 8.12 | 32.63 |  |  |  |
| 30-49 | fair or poor self-rated health | No longer employed | ALSPAC(G0) | |  |  |  |  | no representation |  |
| 30-49 | fair or poor self-rated health | No longer employed | Twins UK |  |  |  |  |  | low counts |  |
| 30-49 | fair or poor self-rated health | No longer employed | Overall | 2.86 | 1.28 | 6.36 |  | 0.00 |  |  |
| 30-49 | fair or poor self-rated health | Stable unemployed | MCS |  |  |  |  |  | no representation | REML |
| 30-49 | fair or poor self-rated health | Stable unemployed | ALSPAC(G1) | |  |  |  |  | no representation |  |
| 30-49 | fair or poor self-rated health | Stable unemployed | NS | 2.00 | 1.03 | 3.86 | 100.00 |  |  |  |
| 30-49 | fair or poor self-rated health | Stable unemployed | BCS70 |  |  |  |  |  | no representation |  |
| 30-49 | fair or poor self-rated health | Stable unemployed | NCDS |  |  |  |  |  | no representation |  |
| 30-49 | fair or poor self-rated health | Stable unemployed | USOC |  |  |  |  |  | no measure |  |
| 30-49 | fair or poor self-rated health | Stable unemployed | ELSA |  |  |  |  |  | no representation |  |
| 30-49 | fair or poor self-rated health | Stable unemployed | GS |  |  |  |  |  |  |  |
| 30-49 | fair or poor self-rated health | Stable unemployed | ALSPAC(G0) | |  |  |  |  | no representation |  |
| 30-49 | fair or poor self-rated health | Stable unemployed | Twins UK |  |  |  |  |  | low counts |  |
| 30-49 | fair or poor self-rated health | Stable unemployed | Overall | 2.00 | 1.03 | 3.86 |  |  |  |  |

# 11. 50+

| **Adjustment** | **Outcome** | **Exposure** | **Study** | **Coefficient** | **lower_ci** | **upper_ci** | **%Weight** | **%I2** | **Reason for missing** | **Method** |
| --- | --- | --- | --- | --- | --- | --- | --- | --- | --- | --- |
| 50+ | low life satisfaction | Furloughed | MCS |  |  |  |  |  | no representation | REML |
| 50+ | low life satisfaction | Furloughed | ALSPAC(G1) | |  |  |  |  | no representation |  |
| 50+ | low life satisfaction | Furloughed | NS |  |  |  |  |  | no representation |  |
| 50+ | low life satisfaction | Furloughed | BCS70 | 1.31 | 1.11 | 1.53 | 25.33 |  |  |  |
| 50+ | low life satisfaction | Furloughed | NCDS | 1.16 | 0.93 | 1.43 | 14.13 |  |  |  |
| 50+ | low life satisfaction | Furloughed | USOC | 1.15 | 0.98 | 1.35 | 25.41 |  |  |  |
| 50+ | low life satisfaction | Furloughed | ELSA | 1.19 | 0.97 | 1.45 | 16.61 |  |  |  |
| 50+ | low life satisfaction | Furloughed | GS | 1.06 | 0.87 | 1.30 | 15.85 |  |  |  |
| 50+ | low life satisfaction | Furloughed | ALSPAC(G0) | |  |  |  |  | no representation |  |
| 50+ | low life satisfaction | Furloughed | Twins UK | 0.97 | 0.59 | 1.59 | 2.68 |  |  |  |
| 50+ | low life satisfaction | Furloughed | Overall | 1.17 | 1.08 | 1.27 |  | 0.00 |  |  |
| 50+ | low life satisfaction | No longer employed | MCS |  |  |  |  |  | no representation | REML |
| 50+ | low life satisfaction | No longer employed | ALSPAC(G1) | |  |  |  |  | no representation |  |
| 50+ | low life satisfaction | No longer employed | NS |  |  |  |  |  | no representation |  |
| 50+ | low life satisfaction | No longer employed | BCS70 | 1.08 | 0.63 | 1.84 | 9.20 |  |  |  |
| 50+ | low life satisfaction | No longer employed | NCDS | 1.82 | 1.37 | 2.41 | 20.97 |  |  |  |
| 50+ | low life satisfaction | No longer employed | USOC | 1.31 | 1.04 | 1.66 | 24.54 |  |  |  |
| 50+ | low life satisfaction | No longer employed | ELSA | 1.70 | 1.29 | 2.24 | 21.19 |  |  |  |
| 50+ | low life satisfaction | No longer employed | GS | 1.19 | 0.93 | 1.51 | 24.10 |  |  |  |
| 50+ | low life satisfaction | No longer employed | ALSPAC(G0) | |  |  |  |  | no representation |  |
| 50+ | low life satisfaction | No longer employed | Twins UK |  |  |  |  |  | low counts |  |
| 50+ | low life satisfaction | No longer employed | Overall | 1.42 | 1.18 | 1.71 |  | 51.04 |  |  |
| 50+ | low life satisfaction | Stable unemployed | MCS |  |  |  |  |  | no representation | REML |
| 50+ | low life satisfaction | Stable unemployed | ALSPAC(G1) | |  |  |  |  | no representation |  |
| 50+ | low life satisfaction | Stable unemployed | NS |  |  |  |  |  | no representation |  |
| 50+ | low life satisfaction | Stable unemployed | BCS70 | 0.90 | 0.46 | 1.76 | 14.52 |  |  |  |
| 50+ | low life satisfaction | Stable unemployed | NCDS | 2.39 | 1.83 | 3.12 | 24.39 |  |  |  |
| 50+ | low life satisfaction | Stable unemployed | USOC | 1.17 | 0.85 | 1.60 | 23.29 |  |  |  |
| 50+ | low life satisfaction | Stable unemployed | ELSA | 1.64 | 1.23 | 2.20 | 23.77 |  |  |  |
| 50+ | low life satisfaction | Stable unemployed | GS | 0.91 | 0.45 | 1.82 | 14.03 |  |  |  |
| 50+ | low life satisfaction | Stable unemployed | ALSPAC(G0) | |  |  |  |  | no representation |  |
| 50+ | low life satisfaction | Stable unemployed | Twins UK |  |  |  |  |  | low counts |  |
| 50+ | low life satisfaction | Stable unemployed | Overall | 1.40 | 0.97 | 2.03 |  | 77.89 |  |  |
| 50+ | often lonely | Furloughed | MCS |  |  |  |  |  | no representation | REML |
| 50+ | often lonely | Furloughed | ALSPAC(G1) | |  |  |  |  | no representation |  |
| 50+ | often lonely | Furloughed | NS |  |  |  |  |  | no representation |  |
| 50+ | often lonely | Furloughed | BCS70 | 1.16 | 0.74 | 1.81 | 25.86 |  |  |  |
| 50+ | often lonely | Furloughed | NCDS | 0.84 | 0.51 | 1.38 | 23.46 |  |  |  |
| 50+ | often lonely | Furloughed | USOC | 2.21 | 1.33 | 3.68 | 23.12 |  |  |  |
| 50+ | often lonely | Furloughed | ELSA | 1.22 | 0.69 | 2.15 | 20.78 |  |  |  |
| 50+ | often lonely | Furloughed | GS | 1.35 | 0.37 | 4.87 | 6.78 |  |  |  |
| 50+ | often lonely | Furloughed | ALSPAC(G0) | |  |  |  |  | no representation |  |
| 50+ | often lonely | Furloughed | Twins UK |  |  |  |  |  | low counts |  |
| 50+ | often lonely | Furloughed | Overall | 1.27 | 0.88 | 1.83 |  | 49.76 |  |  |
| 50+ | often lonely | No longer employed | MCS |  |  |  |  |  | no representation | REML |
| 50+ | often lonely | No longer employed | ALSPAC(G1) | |  |  |  |  | no representation |  |
| 50+ | often lonely | No longer employed | NS |  |  |  |  |  | no representation |  |
| 50+ | often lonely | No longer employed | BCS70 | 3.87 | 2.00 | 7.49 | 32.15 |  |  |  |
| 50+ | often lonely | No longer employed | NCDS | 2.76 | 1.30 | 5.89 | 24.55 |  |  |  |
| 50+ | often lonely | No longer employed | USOC | 2.31 | 1.16 | 4.60 | 29.64 |  |  |  |
| 50+ | often lonely | No longer employed | ELSA | 1.15 | 0.37 | 3.63 | 10.64 |  |  |  |
| 50+ | often lonely | No longer employed | GS | 2.37 | 0.27 | 20.39 | 3.02 |  |  |  |
| 50+ | often lonely | No longer employed | ALSPAC(G0) | |  |  |  |  | no representation |  |
| 50+ | often lonely | No longer employed | Twins UK |  |  |  |  |  | low counts |  |
| 50+ | often lonely | No longer employed | Overall | 2.65 | 1.82 | 3.85 |  | 0.00 |  |  |
| 50+ | often lonely | Stable unemployed | MCS |  |  |  |  |  | no representation | REML |
| 50+ | often lonely | Stable unemployed | ALSPAC(G1) | |  |  |  |  | no representation |  |
| 50+ | often lonely | Stable unemployed | NS |  |  |  |  |  | no representation |  |
| 50+ | often lonely | Stable unemployed | BCS70 | 0.72 | 0.19 | 2.69 | 13.35 |  |  |  |
| 50+ | often lonely | Stable unemployed | NCDS | 2.73 | 1.51 | 4.91 | 39.71 |  |  |  |
| 50+ | often lonely | Stable unemployed | USOC | 1.41 | 0.56 | 3.54 | 23.18 |  |  |  |
| 50+ | often lonely | Stable unemployed | ELSA | 1.46 | 0.59 | 3.61 | 23.75 |  |  |  |
| 50+ | often lonely | Stable unemployed | GS |  |  |  |  |  |  |  |
| 50+ | often lonely | Stable unemployed | ALSPAC(G0) | |  |  |  |  | no representation |  |
| 50+ | often lonely | Stable unemployed | Twins UK |  |  |  |  |  | low counts |  |
| 50+ | often lonely | Stable unemployed | Overall | 1.69 | 0.99 | 2.87 |  | 31.77 |  |  |
| 50+ | high loneliness | Furloughed | MCS |  |  |  |  |  | no representation | REML |
| 50+ | high loneliness | Furloughed | ALSPAC(G1) | |  |  |  |  | no representation |  |
| 50+ | high loneliness | Furloughed | NS |  |  |  |  |  | no representation |  |
| 50+ | high loneliness | Furloughed | BCS70 | 1.26 | 1.03 | 1.54 | 30.82 |  |  |  |
| 50+ | high loneliness | Furloughed | NCDS | 1.12 | 0.87 | 1.42 | 27.45 |  |  |  |
| 50+ | high loneliness | Furloughed | USOC |  |  |  |  |  | no measure |  |
| 50+ | high loneliness | Furloughed | ELSA | 0.94 | 0.72 | 1.24 | 25.73 |  |  |  |
| 50+ | high loneliness | Furloughed | GS |  |  |  |  |  | no measure |  |
| 50+ | high loneliness | Furloughed | ALSPAC(G0) | |  |  |  |  | no representation |  |
| 50+ | high loneliness | Furloughed | Twins UK | 0.66 | 0.43 | 1.03 | 16.00 |  |  |  |
| 50+ | high loneliness | Furloughed | Overall | 1.02 | 0.81 | 1.28 |  | 64.49 |  |  |
| 50+ | high loneliness | No longer employed | MCS |  |  |  |  |  | no representation | REML |
| 50+ | high loneliness | No longer employed | ALSPAC(G1) | |  |  |  |  | no representation |  |
| 50+ | high loneliness | No longer employed | NS |  |  |  |  |  | no representation |  |
| 50+ | high loneliness | No longer employed | BCS70 | 0.77 | 0.39 | 1.50 | 16.23 |  |  |  |
| 50+ | high loneliness | No longer employed | NCDS | 1.36 | 0.89 | 2.07 | 36.20 |  |  |  |
| 50+ | high loneliness | No longer employed | USOC |  |  |  |  |  | no measure |  |
| 50+ | high loneliness | No longer employed | ELSA | 0.90 | 0.55 | 1.47 | 28.40 |  |  |  |
| 50+ | high loneliness | No longer employed | GS |  |  |  |  |  | no measure |  |
| 50+ | high loneliness | No longer employed | ALSPAC(G0) | |  |  |  |  | no representation |  |
| 50+ | high loneliness | No longer employed | Twins UK | 0.87 | 0.47 | 1.60 | 19.17 |  |  |  |
| 50+ | high loneliness | No longer employed | Overall | 1.01 | 0.76 | 1.34 |  | 14.16 |  |  |
| 50+ | high loneliness | Stable unemployed | MCS |  |  |  |  |  | no representation | REML |
| 50+ | high loneliness | Stable unemployed | ALSPAC(G1) | |  |  |  |  | no representation |  |
| 50+ | high loneliness | Stable unemployed | NS |  |  |  |  |  | no representation |  |
| 50+ | high loneliness | Stable unemployed | BCS70 | 0.49 | 0.16 | 1.53 | 8.68 |  |  |  |
| 50+ | high loneliness | Stable unemployed | NCDS | 1.66 | 1.16 | 2.36 | 38.10 |  |  |  |
| 50+ | high loneliness | Stable unemployed | USOC |  |  |  |  |  | no measure |  |
| 50+ | high loneliness | Stable unemployed | ELSA | 1.14 | 0.71 | 1.82 | 30.09 |  |  |  |
| 50+ | high loneliness | Stable unemployed | GS |  |  |  |  |  | no measure |  |
| 50+ | high loneliness | Stable unemployed | ALSPAC(G0) | |  |  |  |  | no representation |  |
| 50+ | high loneliness | Stable unemployed | Twins UK | 0.99 | 0.55 | 1.80 | 23.14 |  |  |  |
| 50+ | high loneliness | Stable unemployed | Overall | 1.18 | 0.82 | 1.70 |  | 43.42 |  |  |
| 50+ | distressed (bin.) | Furloughed | MCS |  |  |  |  |  | no representation | REML |
| 50+ | distressed (bin.) | Furloughed | ALSPAC(G1) | |  |  |  |  | no representation |  |
| 50+ | distressed (bin.) | Furloughed | NS |  |  |  |  |  | no representation |  |
| 50+ | distressed (bin.) | Furloughed | BCS70 | 1.30 | 1.04 | 1.62 | 29.61 |  |  |  |
| 50+ | distressed (bin.) | Furloughed | NCDS | 0.90 | 0.65 | 1.26 | 15.57 |  |  |  |
| 50+ | distressed (bin.) | Furloughed | USOC | 1.04 | 0.85 | 1.29 | 31.73 |  |  |  |
| 50+ | distressed (bin.) | Furloughed | ELSA | 1.00 | 0.75 | 1.34 | 19.54 |  |  |  |
| 50+ | distressed (bin.) | Furloughed | GS | 1.02 | 0.48 | 2.17 | 3.54 |  |  |  |
| 50+ | distressed (bin.) | Furloughed | ALSPAC(G0) | |  |  |  |  | no representation |  |
| 50+ | distressed (bin.) | Furloughed | Twins UK |  |  |  |  |  | low counts |  |
| 50+ | distressed (bin.) | Furloughed | Overall | 1.08 | 0.93 | 1.25 |  | 22.07 |  |  |
| 50+ | distressed (bin.) | No longer employed | MCS |  |  |  |  |  | no representation | REML |
| 50+ | distressed (bin.) | No longer employed | ALSPAC(G1) | |  |  |  |  | no representation |  |
| 50+ | distressed (bin.) | No longer employed | NS |  |  |  |  |  | no representation |  |
| 50+ | distressed (bin.) | No longer employed | BCS70 | 1.45 | 0.90 | 2.34 | 18.23 |  |  |  |
| 50+ | distressed (bin.) | No longer employed | NCDS | 1.84 | 1.02 | 3.32 | 11.84 |  |  |  |
| 50+ | distressed (bin.) | No longer employed | USOC | 1.17 | 0.86 | 1.60 | 43.37 |  |  |  |
| 50+ | distressed (bin.) | No longer employed | ELSA | 1.24 | 0.80 | 1.94 | 20.75 |  |  |  |
| 50+ | distressed (bin.) | No longer employed | GS | 1.91 | 0.82 | 4.45 | 5.80 |  |  |  |
| 50+ | distressed (bin.) | No longer employed | ALSPAC(G0) | |  |  |  |  | no representation |  |
| 50+ | distressed (bin.) | No longer employed | Twins UK |  |  |  |  |  | low counts |  |
| 50+ | distressed (bin.) | No longer employed | Overall | 1.34 | 1.09 | 1.64 |  | 0.00 |  |  |
| 50+ | distressed (bin.) | Stable unemployed | MCS |  |  |  |  |  | no representation | REML |
| 50+ | distressed (bin.) | Stable unemployed | ALSPAC(G1) | |  |  |  |  | no representation |  |
| 50+ | distressed (bin.) | Stable unemployed | NS |  |  |  |  |  | no representation |  |
| 50+ | distressed (bin.) | Stable unemployed | BCS70 | 0.64 | 0.18 | 2.25 | 7.55 |  |  |  |
| 50+ | distressed (bin.) | Stable unemployed | NCDS | 2.17 | 1.03 | 4.59 | 18.16 |  |  |  |
| 50+ | distressed (bin.) | Stable unemployed | USOC | 1.03 | 0.68 | 1.57 | 38.51 |  |  |  |
| 50+ | distressed (bin.) | Stable unemployed | ELSA | 1.55 | 0.99 | 2.43 | 35.77 |  |  |  |
| 50+ | distressed (bin.) | Stable unemployed | GS |  |  |  |  |  | low counts |  |
| 50+ | distressed (bin.) | Stable unemployed | ALSPAC(G0) | |  |  |  |  | no representation |  |
| 50+ | distressed (bin.) | Stable unemployed | Twins UK |  |  |  |  |  | low counts |  |
| 50+ | distressed (bin.) | Stable unemployed | Overall | 1.32 | 0.92 | 1.89 |  | 31.78 |  |  |
| 50+ | distressed (cont.) | Furloughed | MCS |  |  |  |  |  | no representation | REML |
| 50+ | distressed (cont.) | Furloughed | ALSPAC(G1) | |  |  |  |  | no representation |  |
| 50+ | distressed (cont.) | Furloughed | NS |  |  |  |  |  | no representation |  |
| 50+ | distressed (cont.) | Furloughed | BCS70 | 0.13 | 0.02 | 0.24 | 21.76 |  |  |  |
| 50+ | distressed (cont.) | Furloughed | NCDS | -0.02 | -0.13 | 0.09 | 22.08 |  |  |  |
| 50+ | distressed (cont.) | Furloughed | USOC | -0.05 | -0.11 | 0.02 | 29.80 |  |  |  |
| 50+ | distressed (cont.) | Furloughed | ELSA | 0.06 | -0.06 | 0.18 | 20.34 |  |  |  |
| 50+ | distressed (cont.) | Furloughed | GS | -0.14 | -0.84 | 0.56 | 1.26 |  |  |  |
| 50+ | distressed (cont.) | Furloughed | ALSPAC(G0) | |  |  |  |  | no representation |  |
| 50+ | distressed (cont.) | Furloughed | Twins UK | -0.27 | -0.62 | 0.07 | 4.76 |  |  |  |
| 50+ | distressed (cont.) | Furloughed | Overall | 0.01 | -0.07 | 0.09 |  | 52.55 |  |  |
| 50+ | distressed (cont.) | No longer employed | MCS |  |  |  |  |  | no representation | REML |
| 50+ | distressed (cont.) | No longer employed | ALSPAC(G1) | |  |  |  |  | no representation |  |
| 50+ | distressed (cont.) | No longer employed | NS |  |  |  |  |  | no representation |  |
| 50+ | distressed (cont.) | No longer employed | BCS70 | -0.02 | -0.27 | 0.24 | 19.30 |  |  |  |
| 50+ | distressed (cont.) | No longer employed | NCDS | 0.20 | -0.06 | 0.47 | 18.29 |  |  |  |
| 50+ | distressed (cont.) | No longer employed | USOC | 0.11 | -0.05 | 0.27 | 29.36 |  |  |  |
| 50+ | distressed (cont.) | No longer employed | ELSA | 0.12 | -0.13 | 0.37 | 19.53 |  |  |  |
| 50+ | distressed (cont.) | No longer employed | GS | 1.33 | -0.09 | 2.75 | 1.07 |  |  |  |
| 50+ | distressed (cont.) | No longer employed | ALSPAC(G0) | |  |  |  |  | no representation |  |
| 50+ | distressed (cont.) | No longer employed | Twins UK | -0.39 | -0.75 | -0.03 | 12.44 |  |  |  |
| 50+ | distressed (cont.) | No longer employed | Overall | 0.06 | -0.09 | 0.20 |  | 40.52 |  |  |
| 50+ | distressed (cont.) | Stable unemployed | MCS |  |  |  |  |  | no representation | REML |
| 50+ | distressed (cont.) | Stable unemployed | ALSPAC(G1) | |  |  |  |  | no representation |  |
| 50+ | distressed (cont.) | Stable unemployed | NS |  |  |  |  |  | no representation |  |
| 50+ | distressed (cont.) | Stable unemployed | BCS70 | -0.31 | -0.71 | 0.09 | 18.51 |  |  |  |
| 50+ | distressed (cont.) | Stable unemployed | NCDS | 0.35 | 0.00 | 0.70 | 21.03 |  |  |  |
| 50+ | distressed (cont.) | Stable unemployed | USOC | 0.09 | -0.12 | 0.29 | 29.55 |  |  |  |
| 50+ | distressed (cont.) | Stable unemployed | ELSA | 0.40 | 0.02 | 0.78 | 19.54 |  |  |  |
| 50+ | distressed (cont.) | Stable unemployed | GS | 1.33 | -1.77 | 4.43 | 0.60 |  |  |  |
| 50+ | distressed (cont.) | Stable unemployed | ALSPAC(G0) | |  |  |  |  | no representation |  |
| 50+ | distressed (cont.) | Stable unemployed | Twins UK | 0.26 | -0.36 | 0.89 | 10.77 |  |  |  |
| 50+ | distressed (cont.) | Stable unemployed | Overall | 0.16 | -0.08 | 0.40 |  | 50.48 |  |  |
| 50+ | less than daily contact | Furloughed | MCS |  |  |  |  |  | no representation | REML |
| 50+ | less than daily contact | Furloughed | ALSPAC(G1) | |  |  |  |  | no representation |  |
| 50+ | less than daily contact | Furloughed | NS |  |  |  |  |  | no representation |  |
| 50+ | less than daily contact | Furloughed | BCS70 | 0.91 | 0.84 | 0.99 | 23.93 |  |  |  |
| 50+ | less than daily contact | Furloughed | NCDS | 0.91 | 0.82 | 1.02 | 18.43 |  |  |  |
| 50+ | less than daily contact | Furloughed | USOC | 0.98 | 0.92 | 1.05 | 30.53 |  |  |  |
| 50+ | less than daily contact | Furloughed | ELSA | 1.06 | 0.95 | 1.18 | 18.45 |  |  |  |
| 50+ | less than daily contact | Furloughed | GS | 0.87 | 0.72 | 1.05 | 8.04 |  |  |  |
| 50+ | less than daily contact | Furloughed | ALSPAC(G0) | |  |  |  |  | no representation |  |
| 50+ | less than daily contact | Furloughed | Twins UK | 0.60 | 0.29 | 1.25 | 0.61 |  |  |  |
| 50+ | less than daily contact | Furloughed | Overall | 0.95 | 0.90 | 1.01 |  | 37.13 |  |  |
| 50+ | less than daily contact | No longer employed | MCS |  |  |  |  |  | no representation | REML |
| 50+ | less than daily contact | No longer employed | ALSPAC(G1) | |  |  |  |  | no representation |  |
| 50+ | less than daily contact | No longer employed | NS |  |  |  |  |  | no representation |  |
| 50+ | less than daily contact | No longer employed | BCS70 | 0.87 | 0.67 | 1.13 | 6.99 |  |  |  |
| 50+ | less than daily contact | No longer employed | NCDS | 0.97 | 0.81 | 1.17 | 13.49 |  |  |  |
| 50+ | less than daily contact | No longer employed | USOC | 1.00 | 0.92 | 1.09 | 62.44 |  |  |  |
| 50+ | less than daily contact | No longer employed | ELSA | 0.92 | 0.70 | 1.21 | 6.13 |  |  |  |
| 50+ | less than daily contact | No longer employed | GS | 1.14 | 0.92 | 1.40 | 10.96 |  |  |  |
| 50+ | less than daily contact | No longer employed | ALSPAC(G0) | |  |  |  |  | no representation |  |
| 50+ | less than daily contact | No longer employed | Twins UK |  |  |  |  |  | low counts |  |
| 50+ | less than daily contact | No longer employed | Overall | 1.00 | 0.93 | 1.07 |  | 0.00 |  |  |
| 50+ | less than daily contact | Stable unemployed | MCS |  |  |  |  |  | no representation | REML |
| 50+ | less than daily contact | Stable unemployed | ALSPAC(G1) | |  |  |  |  | no representation |  |
| 50+ | less than daily contact | Stable unemployed | NS |  |  |  |  |  | no representation |  |
| 50+ | less than daily contact | Stable unemployed | BCS70 | 0.99 | 0.57 | 1.73 | 9.48 |  |  |  |
| 50+ | less than daily contact | Stable unemployed | NCDS | 1.31 | 1.11 | 1.56 | 31.06 |  |  |  |
| 50+ | less than daily contact | Stable unemployed | USOC | 0.89 | 0.70 | 1.12 | 25.82 |  |  |  |
| 50+ | less than daily contact | Stable unemployed | ELSA | 0.92 | 0.70 | 1.22 | 22.53 |  |  |  |
| 50+ | less than daily contact | Stable unemployed | GS | 1.02 | 0.62 | 1.68 | 11.12 |  |  |  |
| 50+ | less than daily contact | Stable unemployed | ALSPAC(G0) | |  |  |  |  | no representation |  |
| 50+ | less than daily contact | Stable unemployed | Twins UK |  |  |  |  |  | low counts |  |
| 50+ | less than daily contact | Stable unemployed | Overall | 1.04 | 0.85 | 1.26 |  | 54.07 |  |  |
| 50+ | fair or poor self-rated health | Furloughed | MCS |  |  |  |  |  | no representation | MLE |
| 50+ | fair or poor self-rated health | Furloughed | ALSPAC(G1) | |  |  |  |  | no representation |  |
| 50+ | fair or poor self-rated health | Furloughed | NS |  |  |  |  |  | no representation |  |
| 50+ | fair or poor self-rated health | Furloughed | BCS70 | 1.39 | 1.06 | 1.83 | 25.20 |  |  |  |
| 50+ | fair or poor self-rated health | Furloughed | NCDS | 1.48 | 1.05 | 2.08 | 20.11 |  |  |  |
| 50+ | fair or poor self-rated health | Furloughed | USOC |  |  |  |  |  | no measure |  |
| 50+ | fair or poor self-rated health | Furloughed | ELSA | 1.05 | 1.00 | 1.09 | 46.02 |  |  |  |
| 50+ | fair or poor self-rated health | Furloughed | GS | 0.98 | 0.39 | 2.45 | 4.40 |  |  |  |
| 50+ | fair or poor self-rated health | Furloughed | ALSPAC(G0) | |  |  |  |  | no representation |  |
| 50+ | fair or poor self-rated health | Furloughed | Twins UK | 3.15 | 1.24 | 8.00 | 4.28 |  |  |  |
| 50+ | fair or poor self-rated health | Furloughed | Overall | 1.26 | 1.03 | 1.54 |  | 50.46 |  |  |
| 50+ | fair or poor self-rated health | No longer employed | MCS |  |  |  |  |  | no representation | REML |
| 50+ | fair or poor self-rated health | No longer employed | ALSPAC(G1) | |  |  |  |  | no representation |  |
| 50+ | fair or poor self-rated health | No longer employed | NS |  |  |  |  |  | no representation |  |
| 50+ | fair or poor self-rated health | No longer employed | BCS70 | 1.50 | 0.82 | 2.76 | 16.26 |  |  |  |
| 50+ | fair or poor self-rated health | No longer employed | NCDS | 1.44 | 0.81 | 2.57 | 17.46 |  |  |  |
| 50+ | fair or poor self-rated health | No longer employed | USOC |  |  |  |  |  | no measure |  |
| 50+ | fair or poor self-rated health | No longer employed | ELSA | 1.06 | 0.96 | 1.17 | 53.72 |  |  |  |
| 50+ | fair or poor self-rated health | No longer employed | GS | 1.93 | 0.94 | 3.98 | 12.57 |  |  |  |
| 50+ | fair or poor self-rated health | No longer employed | ALSPAC(G0) | |  |  |  |  | no representation |  |
| 50+ | fair or poor self-rated health | No longer employed | Twins UK |  |  |  |  |  | low counts |  |
| 50+ | fair or poor self-rated health | No longer employed | Overall | 1.28 | 0.95 | 1.71 |  | 41.53 |  |  |
| 50+ | fair or poor self-rated health | Stable unemployed | MCS |  |  |  |  |  | no representation | REML |
| 50+ | fair or poor self-rated health | Stable unemployed | ALSPAC(G1) | |  |  |  |  | no representation |  |
| 50+ | fair or poor self-rated health | Stable unemployed | NS |  |  |  |  |  | no representation |  |
| 50+ | fair or poor self-rated health | Stable unemployed | BCS70 | 3.90 | 1.10 | 13.76 | 14.54 |  |  |  |
| 50+ | fair or poor self-rated health | Stable unemployed | NCDS | 1.98 | 1.31 | 2.99 | 38.49 |  |  |  |
| 50+ | fair or poor self-rated health | Stable unemployed | USOC |  |  |  |  |  | no measure |  |
| 50+ | fair or poor self-rated health | Stable unemployed | ELSA | 1.14 | 1.01 | 1.28 | 46.96 |  |  |  |
| 50+ | fair or poor self-rated health | Stable unemployed | GS |  |  |  |  |  | low counts |  |
| 50+ | fair or poor self-rated health | Stable unemployed | ALSPAC(G0) | |  |  |  |  | no representation |  |
| 50+ | fair or poor self-rated health | Stable unemployed | Twins UK |  |  |  |  |  | low counts |  |
| 50+ | fair or poor self-rated health | Stable unemployed | Overall | 1.68 | 0.95 | 3.00 |  | 80.57 |  |  |

### 12. Alone

| **Adjustment** | **Outcome** | **Exposure** | **Study** | **Coefficient** | **lower_ci** | **upper_ci** | **%Weight** | **%I2** | **Reason for missing** | **Method** |
| --- | --- | --- | --- | --- | --- | --- | --- | --- | --- | --- |
| Alone | low life satisfaction | Furloughed | MCS |  |  |  |  |  | low counts | REML |
| Alone | low life satisfaction | Furloughed | ALSPAC(G1) | |  |  |  |  | no measure |  |
| Alone | low life satisfaction | Furloughed | NS | 1.41 | 0.98 | 2.03 | 16.83 |  |  |  |
| Alone | low life satisfaction | Furloughed | BCS70 | 0.95 | 0.66 | 1.38 | 16.63 |  |  |  |
| Alone | low life satisfaction | Furloughed | NCDS | 1.23 | 0.82 | 1.83 | 15.47 |  |  |  |
| Alone | low life satisfaction | Furloughed | USOC | 1.47 | 1.13 | 1.90 | 21.27 |  |  |  |
| Alone | low life satisfaction | Furloughed | ELSA | 1.87 | 1.22 | 2.88 | 14.40 |  |  |  |
| Alone | low life satisfaction | Furloughed | GS | 0.81 | 0.54 | 1.22 | 15.40 |  |  |  |
| Alone | low life satisfaction | Furloughed | ALSPAC(G0) | |  |  |  |  | no measure |  |
| Alone | low life satisfaction | Furloughed | Twins UK |  |  |  |  |  | low counts |  |
| Alone | low life satisfaction | Furloughed | Overall | 1.25 | 0.99 | 1.57 |  | 57.94 |  |  |
| Alone | low life satisfaction | No longer employed | MCS |  |  |  |  |  | low counts | REML |
| Alone | low life satisfaction | No longer employed | ALSPAC(G1) | |  |  |  |  | no measure |  |
| Alone | low life satisfaction | No longer employed | NS | 1.31 | 0.77 | 2.23 | 18.12 |  |  |  |
| Alone | low life satisfaction | No longer employed | BCS70 | 1.67 | 0.98 | 2.83 | 18.19 |  |  |  |
| Alone | low life satisfaction | No longer employed | NCDS | 1.75 | 0.96 | 3.20 | 15.91 |  |  |  |
| Alone | low life satisfaction | No longer employed | USOC | 1.50 | 0.86 | 2.64 | 17.09 |  |  |  |
| Alone | low life satisfaction | No longer employed | ELSA | 3.19 | 1.64 | 6.19 | 14.25 |  |  |  |
| Alone | low life satisfaction | No longer employed | GS | 0.83 | 0.46 | 1.49 | 16.45 |  |  |  |
| Alone | low life satisfaction | No longer employed | ALSPAC(G0) | |  |  |  |  | no measure |  |
| Alone | low life satisfaction | No longer employed | Twins UK |  |  |  |  |  | low counts |  |
| Alone | low life satisfaction | No longer employed | Overall | 1.55 | 1.12 | 2.13 |  | 46.83 |  |  |
| Alone | low life satisfaction | Stable unemployed | MCS |  |  |  |  |  | low counts | REML |
| Alone | low life satisfaction | Stable unemployed | ALSPAC(G1) | |  |  |  |  | no measure |  |
| Alone | low life satisfaction | Stable unemployed | NS |  |  |  |  |  | low counts |  |
| Alone | low life satisfaction | Stable unemployed | BCS70 | 1.45 | 1.01 | 2.07 | 22.54 |  |  |  |
| Alone | low life satisfaction | Stable unemployed | NCDS | 2.36 | 1.66 | 3.36 | 22.75 |  |  |  |
| Alone | low life satisfaction | Stable unemployed | USOC | 1.14 | 0.75 | 1.72 | 20.63 |  |  |  |
| Alone | low life satisfaction | Stable unemployed | ELSA | 1.87 | 1.16 | 3.02 | 18.63 |  |  |  |
| Alone | low life satisfaction | Stable unemployed | GS | 0.88 | 0.49 | 1.59 | 15.46 |  |  |  |
| Alone | low life satisfaction | Stable unemployed | ALSPAC(G0) | |  |  |  |  | no measure |  |
| Alone | low life satisfaction | Stable unemployed | Twins UK |  |  |  |  |  | low counts |  |
| Alone | low life satisfaction | Stable unemployed | Overall | 1.50 | 1.08 | 2.08 |  | 66.56 |  |  |
| Alone | often lonely | Furloughed | MCS |  |  |  |  |  | low counts | REML |
| Alone | often lonely | Furloughed | ALSPAC(G1) | |  |  |  |  | no measure |  |
| Alone | often lonely | Furloughed | NS | 1.28 | 0.54 | 3.03 | 15.80 |  |  |  |
| Alone | often lonely | Furloughed | BCS70 | 0.94 | 0.47 | 1.89 | 18.32 |  |  |  |
| Alone | often lonely | Furloughed | NCDS | 0.55 | 0.27 | 1.10 | 18.37 |  |  |  |
| Alone | often lonely | Furloughed | USOC | 2.86 | 1.76 | 4.65 | 21.91 |  |  |  |
| Alone | often lonely | Furloughed | ELSA | 1.61 | 0.64 | 4.04 | 14.90 |  |  |  |
| Alone | often lonely | Furloughed | GS | 0.95 | 0.27 | 3.36 | 10.69 |  |  |  |
| Alone | often lonely | Furloughed | ALSPAC(G0) | |  |  |  |  | no measure |  |
| Alone | often lonely | Furloughed | Twins UK |  |  |  |  |  |  |  |
| Alone | often lonely | Furloughed | Overall | 1.24 | 0.73 | 2.11 |  | 65.26 |  |  |
| Alone | often lonely | No longer employed | MCS |  |  |  |  |  | low counts | REML |
| Alone | often lonely | No longer employed | ALSPAC(G1) | |  |  |  |  | no measure |  |
| Alone | often lonely | No longer employed | NS |  |  |  |  |  | low counts |  |
| Alone | often lonely | No longer employed | BCS70 |  |  |  |  |  | low counts |  |
| Alone | often lonely | No longer employed | NCDS | 1.64 | 0.55 | 4.89 | 52.83 |  |  |  |
| Alone | often lonely | No longer employed | USOC | 1.53 | 0.48 | 4.88 | 47.17 |  |  |  |
| Alone | often lonely | No longer employed | ELSA |  |  |  |  |  | low counts |  |
| Alone | often lonely | No longer employed | GS |  |  |  |  |  | low counts |  |
| Alone | often lonely | No longer employed | ALSPAC(G0) | |  |  |  |  | no measure |  |
| Alone | often lonely | No longer employed | Twins UK |  |  |  |  |  |  |  |
| Alone | often lonely | No longer employed | Overall | 1.59 | 0.72 | 3.52 |  | 0.00 |  |  |
| Alone | often lonely | Stable unemployed | MCS |  |  |  |  |  | low counts | REML |
| Alone | often lonely | Stable unemployed | ALSPAC(G1) | |  |  |  |  | no measure |  |
| Alone | often lonely | Stable unemployed | NS |  |  |  |  |  | low counts |  |
| Alone | often lonely | Stable unemployed | BCS70 | 1.94 | 0.75 | 5.02 | 22.25 |  |  |  |
| Alone | often lonely | Stable unemployed | NCDS | 2.09 | 1.10 | 3.95 | 49.44 |  |  |  |
| Alone | often lonely | Stable unemployed | USOC |  |  |  |  |  | low counts |  |
| Alone | often lonely | Stable unemployed | ELSA | 2.18 | 0.75 | 6.30 | 17.82 |  |  |  |
| Alone | often lonely | Stable unemployed | GS | 2.11 | 0.53 | 8.41 | 10.49 |  |  |  |
| Alone | often lonely | Stable unemployed | ALSPAC(G0) | |  |  |  |  | no measure |  |
| Alone | often lonely | Stable unemployed | Twins UK |  |  |  |  |  | low counts |  |
| Alone | often lonely | Stable unemployed | Overall | 2.07 | 1.32 | 3.25 |  | 0.00 |  |  |
| Alone | high loneliness | Furloughed | MCS |  |  |  |  |  | low counts | REML |
| Alone | high loneliness | Furloughed | ALSPAC(G1) | |  |  |  |  | no measure |  |
| Alone | high loneliness | Furloughed | NS | 1.81 | 1.15 | 2.85 | 16.69 |  |  |  |
| Alone | high loneliness | Furloughed | BCS70 | 1.08 | 0.75 | 1.55 | 26.21 |  |  |  |
| Alone | high loneliness | Furloughed | NCDS | 1.16 | 0.79 | 1.69 | 23.93 |  |  |  |
| Alone | high loneliness | Furloughed | USOC |  |  |  |  |  | no measure |  |
| Alone | high loneliness | Furloughed | ELSA | 1.09 | 0.72 | 1.63 | 20.77 |  |  |  |
| Alone | high loneliness | Furloughed | GS |  |  |  |  |  | no measure |  |
| Alone | high loneliness | Furloughed | ALSPAC(G0) | |  |  |  |  | no measure |  |
| Alone | high loneliness | Furloughed | Twins UK | 1.14 | 0.67 | 1.92 | 12.40 |  |  |  |
| Alone | high loneliness | Furloughed | Overall | 1.21 | 1.00 | 1.45 |  | 0.00 |  |  |
| Alone | high loneliness | No longer employed | MCS |  |  |  |  |  | low counts | REML |
| Alone | high loneliness | No longer employed | ALSPAC(G1) | |  |  |  |  | no measure |  |
| Alone | high loneliness | No longer employed | NS | 1.52 | 0.71 | 3.24 | 20.85 |  |  |  |
| Alone | high loneliness | No longer employed | BCS70 | 1.62 | 0.97 | 2.71 | 45.37 |  |  |  |
| Alone | high loneliness | No longer employed | NCDS | 1.30 | 0.65 | 2.58 | 25.25 |  |  |  |
| Alone | high loneliness | No longer employed | USOC |  |  |  |  |  | no measure |  |
| Alone | high loneliness | No longer employed | ELSA | 1.29 | 0.40 | 4.23 | 8.52 |  |  |  |
| Alone | high loneliness | No longer employed | GS |  |  |  |  |  | no measure |  |
| Alone | high loneliness | No longer employed | ALSPAC(G0) | |  |  |  |  |  |  |
| Alone | high loneliness | No longer employed | Twins UK |  |  |  |  |  | low counts |  |
| Alone | high loneliness | No longer employed | Overall | 1.48 | 1.05 | 2.10 |  | 0.00 |  |  |
| Alone | high loneliness | Stable unemployed | MCS |  |  |  |  |  | low counts | REML |
| Alone | high loneliness | Stable unemployed | ALSPAC(G1) | |  |  |  |  | no measure |  |
| Alone | high loneliness | Stable unemployed | NS |  |  |  |  |  | low counts |  |
| Alone | high loneliness | Stable unemployed | BCS70 | 1.64 | 1.10 | 2.44 | 41.52 |  |  |  |
| Alone | high loneliness | Stable unemployed | NCDS | 1.42 | 0.93 | 2.19 | 35.88 |  |  |  |
| Alone | high loneliness | Stable unemployed | USOC |  |  |  |  |  | no measure |  |
| Alone | high loneliness | Stable unemployed | ELSA | 1.46 | 0.85 | 2.51 | 22.59 |  |  |  |
| Alone | high loneliness | Stable unemployed | GS |  |  |  |  |  | no measure |  |
| Alone | high loneliness | Stable unemployed | ALSPAC(G0) | |  |  |  |  | no measure |  |
| Alone | high loneliness | Stable unemployed | Twins UK |  |  |  |  |  | low counts |  |
| Alone | high loneliness | Stable unemployed | Overall | 1.52 | 1.17 | 1.96 |  | 0.00 |  |  |
| Alone | distressed (bin.) | Furloughed | MCS |  |  |  |  |  | low counts | REML |
| Alone | distressed (bin.) | Furloughed | ALSPAC(G1) | 2.07 | 0.69 | 6.21 | 4.47 |  |  |  |
| Alone | distressed (bin.) | Furloughed | NS |  |  |  |  |  | low counts |  |
| Alone | distressed (bin.) | Furloughed | BCS70 | 1.06 | 0.60 | 1.86 | 16.64 |  |  |  |
| Alone | distressed (bin.) | Furloughed | NCDS | 1.05 | 0.58 | 1.91 | 14.90 |  |  |  |
| Alone | distressed (bin.) | Furloughed | USOC | 1.67 | 1.18 | 2.35 | 43.53 |  |  |  |
| Alone | distressed (bin.) | Furloughed | ELSA | 1.30 | 0.74 | 2.28 | 16.92 |  |  |  |
| Alone | distressed (bin.) | Furloughed | GS |  |  |  |  |  | low counts |  |
| Alone | distressed (bin.) | Furloughed | ALSPAC(G0) | 1.81 | 0.53 | 6.22 | 3.55 |  |  |  |
| Alone | distressed (bin.) | Furloughed | Twins UK |  |  |  |  |  | low counts |  |
| Alone | distressed (bin.) | Furloughed | Overall | 1.40 | 1.11 | 1.77 |  | 1.84 |  |  |
| Alone | distressed (bin.) | No longer employed | MCS |  |  |  |  |  | low counts | REML |
| Alone | distressed (bin.) | No longer employed | ALSPAC(G1) | 2.33 | 1.03 | 5.28 | 21.59 |  |  |  |
| Alone | distressed (bin.) | No longer employed | NS |  |  |  |  |  | low counts |  |
| Alone | distressed (bin.) | No longer employed | BCS70 |  |  |  |  |  | low counts |  |
| Alone | distressed (bin.) | No longer employed | NCDS | 1.24 | 0.41 | 3.70 | 13.94 |  |  |  |
| Alone | distressed (bin.) | No longer employed | USOC | 1.63 | 1.01 | 2.65 | 39.17 |  |  |  |
| Alone | distressed (bin.) | No longer employed | ELSA | 3.84 | 1.86 | 7.95 | 25.31 |  |  |  |
| Alone | distressed (bin.) | No longer employed | GS |  |  |  |  |  | low counts |  |
| Alone | distressed (bin.) | No longer employed | ALSPAC(G0) | |  |  |  |  | low counts |  |
| Alone | distressed (bin.) | No longer employed | Twins UK |  |  |  |  |  | low counts |  |
| Alone | distressed (bin.) | No longer employed | Overall | 2.11 | 1.33 | 3.33 |  | 35.86 |  |  |
| Alone | distressed (bin.) | Stable unemployed | MCS |  |  |  |  |  | low counts | REML |
| Alone | distressed (bin.) | Stable unemployed | ALSPAC(G1) | |  |  |  |  | low counts |  |
| Alone | distressed (bin.) | Stable unemployed | NS |  |  |  |  |  | low counts |  |
| Alone | distressed (bin.) | Stable unemployed | BCS70 | 3.06 | 1.22 | 7.68 | 20.79 |  |  |  |
| Alone | distressed (bin.) | Stable unemployed | NCDS | 1.43 | 0.47 | 4.36 | 15.46 |  |  |  |
| Alone | distressed (bin.) | Stable unemployed | USOC | 0.97 | 0.39 | 2.41 | 21.29 |  |  |  |
| Alone | distressed (bin.) | Stable unemployed | ELSA | 2.49 | 1.49 | 4.17 | 42.45 |  |  |  |
| Alone | distressed (bin.) | Stable unemployed | GS |  |  |  |  |  | low counts |  |
| Alone | distressed (bin.) | Stable unemployed | ALSPAC(G0) | |  |  |  |  | low counts |  |
| Alone | distressed (bin.) | Stable unemployed | Twins UK |  |  |  |  |  | low counts |  |
| Alone | distressed (bin.) | Stable unemployed | Overall | 1.95 | 1.20 | 3.18 |  | 30.07 |  |  |
| Alone | distressed (cont.) | Furloughed | MCS | -1.90 | -3.47 | -0.33 | 0.43 |  |  | REML |
| Alone | distressed (cont.) | Furloughed | ALSPAC(G1) | 0.09 | -0.70 | 0.89 | 1.66 |  |  |  |
| Alone | distressed (cont.) | Furloughed | NS | 0.50 | 0.12 | 0.88 | 7.35 |  |  |  |
| Alone | distressed (cont.) | Furloughed | BCS70 | 0.11 | -0.15 | 0.36 | 15.88 |  |  |  |
| Alone | distressed (cont.) | Furloughed | NCDS | 0.09 | -0.12 | 0.29 | 25.74 |  |  |  |
| Alone | distressed (cont.) | Furloughed | USOC | 0.17 | -0.02 | 0.36 | 29.39 |  |  |  |
| Alone | distressed (cont.) | Furloughed | ELSA | 0.12 | -0.12 | 0.37 | 17.43 |  |  |  |
| Alone | distressed (cont.) | Furloughed | GS | -0.89 | -2.77 | 0.99 | 0.30 |  |  |  |
| Alone | distressed (cont.) | Furloughed | ALSPAC(G0) | 1.42 | 0.66 | 2.18 | 1.82 |  |  |  |
| Alone | distressed (cont.) | Furloughed | Twins UK |  |  |  |  |  | low counts |  |
| Alone | distressed (cont.) | Furloughed | Overall | 0.16 | 0.06 | 0.27 |  | 0.00 |  |  |
| Alone | distressed (cont.) | No longer employed | MCS | -2.58 | -5.06 | -0.10 | 2.81 |  |  | REML |
| Alone | distressed (cont.) | No longer employed | ALSPAC(G1) | -0.03 | -0.70 | 0.64 | 14.67 |  |  |  |
| Alone | distressed (cont.) | No longer employed | NS | 1.20 | 0.17 | 2.23 | 10.13 |  |  |  |
| Alone | distressed (cont.) | No longer employed | BCS70 | 0.78 | -0.32 | 1.88 | 9.35 |  |  |  |
| Alone | distressed (cont.) | No longer employed | NCDS | 0.13 | -0.23 | 0.49 | 19.23 |  |  |  |
| Alone | distressed (cont.) | No longer employed | USOC | 0.49 | -0.09 | 1.08 | 15.93 |  |  |  |
| Alone | distressed (cont.) | No longer employed | ELSA | 0.97 | 0.30 | 1.64 | 14.68 |  |  |  |
| Alone | distressed (cont.) | No longer employed | GS | 1.13 | -2.74 | 5.00 | 1.25 |  |  |  |
| Alone | distressed (cont.) | No longer employed | ALSPAC(G0) | 1.55 | 0.68 | 2.42 | 11.94 |  |  |  |
| Alone | distressed (cont.) | No longer employed | Twins UK |  |  |  |  |  | low counts |  |
| Alone | distressed (cont.) | No longer employed | Overall | 0.57 | 0.12 | 1.01 |  | 61.16 |  |  |
| Alone | distressed (cont.) | Stable unemployed | MCS |  |  |  |  |  | low counts | REML |
| Alone | distressed (cont.) | Stable unemployed | ALSPAC(G1) | |  |  |  |  | low counts |  |
| Alone | distressed (cont.) | Stable unemployed | NS | -1.99 | -3.98 | 0.00 | 6.13 |  |  |  |
| Alone | distressed (cont.) | Stable unemployed | BCS70 | 1.45 | 0.52 | 2.37 | 14.81 |  |  |  |
| Alone | distressed (cont.) | Stable unemployed | NCDS | 0.41 | 0.01 | 0.81 | 21.72 |  |  |  |
| Alone | distressed (cont.) | Stable unemployed | USOC | 0.11 | -0.26 | 0.47 | 22.11 |  |  |  |
| Alone | distressed (cont.) | Stable unemployed | ELSA | 0.63 | -0.01 | 1.28 | 18.59 |  |  |  |
| Alone | distressed (cont.) | Stable unemployed | GS | 2.62 | -3.87 | 9.11 | 0.75 |  |  |  |
| Alone | distressed (cont.) | Stable unemployed | ALSPAC(G0) | 1.35 | 0.51 | 2.19 | 15.91 |  |  |  |
| Alone | distressed (cont.) | Stable unemployed | Twins UK |  |  |  |  |  | low counts |  |
| Alone | distressed (cont.) | Stable unemployed | Overall | 0.56 | -0.01 | 1.13 |  | 75.21 |  |  |
| Alone | less than daily contact | Furloughed | MCS |  |  |  |  |  | low counts | REML |
| Alone | less than daily contact | Furloughed | ALSPAC(G1) | 2.61 | 0.63 | 10.89 | 1.00 |  |  |  |
| Alone | less than daily contact | Furloughed | NS | 0.79 | 0.51 | 1.20 | 8.91 |  |  |  |
| Alone | less than daily contact | Furloughed | BCS70 | 0.67 | 0.49 | 0.90 | 14.67 |  |  |  |
| Alone | less than daily contact | Furloughed | NCDS | 0.88 | 0.68 | 1.14 | 17.54 |  |  |  |
| Alone | less than daily contact | Furloughed | USOC | 0.96 | 0.83 | 1.11 | 28.53 |  |  |  |
| Alone | less than daily contact | Furloughed | ELSA | 1.17 | 0.90 | 1.53 | 17.01 |  |  |  |
| Alone | less than daily contact | Furloughed | GS | 1.01 | 0.66 | 1.57 | 8.67 |  |  |  |
| Alone | less than daily contact | Furloughed | ALSPAC(G0) | |  |  |  |  | low counts |  |
| Alone | less than daily contact | Furloughed | Twins UK | 1.09 | 0.53 | 2.23 | 3.67 |  |  |  |
| Alone | less than daily contact | Furloughed | Overall | 0.93 | 0.80 | 1.07 |  | 35.04 |  |  |
| Alone | less than daily contact | No longer employed | MCS |  |  |  |  |  | low counts | REML |
| Alone | less than daily contact | No longer employed | ALSPAC(G1) | |  |  |  |  | low counts |  |
| Alone | less than daily contact | No longer employed | NS | 1.13 | 0.61 | 2.08 | 10.27 |  |  |  |
| Alone | less than daily contact | No longer employed | BCS70 |  |  |  |  |  | low counts |  |
| Alone | less than daily contact | No longer employed | NCDS | 1.21 | 0.92 | 1.59 | 35.78 |  |  |  |
| Alone | less than daily contact | No longer employed | USOC | 0.90 | 0.71 | 1.15 | 41.48 |  |  |  |
| Alone | less than daily contact | No longer employed | ELSA |  |  |  |  |  | low counts |  |
| Alone | less than daily contact | No longer employed | GS | 1.33 | 0.77 | 2.31 | 12.48 |  |  |  |
| Alone | less than daily contact | No longer employed | ALSPAC(G0) | |  |  |  |  | low counts |  |
| Alone | less than daily contact | No longer employed | Twins UK |  |  |  |  |  | low counts |  |
| Alone | less than daily contact | No longer employed | Overall | 1.08 | 0.87 | 1.33 |  | 27.54 |  |  |
| Alone | less than daily contact | Stable unemployed | MCS |  |  |  |  |  | low counts | REML |
| Alone | less than daily contact | Stable unemployed | ALSPAC(G1) | |  |  |  |  | low counts |  |
| Alone | less than daily contact | Stable unemployed | NS |  |  |  |  |  | low counts |  |
| Alone | less than daily contact | Stable unemployed | BCS70 | 1.22 | 0.58 | 2.56 | 6.13 |  |  |  |
| Alone | less than daily contact | Stable unemployed | NCDS | 1.26 | 0.89 | 1.76 | 29.05 |  |  |  |
| Alone | less than daily contact | Stable unemployed | USOC | 0.98 | 0.74 | 1.30 | 41.74 |  |  |  |
| Alone | less than daily contact | Stable unemployed | ELSA | 1.02 | 0.67 | 1.55 | 18.76 |  |  |  |
| Alone | less than daily contact | Stable unemployed | GS | 1.06 | 0.44 | 2.56 | 4.32 |  |  |  |
| Alone | less than daily contact | Stable unemployed | ALSPAC(G0) | |  |  |  |  | low counts |  |
| Alone | less than daily contact | Stable unemployed | Twins UK |  |  |  |  |  | low counts |  |
| Alone | less than daily contact | Stable unemployed | Overall | 1.08 | 0.90 | 1.30 |  | 0.00 |  |  |
| Alone | fair or poor self-rated health | Furloughed | MCS |  |  |  |  |  | low counts | MLE |
| Alone | fair or poor self-rated health | Furloughed | ALSPAC(G1) | |  |  |  |  | no measure |  |
| Alone | fair or poor self-rated health | Furloughed | NS |  |  |  |  |  | low counts |  |
| Alone | fair or poor self-rated health | Furloughed | BCS70 | 0.72 | 0.32 | 1.65 | 1.71 |  |  |  |
| Alone | fair or poor self-rated health | Furloughed | NCDS | 2.24 | 1.10 | 4.55 | 2.31 |  |  |  |
| Alone | fair or poor self-rated health | Furloughed | USOC |  |  |  |  |  | no measure |  |
| Alone | fair or poor self-rated health | Furloughed | ELSA | 1.01 | 0.91 | 1.13 | 95.98 |  |  |  |
| Alone | fair or poor self-rated health | Furloughed | GS |  |  |  |  |  | low counts |  |
| Alone | fair or poor self-rated health | Furloughed | ALSPAC(G0) | |  |  |  |  | no measure |  |
| Alone | fair or poor self-rated health | Furloughed | Twins UK |  |  |  |  |  | low counts |  |
| Alone | fair or poor self-rated health | Furloughed | Overall | 1.03 | 0.92 | 1.14 |  | 0.00 |  |  |
| Alone | fair or poor self-rated health | No longer employed | MCS |  |  |  |  |  | low counts | REML |
| Alone | fair or poor self-rated health | No longer employed | ALSPAC(G1) | |  |  |  |  | no measure |  |
| Alone | fair or poor self-rated health | No longer employed | NS |  |  |  |  |  | low counts |  |
| Alone | fair or poor self-rated health | No longer employed | BCS70 |  |  |  |  |  | low counts |  |
| Alone | fair or poor self-rated health | No longer employed | NCDS | 1.44 | 0.51 | 4.01 | 100.00 |  |  |  |
| Alone | fair or poor self-rated health | No longer employed | USOC |  |  |  |  |  | no measure |  |
| Alone | fair or poor self-rated health | No longer employed | ELSA |  |  |  |  |  | low counts |  |
| Alone | fair or poor self-rated health | No longer employed | GS |  |  |  |  |  | low counts |  |
| Alone | fair or poor self-rated health | No longer employed | ALSPAC(G0) | |  |  |  |  | no measure |  |
| Alone | fair or poor self-rated health | No longer employed | Twins UK |  |  |  |  |  | low counts |  |
| Alone | fair or poor self-rated health | No longer employed | Overall | 1.44 | 0.51 | 4.01 |  |  |  |  |
| Alone | fair or poor self-rated health | Stable unemployed | MCS |  |  |  |  |  | low counts | REML |
| Alone | fair or poor self-rated health | Stable unemployed | ALSPAC(G1) | |  |  |  |  | no measure |  |
| Alone | fair or poor self-rated health | Stable unemployed | NS |  |  |  |  |  | low counts |  |
| Alone | fair or poor self-rated health | Stable unemployed | BCS70 |  |  |  |  |  | low counts |  |
| Alone | fair or poor self-rated health | Stable unemployed | NCDS | 2.59 | 1.43 | 4.70 | 44.45 |  |  |  |
| Alone | fair or poor self-rated health | Stable unemployed | USOC |  |  |  |  |  | no measure |  |
| Alone | fair or poor self-rated health | Stable unemployed | ELSA | 1.06 | 0.93 | 1.22 | 55.55 |  |  |  |
| Alone | fair or poor self-rated health | Stable unemployed | GS |  |  |  |  |  | low counts |  |
| Alone | fair or poor self-rated health | Stable unemployed | ALSPAC(G0) | |  |  |  |  | no measure |  |
| Alone | fair or poor self-rated health | Stable unemployed | Twins UK |  |  |  |  |  | low counts |  |
| Alone | fair or poor self-rated health | Stable unemployed | Overall | 1.58 | 0.66 | 3.76 |  | 87.70 |  |  |

### 13. Partner

| **Adjustment** | **Outcome** | **Exposure** | **Study** | **Coefficient** | **lower_ci** | **upper_ci** | **%Weight** | **%I2** | **Reason for missing** | **Method** |
| --- | --- | --- | --- | --- | --- | --- | --- | --- | --- | --- |
| Partner | low life satisfaction | Furloughed | MCS | 2.80 | 1.13 | 6.91 | 1.74 |  |  | REML |
| Partner | low life satisfaction | Furloughed | ALSPAC(G1) | |  |  |  |  | no measure |  |
| Partner | low life satisfaction | Furloughed | NS | 1.02 | 0.67 | 1.57 | 6.54 |  |  |  |
| Partner | low life satisfaction | Furloughed | BCS70 | 1.38 | 1.14 | 1.67 | 17.45 |  |  |  |
| Partner | low life satisfaction | Furloughed | NCDS | 1.03 | 0.79 | 1.35 | 12.65 |  |  |  |
| Partner | low life satisfaction | Furloughed | USOC | 0.99 | 0.85 | 1.16 | 20.91 |  |  |  |
| Partner | low life satisfaction | Furloughed | ELSA | 1.02 | 0.79 | 1.32 | 13.16 |  |  |  |
| Partner | low life satisfaction | Furloughed | GS | 1.13 | 0.95 | 1.34 | 19.12 |  |  |  |
| Partner | low life satisfaction | Furloughed | ALSPAC(G0) | |  |  |  |  | no measure |  |
| Partner | low life satisfaction | Furloughed | Twins UK | 1.55 | 1.08 | 2.22 | 8.42 |  |  |  |
| Partner | low life satisfaction | Furloughed | Overall | 1.15 | 1.02 | 1.30 |  | 45.31 |  |  |
| Partner | low life satisfaction | No longer employed | MCS |  |  |  |  |  | low counts | REML |
| Partner | low life satisfaction | No longer employed | ALSPAC(G1) | |  |  |  |  | no measure |  |
| Partner | low life satisfaction | No longer employed | NS | 1.31 | 0.67 | 2.58 | 4.36 |  |  |  |
| Partner | low life satisfaction | No longer employed | BCS70 | 0.49 | 0.21 | 1.14 | 2.78 |  |  |  |
| Partner | low life satisfaction | No longer employed | NCDS | 1.73 | 1.25 | 2.38 | 18.59 |  |  |  |
| Partner | low life satisfaction | No longer employed | USOC | 1.34 | 1.04 | 1.72 | 29.47 |  |  |  |
| Partner | low life satisfaction | No longer employed | ELSA | 1.48 | 1.04 | 2.13 | 14.94 |  |  |  |
| Partner | low life satisfaction | No longer employed | GS | 1.22 | 0.95 | 1.56 | 29.85 |  |  |  |
| Partner | low life satisfaction | No longer employed | ALSPAC(G0) | |  |  |  |  | no measure |  |
| Partner | low life satisfaction | No longer employed | Twins UK |  |  |  |  |  | low counts |  |
| Partner | low life satisfaction | No longer employed | Overall | 1.35 | 1.17 | 1.55 |  | 5.29 |  |  |
| Partner | low life satisfaction | Stable unemployed | MCS |  |  |  |  |  | low counts | REML |
| Partner | low life satisfaction | Stable unemployed | ALSPAC(G1) | |  |  |  |  | no measure |  |
| Partner | low life satisfaction | Stable unemployed | NS | 2.79 | 1.59 | 4.93 | 15.73 |  |  |  |
| Partner | low life satisfaction | Stable unemployed | BCS70 |  |  |  |  |  | low counts |  |
| Partner | low life satisfaction | Stable unemployed | NCDS | 2.71 | 1.69 | 4.36 | 20.33 |  |  |  |
| Partner | low life satisfaction | Stable unemployed | USOC | 1.78 | 1.41 | 2.24 | 42.61 |  |  |  |
| Partner | low life satisfaction | Stable unemployed | ELSA | 1.56 | 0.99 | 2.47 | 21.33 |  |  |  |
| Partner | low life satisfaction | Stable unemployed | GS |  |  |  |  |  | low counts |  |
| Partner | low life satisfaction | Stable unemployed | ALSPAC(G0) | |  |  |  |  | no measure |  |
| Partner | low life satisfaction | Stable unemployed | Twins UK |  |  |  |  |  | low counts |  |
| Partner | low life satisfaction | Stable unemployed | Overall | 2.02 | 1.56 | 2.62 |  | 37.78 |  |  |
| Partner | often lonely | Furloughed | MCS | 0.16 | 0.01 | 2.15 | 2.02 |  |  | REML |
| Partner | often lonely | Furloughed | ALSPAC(G1) | |  |  |  |  | no measure |  |
| Partner | often lonely | Furloughed | NS | 2.67 | 1.04 | 6.84 | 12.95 |  |  |  |
| Partner | often lonely | Furloughed | BCS70 | 1.60 | 0.84 | 3.08 | 22.74 |  |  |  |
| Partner | often lonely | Furloughed | NCDS | 0.75 | 0.37 | 1.53 | 20.14 |  |  |  |
| Partner | often lonely | Furloughed | USOC | 1.25 | 0.73 | 2.15 | 28.97 |  |  |  |
| Partner | often lonely | Furloughed | ELSA | 0.82 | 0.32 | 2.07 | 13.19 |  |  |  |
| Partner | often lonely | Furloughed | GS |  |  |  |  |  |  |  |
| Partner | often lonely | Furloughed | ALSPAC(G0) | |  |  |  |  | no measure |  |
| Partner | often lonely | Furloughed | Twins UK |  |  |  |  |  | low counts |  |
| Partner | often lonely | Furloughed | Overall | 1.20 | 0.83 | 1.73 |  | 22.01 |  |  |
| Partner | often lonely | No longer employed | MCS |  |  |  |  |  | low counts | REML |
| Partner | often lonely | No longer employed | ALSPAC(G1) | |  |  |  |  | no measure |  |
| Partner | often lonely | No longer employed | NS | 1.16 | 0.30 | 4.47 | 13.25 |  |  |  |
| Partner | often lonely | No longer employed | BCS70 | 2.56 | 0.75 | 8.80 | 15.82 |  |  |  |
| Partner | often lonely | No longer employed | NCDS | 3.14 | 1.11 | 8.89 | 22.18 |  |  |  |
| Partner | often lonely | No longer employed | USOC | 1.76 | 0.67 | 4.66 | 25.54 |  |  |  |
| Partner | often lonely | No longer employed | ELSA | 1.39 | 0.33 | 5.87 | 11.63 |  |  |  |
| Partner | often lonely | No longer employed | GS | 4.06 | 0.96 | 17.17 | 11.58 |  |  |  |
| Partner | often lonely | No longer employed | ALSPAC(G0) | |  |  |  |  | no measure |  |
| Partner | often lonely | No longer employed | Twins UK |  |  |  |  |  | low counts |  |
| Partner | often lonely | No longer employed | Overall | 2.16 | 1.32 | 3.52 |  | 0.00 |  |  |
| Partner | often lonely | Stable unemployed | MCS |  |  |  |  |  | low counts | REML |
| Partner | often lonely | Stable unemployed | ALSPAC(G1) | |  |  |  |  | no measure |  |
| Partner | often lonely | Stable unemployed | NS | 7.93 | 2.12 | 29.70 | 18.30 |  |  |  |
| Partner | often lonely | Stable unemployed | BCS70 |  |  |  |  |  | low counts |  |
| Partner | often lonely | Stable unemployed | NCDS | 5.43 | 2.21 | 13.30 | 38.31 |  |  |  |
| Partner | often lonely | Stable unemployed | USOC | 2.55 | 0.99 | 6.56 | 34.71 |  |  |  |
| Partner | often lonely | Stable unemployed | ELSA | 1.68 | 0.24 | 11.64 | 8.68 |  |  |  |
| Partner | often lonely | Stable unemployed | GS |  |  |  |  |  | low counts |  |
| Partner | often lonely | Stable unemployed | ALSPAC(G0) | |  |  |  |  | no measure |  |
| Partner | often lonely | Stable unemployed | Twins UK |  |  |  |  |  | low counts |  |
| Partner | often lonely | Stable unemployed | Overall | 4.04 | 2.28 | 7.18 |  | 3.89 |  |  |
| Partner | high loneliness | Furloughed | MCS | 0.98 | 0.31 | 3.15 | 3.00 |  |  | REML |
| Partner | high loneliness | Furloughed | ALSPAC(G1) | |  |  |  |  | no measure |  |
| Partner | high loneliness | Furloughed | NS | 1.24 | 0.80 | 1.90 | 15.40 |  |  |  |
| Partner | high loneliness | Furloughed | BCS70 | 1.37 | 1.05 | 1.79 | 25.91 |  |  |  |
| Partner | high loneliness | Furloughed | NCDS | 0.87 | 0.63 | 1.18 | 22.14 |  |  |  |
| Partner | high loneliness | Furloughed | USOC |  |  |  |  |  | no measure |  |
| Partner | high loneliness | Furloughed | ELSA | 0.94 | 0.64 | 1.40 | 17.22 |  |  |  |
| Partner | high loneliness | Furloughed | GS |  |  |  |  |  | no measure |  |
| Partner | high loneliness | Furloughed | ALSPAC(G0) | |  |  |  |  | no measure |  |
| Partner | high loneliness | Furloughed | Twins UK | 0.83 | 0.55 | 1.25 | 16.33 |  |  |  |
| Partner | high loneliness | Furloughed | Overall | 1.04 | 0.85 | 1.28 |  | 39.80 |  |  |
| Partner | high loneliness | No longer employed | MCS |  |  |  |  |  | low counts | REML |
| Partner | high loneliness | No longer employed | ALSPAC(G1) | |  |  |  |  | no measure |  |
| Partner | high loneliness | No longer employed | NS | 0.96 | 0.46 | 2.02 | 14.18 |  |  |  |
| Partner | high loneliness | No longer employed | BCS70 | 0.79 | 0.29 | 2.14 | 7.78 |  |  |  |
| Partner | high loneliness | No longer employed | NCDS | 1.31 | 0.77 | 2.23 | 27.45 |  |  |  |
| Partner | high loneliness | No longer employed | USOC |  |  |  |  |  | no measure |  |
| Partner | high loneliness | No longer employed | ELSA | 0.97 | 0.61 | 1.55 | 35.85 |  |  |  |
| Partner | high loneliness | No longer employed | GS |  |  |  |  |  | no measure |  |
| Partner | high loneliness | No longer employed | ALSPAC(G0) | |  |  |  |  | no measure |  |
| Partner | high loneliness | No longer employed | Twins UK | 0.74 | 0.36 | 1.54 | 14.73 |  |  |  |
| Partner | high loneliness | No longer employed | Overall | 1.00 | 0.75 | 1.32 |  | 0.00 |  |  |
| Partner | high loneliness | Stable unemployed | MCS |  |  |  |  |  | low counts | REML |
| Partner | high loneliness | Stable unemployed | ALSPAC(G1) | |  |  |  |  | no measure |  |
| Partner | high loneliness | Stable unemployed | NS |  |  |  |  |  | low counts |  |
| Partner | high loneliness | Stable unemployed | BCS70 |  |  |  |  |  | low counts |  |
| Partner | high loneliness | Stable unemployed | NCDS | 1.76 | 0.91 | 3.40 | 58.65 |  |  |  |
| Partner | high loneliness | Stable unemployed | USOC |  |  |  |  |  | no measure |  |
| Partner | high loneliness | Stable unemployed | ELSA |  |  |  |  |  | low counts |  |
| Partner | high loneliness | Stable unemployed | GS |  |  |  |  |  | no measure |  |
| Partner | high loneliness | Stable unemployed | ALSPAC(G0) | |  |  |  |  | no measure |  |
| Partner | high loneliness | Stable unemployed | Twins UK | 1.18 | 0.54 | 2.59 | 41.35 |  |  |  |
| Partner | high loneliness | Stable unemployed | Overall | 1.49 | 0.90 | 2.47 |  | 0.00 |  |  |
| Partner | distressed (bin.) | Furloughed | MCS | 1.14 | 0.21 | 6.25 | 1.16 |  |  | REML |
| Partner | distressed (bin.) | Furloughed | ALSPAC(G1) | 1.65 | 1.03 | 2.64 | 9.73 |  |  |  |
| Partner | distressed (bin.) | Furloughed | NS | 1.41 | 0.96 | 2.06 | 12.36 |  |  |  |
| Partner | distressed (bin.) | Furloughed | BCS70 | 1.26 | 0.97 | 1.63 | 17.00 |  |  |  |
| Partner | distressed (bin.) | Furloughed | NCDS | 0.72 | 0.49 | 1.04 | 12.58 |  |  |  |
| Partner | distressed (bin.) | Furloughed | USOC | 0.89 | 0.74 | 1.06 | 20.53 |  |  |  |
| Partner | distressed (bin.) | Furloughed | ELSA | 1.04 | 0.72 | 1.51 | 12.75 |  |  |  |
| Partner | distressed (bin.) | Furloughed | GS | 1.01 | 0.54 | 1.92 | 6.46 |  |  |  |
| Partner | distressed (bin.) | Furloughed | ALSPAC(G0) | 1.12 | 0.45 | 2.82 | 3.55 |  |  |  |
| Partner | distressed (bin.) | Furloughed | Twins UK | 0.72 | 0.30 | 1.74 | 3.88 |  |  |  |
| Partner | distressed (bin.) | Furloughed | Overall | 1.06 | 0.88 | 1.28 |  | 47.32 |  |  |
| Partner | distressed (bin.) | No longer employed | MCS |  |  |  |  |  | low counts | REML |
| Partner | distressed (bin.) | No longer employed | ALSPAC(G1) | 1.75 | 0.83 | 3.68 | 7.21 |  |  |  |
| Partner | distressed (bin.) | No longer employed | NS | 1.36 | 0.73 | 2.54 | 10.17 |  |  |  |
| Partner | distressed (bin.) | No longer employed | BCS70 | 0.66 | 0.22 | 1.99 | 3.27 |  |  |  |
| Partner | distressed (bin.) | No longer employed | NCDS | 1.57 | 0.76 | 3.28 | 7.38 |  |  |  |
| Partner | distressed (bin.) | No longer employed | USOC | 1.23 | 0.93 | 1.63 | 49.41 |  |  |  |
| Partner | distressed (bin.) | No longer employed | ELSA | 1.12 | 0.68 | 1.84 | 16.32 |  |  |  |
| Partner | distressed (bin.) | No longer employed | GS | 1.53 | 0.57 | 4.10 | 4.10 |  |  |  |
| Partner | distressed (bin.) | No longer employed | ALSPAC(G0) | 3.08 | 0.79 | 12.07 | 2.14 |  |  |  |
| Partner | distressed (bin.) | No longer employed | Twins UK |  |  |  |  |  | low counts |  |
| Partner | distressed (bin.) | No longer employed | Overall | 1.29 | 1.06 | 1.57 |  | 0.00 |  |  |
| Partner | distressed (bin.) | Stable unemployed | MCS |  |  |  |  |  | low counts | REML |
| Partner | distressed (bin.) | Stable unemployed | ALSPAC(G1) | 0.66 | 0.14 | 3.03 | 4.00 |  |  |  |
| Partner | distressed (bin.) | Stable unemployed | NS | 1.58 | 0.77 | 3.25 | 17.85 |  |  |  |
| Partner | distressed (bin.) | Stable unemployed | BCS70 |  |  |  |  |  | low counts |  |
| Partner | distressed (bin.) | Stable unemployed | NCDS | 1.59 | 0.69 | 3.63 | 13.61 |  |  |  |
| Partner | distressed (bin.) | Stable unemployed | USOC | 1.15 | 0.69 | 1.90 | 36.69 |  |  |  |
| Partner | distressed (bin.) | Stable unemployed | ELSA | 1.11 | 0.54 | 2.27 | 18.06 |  |  |  |
| Partner | distressed (bin.) | Stable unemployed | GS |  |  |  |  |  | low counts |  |
| Partner | distressed (bin.) | Stable unemployed | ALSPAC(G0) | 1.42 | 0.54 | 3.77 | 9.79 |  |  |  |
| Partner | distressed (bin.) | Stable unemployed | Twins UK |  |  |  |  |  | low counts |  |
| Partner | distressed (bin.) | Stable unemployed | Overall | 1.26 | 0.93 | 1.71 |  | 0.00 |  |  |
| Partner | distressed (cont.) | Furloughed | MCS | 0.11 | -0.57 | 0.79 | 1.17 |  |  | REML |
| Partner | distressed (cont.) | Furloughed | ALSPAC(G1) | 0.20 | -0.01 | 0.42 | 8.45 |  |  |  |
| Partner | distressed (cont.) | Furloughed | NS | 0.03 | -0.11 | 0.17 | 13.92 |  |  |  |
| Partner | distressed (cont.) | Furloughed | BCS70 | 0.08 | -0.05 | 0.21 | 15.01 |  |  |  |
| Partner | distressed (cont.) | Furloughed | NCDS | -0.11 | -0.22 | -0.01 | 17.26 |  |  |  |
| Partner | distressed (cont.) | Furloughed | USOC | -0.05 | -0.11 | 0.01 | 23.12 |  |  |  |
| Partner | distressed (cont.) | Furloughed | ELSA | 0.07 | -0.07 | 0.22 | 13.89 |  |  |  |
| Partner | distressed (cont.) | Furloughed | GS | -0.16 | -0.83 | 0.51 | 1.19 |  |  |  |
| Partner | distressed (cont.) | Furloughed | ALSPAC(G0) | 0.15 | -0.12 | 0.42 | 5.99 |  |  |  |
| Partner | distressed (cont.) | Furloughed | Twins UK |  |  |  |  |  | low counts |  |
| Partner | distressed (cont.) | Furloughed | Overall | 0.02 | -0.05 | 0.10 |  | 50.19 |  |  |
| Partner | distressed (cont.) | No longer employed | MCS | -0.31 | -1.33 | 0.71 | 2.45 |  |  | REML |
| Partner | distressed (cont.) | No longer employed | ALSPAC(G1) | 0.28 | -0.20 | 0.75 | 8.19 |  |  |  |
| Partner | distressed (cont.) | No longer employed | NS | 0.35 | 0.05 | 0.66 | 13.19 |  |  |  |
| Partner | distressed (cont.) | No longer employed | BCS70 | -0.30 | -0.52 | -0.08 | 16.49 |  |  |  |
| Partner | distressed (cont.) | No longer employed | NCDS | 0.12 | -0.22 | 0.45 | 12.13 |  |  |  |
| Partner | distressed (cont.) | No longer employed | USOC | 0.11 | -0.02 | 0.25 | 20.11 |  |  |  |
| Partner | distressed (cont.) | No longer employed | ELSA | 0.14 | -0.13 | 0.41 | 14.54 |  |  |  |
| Partner | distressed (cont.) | No longer employed | GS | 1.33 | -0.03 | 2.68 | 1.46 |  |  |  |
| Partner | distressed (cont.) | No longer employed | ALSPAC(G0) | 0.19 | -0.17 | 0.54 | 11.43 |  |  |  |
| Partner | distressed (cont.) | No longer employed | Twins UK |  |  |  |  |  | low counts |  |
| Partner | distressed (cont.) | No longer employed | Overall | 0.11 | -0.06 | 0.28 |  | 58.62 |  |  |
| Partner | distressed (cont.) | Stable unemployed | MCS | 0.02 | -1.14 | 1.18 | 1.21 |  |  | REML |
| Partner | distressed (cont.) | Stable unemployed | ALSPAC(G1) | -0.23 | -0.57 | 0.12 | 12.60 |  |  |  |
| Partner | distressed (cont.) | Stable unemployed | NS | 0.43 | -0.10 | 0.96 | 5.73 |  |  |  |
| Partner | distressed (cont.) | Stable unemployed | BCS70 | -0.50 | -1.05 | 0.04 | 5.44 |  |  |  |
| Partner | distressed (cont.) | Stable unemployed | NCDS | 0.07 | -0.25 | 0.40 | 14.33 |  |  |  |
| Partner | distressed (cont.) | Stable unemployed | USOC | 0.14 | -0.08 | 0.36 | 28.17 |  |  |  |
| Partner | distressed (cont.) | Stable unemployed | ELSA | 0.18 | -0.23 | 0.59 | 9.10 |  |  |  |
| Partner | distressed (cont.) | Stable unemployed | GS | -0.69 | -3.35 | 1.98 | 0.23 |  |  |  |
| Partner | distressed (cont.) | Stable unemployed | ALSPAC(G0) | 0.04 | -0.21 | 0.28 | 23.19 |  |  |  |
| Partner | distressed (cont.) | Stable unemployed | Twins UK |  |  |  |  |  | low counts |  |
| Partner | distressed (cont.) | Stable unemployed | Overall | 0.04 | -0.09 | 0.17 |  | 6.87 |  |  |
| Partner | less than daily contact | Furloughed | MCS | 1.22 | 0.71 | 2.11 | 0.59 |  |  | REML |
| Partner | less than daily contact | Furloughed | ALSPAC(G1) | 1.04 | 0.45 | 2.39 | 0.25 |  |  |  |
| Partner | less than daily contact | Furloughed | NS | 0.94 | 0.79 | 1.12 | 5.49 |  |  |  |
| Partner | less than daily contact | Furloughed | BCS70 | 0.91 | 0.84 | 1.00 | 17.64 |  |  |  |
| Partner | less than daily contact | Furloughed | NCDS | 0.94 | 0.85 | 1.05 | 12.52 |  |  |  |
| Partner | less than daily contact | Furloughed | USOC | 1.00 | 0.95 | 1.04 | 45.18 |  |  |  |
| Partner | less than daily contact | Furloughed | ELSA | 1.03 | 0.91 | 1.16 | 10.42 |  |  |  |
| Partner | less than daily contact | Furloughed | GS | 0.92 | 0.79 | 1.07 | 7.05 |  |  |  |
| Partner | less than daily contact | Furloughed | ALSPAC(G0) | 0.91 | 0.58 | 1.41 | 0.87 |  |  |  |
| Partner | less than daily contact | Furloughed | Twins UK |  |  |  |  |  | low counts |  |
| Partner | less than daily contact | Furloughed | Overall | 0.97 | 0.93 | 1.01 |  | 10.22 |  |  |
| Partner | less than daily contact | No longer employed | MCS | 1.24 | 0.42 | 3.67 | 0.38 |  |  | REML |
| Partner | less than daily contact | No longer employed | ALSPAC(G1) | 0.83 | 0.19 | 3.75 | 0.20 |  |  |  |
| Partner | less than daily contact | No longer employed | NS | 1.06 | 0.72 | 1.55 | 3.04 |  |  |  |
| Partner | less than daily contact | No longer employed | BCS70 | 1.07 | 0.90 | 1.29 | 13.56 |  |  |  |
| Partner | less than daily contact | No longer employed | NCDS | 0.98 | 0.79 | 1.22 | 9.64 |  |  |  |
| Partner | less than daily contact | No longer employed | USOC | 1.00 | 0.92 | 1.09 | 57.20 |  |  |  |
| Partner | less than daily contact | No longer employed | ELSA | 0.94 | 0.70 | 1.28 | 4.82 |  |  |  |
| Partner | less than daily contact | No longer employed | GS | 1.07 | 0.87 | 1.32 | 10.04 |  |  |  |
| Partner | less than daily contact | No longer employed | ALSPAC(G0) | 0.91 | 0.48 | 1.71 | 1.12 |  |  |  |
| Partner | less than daily contact | No longer employed | Twins UK |  |  |  |  |  | low counts |  |
| Partner | less than daily contact | No longer employed | Overall | 1.01 | 0.95 | 1.08 |  | 0.00 |  |  |
| Partner | less than daily contact | Stable unemployed | MCS |  |  |  |  |  | low counts | REML |
| Partner | less than daily contact | Stable unemployed | ALSPAC(G1) | |  |  |  |  | low counts |  |
| Partner | less than daily contact | Stable unemployed | NS | 1.04 | 0.74 | 1.48 | 14.90 |  |  |  |
| Partner | less than daily contact | Stable unemployed | BCS70 |  |  |  |  |  | low counts |  |
| Partner | less than daily contact | Stable unemployed | NCDS | 1.18 | 0.96 | 1.43 | 32.53 |  |  |  |
| Partner | less than daily contact | Stable unemployed | USOC | 0.83 | 0.63 | 1.09 | 21.01 |  |  |  |
| Partner | less than daily contact | Stable unemployed | ELSA | 1.05 | 0.75 | 1.47 | 16.05 |  |  |  |
| Partner | less than daily contact | Stable unemployed | GS | 1.30 | 0.80 | 2.12 | 8.35 |  |  |  |
| Partner | less than daily contact | Stable unemployed | ALSPAC(G0) | 0.82 | 0.48 | 1.39 | 7.16 |  |  |  |
| Partner | less than daily contact | Stable unemployed | Twins UK |  |  |  |  |  | low counts |  |
| Partner | less than daily contact | Stable unemployed | Overall | 1.03 | 0.89 | 1.20 |  | 21.86 |  |  |
| Partner | fair or poor self-rated health | Furloughed | MCS | 5.26 | 1.04 | 26.59 | 1.02 |  |  | MLE |
| Partner | fair or poor self-rated health | Furloughed | ALSPAC(G1) | |  |  |  |  | no measure |  |
| Partner | fair or poor self-rated health | Furloughed | NS | 0.90 | 0.46 | 1.78 | 5.35 |  |  |  |
| Partner | fair or poor self-rated health | Furloughed | BCS70 | 1.39 | 1.03 | 1.88 | 19.38 |  |  |  |
| Partner | fair or poor self-rated health | Furloughed | NCDS | 1.26 | 0.87 | 1.83 | 14.51 |  |  |  |
| Partner | fair or poor self-rated health | Furloughed | USOC |  |  |  |  |  | no measure |  |
| Partner | fair or poor self-rated health | Furloughed | ELSA | 1.05 | 1.00 | 1.10 | 53.14 |  |  |  |
| Partner | fair or poor self-rated health | Furloughed | GS | 1.56 | 0.69 | 3.52 | 3.81 |  |  |  |
| Partner | fair or poor self-rated health | Furloughed | ALSPAC(G0) | |  |  |  |  | no measure |  |
| Partner | fair or poor self-rated health | Furloughed | Twins UK | 2.98 | 1.14 | 7.83 | 2.79 |  |  |  |
| Partner | fair or poor self-rated health | Furloughed | Overall | 1.20 | 1.02 | 1.41 |  | 26.94 |  |  |
| Partner | fair or poor self-rated health | No longer employed | MCS |  |  |  |  |  | low counts | REML |
| Partner | fair or poor self-rated health | No longer employed | ALSPAC(G1) | |  |  |  |  | no measure |  |
| Partner | fair or poor self-rated health | No longer employed | NS | 3.96 | 1.59 | 9.86 | 13.86 |  |  |  |
| Partner | fair or poor self-rated health | No longer employed | BCS70 | 1.32 | 0.62 | 2.81 | 17.24 |  |  |  |
| Partner | fair or poor self-rated health | No longer employed | NCDS | 1.37 | 0.66 | 2.83 | 17.86 |  |  |  |
| Partner | fair or poor self-rated health | No longer employed | USOC |  |  |  |  |  | no measure |  |
| Partner | fair or poor self-rated health | No longer employed | ELSA | 1.02 | 0.93 | 1.12 | 35.79 |  |  |  |
| Partner | fair or poor self-rated health | No longer employed | GS | 1.78 | 0.76 | 4.13 | 15.25 |  |  |  |
| Partner | fair or poor self-rated health | No longer employed | ALSPAC(G0) | |  |  |  |  | no measure |  |
| Partner | fair or poor self-rated health | No longer employed | Twins UK |  |  |  |  |  | low counts |  |
| Partner | fair or poor self-rated health | No longer employed | Overall | 1.48 | 0.96 | 2.28 |  | 60.71 |  |  |
| Partner | fair or poor self-rated health | Stable unemployed | MCS |  |  |  |  |  | low counts | REML |
| Partner | fair or poor self-rated health | Stable unemployed | ALSPAC(G1) | |  |  |  |  | no measure |  |
| Partner | fair or poor self-rated health | Stable unemployed | NS |  |  |  |  |  | low counts |  |
| Partner | fair or poor self-rated health | Stable unemployed | BCS70 |  |  |  |  |  | low counts |  |
| Partner | fair or poor self-rated health | Stable unemployed | NCDS | 1.34 | 0.63 | 2.86 | 6.87 |  |  |  |
| Partner | fair or poor self-rated health | Stable unemployed | USOC |  |  |  |  |  | no measure |  |
| Partner | fair or poor self-rated health | Stable unemployed | ELSA | 1.19 | 0.97 | 1.46 | 93.13 |  |  |  |
| Partner | fair or poor self-rated health | Stable unemployed | GS |  |  |  |  |  | low counts |  |
| Partner | fair or poor self-rated health | Stable unemployed | ALSPAC(G0) | |  |  |  |  | no measure |  |
| Partner | fair or poor self-rated health | Stable unemployed | Twins UK |  |  |  |  |  | low counts |  |
| Partner | fair or poor self-rated health | Stable unemployed | Overall | 1.20 | 0.98 | 1.46 |  | 0.00 |  |  |

### 14. Others

| **Adjustment** | **Outcome** | **Exposure** | **Study** | **Coefficient** | **lower_ci** | **upper_ci** | **%Weight** | **%I2** | **Reason for missing** | **Method** |
| --- | --- | --- | --- | --- | --- | --- | --- | --- | --- | --- |
| Others | low life satisfaction | Furloughed | MCS | 1.18 | 0.89 | 1.55 | 19.64 |  |  | REML |
| Others | low life satisfaction | Furloughed | ALSPAC(G1) | |  |  |  |  | no measure |  |
| Others | low life satisfaction | Furloughed | NS | 0.86 | 0.57 | 1.30 | 11.01 |  |  |  |
| Others | low life satisfaction | Furloughed | BCS70 | 1.56 | 1.07 | 2.27 | 12.65 |  |  |  |
| Others | low life satisfaction | Furloughed | NCDS | 1.84 | 1.09 | 3.10 | 7.28 |  |  |  |
| Others | low life satisfaction | Furloughed | USOC | 1.01 | 0.79 | 1.28 | 23.55 |  |  |  |
| Others | low life satisfaction | Furloughed | ELSA | 1.26 | 0.81 | 1.96 | 9.69 |  |  |  |
| Others | low life satisfaction | Furloughed | GS | 1.19 | 0.84 | 1.70 | 13.93 |  |  |  |
| Others | low life satisfaction | Furloughed | ALSPAC(G0) | |  |  |  |  | no measure |  |
| Others | low life satisfaction | Furloughed | Twins UK | 0.64 | 0.24 | 1.72 | 2.25 |  |  |  |
| Others | low life satisfaction | Furloughed | Overall | 1.17 | 1.00 | 1.36 |  | 21.15 |  |  |
| Others | low life satisfaction | No longer employed | MCS | 1.07 | 0.75 | 1.53 | 27.35 |  |  | REML |
| Others | low life satisfaction | No longer employed | ALSPAC(G1) | |  |  |  |  | no measure |  |
| Others | low life satisfaction | No longer employed | NS | 1.13 | 0.59 | 2.17 | 8.09 |  |  |  |
| Others | low life satisfaction | No longer employed | BCS70 | 2.42 | 0.96 | 6.14 | 3.97 |  |  |  |
| Others | low life satisfaction | No longer employed | NCDS | 2.19 | 0.96 | 5.02 | 5.01 |  |  |  |
| Others | low life satisfaction | No longer employed | USOC | 1.37 | 1.02 | 1.85 | 38.66 |  |  |  |
| Others | low life satisfaction | No longer employed | ELSA | 2.04 | 0.85 | 4.93 | 4.43 |  |  |  |
| Others | low life satisfaction | No longer employed | GS | 1.25 | 0.74 | 2.11 | 12.48 |  |  |  |
| Others | low life satisfaction | No longer employed | ALSPAC(G0) | |  |  |  |  | no measure |  |
| Others | low life satisfaction | No longer employed | Twins UK |  |  |  |  |  | low counts |  |
| Others | low life satisfaction | No longer employed | Overall | 1.33 | 1.11 | 1.60 |  | 0.00 |  |  |
| Others | low life satisfaction | Stable unemployed | MCS | 1.25 | 0.97 | 1.60 | 22.70 |  |  | REML |
| Others | low life satisfaction | Stable unemployed | ALSPAC(G1) | |  |  |  |  | no measure |  |
| Others | low life satisfaction | Stable unemployed | NS | 1.29 | 0.88 | 1.90 | 21.24 |  |  |  |
| Others | low life satisfaction | Stable unemployed | BCS70 |  |  |  |  |  | low counts |  |
| Others | low life satisfaction | Stable unemployed | NCDS | 5.08 | 2.96 | 8.71 | 19.19 |  |  |  |
| Others | low life satisfaction | Stable unemployed | USOC | 1.10 | 0.60 | 2.00 | 18.30 |  |  |  |
| Others | low life satisfaction | Stable unemployed | ELSA | 1.73 | 0.97 | 3.10 | 18.58 |  |  |  |
| Others | low life satisfaction | Stable unemployed | GS |  |  |  |  |  | low counts |  |
| Others | low life satisfaction | Stable unemployed | ALSPAC(G0) | |  |  |  |  | no measure |  |
| Others | low life satisfaction | Stable unemployed | Twins UK |  |  |  |  |  | low counts |  |
| Others | low life satisfaction | Stable unemployed | Overall | 1.71 | 1.00 | 2.91 |  | 86.52 |  |  |
| Others | often lonely | Furloughed | MCS | 0.63 | 0.44 | 0.91 | 31.64 |  |  | REML |
| Others | often lonely | Furloughed | ALSPAC(G1) | |  |  |  |  | no measure |  |
| Others | often lonely | Furloughed | NS | 1.08 | 0.57 | 2.05 | 19.15 |  |  |  |
| Others | often lonely | Furloughed | BCS70 | 0.59 | 0.24 | 1.45 | 12.32 |  |  |  |
| Others | often lonely | Furloughed | NCDS | 9.29 | 0.94 | 92.14 | 2.44 |  |  |  |
| Others | often lonely | Furloughed | USOC | 1.17 | 0.75 | 1.83 | 27.44 |  |  |  |
| Others | often lonely | Furloughed | ELSA | 1.41 | 0.39 | 5.09 | 7.02 |  |  |  |
| Others | often lonely | Furloughed | GS |  |  |  |  |  | low counts |  |
| Others | often lonely | Furloughed | ALSPAC(G0) | |  |  |  |  | no measure |  |
| Others | often lonely | Furloughed | Twins UK |  |  |  |  |  | low counts |  |
| Others | often lonely | Furloughed | Overall | 0.93 | 0.64 | 1.34 |  | 40.85 |  |  |
| Others | often lonely | No longer employed | MCS | 0.63 | 0.32 | 1.23 | 48.11 |  |  | REML |
| Others | often lonely | No longer employed | ALSPAC(G1) | |  |  |  |  | no measure |  |
| Others | often lonely | No longer employed | NS |  |  |  |  |  | low counts |  |
| Others | often lonely | No longer employed | BCS70 |  |  |  |  |  | low counts |  |
| Others | often lonely | No longer employed | NCDS |  |  |  |  |  | low counts |  |
| Others | often lonely | No longer employed | USOC | 2.34 | 1.49 | 3.68 | 51.89 |  |  |  |
| Others | often lonely | No longer employed | ELSA |  |  |  |  |  | low counts |  |
| Others | often lonely | No longer employed | GS |  |  |  |  |  | low counts |  |
| Others | often lonely | No longer employed | ALSPAC(G0) | |  |  |  |  | no measure |  |
| Others | often lonely | No longer employed | Twins UK |  |  |  |  |  | low counts |  |
| Others | often lonely | No longer employed | Overall | 1.24 | 0.34 | 4.51 |  | 90.06 |  |  |
| Others | often lonely | Stable unemployed | MCS | 1.12 | 0.69 | 1.83 | 55.55 |  |  | REML |
| Others | often lonely | Stable unemployed | ALSPAC(G1) | |  |  |  |  | no measure |  |
| Others | often lonely | Stable unemployed | NS | 1.12 | 0.58 | 2.18 | 30.17 |  |  |  |
| Others | often lonely | Stable unemployed | BCS70 |  |  |  |  |  | low counts |  |
| Others | often lonely | Stable unemployed | NCDS |  |  |  |  |  | low counts |  |
| Others | often lonely | Stable unemployed | USOC | 0.50 | 0.19 | 1.32 | 14.29 |  |  |  |
| Others | often lonely | Stable unemployed | ELSA |  |  |  |  |  | low counts |  |
| Others | often lonely | Stable unemployed | GS |  |  |  |  |  | low counts |  |
| Others | often lonely | Stable unemployed | ALSPAC(G0) | |  |  |  |  | no measure |  |
| Others | often lonely | Stable unemployed | Twins UK |  |  |  |  |  | low counts |  |
| Others | often lonely | Stable unemployed | Overall | 1.00 | 0.69 | 1.44 |  | 0.00 |  |  |
| Others | high loneliness | Furloughed | MCS | 1.10 | 0.85 | 1.42 | 42.25 |  |  | REML |
| Others | high loneliness | Furloughed | ALSPAC(G1) | |  |  |  |  | no measure |  |
| Others | high loneliness | Furloughed | NS | 0.89 | 0.61 | 1.31 | 19.17 |  |  |  |
| Others | high loneliness | Furloughed | BCS70 | 1.23 | 0.81 | 1.86 | 16.29 |  |  |  |
| Others | high loneliness | Furloughed | NCDS | 1.98 | 1.02 | 3.83 | 6.48 |  |  |  |
| Others | high loneliness | Furloughed | USOC |  |  |  |  |  | no measure |  |
| Others | high loneliness | Furloughed | ELSA | 0.82 | 0.43 | 1.57 | 6.64 |  |  |  |
| Others | high loneliness | Furloughed | GS |  |  |  |  |  | no measure |  |
| Others | high loneliness | Furloughed | ALSPAC(G0) | |  |  |  |  | no measure |  |
| Others | high loneliness | Furloughed | Twins UK | 1.30 | 0.74 | 2.26 | 9.18 |  |  |  |
| Others | high loneliness | Furloughed | Overall | 1.11 | 0.94 | 1.32 |  | 0.00 |  |  |
| Others | high loneliness | No longer employed | MCS | 1.19 | 0.77 | 1.84 | 50.65 |  |  | REML |
| Others | high loneliness | No longer employed | ALSPAC(G1) | |  |  |  |  | no measure |  |
| Others | high loneliness | No longer employed | NS | 1.49 | 0.94 | 2.38 | 44.46 |  |  |  |
| Others | high loneliness | No longer employed | BCS70 |  |  |  |  |  | low counts |  |
| Others | high loneliness | No longer employed | NCDS | 0.91 | 0.22 | 3.73 | 4.90 |  |  |  |
| Others | high loneliness | No longer employed | USOC |  |  |  |  |  | no measure |  |
| Others | high loneliness | No longer employed | ELSA |  |  |  |  |  | low counts |  |
| Others | high loneliness | No longer employed | GS |  |  |  |  |  | no measure |  |
| Others | high loneliness | No longer employed | ALSPAC(G0) | |  |  |  |  | no measure |  |
| Others | high loneliness | No longer employed | Twins UK |  |  |  |  |  | low counts |  |
| Others | high loneliness | No longer employed | Overall | 1.30 | 0.95 | 1.77 |  | 0.00 |  |  |
| Others | high loneliness | Stable unemployed | MCS | 0.88 | 0.53 | 1.44 | 29.53 |  |  | REML |
| Others | high loneliness | Stable unemployed | ALSPAC(G1) | |  |  |  |  | no measure |  |
| Others | high loneliness | Stable unemployed | NS | 1.13 | 0.72 | 1.78 | 31.04 |  |  |  |
| Others | high loneliness | Stable unemployed | BCS70 |  |  |  |  |  | low counts |  |
| Others | high loneliness | Stable unemployed | NCDS | 3.59 | 1.47 | 8.75 | 18.25 |  |  |  |
| Others | high loneliness | Stable unemployed | USOC |  |  |  |  |  | no measure |  |
| Others | high loneliness | Stable unemployed | ELSA | 1.29 | 0.60 | 2.79 | 21.19 |  |  |  |
| Others | high loneliness | Stable unemployed | GS |  |  |  |  |  | no measure |  |
| Others | high loneliness | Stable unemployed | ALSPAC(G0) | |  |  |  |  | no measure |  |
| Others | high loneliness | Stable unemployed | Twins UK |  |  |  |  |  | low counts |  |
| Others | high loneliness | Stable unemployed | Overall | 1.33 | 0.80 | 2.23 |  | 63.84 |  |  |
| Others | distressed (bin.) | Furloughed | MCS | 0.81 | 0.48 | 1.39 | 13.10 |  |  | REML |
| Others | distressed (bin.) | Furloughed | ALSPAC(G1) | 1.52 | 0.91 | 2.56 | 13.65 |  |  |  |
| Others | distressed (bin.) | Furloughed | NS | 0.93 | 0.58 | 1.47 | 15.34 |  |  |  |
| Others | distressed (bin.) | Furloughed | BCS70 | 2.21 | 1.19 | 4.12 | 10.97 |  |  |  |
| Others | distressed (bin.) | Furloughed | NCDS | 1.25 | 0.31 | 4.97 | 3.12 |  |  |  |
| Others | distressed (bin.) | Furloughed | USOC | 0.90 | 0.70 | 1.15 | 23.83 |  |  |  |
| Others | distressed (bin.) | Furloughed | ELSA | 0.81 | 0.40 | 1.63 | 9.38 |  |  |  |
| Others | distressed (bin.) | Furloughed | GS | 1.30 | 0.53 | 3.16 | 6.53 |  |  |  |
| Others | distressed (bin.) | Furloughed | ALSPAC(G0) | 2.39 | 0.73 | 7.81 | 4.08 |  |  |  |
| Others | distressed (bin.) | Furloughed | Twins UK |  |  |  |  |  | low counts |  |
| Others | distressed (bin.) | Furloughed | Overall | 1.13 | 0.87 | 1.46 |  | 41.55 |  |  |
| Others | distressed (bin.) | No longer employed | MCS | 1.00 | 0.47 | 2.15 | 9.66 |  |  | REML |
| Others | distressed (bin.) | No longer employed | ALSPAC(G1) | 1.21 | 0.55 | 2.67 | 8.92 |  |  |  |
| Others | distressed (bin.) | No longer employed | NS | 0.68 | 0.20 | 2.27 | 3.81 |  |  |  |
| Others | distressed (bin.) | No longer employed | BCS70 | 2.64 | 1.21 | 5.75 | 9.22 |  |  |  |
| Others | distressed (bin.) | No longer employed | NCDS |  |  |  |  |  | low counts |  |
| Others | distressed (bin.) | No longer employed | USOC | 1.33 | 0.99 | 1.79 | 64.74 |  |  |  |
| Others | distressed (bin.) | No longer employed | ELSA |  |  |  |  |  | low counts |  |
| Others | distressed (bin.) | No longer employed | GS | 2.87 | 0.83 | 9.87 | 3.65 |  |  |  |
| Others | distressed (bin.) | No longer employed | ALSPAC(G0) | |  |  |  |  | low counts |  |
| Others | distressed (bin.) | No longer employed | Twins UK |  |  |  |  |  | low counts |  |
| Others | distressed (bin.) | No longer employed | Overall | 1.37 | 1.08 | 1.74 |  | 0.00 |  |  |
| Others | distressed (bin.) | Stable unemployed | MCS | 1.22 | 0.60 | 2.49 | 15.40 |  |  | REML |
| Others | distressed (bin.) | Stable unemployed | ALSPAC(G1) | 1.61 | 0.71 | 3.67 | 14.02 |  |  |  |
| Others | distressed (bin.) | Stable unemployed | NS | 1.82 | 1.05 | 3.17 | 17.48 |  |  |  |
| Others | distressed (bin.) | Stable unemployed | BCS70 |  |  |  |  |  | low counts |  |
| Others | distressed (bin.) | Stable unemployed | NCDS | 9.60 | 3.55 | 25.93 | 11.97 |  |  |  |
| Others | distressed (bin.) | Stable unemployed | USOC | 0.90 | 0.47 | 1.75 | 16.10 |  |  |  |
| Others | distressed (bin.) | Stable unemployed | ELSA | 2.12 | 0.86 | 5.21 | 13.05 |  |  |  |
| Others | distressed (bin.) | Stable unemployed | GS |  |  |  |  |  | low counts |  |
| Others | distressed (bin.) | Stable unemployed | ALSPAC(G0) | 1.46 | 0.54 | 3.95 | 11.98 |  |  |  |
| Others | distressed (bin.) | Stable unemployed | Twins UK |  |  |  |  |  | low counts |  |
| Others | distressed (bin.) | Stable unemployed | Overall | 1.82 | 1.10 | 3.03 |  | 66.51 |  |  |
| Others | distressed (cont.) | Furloughed | MCS | -0.01 | -0.19 | 0.17 | 16.17 |  |  | REML |
| Others | distressed (cont.) | Furloughed | ALSPAC(G1) | 0.27 | -0.06 | 0.60 | 9.31 |  |  |  |
| Others | distressed (cont.) | Furloughed | NS | 0.10 | -0.16 | 0.36 | 12.05 |  |  |  |
| Others | distressed (cont.) | Furloughed | BCS70 | 0.40 | 0.09 | 0.71 | 10.16 |  |  |  |
| Others | distressed (cont.) | Furloughed | NCDS | 0.20 | -0.18 | 0.58 | 7.84 |  |  |  |
| Others | distressed (cont.) | Furloughed | USOC | -0.08 | -0.20 | 0.03 | 19.97 |  |  |  |
| Others | distressed (cont.) | Furloughed | ELSA | 0.09 | -0.28 | 0.45 | 8.33 |  |  |  |
| Others | distressed (cont.) | Furloughed | GS | 0.57 | -1.30 | 2.45 | 0.47 |  |  |  |
| Others | distressed (cont.) | Furloughed | ALSPAC(G0) | -0.15 | -0.34 | 0.04 | 15.71 |  |  |  |
| Others | distressed (cont.) | Furloughed | Twins UK |  |  |  |  |  | low counts |  |
| Others | distressed (cont.) | Furloughed | Overall | 0.06 | -0.07 | 0.19 |  | 55.14 |  |  |
| Others | distressed (cont.) | No longer employed | MCS | -0.10 | -0.39 | 0.18 | 15.55 |  |  | REML |
| Others | distressed (cont.) | No longer employed | ALSPAC(G1) | 0.13 | -0.27 | 0.54 | 13.63 |  |  |  |
| Others | distressed (cont.) | No longer employed | NS | -0.37 | -1.06 | 0.33 | 9.22 |  |  |  |
| Others | distressed (cont.) | No longer employed | BCS70 | 0.55 | -0.13 | 1.23 | 9.45 |  |  |  |
| Others | distressed (cont.) | No longer employed | NCDS | 0.77 | -0.04 | 1.59 | 7.77 |  |  |  |
| Others | distressed (cont.) | No longer employed | USOC | 0.39 | 0.14 | 0.65 | 15.97 |  |  |  |
| Others | distressed (cont.) | No longer employed | ELSA | -0.55 | -0.94 | -0.17 | 13.92 |  |  |  |
| Others | distressed (cont.) | No longer employed | GS | 3.69 | -1.44 | 8.82 | 0.34 |  |  |  |
| Others | distressed (cont.) | No longer employed | ALSPAC(G0) | 0.46 | 0.09 | 0.83 | 14.15 |  |  |  |
| Others | distressed (cont.) | No longer employed | Twins UK |  |  |  |  |  | low counts |  |
| Others | distressed (cont.) | No longer employed | Overall | 0.14 | -0.16 | 0.45 |  | 73.60 |  |  |
| Others | distressed (cont.) | Stable unemployed | MCS | 0.18 | -0.12 | 0.47 | 11.35 |  |  | REML |
| Others | distressed (cont.) | Stable unemployed | ALSPAC(G1) | -0.03 | -0.47 | 0.41 | 11.33 |  |  |  |
| Others | distressed (cont.) | Stable unemployed | NS | 0.47 | -0.11 | 1.05 | 11.29 |  |  |  |
| Others | distressed (cont.) | Stable unemployed | BCS70 | 0.36 | -0.08 | 0.79 | 11.33 |  |  |  |
| Others | distressed (cont.) | Stable unemployed | NCDS | 1.66 | 0.99 | 2.33 | 11.26 |  |  |  |
| Others | distressed (cont.) | Stable unemployed | USOC | 0.12 | -0.21 | 0.44 | 11.35 |  |  |  |
| Others | distressed (cont.) | Stable unemployed | ELSA | 0.87 | 0.14 | 1.60 | 11.23 |  |  |  |
| Others | distressed (cont.) | Stable unemployed | GS | 11.82 | 8.97 | 14.67 | 9.52 |  |  |  |
| Others | distressed (cont.) | Stable unemployed | ALSPAC(G0) | 0.27 | -0.06 | 0.60 | 11.35 |  |  |  |
| Others | distressed (cont.) | Stable unemployed | Twins UK |  |  |  |  |  | low counts |  |
| Others | distressed (cont.) | Stable unemployed | Overall | 1.56 | -0.62 | 3.75 |  | 99.52 |  |  |
| Others | less than daily contact | Furloughed | MCS | 0.80 | 0.63 | 1.00 | 16.51 |  |  | REML |
| Others | less than daily contact | Furloughed | ALSPAC(G1) | 2.62 | 1.09 | 6.29 | 2.34 |  |  |  |
| Others | less than daily contact | Furloughed | NS | 0.84 | 0.62 | 1.15 | 12.09 |  |  |  |
| Others | less than daily contact | Furloughed | BCS70 | 1.29 | 1.00 | 1.66 | 15.22 |  |  |  |
| Others | less than daily contact | Furloughed | NCDS | 0.99 | 0.70 | 1.40 | 10.70 |  |  |  |
| Others | less than daily contact | Furloughed | USOC | 1.00 | 0.90 | 1.12 | 25.32 |  |  |  |
| Others | less than daily contact | Furloughed | ELSA | 1.16 | 0.81 | 1.65 | 10.30 |  |  |  |
| Others | less than daily contact | Furloughed | GS | 1.14 | 0.72 | 1.80 | 7.08 |  |  |  |
| Others | less than daily contact | Furloughed | ALSPAC(G0) | 0.45 | 0.06 | 3.62 | 0.44 |  |  |  |
| Others | less than daily contact | Furloughed | Twins UK |  |  |  |  |  | low counts |  |
| Others | less than daily contact | Furloughed | Overall | 1.02 | 0.89 | 1.18 |  | 44.05 |  |  |
| Others | less than daily contact | No longer employed | MCS | 0.93 | 0.67 | 1.28 | 24.46 |  |  | REML |
| Others | less than daily contact | No longer employed | ALSPAC(G1) | 2.77 | 0.90 | 8.48 | 2.01 |  |  |  |
| Others | less than daily contact | No longer employed | NS | 0.63 | 0.31 | 1.29 | 4.95 |  |  |  |
| Others | less than daily contact | No longer employed | BCS70 |  |  |  |  |  | low counts |  |
| Others | less than daily contact | No longer employed | NCDS |  |  |  |  |  | low counts |  |
| Others | less than daily contact | No longer employed | USOC | 0.97 | 0.78 | 1.20 | 54.08 |  |  |  |
| Others | less than daily contact | No longer employed | ELSA | 0.80 | 0.43 | 1.47 | 6.68 |  |  |  |
| Others | less than daily contact | No longer employed | GS | 0.76 | 0.40 | 1.43 | 6.33 |  |  |  |
| Others | less than daily contact | No longer employed | ALSPAC(G0) | 1.10 | 0.30 | 4.08 | 1.48 |  |  |  |
| Others | less than daily contact | No longer employed | Twins UK |  |  |  |  |  | low counts |  |
| Others | less than daily contact | No longer employed | Overall | 0.93 | 0.80 | 1.09 |  | 0.00 |  |  |
| Others | less than daily contact | Stable unemployed | MCS | 0.89 | 0.58 | 1.36 | 15.78 |  |  | REML |
| Others | less than daily contact | Stable unemployed | ALSPAC(G1) | 5.20 | 2.17 | 12.49 | 9.52 |  |  |  |
| Others | less than daily contact | Stable unemployed | NS | 0.90 | 0.54 | 1.50 | 14.45 |  |  |  |
| Others | less than daily contact | Stable unemployed | BCS70 | 1.24 | 0.83 | 1.86 | 16.10 |  |  |  |
| Others | less than daily contact | Stable unemployed | NCDS | 1.65 | 1.11 | 2.44 | 16.30 |  |  |  |
| Others | less than daily contact | Stable unemployed | USOC | 1.00 | 0.82 | 1.21 | 18.81 |  |  |  |
| Others | less than daily contact | Stable unemployed | ELSA |  |  |  |  |  | low counts |  |
| Others | less than daily contact | Stable unemployed | GS |  |  |  |  |  | low counts |  |
| Others | less than daily contact | Stable unemployed | ALSPAC(G0) | 2.18 | 0.87 | 5.46 | 9.04 |  |  |  |
| Others | less than daily contact | Stable unemployed | Twins UK |  |  |  |  |  | low counts |  |
| Others | less than daily contact | Stable unemployed | Overall | 1.36 | 0.94 | 1.98 |  | 80.09 |  |  |
| Others | fair or poor self-rated health | Furloughed | MCS | 1.64 | 0.67 | 4.06 | 10.10 |  |  | MLE |
| Others | fair or poor self-rated health | Furloughed | ALSPAC(G1) | |  |  |  |  | no measure |  |
| Others | fair or poor self-rated health | Furloughed | NS | 1.37 | 0.62 | 3.04 | 12.34 |  |  |  |
| Others | fair or poor self-rated health | Furloughed | BCS70 | 3.15 | 1.51 | 6.55 | 13.95 |  |  |  |
| Others | fair or poor self-rated health | Furloughed | NCDS | 1.30 | 0.49 | 3.44 | 8.98 |  |  |  |
| Others | fair or poor self-rated health | Furloughed | USOC |  |  |  |  |  | no measure |  |
| Others | fair or poor self-rated health | Furloughed | ELSA | 1.10 | 0.95 | 1.28 | 46.32 |  |  |  |
| Others | fair or poor self-rated health | Furloughed | GS | 1.49 | 0.54 | 4.12 | 8.31 |  |  |  |
| Others | fair or poor self-rated health | Furloughed | ALSPAC(G0) | |  |  |  |  | no measure |  |
| Others | fair or poor self-rated health | Furloughed | Twins UK |  |  |  |  |  | low counts |  |
| Others | fair or poor self-rated health | Furloughed | Overall | 1.42 | 1.03 | 1.95 |  | 32.96 |  |  |
| Others | fair or poor self-rated health | No longer employed | MCS |  |  |  |  |  | low counts | REML |
| Others | fair or poor self-rated health | No longer employed | ALSPAC(G1) | |  |  |  |  | no measure |  |
| Others | fair or poor self-rated health | No longer employed | NS |  |  |  |  |  | low counts |  |
| Others | fair or poor self-rated health | No longer employed | BCS70 |  |  |  |  |  | low counts |  |
| Others | fair or poor self-rated health | No longer employed | NCDS |  |  |  |  |  | low counts |  |
| Others | fair or poor self-rated health | No longer employed | USOC |  |  |  |  |  | no measure |  |
| Others | fair or poor self-rated health | No longer employed | ELSA | 1.33 | 1.04 | 1.69 | 100.00 |  |  |  |
| Others | fair or poor self-rated health | No longer employed | GS |  |  |  |  |  | low counts |  |
| Others | fair or poor self-rated health | No longer employed | ALSPAC(G0) | |  |  |  |  | no measure |  |
| Others | fair or poor self-rated health | No longer employed | Twins UK |  |  |  |  |  | low counts |  |
| Others | fair or poor self-rated health | No longer employed | Overall | 1.33 | 1.04 | 1.69 |  |  |  |  |
| Others | fair or poor self-rated health | Stable unemployed | MCS | 4.54 | 1.69 | 12.20 | 47.20 |  |  | REML |
| Others | fair or poor self-rated health | Stable unemployed | ALSPAC(G1) | |  |  |  |  | no measure |  |
| Others | fair or poor self-rated health | Stable unemployed | NS | 1.80 | 0.74 | 4.37 | 52.80 |  |  |  |
| Others | fair or poor self-rated health | Stable unemployed | BCS70 |  |  |  |  |  | low counts |  |
| Others | fair or poor self-rated health | Stable unemployed | NCDS |  |  |  |  |  | low counts |  |
| Others | fair or poor self-rated health | Stable unemployed | USOC |  |  |  |  |  | no measure |  |
| Others | fair or poor self-rated health | Stable unemployed | ELSA |  |  |  |  |  | low counts |  |
| Others | fair or poor self-rated health | Stable unemployed | GS |  |  |  |  |  | low counts |  |
| Others | fair or poor self-rated health | Stable unemployed | ALSPAC(G0) | |  |  |  |  | no measure |  |
| Others | fair or poor self-rated health | Stable unemployed | Twins UK |  |  |  |  |  | low counts |  |
| Others | fair or poor self-rated health | Stable unemployed | Overall | 2.78 | 1.12 | 6.89 |  | 46.48 |  |  |
