## Supplementary File 4 for "Mental and social wellbeing and the UK Coronavirus Job Retention Scheme: evidence from nine longitudinal studies"

**Supplementary File 4** – **figures for subgroup analyses**

**Figure a.1. Stratification by gender in the fully adjusted model**

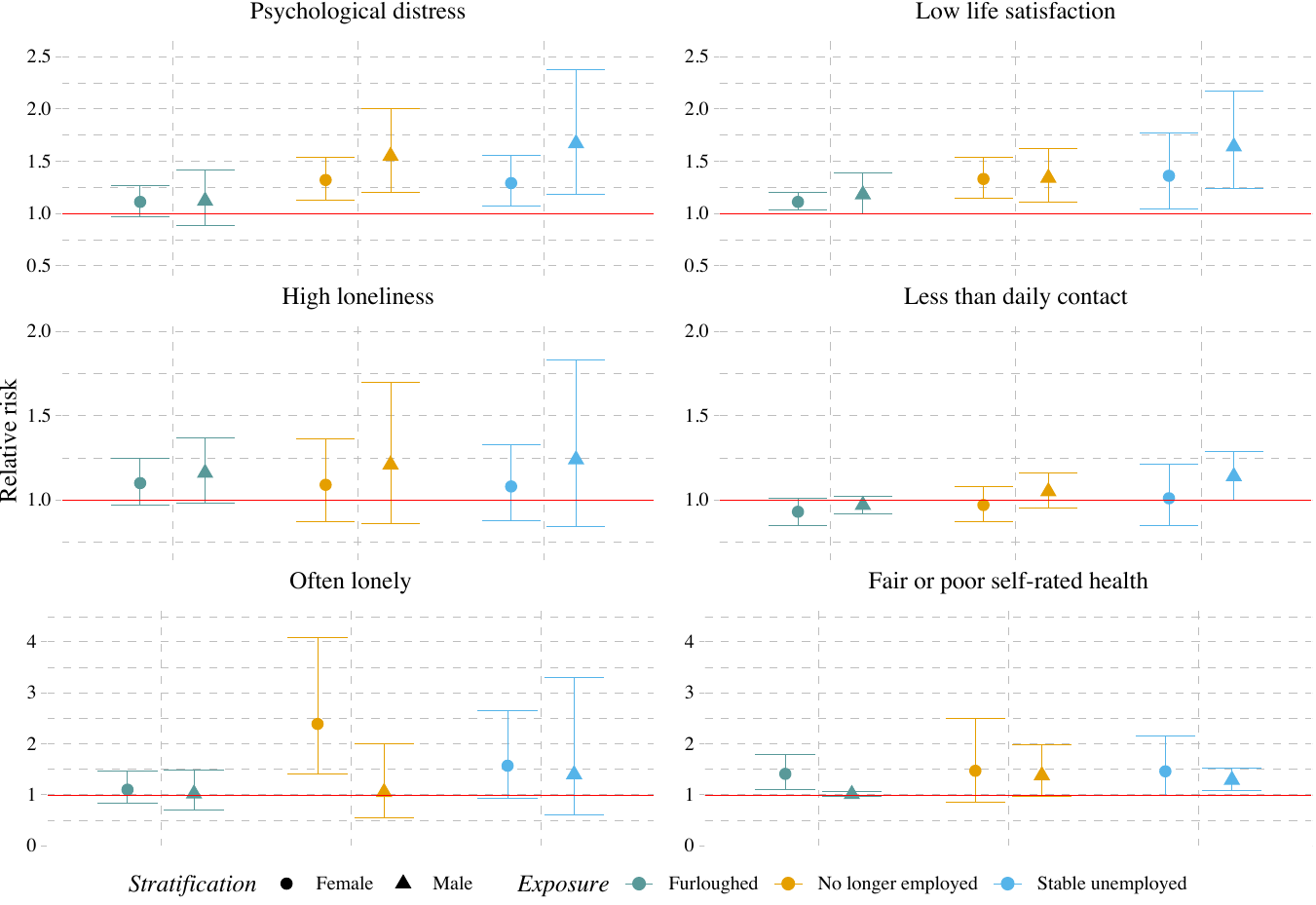

**Figure a.2. Stratification by living arrangements in the fully adjusted model**

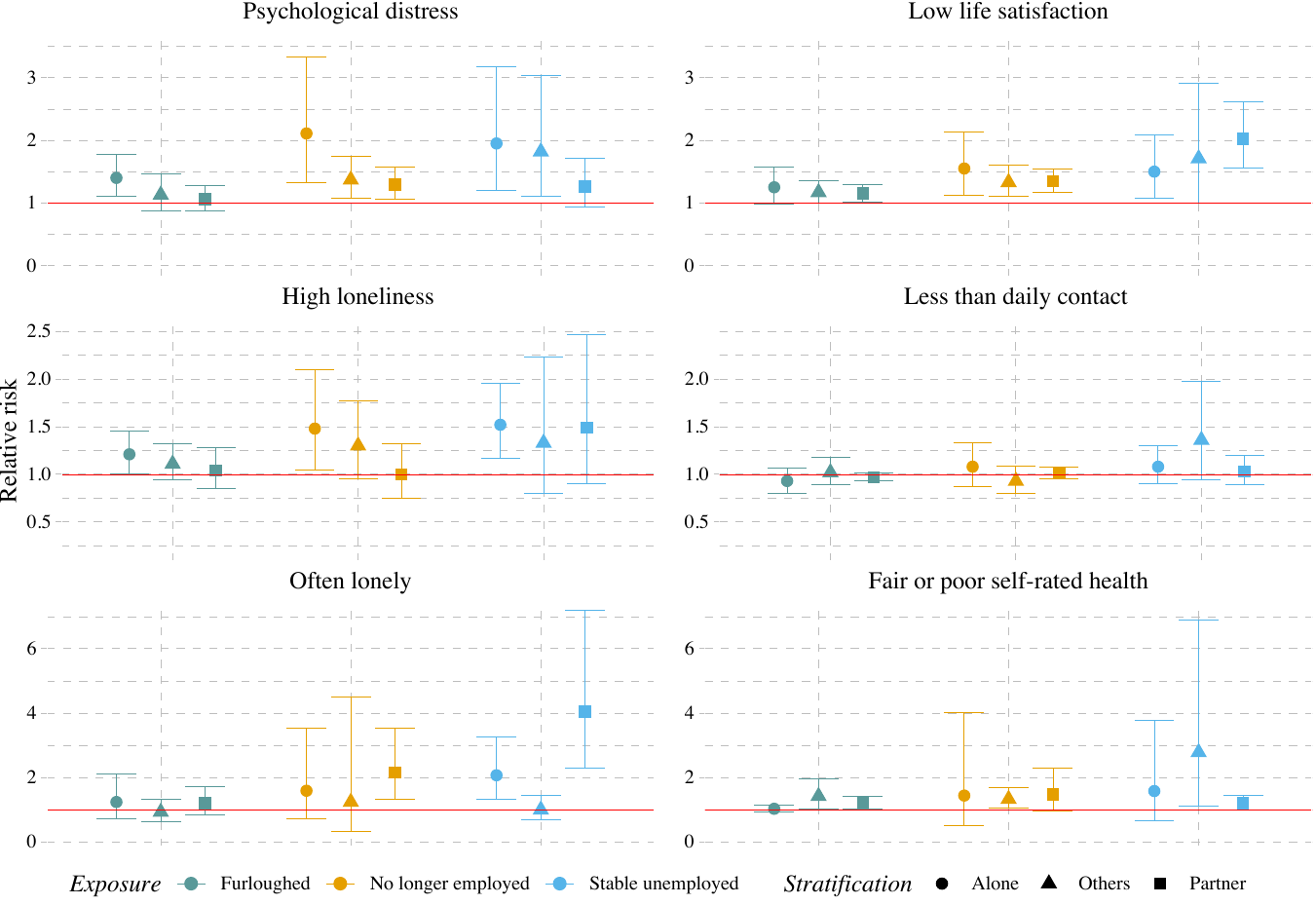

**Figure a.3. Stratification by highest level of education in the fully adjusted model**

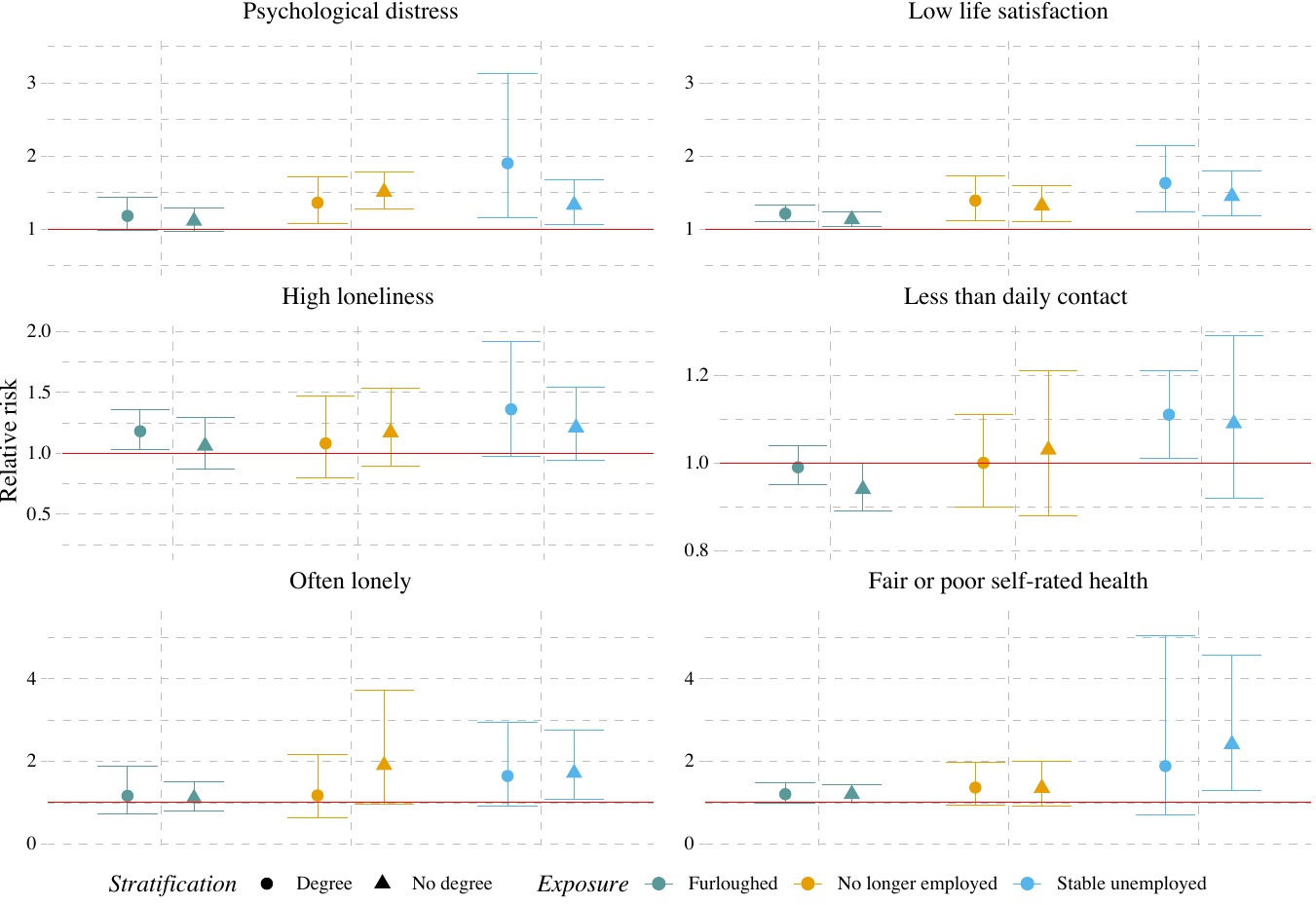

**Figure a.4. Stratification by age group in the fully adjusted model**

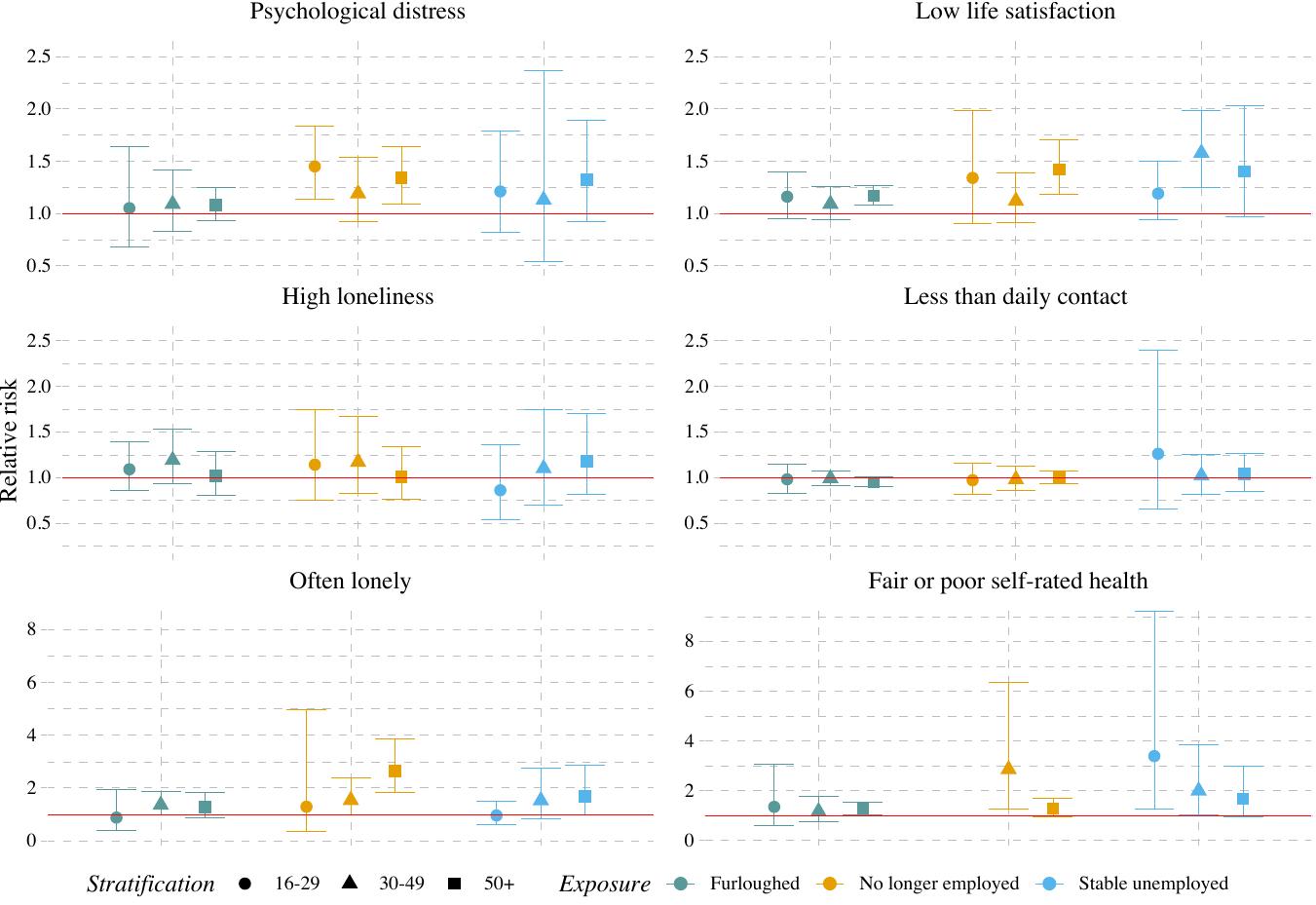
